# Integrative Genetic, Proteogenomic, and Multi-omics Analyses Reveal Sex-Biased Causal Genes and Drug Targets in Alzheimer’s Disease

**DOI:** 10.1101/2025.10.31.25339089

**Authors:** Noah Cook, Chenyu Yang, Youjie Zeng, Sathesh K. Sivasankaran, Soomin Song, Lia Talozzi, Daniel Western, Chengran Yang, Yue Liu, Yann Le Guen, Ilaria Stewart, Christina Young, FinnGen, Elizabeth C. Mormino, Andre Altmann, Zihuai He, Valerio Napolioni, Aliza P. Wingo, Thomas S. Wingo, Carlos Cruchaga, Yun Ju Sung, Michael D. Greicius, Michael E. Belloy

## Abstract

Sex differences are pervasive in Alzheimer’s disease, but the underlying drivers remain poorly understood. To address this, we performed sex-stratified genome-wide association studies of Alzheimer’s disease in ∼1,000,000 individuals, which we subsequently integrated with proteogenomics datasets from neurological tissues to identify candidate causal genes. We further prioritized genes through additional multi-omics approaches, including quantitative trait locus summary-based mendelian randomization and colocalization. Altogether, we prioritized 125 female-biased and 21 male-biased risk genes. Female-biased pathways included amyloid, neurite, stress, clearance, and immune processes, with genes enriched for microglia and astrocyte expression. Through computational drug repurposing analyses, a set of sex hormone related drugs, converging on *Epidermal Growth Factor Receptor* (*EGFR*), were uniquely prioritized in women. Finally, we identified *Haptoglobin* (*HP*) as a female-specific gene, leveraging long-read sequencing approaches to implicate a link to oxidative stress, APOE, and hemoglobin biology. Altogether, our findings provide a portal into sex-specific precision medicine for Alzheimer’s disease.

## Introduction

Sex differences are central to late-onset Alzheimer’s disease (AD)^1,2^, with women displaying higher lifetime prevalence^3^, higher incidence rates at old age^4^, and greater risk related to *APOE* ε4^5–7^–the strongest, common genetic risk factor for AD–while also differing from men in clinicopathological trajectories^8–10^, resilience^10,11^, and response to anti-amyloid therapies^12^. These differences have been tied to sex hormones and chromosomes, and pathways including immune function, metabolism, and autophagy^13,14^, yet major gaps remain in understanding the causal genetic and molecular mechanisms underlying this dimorphism^15^.

While genome-wide association studies (GWAS) have transformed our understanding of AD, identifying over 70 risk loci^16,17^, sex-stratified GWAS have remained comparatively small, revealing only select sex-biased genetic loci^18–22^. Most omics studies investigating AD sex differences highlight female-specific molecular changes in brain, cerebrospinal fluid (CSF), and plasma, but these remain to be causally anchored to AD genetic factors^23^. To start bridging this gap, Wingo *et al*. conducted the largest-to-date brain-tissue study to identify genes displaying sex-biased genetic regulation of their expression (expression quantitative trait loci, eQTLs) or protein levels (pQTLs) and integrated these with neurodegenerative GWAS, identifying 6 sex-biased AD causal proteins^24^. Variant-to-protein studies, i.e. proteogenomics, are especially important given low protein-transcript correlation and proteins being more direct disease actuators^25,26^. Along this line, Western *et al*. and Song *et al*. built the largest-to-date CSF proteogenomics resource and integrated it with AD GWAS, mapping many causal gene-AD associations including a handful that were sex-biased^27,28^. Across these and other sex-aware studies^29,30^, shared limitations included the lack or small size of sex-stratified GWAS and incomplete assessment of how sex-biases in QTLs influence gene prioritization, likely explaining the relative paucity of causal sex-biased gene-AD findings.

Here, we addressed these gaps by performing a sex-stratified GWAS of AD in nearly 1,000,000 individuals and integrating findings with brain and CSF proteogenomics data, conducting proteome-wide association studies (PWAS) to implicate sex-biased AD proteins. Moving beyond prior work, we evaluated colocalization of sex-biased AD genetic signals with multi-tissue multi-QTL datasets, broadening gene prioritization and composing a comprehensive set of likely causal sex-biased genes. We then leveraged these to identify sex-biased biological pathways, cellular contexts, and repurposable drugs. Finally, we highlight *Haptoglobin*, leveraging long-read sequencing (LRS) data and functional genomics, to identify causal variation and mechanisms linking *Haptoglobin* to AD risk uniquely in women.

## Results

### Sex-stratified Alzheimer’s disease GWAS

We conducted multi-stage meta-analyses of European ancestry (EUR) sex-stratified AD GWAS (**Fig.1A**). Stage-1 included clinically diagnosed cases (40% pathology confirmed) and controls from the Alzheimer’s Disease Genetics Consortium (ADGC) and Alzheimer’s Disease Sequencing Project (ADSP; **Tables-S1-2**, admixture **Fig.S1**)^31,32^. Stage-2 used UK Biobank, incorporating health-registry defined cases and questionnaire-identified proxy-cases whose parents were affected by AD or dementia^33^. The “proxy-AD” GWAS approach has proven powerful in the study of AD genetics and was tailored here to map sex-specificity (cf. Methods)^17,34,35^. Stage-3 leveraged health-registry defined cases from FinnGen^36^. Total sample sizes were N_Females_=636,155 and N_Males_=507,678, while effective sample sizes accounting for case-control imbalance and proxy-AD GWAS were N_Females-effective_=111,477 and N_Males-effective_=86,200 (case counts **Fig.1A**, demographics **Table-S3**, variant counts **Table-S4**, quantile-quantile plots **Fig.S2**). Considering the established female-biased effect of *APOE* ε4 in AD^5–7^, GWAS excluded the *APOE* locus and adjusted for *APOE* ε4 and *APOE* ε2 dosage to account for sex-by-*APOE* interactions (cf. Methods).

**Figure 1.**
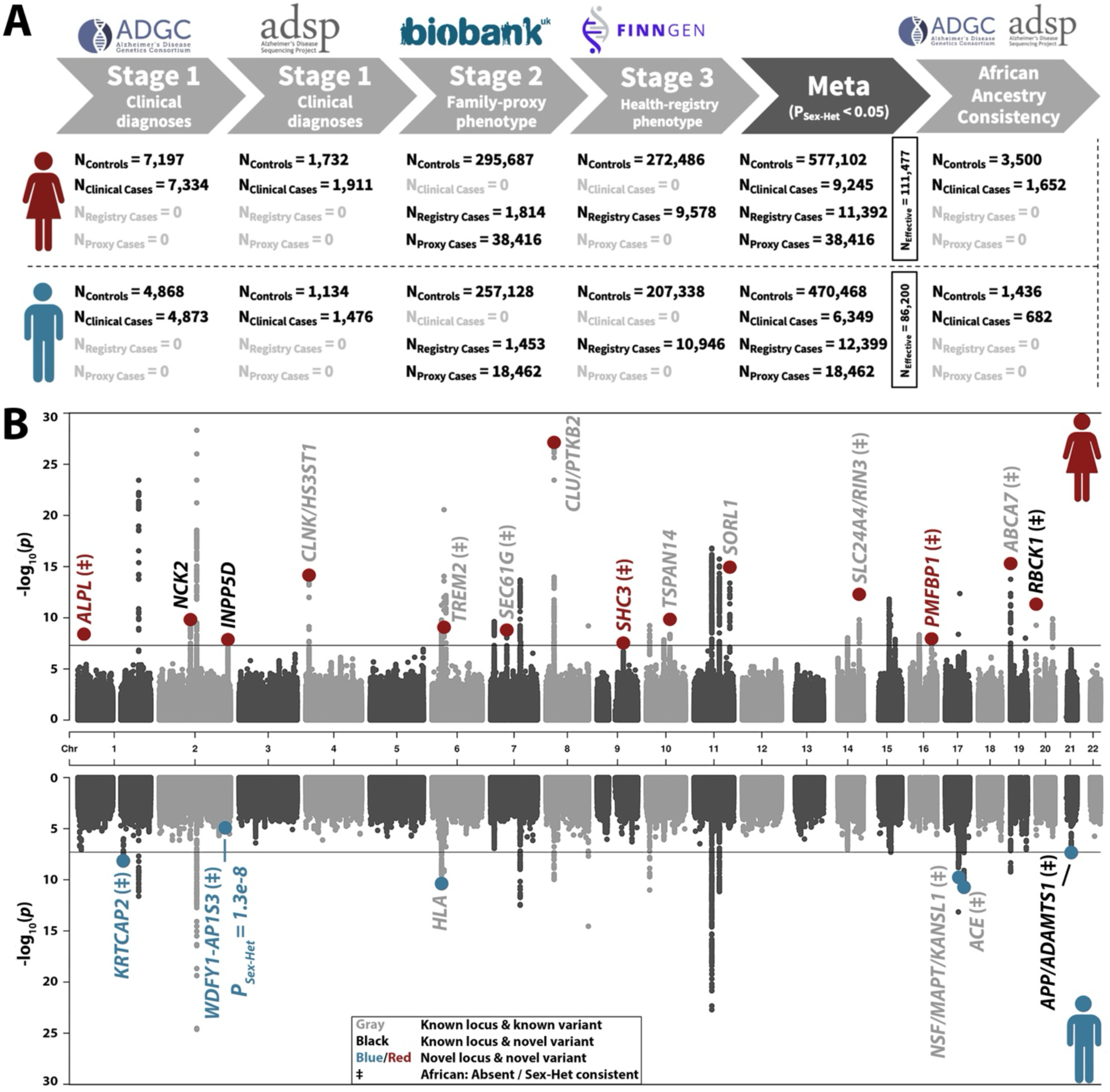
Sex-stratified genome-wide association study of Alzheimer’s disease. **A)** Overview of study design and sample sizes. **B)** Miami plot for the GWAS meta-analyses; top, women; bottom, men. The solid lines indicate genome-wide significance (*P*<5×10^-8^); plots are truncated at *P*<1×10^-30^; Colored dots indicate index variants at significant loci that also passed sex heterogeneity criteria; one variant is marked below genome-wide significance in men but passed genome-wide significance in the sex heterogeneity GWAS. The legend details annotations regarding variant novelty and sex heterogeneity consistency in African ancestry (AFR). Lead variants for independent loci are annotated with their nearest protein-coding gene (Gencode v47) or known AD risk locus.

We then mapped independent sex-stratified signals whose lead variants had P<5e−8 and sex-heterogeneity P_Sex-Het_<0.05, which identified 14 female and 5 male-biased loci with pronounced sex differences in effect magnitude (ratios ≥1.5; **Fig.1B, Table-1**). One additional locus, *WDFY1-AP1S3*, was identified in sex-heterogeneity GWAS (P_Sex-Het_=1.3e-8; **Fig.S3**). Of the 20 loci, 11 were signals reported in previous AD GWAS, 4 were novel independent signals at known loci (**Table-S5**), and 5 were novel loci, including female-biased *ALPL*, *SHC3*, *PMFBP1* and male-biased *KRTCAP2*, *WDFY1-AP1S3* (locus names reflect most proximal genes; locus plots with all mapped signals **Fig.S4;** variant forest plots **Fig.S5**). Across novel loci, only *ALPL* was marked by a rare missense variant, rs121918007 (OR_Females_=0.68, P_Females_=4.0e-9, OR_Males_=0.90, P_Males_=0.067), with a high predicted deleteriousness score (**Table-S6**) and detection enabled by inclusion of FinnGen given elevated frequencies in Finnish versus non-Finnish Europeans (gnomAD AF 1.70% versus 0.14%). Sex-stratified GWAS in a small African ancestry dataset (AFR; N=7,270; **Tables-S7-8**, **Fig.S6**) showed 13/20 loci had consistent sex-heterogeneity or were unique to EUR subjects (**Table-1, Table-S9**). The *HLA* locus, already displaying the weakest sex-heterogeneity in EUR GWAS (P_Sex-Het_=0.049), was distinctly discordant for sex-heterogeneity in AFR GWAS, indicating it was not robustly sex-biased. Notably, the male-biased rs12726330 variant in the *KRTCAP2* locus (OR_Females_=1.04, P_Females_=0.40, OR_Males_=1.20, P_Males_=7.1e-9) colocalized with the *GBA* risk locus for Parkinson’s disease (PD; **Fig.S7A**)^37^. In follow-up analyses, we confirmed this related to Lewy Body dementia (LBD) cases being part of AD phenotypes, even in Stage-1 clinically diagnosed cohorts (**Fig.S7B**). Nonetheless, we observed other sex-biased loci to have highly consistent effect sizes across GWAS Stages (**Fig.S8**), while their AD relevance is corroborated in subsequent gene prioritizations.

### Sex-stratified Alzheimer’s disease PWAS

Next, we performed PWAS integrating our sex-stratified AD GWAS with proteogenomics resources in brain and CSF to identify proteins whose genetically predicted levels likely exert causal, sex-biased effects on AD. Primary analyses matched GWAS and proteogenomics data by sex, similar to prior studies^24,28,29^, while secondary analyses did not stratify proteogenomics data by sex to assess to what extent this helps identify additional sex-biased proteins; Both discoveries were further filtered to ensure sex-specificity (**Fig.2A**).

**Figure 2.**
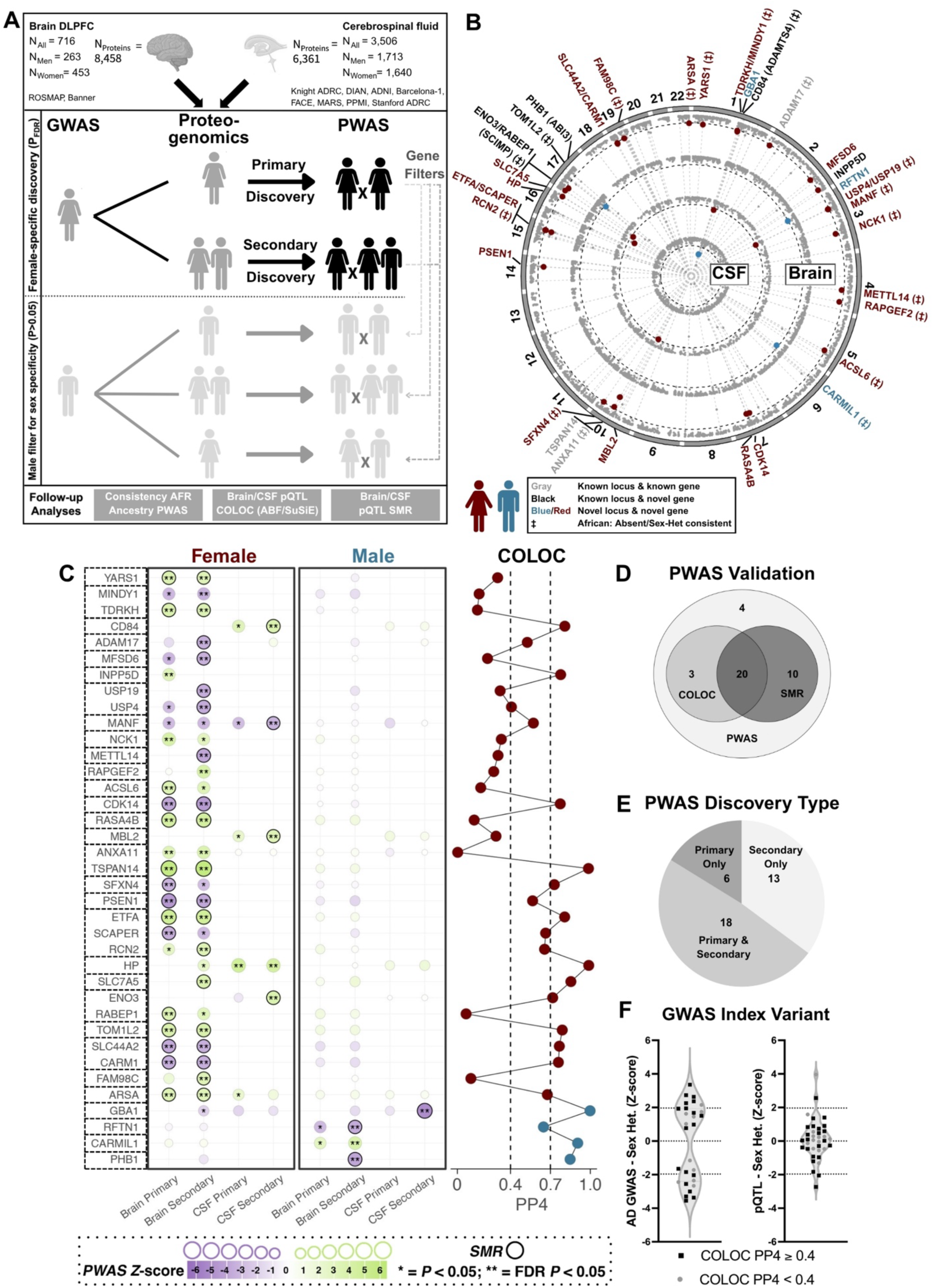
Sex-stratified proteome-wide association studies of Alzheimer’s disease in brain and Cerebrospinal fluid. **A)** Illustration of the PWAS study design for identification of female-specific AD risk genes. The discovery design for men is equivalent but inverted. **B)** Circos plot of AD PWAS analyses, showing from outer to inner layers: female-brain, male-brain, female-CSF, male-CSF. Per protein, only the best findings across discoveries are displayed. Plots are truncated at *P*<1x10^-15^. The black dotted lines denote the proteome-wide significance thresholds at *P_FDR_*<0.05. Proteins passing sex-specificity criteria are annotated regarding novelty and sex heterogeneity consistency in African ancestry data (cf. legend). **C)** Left. Dot matrix summarizes PWAS Z-scores for all identified sex-biased proteins; positive and negative Z-scores respectively indicate that higher or lower proteins levels increase risk. Proteins with pQTL SMR support are marked by black circles. Right. pQTL COLOC support was considered suggestive at PP4>0.4 and strong at PP4>0.7. **D)** Venn diagram summarizes SMR and COLOC support, indicating 33 out of 37 proteins had at least 1 level of support. **E)** Venn diagram summarizes the types of PWAS discoveries, showing their complimentary value. **F)** Sex-heterogeneity is shown for top AD genetic risk variants and top pQTLs at respective PWAS loci, with dotted lines reflecting P_Sex-Het_<0.05. While top AD genetic risk variants at respective PWAS loci showed substantial sex heterogeneity in AD, this was less apparent for top pQTLs.

We identified 37 proteins (28 female-brain, 5 female-CSF, 3 male-brain, 1 male-CSF) significant in one sex (P_FDR_<0.05) but not the other (P>0.05) across 30 unique loci (**Fig. 2B**; **Tables-S10-17**, **Figs.S9-10**). These comprised 28 novel genes at novel AD loci, 6 novel genes at known loci, and 3 known AD genes (**Table 2**), with 4 loci overlapping the 20 sex-biased GWAS loci (**Fig.1**; *INPP5D*, *TSPAN14*, *KRTCAP2/GBA1, PMBFBP1/HP*). Among the strongest associations in women were TSPAN14 (Z=5.88), a known AD gene implicated in amyloid processing^17^, PSEN1 (Z=-4.57), linked to early-onset AD but discovered here in the context of late-onset AD^38^, ETFA (Z=4.84), relevant to mitochondrial metabolism, and HP (Z=4.76), which impacts APOE biology^39^. The proportion of female-biased proteins was high (89%) and remained high (65%) even when male significance thresholds were adjusted to balance power (**Fig.S11**). While heritable proteins across brain and CSF showed limited overlap (9%; **Fig.S12**), 5/6 overlapping sex-biased proteins had consistent effect directions with 3 replicating nominally (**Table-S18**). Out of 37 proteins, 20 also displayed consistent sex-heterogeneity in AFR PWAS (**Table-2**, **Table-S19**). Importantly, when repeating PWAS after excluding Stage-2 from the sex-stratified GWAS to avoid potential biases due to proxy-AD samples^40^, 34/37 genes showed consistent associations (**Table-S20**).

**Table 1.**
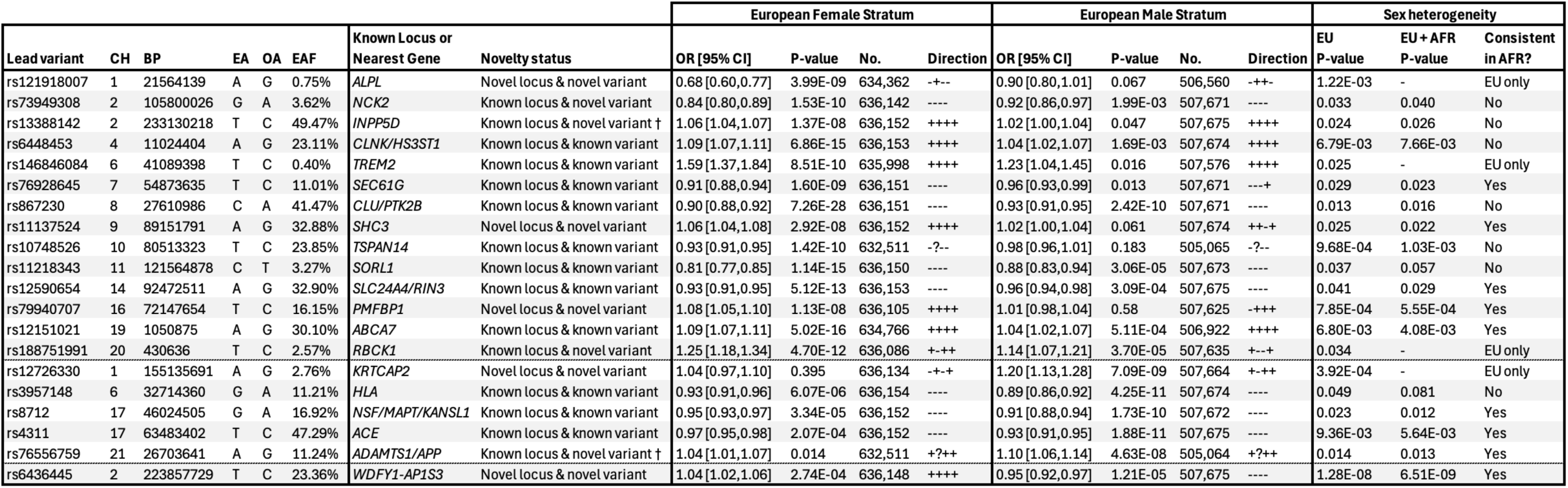
Sex-stratified genome-wide association study of Alzheimer’s disease: Associated lead variants. Listed lead variants passed GWAS significance thresholds (*P*<5e^-8^) in a respective stratum as well as sex heterogeneity significances thresholds (*P_Sex-Het_*<0.05), or, passed GWAS significance thresholds for sex heterogeneity tests (*P*_Sex-Het_<5e^-8^). The ‘Direction’ columns indicate the association effect direction across meta-analyzed cohorts following the order of ADGC, ADSP, UKB, and FinnGen; A question mark indicates the variant was not available in the respective cohort. Variants are annotated using dbSNP153, while novel genes are annotated using GENCODE v47. Sex heterogeneity P-values are shown for the main European ancestry meta-analyses, as well as additional meta-analyses across European and African ancestry to assess consistency (i.e. improved *P*_Sex-Het_ upon meta-analysis). † Two respective lead variants showed linkage disequilibrium at R^2^>0.01 with known AD lead variants from prior GWAS but still appeared to reflect novel independent signals (cf. Fig.S4). *Abbreviations: OR, odds ratio; CI, confidence interval; EA, effect allele; OA, other allele; EAF, effect allele frequency; CH, chromosome; BP, base pair; No., sample size; P*_Sex-Het_*, Sex heterogeneity P-value*.

**Table 2.** Sex-stratified proteome-wide association study of Alzheimer’s disease: Associated genes. Listed proteins/genes passed FDR significance thresholds in respective strata, tissues, and discovery types in the European sex-stratified PWAS, while not reaching nominal significance (*P*>0.05) in the opposite sex (the most significant findings are shown for each protein). The two discovery types are described in Figure 2A, with “Prim+Sec” indicating a protein was associated in both types of discoveries. Shaded cells indicate respective loci (with some loci corroborating multiple protein associations). Gene novelty status is indicated as “Known locus & known gene”, i.e. an AD-associated gene from prior AD GWAS that has had functional prioritization support, a “Known locus & novel gene”, i.e. an AD-associated gene from prior AD GWAS that has not had functional prioritization support, and “Novel locus & novel gene” with regard to prior AD GWAS. European findings were considered consistent in African ancestry analyses if the P-value in the discovery sex improved after sample-size weighted meta-analyses of the EUR and AFR findings, while the opposite sex maintained *P*>0.05. † Protein was analyzed in female-specific proteogenomic data but not in male or non-stratified data as it did not display significant SNP-heritability in those strata. PWAS values reported for the male strata were therefore derived from combining male AD GWAS with female-derived protein weights. *Abbreviations: PWAS, protein wide association studies; GWAS, genome-wide association study; FDR, false-discovery rate corrected; EUR, European; AFR, African*.

|  |  |  |  |  |  |  | European PWAS |  |  |  |  |  | Sex-het. |
| --- | --- | --- | --- | --- | --- | --- | --- | --- | --- | --- | --- | --- | --- |
|  |  |  |  |  |  |  | Female Stratum |  |  | Male Stratum |  |  | consistent<br>in AFR? |
| Protein | CH | Gene start | Gene end | Novelty status | Discovery | Tissue | Z-score | P-value | FDR-P | Z-score | P-value | FDR-P |  |
| YARS1 | 1 | 32775237 | 32818031 | Novel locus & novel gene | Prim+Sec | Brain | 3.66 | 2.55E-04 | 1.81E-02 | -0.63 | 0.53 | 0.90 | Yes |
| MINDY1 | 1 | 150996549 | 151008376 | Novel locus & novel gene | Secondary | Brain | -3.48 | 5.04E-04 | 3.25E-02 | -1.47 | 0.14 | 0.68 | Yes |
| TDRKH | 1 | 151770107 | 151791416 | Novel locus & novel gene | Prim+Sec | Brain | 3.75 | 1.78E-04 | 1.35E-02 | -0.39 | 0.70 | 0.92 | Yes |
| CD84 | 1 | 160541095 | 160579516 | Known locus & novel gene | Secondary | CSF | 3.81 | 1.40E-04 | 9.02E-03 | 0.90 | 0.37 | 0.86 | Yes |
| ADAM17 | 2 | 9488486 | 9556732 | Known locus & known gene | Secondary | Brain | -3.54 | 3.96E-04 | 2.93E-02 | -0.72 | 0.47 | 0.87 | Yes |
| MFSD6 | 2 | 190408355 | 190509205 | Novel locus & novel gene | Secondary | Brain | -3.46 | 5.34E-04 | 3.37E-02 | 0.16 | 0.87 | 0.99 | No |
| INPP5D<sup>†</sup> | 2 | 233059967 | 233207903 | Known locus & novel gene | Primary | Brain | 3.36 | 7.72E-04 | 3.81E-02 | 1.74 | 0.082 | 0.58 | No |
| USP19 | 3 | 49108046 | 49120938 | Novel locus & novel gene | Secondary | Brain | -3.62 | 2.95E-04 | 2.43E-02 | -1.24 | 0.22 | 0.75 | Yes |
| USP4 | 3 | 49277144 | 49340712 | Novel locus & novel gene | Secondary | Brain | -3.70 | 2.17E-04 | 1.84E-02 | -0.42 | 0.68 | 0.94 | No |
| MANF | 3 | 51385291 | 51389397 | Novel locus & novel gene | Secondary | CSF | -3.36 | 7.88E-04 | 3.97E-02 | -1.63 | 0.10 | 0.73 | Yes |
| NCK1 | 3 | 136862208 | 136951606 | Novel locus & novel gene | Primary | Brain | 4.08 | 4.48E-05 | 4.92E-03 | 0.83 | 0.41 | 0.78 | Yes |
| METTL14 | 4 | 118685392 | 118715433 | Novel locus & novel gene | Secondary | Brain | -3.94 | 8.01E-05 | 9.89E-03 | 0.16 | 0.87 | 0.99 | Yes |
| RAPGEF2 | 4 | 159103013 | 159360174 | Novel locus & novel gene | Secondary | Brain | 4.09 | 4.33E-05 | 5.83E-03 | 0.36 | 0.72 | 0.92 | Yes |
| ACSL6 | 5 | 131949973 | 132012243 | Novel locus & novel gene | Primary | Brain | 3.58 | 3.40E-04 | 2.06E-02 | 0.20 | 0.84 | 0.98 | Yes |
| CDK14 | 7 | 90466424 | 91210590 | Novel locus & novel gene | Prim+Sec | Brain | -4.41 | 1.01E-05 | 1.50E-03 | -0.46 | 0.65 | 0.93 | No |
| RASA4B | 7 | 102479976 | 102517777 | Novel locus & novel gene | Prim+Sec | Brain | 4.50 | 6.84E-06 | 1.27E-03 | 1.51 | 0.13 | 0.68 | No |
| MBL2 | 10 | 52765380 | 52772784 | Novel locus & novel gene | Secondary | CSF | 3.47 | 5.25E-04 | 2.90E-02 | 1.48 | 0.14 | 0.81 | No |
| ANXA11 | 10 | 80150889 | 80205572 | Known locus & known gene | Prim+Sec | Brain | 3.74 | 1.81E-04 | 1.62E-02 | 0.58 | 0.56 | 0.90 | No |
| TSPAN14 | 10 | 80454265 | 80533124 | Known locus & known gene | Prim+Sec | Brain | 5.88 | 4.04E-09 | 3.99E-06 | 1.27 | 0.20 | 0.74 | Yes |
| SFXN4 | 10 | 119140767 | 119165714 | Novel locus & novel gene | Primary | Brain | -3.38 | 7.17E-04 | 3.61E-02 | -0.46 | 0.65 | 0.90 | Yes |
| PSEN1 | 14 | 73136418 | 73223691 | Novel locus & novel gene | Prim+Sec | Brain | -4.57 | 4.78E-06 | 7.71E-04 | -1.77 | 0.077 | 0.61 | No |
| ETFA | 15 | 76188555 | 76311730 | Novel locus & novel gene | Prim+Sec | Brain | 4.84 | 1.31E-06 | 3.52E-04 | 0.94 | 0.35 | 0.82 | No |
| SCAPER | 15 | 76347904 | 76905444 | Novel locus & novel gene | Primary | Brain | -3.57 | 3.54E-04 | 2.09E-02 | -0.95 | 0.34 | 0.82 | Yes |
| RCN2 | 15 | 76931738 | 76954393 | Novel locus & novel gene | Secondary | Brain | 3.45 | 5.66E-04 | 3.49E-02 | 1.24 | 0.22 | 0.69 | No |
| HP | 16 | 72054505 | 72061055 | Novel locus & novel gene | Prim+Sec | CSF | 4.76 | 1.96E-06 | 2.27E-04 | 1.76 | 0.079 | 0.60 | No |
| SLC7A5 | 16 | 87830016 | 87869507 | Novel locus & novel gene | Secondary | Brain | 3.80 | 1.46E-04 | 1.40E-02 | 1.82 | 0.069 | 0.59 | No |
| ENO3 | 17 | 4948092 | 4957131 | Known locus & novel gene | Secondary | CSF | 3.78 | 1.55E-04 | 9.46E-03 | 0.17 | 0.86 | 0.99 | Yes |
| RABEP1 | 17 | 5282265 | 5386340 | Known locus & novel gene | Primary | Brain | 3.59 | 3.25E-04 | 2.05E-02 | 1.47 | 0.14 | 0.63 | Yes |
| TOM1L2 | 17 | 17843507 | 17972422 | Known locus & novel gene | Prim+Sec | Brain | 3.92 | 8.82E-05 | 7.77E-03 | 1.44 | 0.15 | 0.70 | Yes |
| SLC44A2 | 19 | 10602457 | 10644557 | Novel locus & novel gene | Prim+Sec | Brain | -3.77 | 1.63E-04 | 1.27E-02 | -1.41 | 0.16 | 0.64 | No |
| CARM1 | 19 | 10871553 | 10923075 | Novel locus & novel gene | Prim+Sec | Brain | -3.60 | 3.19E-04 | 2.05E-02 | -1.46 | 0.14 | 0.69 | No |
| FAM98C | 19 | 38403093 | 38409088 | Novel locus & novel gene | Secondary | Brain | 3.38 | 7.36E-04 | 4.36E-02 | 0.23 | 0.82 | 0.98 | Yes |
| ARSA | 22 | 50622754 | 50628173 | Novel locus & novel gene | Prim+Sec | Brain | 3.44 | 5.77E-04 | 3.10E-02 | 0.97 | 0.33 | 0.82 | Yes |
| GBA1 | 1 | 155234452 | 155244699 | Novel locus & novel gene | Secondary | CSF | -1.77 | 0.076 | 0.61 | -5.08 | 3.86E-07 | 4.98E-05 | No |
| RFTN1 | 3 | 16313574 | 16514026 | Novel locus & novel gene | Secondary | Brain | -0.54 | 0.59 | 0.91 | -3.68 | 2.33E-04 | 4.06E-02 | No |
| CARMIL1 | 6 | 25279078 | 25620530 | Novel locus & novel gene | Secondary | Brain | 0.72 | 0.47 | 0.87 | 3.96 | 7.43E-05 | 2.19E-02 | Yes |
| PHB1 | 17 | 49404049 | 49414905 | Known locus & novel gene | Secondary | Brain | -1.09 | 0.27 | 0.76 | -3.94 | 8.14E-05 | 2.19E-02 | No |

We then sought to further validate the sex-biased proteins by comparing AD GWAS and proteogenomics datasets through two complementary methods: pQTL summary-based mendelian randomization (SMR) and genetic colocalization (COLOC; cf. Methods; **Fig.2C**)^41–43^. Since COLOC may be less sensitive for subthreshold GWAS signals leveraged in PWAS, we considered suggestive COLOC posterior probabilities (PP4>0.4) as supportive. Out of 37 proteins, 33 were supported by at least one method, including 30 by SMR (P_FDR_<0.05 and P_HEIDI_>0.05), 23 by COLOC, and 20 by both (**Fig.2D**, COLOC **Fig.S13**, SMR **Table-S21**, locus plots **Fig.S14**). Annotations of the proteins’ lead pQTLs–and those in close linkage disequilibrium (LD)–further supported the genetic signals were likely to impact protein levels, marking many regulatory region or 5’/3’ untranslated region variants, while 4 proteins also had missense variants including 3 strongly predicted to be deleterious (GBA1, RTFN1, ARSA; **Table-S22; Fig.S15**).

Regarding the impact of sex-stratification of the proteogenomics data, 14/37 proteins were more significant in primary (sex-matched proteogenomics) than secondary (non-sex-stratified proteogenomics) PWAS discoveries (**Fig.S16**), with 6 reaching P_FDR_<0.05 only in primary discoveries (**Fig.2E**), indicating added value for sex-matching GWAS and proteogenomics data. However, 23/37 proteins were more significant in secondary discoveries, with 13 reaching P_FDR_<0.05 only in secondary discoveries. Primary compared to secondary discoveries also tended to confer only minor increases in significance (**Fig.S16**), with the exceptions of INPP5D (immune signaling^44^), PSEN1 (amyloid pathology), and NCK1 (autophagy), a paralog of the known AD gene *NCK2*^17^. Leveraging both sex-stratified and non-sex-stratified proteogenomics data was thus important to comprehensively identify sex-biased AD proteins, yet PWAS discoveries mostly captured sex-biased AD genetic signals for which the causally related proteins were without notable sex-bias in the genetic regulation of their abundance (**Fig.2F**).

### Prioritization of sex-biased causal genes

Many GWAS loci did not have proteins mapped through PWAS, but other gene regulatory mechanisms may explain their sex-biased associations with AD. Even for loci uniquely identified through PWAS, gene prioritization remains incomplete since co-regulation–a regulatory genetic feature affecting multiple nearby genes^45,46^–is common and proteomics methods cover fewer genes and regulatory features than other omics approaches. We therefore performed COLOC analyses for all identified sex-biased AD loci with extensive multi-tissue, multi-QTL (i.e. xQTL) datasets to comprehensively prioritize likely causal genes and epigenetic features (with strong colocalization support PP4>0.7; **Tables-S23-25**). For prioritized proteins, we also performed sex-biased differential abundance level analyses. Findings were finally aggregated into gene priority scores (1-high, 2-medium, 3-none; cf. Methods).

Top gene prioritizations for novel sex-biased loci are presented in **Fig.3** (comprehensive results for all loci are in **Figs.S17-18**). Genes were prioritized at 44/46 unique loci (96%) and epigenetic features at 34/46 (74%), corroborating AD-associated loci broadly harbored regulatory signals. Additionally, 27/49 (55%) prioritized proteins showed evidence of sex-biased differential abundance, reinforcing their relevance to AD sex-dimorphism (**Table-S26**). Example GWAS-QTL colocalization plots are in **Fig.S19**, including the novel female-biased *SHC3* locus where *SEMA4D* was prioritized, a glial activation gene already targeted by Pepinemab in phase1/2 AD clinical trials^47^. Example PWAS-QTL colocalization plots are in **Fig.S20**, including female-biased genes *ARSA* (sphingolipid metabolism, innate immunity) and *ETFA* (mitochondrial metabolism). Both illustrate PWAS-identified proteins that were supported by xQTL COLOC and sex-biased differential abundance, conferring priority score-1. Gene prioritization also supported PWAS-identified genes such as female-biased *RAPGEF2*–linked to amyloid-induced synaptic loss^48^–not validated by standard pQTL SMR and COLOC but prioritized through monocyte and microglia QTL colocalizations. It also resolved scenarios such as *RASA4B*, where more robust evidence was instead found for *SH2B2* and *RELN* (**Fig.S21**). Notably, RELN binds APOE receptors and has been linked to synaptic protection and tau phosphorylation in AD^49,50^.

**Figure 3.**
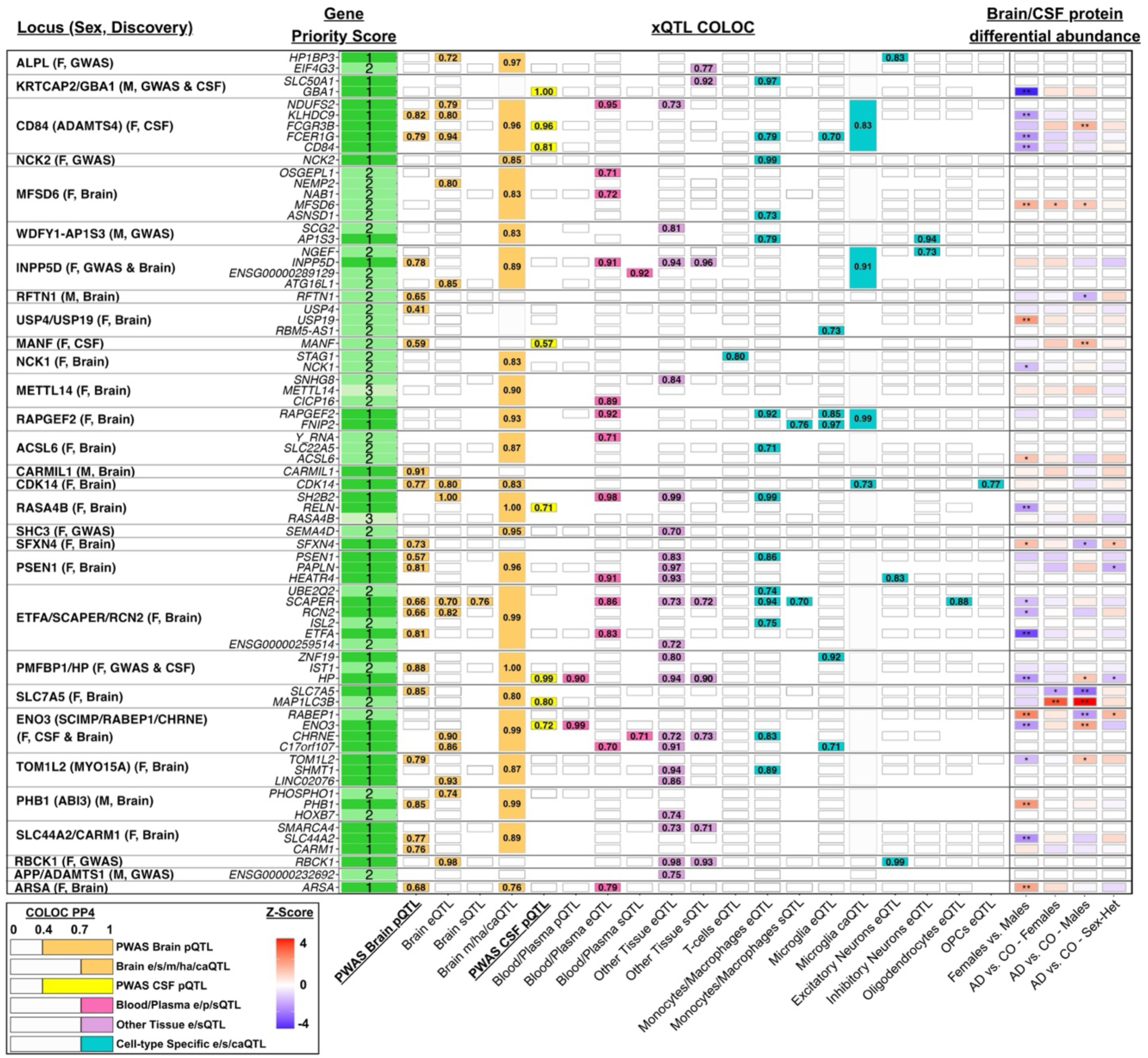
Gene prioritization at novel sex-biased loci through QTL colocalization and protein differential abundance analyses. Genetic COLOC analyses were performed with different QTL types (xQTL) across different tissues and cell-types, including gene expression (eQTL), transcript splicing (sQTL), protein abundance (pQTL), methylation (mQTL), chromatin accessibility (caQTL), and histone acetylation (haQTL). Displayed genes and tissues/cell-types reflect top gene prioritizations marked by priority scores: 1-high, 2-medium, 3-none (cf. Methods for details regarding priority scoring). Indicated values represent best COLOC PP4 findings out of multiple collapsed datasets (e.g. ‘Other Tissue’ shows the best result out of 35 investigated tissues in GTEx v8). PP4≥0.4 in PWAS Brain pQTL and CSF pQTL, and PP4≥0.7 in all other QTL datasets were annotated and contributed to gene prioritization. The heat matrix on the right indicates prioritized proteins that were further investigated for sex differential abundance or sex-biased case-control differential abundance (**, P_FDR_<0.05; *, P<0.05).

A specific strength of PWAS is its ability to integrate multiple regulatory signals to increase power for gene discovery^51^, but obtaining support from pQTL SMR and COLOC may then become challenging and confounded by complex LD structures^52^. To illustrate how xQTL COLOC analyses could resolve such instances, we created novel multi-tag locus plots and visualized the female-biased *PSEN1* finding (**Fig.4A**). Although pQTL COLOC was only suggestive for PSEN1, PP4=0.57, it became clear that 3 independent signals with overlapping LD composed its discovery and respectively showed strong colocalization with different gene QTLs across different tissues. Similarly, 2 independent signals contributed to the female-biased PWAS discovery of *CD84* (immune response regulator), but 2 more independent signals colocalized with other genes including *FCER1G* and *FCGR3B* (innate immunity), expanding gene prioritization and marking sex dimorphism at the larger locus (**Fig.4B**).

**Figure 4.**
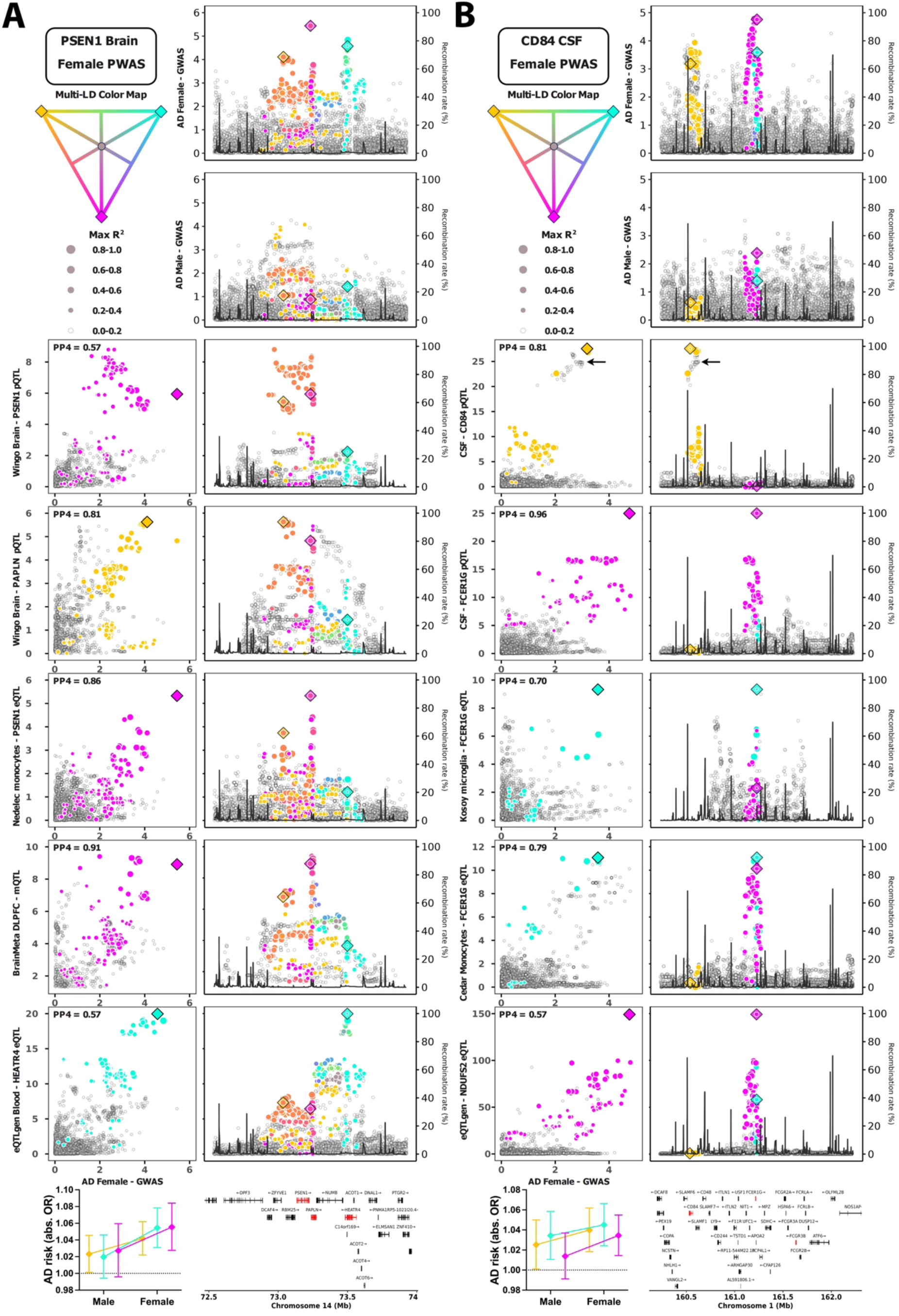
Showcases of sex-biased gene discoveries enabled through combined PWAS and multi-tissue QTL analyses. Multi-tag locus plots display genetic linkage disequilibrium (LD) with 3 tagging variants simultaneously: left sections indicate GWAS-QTL comparisons including COLOC PP4 values in the top left corner, while right sections show GWAS or QTL locus zoom plots. The bottom left corner panels indicate AD sex heterogeneity for the 3 tagging variants. **A)** *PSEN1* was identified as a female-biased risk gene through brain PWAS. The brain PSEN1 pQTL signal is shown on row 3, indicating suggestive colocalization with the female AD GWAS (PP4=0.57). Despite overlapping LD, the 3 tag variants appear to represent independent signals when evaluating QTL colocalization for additional genes at the locus in different tissues, as well as *PSEN1* eQTL data in macrophages (row 5). **B)** *CD84* was identified as a female-biased risk gene through CSF PWAS. The CSF CD84 pQTL signal is shown on row 3, indicating strong colocalization with the female AD GWAS (PP4=0.81). Two independent signals appeared to drive the AD PWAS finding: one signal is marked by the purple tag variant and the second is not directly color-marked (reflecting independent LD) but indicated by the black arrow. Additionally, 2 more independent signals were apparent (yellow and cyan tag variants), marking sex dimorphism at the larger locus and prioritizing additional risk genes through QTL colocalization.

To finalize gene prioritization, 4 genes at sex-biased loci were added given preceding literature strongly implicating them in AD (cf. Methods). We further removed two loci not appearing to be robustly sex-biased, including *HLA* (cf. GWAS results), and *ADAMTS1/APP*, which upon closer inspection displayed evidence of both male and female-biased signals (**Fig.S4.9**). In total, we prioritized 125 female-biased and 21 male-biased likely causal genes, with 62 assigned priority score-1 (**Table.S27**).

### Biological insights into Alzheimer’s disease sex-biases

We next sought to gain a better biological understanding of the newly expanded, sex-biased genetic landscape for AD (**Table.S27**). Pathway enrichment, using Gene Ontology (GO) terms, identified 149 female-biased and 11 male-biased pathways after filtering for significance (P_FDR_<0.05), which we restricted further to 94 female-biased and 10 male-biased pathways with increased sex-specificity (>1.5-fold enrichment ratio across sexes; cf. Methods**; Tables-S28-29**). These pathways formed 7 female-biased clusters, including amyloid, immune, and stress response, and 5 male-biased clusters (**Fig.5A; Figs.S22-23**). Next, using cell-type specific expression data from Zhang et al^43^, we defined cell-type gene sets for enrichment analysis. These revealed significant overrepresentation of female-biased AD genes in astrocytes (fold-enrichment=0.66, P=0.022), and microglia (fold-enrichment=0.73, P=0.015; **Fig.5B**, **Fig.S24**, **Table-S30**).

**Figure 5.**
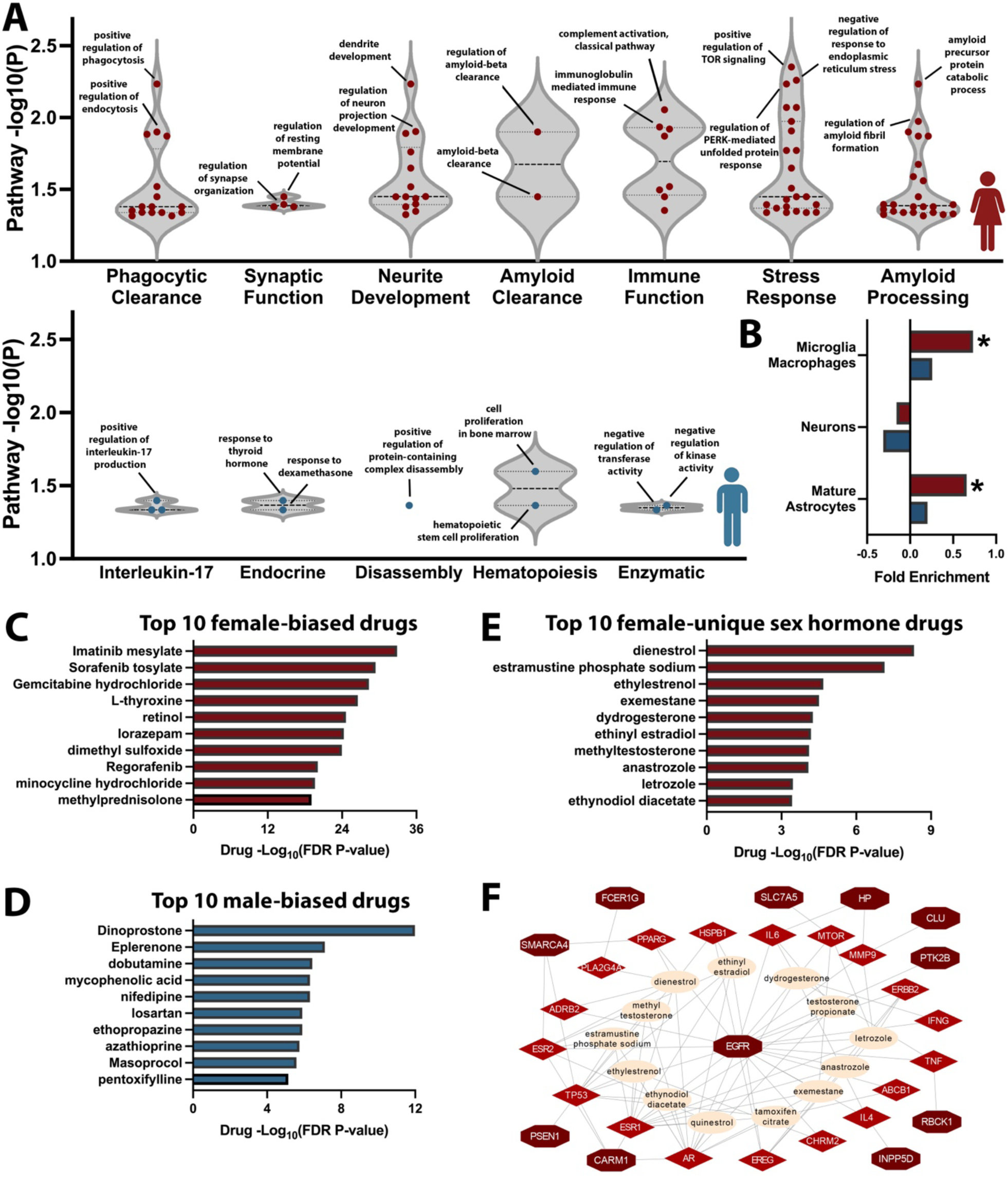
Sex-biased enrichment of biological pathways, cell-type specificity, and drugs. **A)** Sex-biased prioritized AD genes were used for sex-stratified pathway enrichment analyses, followed by retaining FDR significant pathways, filtering for sex specificity, and clustering. Violin plots display final pathway GO terms in their respective clusters (top, female; bottom, male). **B)** Sex-biased prioritized genes were evaluated for cell-type expression specificity, showing significant (*) enrichment for microglia and astrocyte specific genes in women (red bars). **C-F)** Drug enrichment analyses were conducted per sex and filtered for FDR significance and sex specificity. Bar graphs display top 10 enriched drugs based on top prioritized genes (score-1) in **C)** women and **D)** men. **E)** A subset of sex hormone-related drugs was observed solely for female-prioritized genes. **F)** The genes and drug network underlying findings in (E) are displayed, including prioritized female-biased AD genes (dark red), their connected, druggable AD-related genes (bright red), and their connections with sex hormone-related drugs (beige), centered on *Endothelial growth factor receptor* (*EGFR*).

Drug enrichment, focusing on genes with priority score-1 and their connected AD-relevant, druggable genes identified 617 female-biased and 244 male-biased FDA-approved drugs, of which 369 female-biased and 116 male-biased drugs displayed increased sex-specificity (cf. Methods; **Fig.5C-D**, **Fig.S25**, **Tables-S31-32**). We additionally identified a subset of sex hormone-related drugs forming a network with 11 female-prioritized genes converging on *Epidermal Growth Factor Receptor* (*EGFR*, **Fig.5E–F**). The known GWAS top variant at the *SEC61G/EGFR* locus^17^ showed a stronger (P_Sex-Het_=0.029) protective effect in women (OR=0.91, P=1.6e-9) then in men (OR=0.96, P=0.013; **Table1**), with brain and CSF PWAS also indicating strong sex-biases for EGFR (**Table-S33**). Notably, the top female-biased AD signal colocalized with *EGFR* eQTLs in astrocytes, while a second, novel female-specific signal colocalized with *EGFR* eQTLs in oligodendrocyte precursor cells **(Fig.S26**), corroborating sex dimorphism at the locus.

### Haptoglobin

*Haptoglobin* (*HP*) was prioritized as a novel, female-specific AD gene by GWAS (at the *PMFBP1* locus; **Fig.1B**), CSF PWAS (**Fig.2B-C**), and pQTL colocalization in CSF and plasma (**Fig.3**). A common intragenic duplication on *HP* defines two haplotypes, HP1 and HP2, with further sub-haplotypes including the exon 4/6 missense variant F/S^39^. HP1/HP2 are generally invisible to SNP arrays and short-read whole-genome sequencing (WGS) but impact HP quaternary structure and APOE oxidation^39^. To elucidate the relationship between the female-specific genetic signal, HP structural variants, and AD, we leveraged novel long-read sequencing (LRS) data from the Stanford ADRC/SAMS cohorts to assess LD and perform HP1 genotype imputation in Stage-1-2 GWAS and CSF/plasma pQTL data (**Fig.6A**; **Figs.S27-28**, **Table-S34**; cf. Methods)^53^.

**Figure 6.**
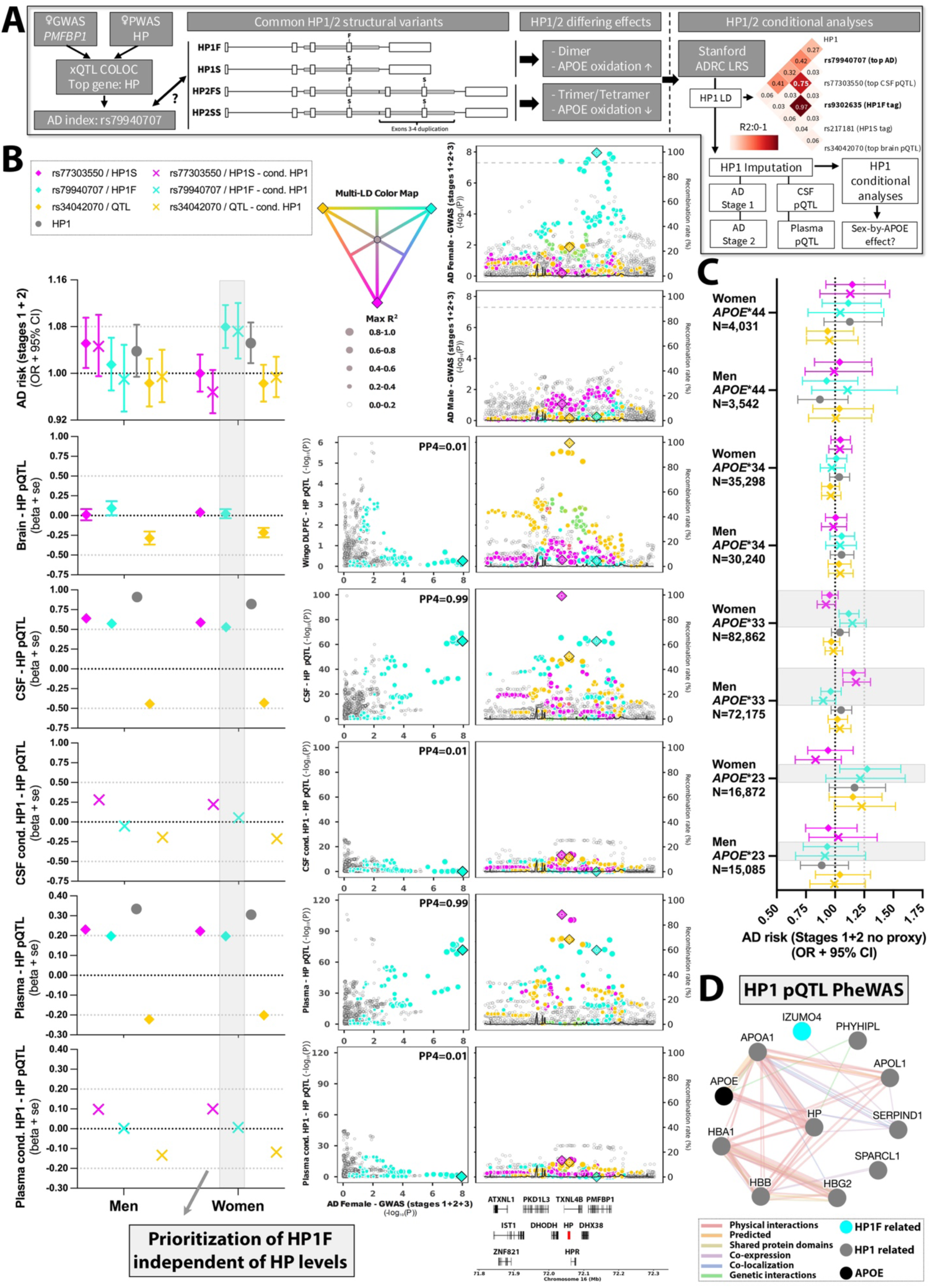
*Haptoglobin* (*HP*) is a female-specific Alzheimer’s disease causal gene that interacts with *APOE* biology. **A)** Schematic of follow-up analyses evaluating the role of structural variants HP1/HP2 and their sub-haplotypes at the novel female-specific *HP* locus. Heatmap on the right shows LD for 3 variants of interest (selected following multi-tissue QTL investigation), HP1, and known tag variants for HP1F and HP1S. Note strong LD for rs79940707 and HP1F, and rs77303550 and HP1S, while rs34042070 was mostly independent. **B)** Left panels display AD risk and pQTL associations for all 3 variants, HP1 genotype, and the 3 variants conditioned on HP1 genotype. The right panels provide matching multi-tag locus plots (pQTL associations are non-sex-stratified; HP1 genotype not depicted). PP4 values are marked to indicate colocalization with the rs79940707/HP1F signal (cyan). The gray bar highlights rs79940707/HP1F remains associated with AD risk in women after HP1 genotype conditioning, but CSF and plasma pQTL associations disappear. **C)** Gray bars highlight the rs79940707/HP1F AD association appears more pronounced in female *APOE* ε33 and *APOE* ε22 carriers (but not *APOE* ε4 carriers). **D)** Protein-protein interaction network shows significantly association proteins from CSF and plasma pQTL PheWAS analyses for all 3 variants and HP1 genotype. APOE was not directly identified in the *trans* pQTL analyses but added given known interactions with HP.

Following GWAS and xQTL investigation, we focused on 3 variants: rs79940707–the female AD index variant–showing strong LD (in EUR) with the known HP1F tag variant (R2=0.75) and some LD with HP1 (R2=0.27), rs77303550–the top CSF HP pQTL–showing strong LD with the known tag variant for HP1S (R2=0.97) and some LD with HP1 (R2=0.41), and rs34042070–the top brain pQTL and multi-tissue eQTL largely LD-independent of HP structural variants (**Fig.6A-B**, **Fig.S29**, **Table-S35**). The association of rs79940707/HP1F with female AD risk was not affected by conditioning on HP1 genotype nor use of proxy-AD GWAS and was stronger than the association of HP1 genotype (**Fig.6B**, **Table-1**, **Table-S36**, **Fig.S30**). However, in CSF and plasma pQTL analyses, HP1 genotype was most strongly associated with HP levels at the locus, while after conditioning on HP1 genotype, rs79940707/HP1F lost its association (**Fig.6B**). Rs79940707/HP1F was never associated in brain HP pQTL analyses which used tandem mass tag mass proteomics (**Fig.6B**)^24^. In contrast, CSF and plasma pQTL analyses used SomaScan aptamer technology for which HP protein measurements may be affected by HP1/HP2-related changes in quaternary structure^27,54^. We concluded HP1F is the likely variation causal to AD but does not impact HP (protein) levels.

There are currently no known functional effects for HP1F versus HP1S, but they are marked by biochemical differences (differential run speed on a protein gel)^39^. Seeking to understand how rs79940707/HP1F may affect AD risk, and in light of the established HP-APOE link^39^, we conducted sex-by-*APOE* stratified analyses which showed the association of rs79940707/HP1F with AD risk was more pronounced in female *APOE* ε33 and ε*23 carriers (OR_APOE*33_=1.12, P_APOE*33_=0.005; OR_APOE*23_=1.28, P_APOE*23_=0.019;* **Fig.6C**, **Table-S37**), corroborating an interaction with *APOE* biology. To further expand insights, we performed CSF and plasma *trans* pQTL phenome-wide association studies (pheWAS) at the *HP* locus, identifying a network of significantly regulated proteins converging on oxidative stress, *APOE*, and hemoglobin pathways (**Fig.6D**, **Figs.S31-33**, **Table-S38**; cf. Methods). Out of 10 associated proteins in the network, 9 were identified through HP1 associations and 1, IZUMO4, through rs79940707/HP1F after conditioning on HP1 genotype. Notably, IZUMO4 may be relevant to TREM2 biology (cf. Discussion).

## Discussion

We performed the largest-to-date sex-stratified GWAS of AD and integrated results with brain and CSF proteogenomics data through PWAS, identifying 46 sex-biased genetic loci and 37 sex-biased proteins. By further integrating GWAS and PWAS with xQTL colocalization we expanded gene prioritization to 125 female-biased and 21 male-biased likely causal genes (**Table-S27**). Enrichment analyses linked these genes to sex-biased AD pathways, cell-types, and repurposable drugs. We finally highlighted *EGFR* and *HP* as important mediators and potential drug targets for female-biased AD risk.

Our sex-stratified AD GWAS substantially extends prior large-scale non-stratified efforts that identified over 70 loci^16,17^, showing many known loci displayed significant sex heterogeneity and further revealing entirely novel sex-biased signals. Two recent X chromosome wide association studies (XWAS) of AD identified 5 female-biased (4 escaping X inactivation) but no male-biased loci^34,55^, complementing our observation that autosomal effects were often female-biased (70%). Acknowledging that proxy-AD GWAS can confer biases to subthreshold polygenic signals^40,56,57^, we performed the sex-stratified proxy-AD GWAS in Stage-2 (UKB) following recommendations from Wu et al to reduce bias^40^. We focused on genome-wide significant loci and observed strong concordance of index variant effect sizes across GWAS Stages, suggesting there were no biases in our discoveries. While PWAS did use subthreshold GWAS signals, 34/37 sex-biased proteins remained consistently associated when excluding proxy-GWAS. Given AD’s oligogenic architecture^58,59^, these PWAS loci thus likely represent true subthreshold, causal genetic signals. Further, biobank phenotypes can reduce AD specificity^57^. While we indeed observed an AD male-biased effect at the *GBA* locus for PD^37^, we identified this association was not specifically driven by biobanks but rather the dependence on non-pathology confirmed AD phenotypes also in clinically diagnosed cohorts. Our prioritization of amyloid and other core AD pathways however suggests we mostly identified AD-relevant genes. Notably, the male-biased AD signal at *GBA* replicates in a recent sex-stratified LBD–but not PD– GWAS (**Fig.S7**)^60,61^, corroborating true sex dimorphism at the *GBA* locus.

We replicated many previous PWAS-prioritized proteins prior to sex-specificity filtering^27,62^, but additionally identified many novel sex-biased proteins. Prior studies focused on sex-biased QTLs as a source of sex differences in human traits, but only a minority of QTLs displayed sex-biases^24,29,30^. We found that by additionally integrating our sex-stratified AD GWAS with non-stratified proteogenomics data, we increased protein discoveries by 54%. We also determined most sex-biased AD genetic loci did not display sex-biased genetic regulation of their causally implicated proteins, but 55% of those proteins did display abundance differences across men and women. Another important extension on prior PWAS was the recognition that proteogenomics data has limitations in gene coverage and misses other important types of gene regulatory information. By implementing xQTL COLOC, we were able to further prioritize implicated proteins or identify other likely causal genes at most loci. Our extensions of the PWAS framework were thus crucial to comprehensively map sex-biased AD genes.

The large amount of female versus male-biased prioritized genes–only partly driven by increased power in females–is in line with the propensity for female-biased rather than male-biased molecular changes observed in sex-aware AD omics studies^23^. Pathway analyses showed female-biased genes converged on immune function, amyloid, and responsive mechanisms, including phagocytosis, endoplasmic reticulum (ER) stress response, neurite development, and synaptic function. The neurite and synaptic pathways, which collectively would relate to neural compensation, are in line with mounting evidence for female-biased brain resilience to AD^10,11,14,63^. Phagocytosis has also been extensively reported to display sex differences and over two-thirds of autophagy proteins have androgen and estrogen receptor binding sites in their promoter regions^13,64^. Phagocytosis in turn may relate to the ER stress and unfolded protein response pathway, proposed to be important to the development of AD^65^. A female-bias for amyloid is perhaps surprising since most studies report female-biases for tau pathology^8,9,66–69^, but this may reflect that sex-dimorphic amyloid changes could happen early in the disease course and would be missed by most observational studies. Notably, using a large real-world sample of amyloid PET data (N=10,361), Raya *et al.* recently showed females have higher risk of amyloid pathology, which was most pronounced at the youngest investigated ages (65-69y)^70^. While we did not observe a female-biased tau pathway, the rs121918007 missense variant on *ALPL,* a.k.a. tissue-nonspecific Alkaline Phosphatase (*TNAP*), causes hypophosphatasia, a metabolic disorder affecting bone mineralization. *TNAP*’s function has also been linked to tau phosphorylation^71^, potentially explaining rs121918007’s female-biased association given amyloid-modulated tau phosphorylation appears to accelerate tau tangle accumulation in females^72^. Finally, the immune pathway likely has the most reported sex-dimorphic support in AD, including both female and male-biased observations^13,73^. It is interesting then that we observed a female immune cluster containing pathways such as complement activation and immunoglobulin-mediated response, while males uniquely displayed enrichment of *Interleukin-17* (*IL-17*), a proinflammatory cytokine linked to inflammation and vascular dysfunction in a mouse model of AD^74^. Extending immune-relevant findings, we observed enrichment of female-biased genes in microglia and astrocytes, consistent with converging evidence that estrogen exerts strong effects on glial function and microglial inflammation disproportionately impacts AD pathogenesis in women^75,76^. Further, we were able to prioritize sex-biased potentially repurposable drugs. The top female-biased drug, Imatinib mesylate, is an anti-cancer drug that already shows some promise for repurposing in AD given its link to amyloidogenesis^77,78^. The top male-biased drug, Dinoprostone (P_FDR_=1.0e-12), or Prostaglandin E2 (PGE2), may relate to AD through neuroinflammatory pathways^79,80^. While primarily used to induce labor, PGE2 interacts with sex hormones and has been shown to influence brain masculinization^81^, indicating Dinoprostone is unlikely to be repurposed for AD in men but instead helps corroborate the prioritized genes interact with male sex hormone biology. We also observed a network of female-biased genes enriched on estrogenic, progestogenic, androgenic, and aromatase inhibitor drugs, corroborating these genes interact with sex hormone biology in women, yet the use of estrogen therapies for AD in women remains uncertain^2,15^. Similar to men–although at much lower levels–women display decreasing testosterone levels across aging which may relate to poorer cognitive function^82,83^, suggesting that targeting testosterone may hold therapeutic relevance in women. At the center of the sex-hormone drugs, we found *EGFR*, known to be transactivated by estrogen^84^, linked to AD in a 2022 GWAS^17^, increasingly implicated in AD pathogenesis^85^, and recently brought forward as a dual molecular target for cancer and AD^86^. Altogether, *EGFR* appears as an important target to be explored for AD with potential increased benefit in women.

The observation of *HP* as a female-unique AD causal gene was particularly interesting given its role in APOE oxidation and the female-biased effect of *APOE* ε4 in AD^5–7^. The female index variant rs79940707, which we showed to be linked to the likely causal structural variant HP1F, showed a stronger effect in *APOE* ε23 and ε33 female subjects, suggesting female-biased interactions with *APOE* biology in AD extends beyond *APOE* ε4. A recent study showed a trending protective effect of HP1 in *APOE* ε23 carriers, but no effect in ε33 or ε34 carriers, and a causal effect in ε44 carriers^87^. However, sample sizes were considerably smaller, and analyses not stratified by sex nor focused on HP1F, likely explaining these differences. Beyond APOE biology, we prioritized a link for *HP* to lipid metabolism and oxidative stress, which have been implicated in AD sex differences^13^, and hemoglobin, of which women have lower mean levels in blood than men^88^. We specifically tied rs79940707/HP1F to *IZUMO4*, which is strongly expressed by testes and ovaries (GTEx v10). Notably, Wang et al showed the AD *MS4A* risk locus affects both TREM2 and IZUMO4 protein levels in CSF^89^. These observations provide initial insights into the potential causal variation and mechanisms linking *HP* to female-specific risk for AD, warranting further experimental validation studies.

There are some limitations to consider when interpreting our findings. While our study was designed to identify genes that are robustly sex-biased, and power differences across sexes only partly accounted for more female-biased findings, our male-stratified analyses nonetheless had less power inherently limiting potential male-biased discoveries. Additionally, although our GWAS incorporated nearly 1,000,000 individuals, integrating more datasets, particularly clinically and pathology confirmed case-control cohorts, will be important to increase power. Future extended sex-stratified GWAS will thus help expand insights into male-biased AD risk mechanisms and refine downstream gene, pathway, and drug prioritizations in both sexes. Further, our main GWAS was focused on EUR subjects. While we did explore consistency of sex-biased loci in a smaller AFR sex-stratified GWAS, those sample sizes were too small to robustly assess cross-population generalizability. Similarly, our AFR PWAS had limited power and LD mismatches between AFR GWAS and EUR proteogenomics data rendered these analyses mostly explorative. The addition of novel multi-ancestry AD GWAS and proteogenomics resources in future studies will thus be relevant to help fine-map causal variants and genes as well as validate AD causal proteins. Further, our gene prioritization analyses did not focus on causal variant fine-mapping but rather on identifying the most likely causal genes for all AD genetic signals at prioritized loci. Although not all prioritized genes will be causal, our findings offer a foundation of the most promising sex-biased targets for further gene prioritization and validation studies.

In conclusion, we present the most comprehensive genetic roadmap of sex-biased potential causal genes for AD. Our prioritized genes, pathways, and repurposed drugs shed light on sex-biased AD pathobiology and steer future computational and experimental validation studies to identify actionable sex-biased drug targets. Altogether, our findings emphasize the crucial importance of sex-aware research and pave the way towards sex-specific precision medicine in AD.

## Online Methods

### Ethics declaration

Participants or their caregivers provided written informed consent in the original studies. The current study protocol was granted an exemption by the Washinton University Institutional Review Board because the analyses were carried out on “de-identified, off-the-shelf” data; therefore, additional informed consent was not required. The FinnGen ethics statement is available in the supplement (Nr HUS/990/2017).

### GWAS samples, phenotypes, and quality control

An overview of the GWAS study design and samples is depicted in **Fig.1A**. In-depth methodology regarding cohort-specific descriptions, genetic QC, imputation, and phenotype definitions are provided in the **Supplementary Methods**, **Fig.S1A**, **Tables-S1-4**, and **Tables-S7-8**^33,36,90–92^. The main sex-stratified AD GWAS meta-analyses were conducted in non-Hispanic White European (EUR) ancestry individuals (>75% EUR^93^) using a 3-stage design. For stage-1, case-control samples with clinical diagnoses (as well as pathology confirmed diagnoses for a subset of individuals; **Table-S3**) were obtained for SNP-array AD GWAS datasets, primarily composed of the Alzheimer’s Disease Genetics Consortium (ADGC; **Table-S1**), along with whole-genome sequencing (WGS) data from the Alzheimer’s Disease Sequencing Project (ADSP R3; **Table-S2**). Stage-1 datasets underwent extensive genetic QC and SNP-arrays were imputed to the Trans-Omics for Precision Medicine (TOPMed) reference panel^94^. For stage 2, the UK Biobank (UKB) provided case-control samples derived primarily through an AD family-proxy phenotype^22,95^, as well as a smaller subset of registry cases determined through ICD codes and health registry data. The proxy phenotype was adapted to support sex-specific analyses (e.g., female-specific analyses leveraged maternal AD status while excluding subjects that reported a father with AD). Considering potential biases when using the AD proxy phenotype, this approach was further tailored according to recent recommendations to reduce bias in AD proxy GWAS (using only parental phenotypes with parental ages ≥65 years)^40^. Imputed genotype data for UKB were available from the combined 1000 Genomes, HRC, and UK10K reference panels^33^. For stage-3, AD health-registry phenotypes were leveraged from the FinnGen (R12) biobank with SNP-array data imputed to the SISu (v.4.2) reference panel^90^. Lastly, admixed African ancestry (> 25% AFR) sex-stratified GWAS were conducted in non-Hispanic and Hispanic case-control samples from the ADGC and ADSP (**Tables-S7-8**). Across all analyses, females and males were respectively XX or XY with concordant self-reported sex.

### Sex-stratified AD GWAS

Across all samples, GWAS were performed in respective sex strata using an additive model, variants with an imputation score > 0.3, and binary case-control status at the outcome measure, while adjusting for age, technical covariates, *APOE* ε4 and *APOE* ε2 dosages (Stages-1-2), and genetic principal components (capturing population stratification), as applicable per dataset (cf. Supplemental methods). The *APOE* region (±5 Mb) was excluded from analyses. In EUR GWAS, mixed models were leveraged to include related subjects (BOLT-LMM^96^ v.2.4 in stage-1 and stage-2; Regenie^97^ in stage-3), while in AFR GWAS, subjects were unrelated down to 2^nd^ degree relatedness (using PLINK v.2.0^98^ for associating testing). Association beta coefficients and standard errors from BOLT-LMM^96^ were rescaled to a standard logistic scale.

In Stage-1, GWAS were performed respectively in ADGC and ADSP. In ADGC, variants were filtered to a minor allele count (MAC) > 20. In ADSP, variants were included with a MAC > 1 but restricted to those evaluated in ADGC. In stage-2 and stage-3, variants were filtered to a minor allele frequency (MAF) > 0.05%. In the AFR samples, GWAS were performed respectively in ADGC and ADSP, which were further subdivided into Hispanic and non-Hispanic groups (4 total analyses). Given the substantially smaller samples sizes in AFR GWAS, variants were filtered to a MAF > 1%.

GWAS meta-analyses per sex were conducted using GWAMA^99^. Fixed effects meta-analyses were used for EUR cohorts, while random effects meta-analyses were used for admixed AFR cohorts. After meta-analyses, all variants were restricted to those observed in ADGC to ensure effect estimates could be evaluated with regard to “clean” AD case-control samples and that associations were not driven solely by stage-2 and stage-3 biobank analyses. Variants were further restricted to those having 90% genotyping rates across all meta-analyzed cohorts, as well as those not displaying frequency deviations > 20% across non-Finnish European cohorts. Effective sample sizes were calculated using the formulation N_Eff_ = 4*v*(1-v)^100^, where v = N_Cases_/(N_Cases_+N_Controls_) for which sample contributions from stage-2 were divided by 4 to account for reduced power when using proxy GWAS^100^. Lastly, GWAMA^99^ was used to perform sex-heterogeneity GWAS to evaluate if sex-stratified associations displayed evidence of significant sex-biases.

### GWAS locus sex-bias and novelty

Independent GWAS loci, per sex, were determined through a 1 Mb sliding window analysis for all variants with *P* < 1×10^-5^. Loci were retained if they reached genome-wide significance (*P* < 5×10^-8^) in a respective sex and showed nominal sex-heterogeneity (*P*_Sex-Het_ < 0.05). The resultant sex-biased variants were further split into independent signals through clumping (using LD data from the ADGC cohort, considering independent signals at R^2^ < 0.01). To ensure these signals were robustly sex-biased, we removed signals where the lead variant was in LD with a previously reported, common AD variant (R^2^ > 0.01, MAF>1%) that was more significant in the same sex but not sex-heterogeneous (*P*_Sex-Het_ > 0.05); this excluded 4 loci. Further, if loci contained multiple independent sex-biased signals, we retained only the most significant one at the locus; this excluded secondary signals in 2 loci (cf. **Fig.S4.10-11**). All lead variants obtained through these filters displayed pronounced sex differences in effect magnitude (cross-sex effect-size ratios ≥1.5); we concluded there was no need to implement multiple-comparison corrections to *P*_Sex-Het_.

Variant and locus novelty was determined by evaluating both proximity and LD to previously reported lead variants from various AD GWAS^17,35,59,101–109^. Loci > 1 Mb away from prior lead variants were considered “novel locus & novel variant”, those < 1 Mb away but with R^2^ < 0.01 were considered “known locus & novel variant”, and those < 1 Mb away with R^2^ > 0.01 were considered “known locus & known variant” (with the exception of 2 variants for which R^2^ > 0.01 was observed but follow-up analyses supported these variants to represent novel independent signals; **Table-S5**).

### GWAS sex-heterogeneity consistency across ancestries

For all sex-biased lead variants, sex heterogeneity was evaluated in a meta-analysis of EUR and admixed AFR sex-specific association results. Given the significantly lower power in the AFR samples, these analyses did not seek to evaluate replication, instead, consistent sex heterogeneity was determined if *P*_Sex-Het_ improved when combining EUR and AFR findings (**Table-S9**).

### Brain proteogenomic samples and analyses

Human brain proteomes were previously generated from the dorsolateral prefrontal cortex (DLPFC) of donated brain samples from EUR participants in the Religious Orders Study (ROS) and Memory Aging Project (MAP) cohorts^110^. Details on cohort information, genomic data QC, sample preparation, and proteomic sequencing, and data processing have been described in previous studies^24,25,62,111,112^. Briefly, proteomic profiling utilized isobaric tandem mass tag (TMT) peptide labelling coupled with liquid chromatography-mass spectrometry (TMT-MS). After QC, 8,458 proteins from 716 proteomic profiles were available for *cis* protein quantitative trait loci (pQTLs) mapping within ± 500 kb of the gene’s boundaries (PLINK v.2.0^98^), under sex-stratified and non-sex-stratified conditions. Covariates included 56 significant surrogate variables derived from surrogate variable analysis (SVA) using the SVA package (v.3.20.0)^113^ while protecting the effect of sex on protein abundance. Non-sex-stratified and sex-stratified brain *cis* proteogenomic weights (i.e., variant protein weights) were generated using the FUSION^51^ package, with sex-stratified weights from prior work^24^, and non-sex-stratified brain protein weights newly added here. Genotype data were restricted to SNPs in the 1,000 Genomes EUR LD reference panel provided with FUSION^51^. Proteins with significant SNP-based heritability (*P* < 0.01) were retained. Prediction models (best linear unbiased prediction [BLUP], Bayesian sparse linear mixed model [BSLMM], least absolute shrinkage and selection operator [LASSO] regression, and elastic net [enet] regression) were evaluated, and variant protein weights were retained from the best performing model.

### CSF proteogenomic samples and analyses

Cerebrospinal fluid (CSF) proteomes were previously generated from 4,989 samples (4,968 participants) with matching genome-wide data from eight cohorts: Alzheimer’s Disease Neuroimaging Initiative (ADNI), Dominantly Inherited Alzheimer’s Network (DIAN), Charles F. and Joanne Knight Alzheimer’s Disease Research Center (Knight-ADRC) Memory and Aging Project (October 2021 and June 2023), Fundació ACE Alzheimer Center Barcelona (FACE), Longitudinal observational study from the Memory and Disorder unit at the University Hospital Mutua de Terrassa (Barcelona-1), Washington University Movement Disorder Clinic (MARS), Parkinson’s Progression Markers Initiative (PPMI), and Stanford Iqbal Farrukh and Asad Jamal ADRC and Aging Memory Study (Stanford)^27^. Protein levels were measured using the SOMAscan 5K platform (PPMI, Stanford) or 7K platform (ADNI, DIAN, Knight-ADRC, MARS, FACE, Barcelona-1). Cohort details, proteomic/genomic data QC, and processing have been described previously^27^. Post-QC, 3,506 individuals were included in non-sex-stratified and sex-stratified analyses. Analyses were restricted to 7,008 aptamers from the largest dataset (ADNI, DIAN, Knight-ADRC [October 2021], FACE, Barcelona-1), mapped to 6,361 unique UniProt IDs through information provided by SomaLogic. *Cis* pQTL analyses were performed within ± 1 Mb of each protein’s transcription start site (PLINK v.2.0^98^) for non-sex-stratified and sex-stratified study designs, adjusting for age, first 10 genetic PCs, and cohort/array. For each set of *cis* variant-aptamer analyses, if the lead variant passed genome-wide significance (*P* < 5×10^-8^) then the window around this lead variant was used to calculate variant aptamer/protein weights using FUSION^51^. If an aptamer reached this significance threshold only in one sex, weights were also generated for the opposite sex. As for brain, variant protein weights were retained from the best performing model.

### Sex-stratified AD PWAS

Proteome-wide association studies (PWAS) were conducted using FUSION^51^, integrating sex-stratified AD GWAS summary statistics with sex-stratified or non-sex-stratified proteogenomic weights from brain and CSF (non-sex-stratified weights were obtained by combining males and females into a single analysis). GWAS statistics were processed using the LD Score (LDSC)^114^ software ‘mungestats’ utility, filtering variants to MAF > 0.1%. For brain PWAS, LD reference panels were generated from the 1000 genomes WGS resource^115^, using EUR samples when integrating the EUR GWAS, and using EUR, AFR, and Amerindian (AMR) samples when integrating the AFR GWAS (**Fig.S1B** for AFR GWAS admixture plot). For CSF PWAS, the genotype data from the CSF proteogenomic samples (EUR ancestry) were used for the LD reference panel. The major histocompatibility complex (MHC) region was excluded from all PWAS analyses. Significant PWAS associations for every combination of GWAS and proteogenomic data were defined at *P*_FDR_ < 0.05.

### PWAS locus sex-bias and novelty

Proteins significantly associated with AD in sex-stratified PWAS were filtered for sex-specificity (schematic in **Fig.2A**). Female biased proteins were identified by integrating female-stratified AD GWAS data with female-stratified (primary discovery) or non-sex-stratified (secondary discovery) variant protein weights, with parallel analyses for male biased proteins. Proteins were considered sex biased if their associations in the opposite sex’s PWAS (using female, male, or non-sex-stratified weights) were discordant in effect direction or did not reach nominal significance (*P* > 0.05). Additionally, if there were loci with multiple significantly associated proteins, and the most significant protein was not sex biased, all proteins at the locus were considered to not be sex biased. Proteins were classified as “novel locus & novel gene” when >1Mb away from previously identified AD lead variants, “known locus & known gene” if they fell within 1Mb and had previously been functionally prioritized in post-GWAS analyses, or “known locus & novel gene” if without clear prior functional prioritization.

### PWAS sex-heterogeneity consistency across ancestries

Sex biased proteins identified in EUR-ancestry PWAS were evaluated in admixed AFR sex-stratified FUSION anlayses^51^. Proteins were considered consistent if the *P*-value in the discovery sex (e.g., female) improved in meta-analysis of EUR and AFR results, while the opposite sex (e.g., male) maintained *P* > 0.05. Meta-analyses were conducted through sample-size weighted combination of *Z*-scores (**Table-S19**).

### PWAS sensitivity analyses

Sex biased proteins identified in EUR-ancestry PWAS (*P*_FDR_ < 0.05) were also evaluated in targeted FUSION^51^ analyses using a sex-stratified EUR GWAS excluding phase-2 proxy-phenotype data to assess potential biases^40^. Proteins were considered consistent in sensitivity analyses if the discovery sex (e.g., female) retained concordant effect direction and *P* < 0.05, while and the opposite sex-maintained *P* > 0.05 (**Table-S20**).

### PWAS gene validation - Brain and CSF pQTL SMR & HEIDI

Consistent with PWAS follow-up approaches in prior studies, we sought to validate sex-biased AD PWAS findings through summary-based mendelian randomization (SMR)^43^, testing causal links between *cis* regulated protein abundance, i.e. *cis* pQTL data (matching brain/CSF PWAS data) and AD risk (GWAS results). The heterogeneity in dependent instruments (HEIDI) test was used to evaluate potential LD bias. Associations were validated if SMR was significant (*P*_FDR_ < 0.05) and HEIDI was non-significant (*P* > 0.05). Similar to the PWAS design, analyses compared sex-stratified AD GWAS with both sex-stratified and non-sex-stratified pQTLs. See **Supplementary Methods** for additional details.

### PWAS gene validation - Brain and CSF pQTL COLOC

Consistent with PWAS follow-up approaches in prior studies, we sought to validate sex biased AD PWAS findings by evaluating genetic colocalization (COLOC) between brain/CSF *cis* pQTL data (matching brain/CSF PWAS data) and the AD GWAS. Variants within ±1 Mb of PWAS genes’ median base pair position (matching windows of variant protein weights) were analyzed using ‘coloc.abf’ with default per-SNP priors, which assumes a single independent signal at the locus^41^, and ‘coloc.susie’, which incorporates SuSiE (Sum of Single Effects) regression to resolve loci with multiple independent signals^42^ (coloc R package v.4.2.1)^41,42^. To further increase sensitivity to detect colocalization, a second set of coloc.abf analyses was performed with adjusted per-SNP priors, assuming increased probabilities of a variant associated with both AD risk and protein abundance; This was merited given loci were prioritized by PWAS. The likelihood of a shared genetic variant affecting both AD risk and protein abundance – COLOC hypothesis 4 – was reported by the posterior probability PP4, ranging 0 to 1. Across COLOC analyses, the best PP4 value was used with values ≥ 0.4 marking suggestive colocalization and ≥ 0.7 marking strong colocalization. Similar to the PWAS design, analyses compared sex-stratified AD GWAS with both sex-stratified and non-sex-stratified pQTLs. See **Supplemental Methods** for additional details.

### Variant annotation

The Ensembl Variant Effect Predictor (VEP; release 115, assembly GRCh38)^116^, was used to annotate consequences of sex biased variants from GWAS and PWAS analyses. For GWAS, the top sex-biased variant per independent locus and additional sex-biased variants in (R² > 0.8) with *P* < 5×10⁻⁸ were annotated. For PWAS, top pQTL variants for sex biased proteins, and those in LD (R² > 0.8) were annotated, with annotations collapsed by locus/gene.

### Gene prioritization - xQTL COLOC

Extending beyond basic pQTL COLOC analyses to corroborate PWAS gene findings (cf. above), a broader approach was used to prioritize potential causal genes at both respective sex-biased AD GWAS and PWAS loci, by performing extensive COLOC analyses with *cis* genetic association signals for a wide range of molecular traits across diverse tissues and cell types. The molecular traits, collectively referred to as “xQTL”, included gene expression (eQTL), transcript splicing (sQTL), protein abundance (pQTL), methylation (mQTL), histone acetylation (haQTL), and chromatin accessibility (caQTL). COLOC analyses evaluated both coloc.abf and coloc.susie and were focused on neurologically relevant tissues such as brain, CSF, plasma/blood, brain cell-type specific data, and peripheral immune cells (monocytes, macrophages, and T-cells), as well as other tissues from across the body available through GTEx v8^117^ (**Table-S23** for comprehensive overview of datasets). Colocalization analyses were restricted to variants located within ±1 Mb of (1) the lead sex-specific associated variant for AD GWAS, or (2) the median base pair position for AD PWAS genes based on the variants considered for variant protein weight calculation. PP4 values ≥ 0.7 marked strong colocalization.

### Gene prioritization - Sex-stratified differential abundance analyses

Any proteins identified as sex-biased through PWAS or displaying PP4 ≥ 0.7 in brain or CSF pQTL COLOC analyses were further interrogated in the brain and CSF proteomics data for evidence of (1) sex differential abundance and (2) sex-biased differential abundance across cases and controls. Sex differential abundance analyses were conducted in cognitively normal, mild cognitive impairment, and dementia subject while adjusting for diagnosis (brain, N=793; CSF, N=2,158). In sex-biased differential abundance analyses, cases and controls were defined based on amyloid and tau positivity status (AT), with controls being A-T- and cases being A+T+, identified through CSF biomarkers for CSF analyses (N_A-T-_=915, N_A+T+_=851) and postmortem CERAD and Braak pathology staging for brain analyses (A-T-: CERAD none or sparse & Braak 0-2; A+T+: CERAD moderate or frequent & Braak 3-6; N_A-T-_=80, N_A+T+_=481). Analyses were adjusted for age and relevant technical covariates.

### Gene prioritization – Gene scoring

Following the xQTL COLOC and differential abundance analyses, a gene prioritization score was designed to rank candidate genes at sex-specific GWAS and PWAS loci.

For sex-specific GWAS loci, candidate genes were prioritized following:

- Score 1: Genes exhibiting more than one gene-specific QTL colocalization (eQTL, sQTL, pQTL) at PP4 ≥ 0.7 across different tissues or datasets, or genes also identified in PWAS with strong colocalization (PP4 ≥ 0.7) in the matching pQTL tissue/dataset.
- Score 2: Genes with only one strong gene-specific QTL colocalization (eQTL, sQTL, pQTL) at PP4 ≥ 0.7.
- Score 3: Genes without strong gene-specific QTL colocalization (PP4 < 0.7).

For sex-specific PWAS loci, candidate genes were prioritized following:

- Score 1: Genes discovered in brain or CSF PWAS with strong colocalization (PP4 ≥ 0.7) for the same gene in the matching pQTL data. Any other gene within the same locus that colocalized at PP4 ≥ 0.7 in the non-matching pQTL tissue (e.g. CSF when locus was discovered through brain PWAS) and was unavailable in the matching pQTL tissue. Additionally, genes at the same locus that showed strong colocalization (PP4 ≥ 0.7) more than once in other gene-specific QTL colocalizations (eQTL, sQTL, pQTL). These extended prioritizations consider that (1) the PWAS prioritized gene may reflect a local coregulated non-causal gene, (2) the PWAS may not represent the causal tissue, (3) the proteomics data may not contain the causal gene, and (4) the identified locus may harbor multiple regulatory signals that cannot be understood through proteogenomics alone.
- Score 2: Genes discovered in brain or CSF PWAS with suggestive colocalization (0.4 ≤ PP4 < 0.7) for the same gene in the matching pQTL tissue; a score of 1, rather than 2, was attributed if the gene showed other gene-specific QTL colocalizations (eQTL, sQTL, pQTL) at PP4 ≥ 0.7. Additionally, genes at the same locus that showed strong colocalization (PP4 ≥ 0.7) in one other gene-specific QTL colocalization (eQTL, sQTL, pQTL). These extended prioritizations consider that subthreshold GWAS signals or multiple independent GWAS signals could lead to low PP4 values decreasing sensitivity to detect colocalization, and that multiple colocalizations at PP4 ≥ 0.7 are more promising than just one colocalization. Finally, genes with nominally significant sex-stratified differential abundance results (*P* < 0.05), but otherwise assigned score 3, were upgraded to score 2, to incorporate complementary gene prioritization support.
- Score 3: Genes that were not scored 1 or 2 reflect genes that were not prioritized and thus received a score of 3.

For 4 sex-biased AD GWAS loci that were already known AD loci, prior studies have implicated likely causal genes–*MAPT*, *HS3ST1*, *SORL1*, and *ABCA7*–but we did not observe xQTL COLOC or differential abundance support for them. For the novel female-biased *ALPL* locus, VEP annotation identified the lead variant to be a missense variant on *ALPL* with a high Combined Annotation Dependent Depletion (CADD) score (29.6). These genes were attributed a priority score of 1. Finally, while GWAS identified the *APP* locus to be male-biased, closer inspection indicated non-specific sex-heterogeneity at this locus (**Fig.S4.9**), leading us to exclude it from further gene prioritization and the final list of sex-biased genes.

### Multi-Tag Locus Plots

Some of the sex biased genetic loci appeared to present multiple genetic signals. To visualize LD with 3 tagging variants simultaneously, the largest LD (max R^2^) of each given variant in a locus with the 3 tagging variants was used to determine dot size, while a color gradient was used to visualize overlapping or distinct LD (R^2^ ≥ 0.2 only with one tagging variant); Variants displaying R^2^ < 0.2 with all 3 tagging variants were colored gray. LD info was derived from stage 1 ADGC genotype data.

### Sex biased pathway enrichment analyses

To investigate biological pathways associated with sex biased prioritized genes (female: N_genes_=125; male: N_genes_=21; **Table-S27**), gene symbols were converted to ENTREZ IDs using the ‘bitr’ function from the clusterProfiler R package (v.4.12.6)^118^. These IDs were analyzed separately for female and male genes using the ‘enrichGO’ function in clusterProfiler to identify enriched Gene Ontology (GO) terms. To ensure pathways were sex biased, GO terms were filtered to include only those with an adjusted p-value (*P*_FDR_) < 0.05 and a gene ratio at least 1.5-fold higher in one sex compared to the other. GO semantic similarity matrices were generated using the ‘GO_similarity’ function from the simplifyEnrichment R package (v.1.14.1)^119^. These matrices were subjected to hierarchical clustering with the ‘hclust’ function, resulting in 7 female-specific and 5 male-specific GO term clusters, which were then manually labeled (**Figs.S22-23** for filtering and clustering details).

### Sex biased cell type enrichment analyses

To determine the cell type specificity for sex-biased prioritized genes, we followed the approached described by Western et al. 2024^27^. Briefly, cell type-specific gene expression data were available from Zhang et al. 2016^120^ for human endothelial cells, oligodendrocytes, mature astrocytes, neurons, and combined microglia and macrophages^27,120^. Average expression levels across each cell type were calculated for each gene^27^. The averages were summed to yield the total expression level of each gene across all five cell types, and the proportion of this total expression attributed to each specific cell type was computed. A gene was considered cell type-specific if the expression level in the predominant cell type was at least 1.5 times higher than in the second most predominant cell type.

This information was then used to identify for female and male prioritized genes respectively the number of genes specific to each cell type. Enrichment was tested by applying the hypergeometric test (‘phyper’ R function) assessing whether the observed number of cell type-specific genes in each subset was greater than the background set of genes. Endothelial cells and oligodendrocytes had less than 10 genes annotated across both sexes, so related results were considered to be unreliable.

### Sex biased drug enrichment and repurposing analyses

To identify potential drug repurposing opportunities, we focused on genes with a priority score of 1 from sex-biased gene lists (female: N_genes_=55; male: N_genes_=7; **Table-S27**). Gene symbols were validated using the ‘checkGeneSymbols’ function from the HGNChelper R package (v.0.8.14)^121^. Per sex, these genes were expanded into networks of AD-related druggable genes. Specifically, for each prioritized gene, protein-protein interaction (PPI) data were queried via the EpiGraphDB API^122^ (/gene/druggability/ppi route) to identify interacting genes. These interacting genes were filtered down to those with AD relevance using literature evidence from EpiGraphDB (/gene/literature route, object: ‘Alzheimer’s disease’). These genes were then queried for overlap with a list of 4,479 druggable genes from Finan et al. 2017 ^123^, and filtered down to the subset of 1,427 most promising, Tier 1, druggable genes.

Drug repurposing enrichment analyses were performed on these expanded gene lists using the DSigDB database^124^ (DSigDB_All.gmt) with the ‘enricher’ function from the clusterProfiler R package (v.4.12.6)^118^. Candidate drugs were further filtered for FDA approval, *P*_FDR_ < 0.05, and sex-specificity (a gene ratio at least 1.5-fold higher in one sex compared to the other). Sex hormone-related drugs in the final female-specific list were reviewed and visualized in a protein-drug interaction network (**Fig.5E-F**). Additional details are provided in the **Supplementary Methods**.

### Haptoglobin – Proteogenomic analyses

Initially, *Haptoglobin* (*HP*) was prioritized through xQTL COLOC analyses of the *PMFBP1* AD GWAS locus but was not observed in AD PWAS. Upon review, it was determined HP was removed during QC in the CSF proteogenomic analyses. We conducted new proteogenomic analyses (consistent with methodology describe above) on a subset of samples where HP passed QC (N=3503), using non-sex-stratified and sex-stratified designs: Variants within a 2 Mb window of the *HP* transcription start site were analyzed to obtain *cis* pQTLs and variant protein weights (though FUSION^51^) for two aptamers targeting HP and the HP1 isoform.

### Haptoglobin – HP1 linkage analyses and imputation

To determine the relationship of *HP* structural variant HP1 (and HP2) with female AD index risk variant rs79940707 and other variants of interest at the locus, we leveraged joint long-read sequencing (LRS) and short-read sequencing (SRS) data from 431 individuals in the Stanford ADRC and Stanford Aging & Memory Study (SAMS) (**Fig.6A**). Briefly, HP1 genotype was obtained through LRS-based structural variant calling using Sniffles2^125^ in population mode, as described in Jensen *et al.*^53^ LD between HP1 and candidate SNVs was computed using the cubic Hill equation and implemented via the CubeX formula in Python, based on Gaunt TR et al. 2007^126^. SNVs with R^2^ > 0.1 were retained for HP1 prediction. A Support Vector Classifier (SVC) was trained with a polynomial kernel (degree = 3), 100 permutations, and 70/30% train/test data splits. A consensus list of 20 SNVs was retained, of which 19 were shared across all stage 1 & 2 AD GWAS cohorts and CSF and plasma pQTL cohorts, in which the model was used to perform HP1 imputation. Additional details are provided in the **Supplementary Methods**. The plasma pQTL data (N_Subj_=2,338), largely overlapping the CSF pQTL data, were obtained in EUR ancestry subjects, and used the same SomaLogic 7K technology with consistent QC steps (cf. Yang *et al.* 2025 for additional details^127^).

### Haptoglobin - HP1 conditional analyses & APOE interaction

For 3 variants of interest at the *HP* locus, stage 1 and 2 AD GWAS and CSF/plasma pQTL analyses were repeated while conditioning on HP1 genotype, while otherwise keeping all model parameters consistent. Additionally, to assess the interaction between *HP* and *APOE* on AD risk, sex by *APOE* genotype (*APOE* ε44, *APOE* ε34, *APOE* ε33, *APOE* ε23) stratified analyses were performed in stage 1 and 2 GWAS data. To increase specificity for *APOE* genotype effects, stage 2 UKB analyses ignored proxy phenotypes and instead leveraged ICD codes and health record data (see **Supplementary Methods**).

### Haptoglobin – CSF and plasma proteome PheWAS & protein-protein interaction network

Phenome-wide association study (pheWAS) analyses were conducted with CSF (N=7,008 aptamers) and plasma (N=6,907 aptamers) proteogenomic data at the *HP* locus (± 1MB of *HP* TSS), including HP1 genotype and analyses both non-conditioned and conditioned on HP1. PheWAS were conducted using PLINK v.2.0^98^ with the --glm option, adjusting for age, genetic PCs, genotyping platform, and HP1 genotype in case of conditional analyses, followed by filtering for MAC ≥ 10 and genotype missingness ≤ 0.1. Significant pheWAS associations were determined for variants of interest (Bonferroni correction corresponding to number of aptamers per tissue). Resultant proteins as well as *APOE* (due to its known link with *HP*^87^) were used to construct a protein-protein interaction (PPI) network using GeneMania^128^. All proteins except TXNL4B were connected in the PPI network. Functional enrichment analysis was performed using the STRING PPI database v.12.0^129^ ‘Functional Enrichment Visualization’ tool to investigate Gene Ontology (GO) biological process and molecular function enrichment for the connected proteins.

## Data availability

Data used in the GWAS analyses are available upon application to:

- dbGaP (https://www.ncbi.nlm.nih.gov/gap/)
- NIAGADS (https://www.niagads.org/)
- LONI (https://ida.loni.usc.edu/)
- AMP-AD knowledge portal / Synapse (https://www.synapse.org/)
- Rush (https://www.radc.rush.edu/)
- NACC (https://naccdata.org/)
- UKB (https://www.ukbiobank.ac.uk/)
- FinnGen (https://www.finngen.fi/en)

The specific data repository and identifier for ADGC and ADSP data are indicated in **Tables-S1-2** of the supplement. Full GWAS summary statistics will be available in NIAGADS and GWAS Catalogue upon publication.

The protein weights used in the study are publicly available:

- Brain (https://www.synapse.org/Synapse:syn51150434)
- CSF (https://neurogenomics.wustl.edu/open-science/raw-data/)

Data used in the xQTL colocalization analyses are publicly available:

- eQTLgen (https://www.eqtlgen.org/cis-eqtls.html)
- UK Biobank PPP (https://metabolomips.org/ukbbpgwas/)
- ARIC (http://nilanjanchatterjeelab.org/pwas/)
- Wingo et al. DLPFC pQTL and eQTL (https://www.synapse.org/Synapse:syn51150434/wiki/621280)
- Western et al. CSF pQTL (https://www.ebi.ac.uk/gwas/publications/39528825)
- MetaBrain (https://download.metabrain.nl/files.html)
- BrainMeta (https://yanglab.westlake.edu.cn/software/smr/#DataResource)
- Brain xQTL serve (http://mostafavilab.stat.ubc.ca/xqtl/)
- Fujita et al. brain single cell eQTL data (https://www.synapse.org/Synapse:syn52335807)
- Kosoy et al. microglia eQTL data (https://www.synapse.org/Synapse:syn30308484)
- Kosoy et al. microclia caQTL data (https://www.synapse.org/Synapse:syn30308248)
- eQTL Catalogue database (https://www.ebi.ac.uk/eqtl/Data_access/)

A table overview of all QTL resources and their public identifiers are indicated in **Table-S23** of the supplement. A public repository of all locus compare plots, with colocalization (PP4 ≥ 0.7) from the xQTL colocalization analyses, can be accessed at **10.5281/zenodo.16989952**.

## Code availability

Codes for this work are available at https://github.com/Belloy-Lab/AD_SexBiased_Genomics_Proteomics. We used publicly available software for all analyses in this study. This included PLINK 2, BOLT-LMM (v.2.5), Regenie, and GWAMA (v.1.2.6), perform sex-stratified GWAS, LDSC (v.2.0.0) to format summary statistics, and FUSION for PWAS. Variant annotation was performed using VEP release 114. SMR (v.1.4.0) was used to perform SMR and HEIDI analyses, and coloc (v.4.2.1) R package to perform colocalization. locuszoomr (v.0.3.8), locuscomparer (v.1.0.0), and ggplot2 (v.3.5.2) were used to create locus zoom, locus compare, and multi-tag locus plots. Enrichment analyses and associated figure generation was performed using clusterProfiler (v.4.12.6), org.Hs.eg.db (v.3.21.0), simplifyEnrichment (v.1.14.1), HGNChelper (v.0.8.14), and ComplexHeatmap (v.2.20.0) R packages. Other figures utilized cowplot (v.1.2.0), scales (v.1.4.0), patchwork (v.1.3.2), ComplexUpset (v.1.3.3), ggrepel (v.0.9.6), and ggpubr (v.0.6.1) R packages.

## Acknowledgements

We thank all study participants and their families as well as many involved institutions and their staff. This work was supported by grants from the National Institutes of Aging (R00AG075238, M.E.B.; R00AG071837, C.B.Y.; R01AG089509, Z.H.; R01AG064614, C.C.; R01AG058501, C.C.; R01AG078964, C.C.; R01AG071706, C.C.; I01BX005686, A.P.W.; IK4BX005219, A.P.W.; R01AG072120, A.P.W., T.S.W.; R01AG075827, A.P.W., T.S.W.; R01AG074007, Y.J.S.) and the Alzheimer’s Association (AARFD-21-849349, C.B.Y.). This work was supported by access to equipment made possible by the Hope Center for Neurological Disorders, the Neurogenomics and Informatics Center (NGI: https://neurogenomics.wustl.edu/) and the Departments of Neurology and Psychiatry at Washington University School of Medicine.

We would like to thank the following resources for allowing access to their data: ADGC, ADSP, UK Biobank, FinnGen, AMP-AD. UK Biobank data were analyzed under Application Number 45420. Detailed acknowledgements for different genetic cohorts and biobanks are provided in the supplementary material.

## Author contributions

N.C. performed data acquisition, designed analyses, performed analyses, and wrote paper. Chenyu.Y., Y.Z., S.S., L.T., D.W., Chengran.Y., and Y.L. performed analyses. L.T. designed and performed HP1 imputation from long-read sequencing data. Y.L.G., I.S., and V.N. performed data acquisition and performed analyses. C.B.Y., E.C.M., A.A., A.P.W., T.S.W., and C.C. contributed data and resources. Y.J.S. contributed data and resources and designed analyses. Z.H. designed analyses. M.D.G. designed study, supervised analyses, and supervised work. M.E.B. performed data acquisition and analyses, designed analyses, designed study, supervised analyses, supervised work, wrote paper, and obtained funding.

