## Supplemental Document for "Integrative Genetic, Proteogenomic, and Multi-omics Analyses Reveal Sex-Biased Causal Genes and Drug Targets in Alzheimer’s Disease"

### Supplementary information

### Supplementary Methods

#### Genome-wide association studies

##### Stage 1 - ADGC & ADSP Phenotypic Data Quality Control and Processing

In the current study, we used data from a variety of cohorts and sequencing projects related to AD[1–22]. All available genetic/phenotypic data were jointly harmonized with the purpose of performing phenotype/covariate harmonization. Details are provided below.

###### *Cohorts and Phenotype Ascertainment*

Details on phenotype ascertainment are described elsewhere[1–5,7]. Briefly, all individuals with a diagnosis of AD met National Institute of Neurological and Communicative Disorders and Stroke/Alzheimer's Disease and Related Disorders Association (NINCDS-ADRDA) criteria for definite, probable, or possible late-onset AD[6], or met Diagnosis and Statistical Manual of Mental Disorders IV-V (DSMIV-V) criteria[8,9], or had a clinical dementia rating (CDR® Dementia Staging Instrument[10]) > 0.5. Some cohorts verified AD diagnoses through neuropathology, using Braak staging[11], CERAD scoring[21], or National Institute on Aging Reagan (NIA-Reagan) 1997 criteria[12]. Cognitively normal subjects did not have AD according to the above clinical AD criteria, did not have a diagnosis of mild-cognitive impairment (MCI), and had a CDR of 0 and/or Mini-Mental State Examination (MMSE[13]) > 25. In MIRAGE, control status was evaluated through a Modified Telephone Interview of Cognitive Status score ≥ 86 (a telephone version of the MMSE)[14].

Further, the National Alzheimer's Coordinating Center (NACC), Rush University Religious Orders Study/Memory and Aging Project (ROSMAP), and Alzheimer's Disease Neuroimaging Initiative (ADNI), are longitudinal cohorts that provide detailed information regarding clinical status (control, MCI, demented) and presumed disease etiology at repeated examinations. Additionally, deceased subjects are assessed for neuropathology. Where possible, in NACC, a final diagnosis of MCI or possible/probable/definite AD was obtained using NIA Alzheimer's Association (NIA-AA) 2011 criteria[15,16]. In all three cohorts, AD diagnoses were verified by neuropathology as middle or high AD likelihood following NIA-Reagan 1997 criteria (moderate to frequent neuritic plaques and Braak stage III-VI)[12]. In concordance with the category "possible AD dementia with evidence of the AD pathophysiological process" from the NIA-AA 2011 criteria[15], we attributed possible AD diagnoses to subjects who met clinical criteria for non-AD dementia but also met AD neuropathological criteria. In concordance with the NIA-AA 2011/2012 framework[16,17], we also evaluated neuropathology in MCI subjects to verify presumed AD etiology. Controls were not re-evaluated based on neuropathology data. Subjects that reverted from dementia to control status during longitudinal follow-up were excluded. Additional cohort-specific details are listed below.

###### *NACC*

Genotyping waves 1 through 7 from the Alzheimer's Disease Centers (ADC1-7) and a subset of the ADSP projects include subjects ascertained and evaluated by the clinical and neuropathological cores of 32 NIA-funded ADCs. NACC coordinates the collection of these phenotypes, implements diagnoses (cognitively normal, cognitively impaired but not MCI, MCI, demented; and presumed disease etiology), and then provides all data to researchers under the form of the Minimum Data Set (MDS), Uniform Data Set (UDS)[18,19,22], and Neuropathology data set (NP)[20]. The MDS represents an older subset of the NACC data and only contains cross-sectional data, while the more recent UDS provides longitudinal phenotypes and covariates. Since 2015, the UDS was updated to incorporate the NIA-AA 2011 criteria for MCI and AD[16,23]. In the current study, we used the UDS and NP for which data was collected between September 2005 and March 2022, to determine phenotypes for subjects in ADC1-7, ADSP WES/WGS, and ADGC Exome arrays.

Subjects that had a diagnosis of Down syndrome, central nervous system neoplasm, bipolar disorder, schizophrenia, alcohol-induced dementia, or substance-abuse-induced dementia, were excluded. Subjects carrying mutations of dominantly inherited AD or frontotemporal lobar degeneration (FTLD) were also excluded. Subjects with a final diagnosis of MCI or dementia, for which the etiology was unknown, not due to AD, or only secondary due to AD (and without AD neuropathological information), were excluded. Subjects with a final diagnosis of "cognitively impaired but not MCI", but having no other neurological disorder, were kept as controls, considering that this more consistently matched control criteria in many of the other cohorts considered in this study.

##### *ROSMAP*

In ROSMAP, subjects were diagnosed at each visit: as possible/probable AD according to NINCDS-ADRDA criteria[6]; as MCI when judged to have cognitive impairment but not meeting dementia criteria according to the clinician; or as control when there was no cognitive impairment or the subject did not meet dementia criteria[24,25]. At time of death, a final clinical diagnosis was made by an expert neurologist, followed by a case conference consensus review (blinded to postmortem data)[26].

##### *ADNI*

In ADNI, subjects were diagnosed at regular visits: as possible/probable AD according to NINCDS-ADRDA criteria[6]; as MCI according to Petersen/Winblad criteria; or as control when not demented, not MCI, CDR = 0, and MMSE > 28. Neuropathology assessments followed the NACC NP framework.

##### *Phenotype Harmonization*

The available sample contained many subjects that were genotyped multiple times across different studies. This largely reflected efforts from the ADGC, ADSP, and AMP-AD, to perform next-generation sequencing (NGS) on existing cohort samples for the purpose of rare variant discovery and AD gene prioritization. In other instances, participants were recruited in different studies at different times. Therefore, to handle potential duplicate discordance and phenotype heterogeneity, we implemented a cross-sample phenotype harmonization procedure aiming to standardize pathology-verified diagnoses where possible, share unique missing information across all duplicate entries of a given subject, resolve longitudinal changes in diagnosis, and flag subjects with unresolvable duplicate discordance for exclusion.

Duplicate samples were identified by determining genetic cryptic relatedness (cf. below), but for sample cross-referencing did not include known identical twins in LOAD and ROSMAP samples. First, duplicate samples were flagged as discordant if their age-at-death information differed by more than 2 years or if pathology measures (Braak or neuritic plaque density) differed. Across all cohorts, where possible, AD diagnoses were verified by neuropathology as middle or high AD likelihood following NIA-Reagan 1997 criteria (moderate to frequent neuritic plaques and Braak stage III-VI)[12]. Additionally, when only either neuritic plaque or Braak information was available and in line with NIA-Reagan 1997 middle or high AD likelihood criteria, and/or the cohort/project demographics provided a diagnosis of definite AD, the subject was considered to have pathology-verified AD status. Cognitively normal (CN) subjects with evidence of AD pathology were kept as CN. Further, if at least one entry across duplicate samples indicated a diagnosis of Down syndrome, central nervous system neoplasm, bipolar disorder, schizophrenia, alcohol-induced dementia, substance-abuse-induced dementia, neurological (not including Parkinson's disease), or systemic disease despite being cognitively normal, or carrying mutations of dominantly inherited AD or frontotemporal lobar degeneration (FTLD), then all duplicate samples were marked as such and flagged for exclusion. Extending on the above, all genetic samples were checked for the presence of known pathogenic mutations on *APP, PSEN1, PSEN2,* and *MAPT*, whereby carriers and their duplicate samples were flagged for exclusion.

Then, duplicate samples with differing age entries (i.e. longitudinal changes) were evaluated. Reversions from AD or dementia to MCI status, or from MCI to cognitively normal (CN) status, were permitted, but reversions from AD or non-AD dementia to CN status were flagged for exclusion. "Reversions" from AD to non-AD dementia status were permitted, unless pathology (cf. above) indicated the presence of AD pathology, thereby marking the subject as AD. Vice versa, "conversions" from non-AD dementia to AD status were permitted, unless pathology (cf. above) indicated no presence of AD pathology, thereby marking the subject as non-AD dementia. All other types of conversions were directly permitted. Then, duplicate samples for which the diagnoses at the oldest shared age entries differed, or for which diagnoses differed but age was consistent (i.e. apparent cross-sectional discordances), were evaluated. Discordances between AD and non-AD dementia status were resolved based on pathology (cf. above) or flagged as discordant if no pathology data was available. Discordances between CN and AD status, or CN and non-AD dementia status, were resolved as respectively AD or non-AD dementia when those dementia diagnoses corresponded to a unique age-at-onset (of symptoms) without other available age information (i.e. indicating that a conversion likely occurred after the subject was lost to follow-up in the cohort that last observed a CN status), or, were flagged as discordant if duplicate entries shared the same age-at-examination and age-at-last-exam. Discordances between CN and MCI status, or MCI and AD status, or MCI and non-AD dementia status, were resolved as respectively MCI, AD, or non-AD dementia (i.e. keeping the most severe diagnosis).

Finally, once all clinical diagnostic and pathological data were unified across duplicate entries, pathological criteria were applied once more to obtain the final diagnoses. Where possible, AD diagnoses were verified by neuropathology as middle or high AD likelihood following NIA-Reagan 1997 criteria (moderate to frequent neuritic plaques and Braak stage III-VI)[12]. In concordance with the category "possible AD dementia with evidence of the AD pathophysiological process" from the NIA-AA 2011 criteria[15], we attributed possible AD diagnoses to subjects who met clinical criteria for non-AD dementia but also met AD neuropathological criteria. In concordance with the NIA-AA 2011/2012 framework[16,17], we also evaluated neuropathology in MCI subjects to verify presumed AD etiology and considered subjects as cases if AD pathology, following NIA-Reagan 1997 criteria (cf. above), was present (i.e. marking high likelihood of AD etiology). Controls were not re-evaluated based on neuropathology data.

Beyond cross-referencing clinical diagnostic and pathological data across subjects, other covariates were considered for cross-referencing or sharing in case of missingness across duplicate entries. These included age-at-onset of cognitive symptoms, age-at-examination providing clinical diagnosis, at-at-last exam, age-at-death, sex, race, ethnicity, *APOE* genotype provided from demographics, *APOE* genotype provided from whole-genome sequencing, and *APOE* genotype provided from whole-exome sequencing. Duplicate entries with discordant sex or race information were flagged for exclusion.

#### Stage 1 - ADGC and ADSP Genetic Data Quality Control and Processing

##### *Ascertainment of Genetic Data*

Genotypes were available from high-density single-nucleotide polymorphism (SNP) genotyping microarrays (Illumina or Affymetrix) for ADGC or whole genome sequencing (WGS) for ADSP (**Tables-S1-2**). Genotype samples had their genetic variants lifted to hg38 using liftOver if not released in hg38 and annotated using dbSNP153 variant identifiers[27].

##### *ADGC Autosomal Quality Control and Imputation*

Autosomal variants were extracted from the SNP array data and further processed in several stages. In each cohort/platform/array, variants were excluded based on genotyping rate (<95%), MAF<1%, and Hardy-Weinberg equilibrium in controls ($p<10^{-6}$) using PLINK v1.9[28]. As in our prior work[29], information derived from the gnomAD v.3.1 database[30] was used to filter out SNPs that met one of the following exclusion criteria: (i) located in a low complexity region, (ii) located within common structural variants (MAF > 1%), (iii) multiallelic SNPs with MAF > 1% for at least two alternate alleles, (iv) located within a common insertion/deletion, (v) having any flag different than PASS in gnomAD, (vi) having potential probe polymorphisms, and (vii) more than 10% MAF difference with gnomAD frequency in non-Finnish Europeans. The remaining SNPs were checked for consistency with the TOPMed panel, flipping of palindromic SNPs, and were imputed on the TOPMed Imputation server[31,32], which uses Minimac 4 for imputation. The following parameters were selected: reference panel TOPMed-r2 (2022), phasing with Eagle v2.4, r-square imputation score cut off 0.3.

##### *ADSP Autosomal Quality Control*

The ADSP WGS data (NG00067.v5) were joint called by the ADSP following the SNP/Indel Variant Calling Pipeline and data management tool used for the analysis of genome and exome sequencing for the Alzheimer's Disease Sequencing Project (VCPA)[33]. The current analyses of

ADSP WGS were restricted to bi-allelic variants, to which we applied the Variant Quality Score Recalibration (VSQR) quality control filter ("PASS" variants; GATK v4.1)[34]. Variants with a genotyping rate less than 80%, deviating from Hardy Weinberg Equilibrium (HWE) in the full sample or in controls ($p<10^{-6}$), and a minor allele count less than 10, were excluded. Consistent with the methodology detailed in Belloy et al. 2022[35], we then applied several filters to remove artifactual variants: (i) variants that represented sequencing center or platform artifacts as identified by Fisher exact testing in controls ($p<10^{-5}$), (ii) variants reported in gnomAD v3.1[30] to have a "non-PASS", falling in a low complexity region, or showing more than 10% allele frequency deviation between our European ancestry control participants in ADSP and non-Finnish European participants in gnomAD, and (iii) duplicate discordance variants that show discrepancies across several 100 technical duplicates present in ADSP.

##### *Genetic Relationship Determination using King*

Across all cohorts, the relatedness of subjects (after QC indicated above) was evaluated through identity-by-descent (IBD) analysis (using directly genotyped non-palindromic SNPs shared across all genetic datasets with a call rate > 95% & minor allele frequency (MAF)>1%)[36]. This outcome was used for duplicate tracking across samples, which in turn was used to enable phenotype harmonization (cf. above).

##### *Ancestry Determination*

Individual ancestries were determined using SNPweights v.2.1 with populations from the 1000 Genomes Consortium as a reference[37,38]. By applying an ancestry percentage cut-off ≥ 75%, the samples were stratified into the five super populations, South-Asians (SAS), East-Asians (EAS), Amerindians (AMR), Africans (AFR) and Europeans (EUR) (**Figure-S1**). When multiple samples were available for a single unique individual, the ancestry was inferred from the sample with the highest genetic coverage.

##### *Relationship Determination and Principal Component Analysis using GENESIS*

For ADGC and ADSP data respectively, the relatedness of subjects and principal components capturing population substructure were determined using IBD and principal component analyses (PCA) as implemented through the R package GENESIS (R v3.6.0)[39]. Specifically, this approach first uses an R-implementation of KING-robust to determine kinship coefficients that take into account ancestry divergence. The derived pairwise kinship coefficients are then used to perform a PCA in related samples (PC-AiR) providing accurate ancestry inference not confounded by family structure. The latter output is then used to estimate kinship coefficients using PC-Relate, which accounts for population structure (ancestry) among sample individuals through the use of ancestry representative principal components (PCs) to provide accurate relatedness estimates due only to recent family (pedigree) structure. For each respective data set, these analyses were performed on pruned SNPs ($R^2 < 0.5$, call rate > 95%, MAF > 1%, and excluding palindromic SNPs) in non-Hispanic White European ancestry individuals.

#### Stage 1 - ADGC & ADSP Statistical Analyses

##### *Case-control GWAS*

All association analyses with AD risk were stratified by sex and adjusted for array type, the first 5 genetic principal components (PC-AiRs), *APOE**4 dosage (0/1/2), and *APOE**2 dosage (0/1/2). Age adjustment in case-control analyses was not performed, given that the current AD genetic samples often showed younger ages for cases than controls due to the use of age-at-onset information (**Table-S3**), which violates the assumption for age adjustment (which is that older age is associated with increased AD incidence). In prior work, we showed that age adjustment in such scenarios leads to significantly decreased power for genetic association analyses[29]. Adjustment for *APOE* genotypes is relevant given the established interactions with sex[40]. Additionally, the case-control clinical cohorts are enriched for *APOE**4 cases compared to population-based studies[40], which may further exacerbate any potential confounding effects.

Cohorts from ADGC were pooled into a mega-analysis. BOLT-LMM was used in both ADGC and ADSP[41], deriving genetic relationship matrices to allow the inclusion of related subjects. Resultant betas were converted to traditional odds ratios using the transformation approach as detailed in the BOLT-LMM manual. Across ADGC and ADSP, subjects were unrelated down to 1st degree.

##### *Age information*

For cases that only had age-at-death (AAD) available, the final ages used for regression analysis were subtracted by 10 years to approximate age-at-onset (AAO). This reflects expected mean delays between AAO and AAD for AD patients[42], and is consistent with the derived age covariate for AD cohorts provided by the Alzheimer's Disease Genetics Consortium (ADGC) on NIAGADS[43]. In cohorts that provide conversion information but not AAO, age-at-examination (AAE) was used and followed a prioritization of age-at-MCI-diagnosis > age-at-dementia-diagnosis (incident) > age-at-dementia diagnosis (prevalent). This was done to most closely approximate AAO. For the remaining control samples, age-at-last-examination (AAL) was used. After implementing these criteria, samples were filtered to have a minimal age of 60 years. Some samples were censored at ages 90+, for which we assumed the age was 90 (since there was no way to estimate the actual age).

#### Stage 2 - UKB Phenotype ascertainment

Detailed descriptions of all the variables and fields provided by UKB are provided elsewhere[44].

In the first round of phenotype ascertainment, we derived health-registry-confirmed AD status and related age information for the individuals directly. Subjects were assumed to be controls if they had no other diagnosis inferred from health registry information relevant to dementia status. We specifically considered the following data fields and entries: *Diagnoses_main_ICD10* [G300,G301,G308,G309,F000,F001,F002,F009], *Diagnoses_secondary_ICD10* [G300,G301,G308,G309,F000,F001,F002,F009], *Date_of_first_in_patient_diagnosis_main_ICD10* [if date provided], *Date_of_first_in_patient_diagnosis_ICD10* [if date provided], *Source_of_alzheimers_disease_report* [0,1,11,12,2,21,22 = self report, hospital admission, death record],

*Date_of_alzheimers_disease_ report* [if date provided], *Source_of_all_cause_dementia_report* [0,1,11,12,2,21,22 = self report, hospital admission, death record], *Source_of_frontotemporal_dementia_report* [any entry], *Source_of_vascular_ dementia_report* [any entry], and *Date_of_all_cause_dementia_report* [if data provided]. The above fields were used to determine dementia status, allowing us to differentiate between late-onset AD individuals (LOAD), early-onset AD (EOAD), vascular dementia, frontotemporal dementia, and other all-cause dementia participants. For the health-registry AD phenotype, cases were restricted to all LOAD individuals. The above fields were further used to determine the earliest available age at which a dementia occurrence or report was made. Age information for controls was available from the variables: *Age_when_attended_assessment_centre* [oldest age entry retrieved] and *Age_at_death*.

In the second round, we derived proxy Alzheimer's disease or dementia case and control status, and related age, by accessing the following fields and entries: *Illnesses_of_father*, *Illnesses_of_mother*, *Illnesses_of_sibblings*, *Fathers_age*, *Fathers_age_at_death*, *Mothers_age*, and *Mothers_age_at_death* (where it should be noted that age and sex info was not available for siblings). The youngest reported age was used for proxy ADD cases, while the oldest reported age was used for proxy controls. Proxy status was ignored if subjects were adopted.

To build the final phenotypes, we took into account recent observations by Wu et al. 2024 that showed AD proxy GWAS can lead to biases in genetic associations[45]. The authors specifically noted that bias could be reduced by using the proxy GWAS approach by Marrioni et al. 2018, which we adapted[46]. This notably entails that parental ages were >65y and that parental age should be added as a covariate in GWAS. We focused on using the parental phenotypes, but when health-registry-confirmed AD status was available (a small fraction of case numbers; cf. **Table-S3**); we used those instead of proxy phenotypes. We finally included additional steps to render phenotypes sex specific. In summary, the following steps were included to obtain sex-specific AD phenotypes:

- Female AD GWAS phenotype:
  - Keep female individuals with health-registry-confirmed AD status and ages > 60y.
  - Keep other individuals with maternal ages > 65y, excluding subjects that have a father or sibling with Alzheimer's disease or dementia (to render the phenotype female specific).
- Male AD GWAS phenotype:
  - Keep male individuals with health-registry-confirmed AD status and ages > 60y.
  - Keep other individuals with paternal ages > 65y, excluding subjects that have a mother or sibling with Alzheimer's disease or dementia (to render the phenotype male specific).

For targeted genetic analyses in the Haptoglobin locus stratified by sex and *APOE* genotype, we considered only health-registry-confirmed AD case control status in subjects ages > 60y.

#### Stage 2 - UKB Genetic Data Quality Control and Processing

A Detailed description of all the UKB genetic data and processing is provided elsewhere[44]. Specifically, we accessed SNP array data imputed to the Haplotype Reference Consortium (HRC) and UK10K haplotype resource. We further filtered to subjects with consent, passing sex check QC, no heterozygosity outliers, having age information available, and belonging to a white ethnic background (field *Ethnic_background* [1001,1002,1003]). We then identified a homogenous ancestry cluster within this group using "aberrant" on the first 20 genetic PCs, as in Schwartzentruber et al. 2021[47].

#### Stage 2 - UKB Statistical Analyses

All AD GWAS were stratified by sex-specific AD phenotypes (cf. above) and adjusted for array type, assessment center, the first 20 genetic principal components provided by UKB, subject age, parental age, and *APOE**4/2 dosage. BOLT-LMM was used (as was done for ADGC and ADSP)[41], using autosomal data to derive genetic relationship matrices to allow the inclusion of related subjects. Resultant betas were converted to traditional odds ratios using the transformation approach as detailed in the BOLT-LMM manual. Additionally, since the UKB GWAS leveraged the proxy phenotype, an additional correction factor (multiply by 2) was needed to rescale beta coefficients and standard errors onto a regular case-control scale (cf. Liu et al. 2017)[48].

#### Stage 3 - FinnGen

The FinnGen study is a large-scale genomics initiative that has analyzed over 500,000 Finnish biobank samples and correlated genetic variation with health data to understand disease mechanisms and predispositions. The project is a collaboration between research organisations and biobanks within Finland and international industry partners.

Ascertainment of FinnGen phenotype and genotype data is described in detail elsewhere[49]. Sex stratified analyses were conducted on the broader AD phenotype "G6_NEURODEGENERATIVE" using data release R12 (in releases prior to R12, this phenotype was referred to as "Alzheimer's disease, wide definition", which is the same one as used in prior AD GWAS[50,51]). Documentation on Genetic data processing and statistical analyses is provided here: https://finngen.gitbook.io/documentation. FinnGen made use of Regenie to include related individuals in the genetic association analyses[52]. Information about phenotypes and endpoints is provided here: https://www.finngen.fi/en/researchers/clinical-endpoints, https://r12.risteys.finngen.fi/

##### *FinnGen ethics statement*

Study subjects in FinnGen provided informed consent for biobank research, based on the Finnish Biobank Act. Alternatively, separate research cohorts, collected prior the Finnish Biobank Act came into effect (in September 2013) and start of FinnGen (August 2017), were collected based on study-specific consents and later transferred to the Finnish biobanks after approval by Fimea (Finnish Medicines Agency), the National Supervisory Authority for Welfare and Health. Recruitment protocols followed the biobank protocols approved by Fimea. The Coordinating Ethics Committee of the Hospital District of Helsinki and Uusimaa (HUS) statement number for the FinnGen study is Nr HUS/990/2017.

The FinnGen study is approved by Finnish Institute for Health and Welfare (permit numbers: THL/2031/6.02.00/2017, THL/1101/5.05.00/2017, THL/341/6.02.00/2018, THL/2222/6.02.00/2018, THL/283/6.02.00/2019, THL/1721/5.05.00/2019 and THL/1524/5.05.00/2020), Digital and population data service agency (permit numbers: VRK43431/2017-3, VRK/6909/2018-3, VRK/4415/2019-3), the Social Insurance Institution (permit numbers: KELA 58/522/2017, KELA 131/522/2018, KELA 70/522/2019, KELA 98/522/2019, KELA 134/522/2019, KELA 138/522/2019, KELA 2/522/2020, KELA 16/522/2020), Findata permit numbers THL/2364/14.02/2020, THL/4055/14.06.00/2020, THL/3433/14.06.00/2020, THL/4432/14.06/2020, THL/5189/14.06/2020, THL/5894/14.06.00/2020, THL/6619/14.06.00/2020, THL/209/14.06.00/2021, THL/688/14.06.00/2021, THL/1284/14.06.00/2021, THL/1965/14.06.00/2021, THL/5546/14.02.00/2020, THL/2658/14.06.00/2021, THL/4235/14.06.00/2021, Statistics Finland (permit numbers: TK-53-1041-17 and TK/143/07.03.00/2020 (earlier TK-53-90-20) TK/1735/07.03.00/2021, TK/3112/07.03.00/2021) and Finnish Registry for Kidney Diseases permission/extract from the meeting minutes on 4th July 2019.

The Biobank Access Decisions for FinnGen samples and data utilized in FinnGen Data Freeze 12 include: THL Biobank BB2017_55, BB2017_111, BB2018_19, BB_2018_34, BB_2018_67, BB2018_71, BB2019_7, BB2019_8, BB2019_26, BB2020_1, BB2021_65, Finnish Red Cross Blood Service Biobank 7.12.2017, Helsinki Biobank HUS/359/2017, HUS/248/2020, HUS/430/2021 §28, §29, HUS/150/2022 §12, §13, §14, §15, §16, §17, §18, §23, §58, §59, HUS/128/2023 §18, Auria Biobank AB17-5154 and amendment #1 (August 17 2020) and amendments BB_2021-0140, BB_2021-0156 (August 26 2021, Feb 2 2022), BB_2021-0169, BB_2021-0179, BB_2021-0161, AB20-5926 and amendment #1 (April 23 2020) and it´s modifications (Sep 22 2021), BB_2022-0262, BB_2022-0256, Biobank Borealis of Northern Finland_2017_1013, 2021_5010, 2021_5010 Amendment, 2021_5018, 2021_5018 Amendment, 2021_5015, 2021_5015 Amendment, 2021_5015 Amendment_2, 2021_5023, 2021_5023 Amendment, 2021_5023 Amendment_2, 2021_5017, 2021_5017 Amendment, 2022_6001, 2022_6001 Amendment, 2022_6006 Amendment, 2022_6006 Amendment, 2022_6006 Amendment_2, BB22-0067, 2022_0262, 2022_0262 Amendment, Biobank of Eastern Finland 1186/2018 and amendment 22§/2020, 53§/2021, 13§/2022, 14§/2022, 15§/2022, 27§/2022, 28§/2022, 29§/2022, 33§/2022, 35§/2022, 36§/2022, 37§/2022, 39§/2022, 7§/2023, 32§/2023, 33§/2023, 34§/2023, 35§/2023, 36§/2023, 37§/2023, 38§/2023, 39§/2023, 40§/2023, 41§/2023, Finnish Clinical Biobank Tampere MH0004 and amendments (21.02.2020 & 06.10.2020), BB2021-0140 8§/2021, 9§/2021, §9/2022, §10/2022, §12/2022, 13§/2022, §20/2022, §21/2022, §22/2022, §23/2022, 28§/2022, 29§/2022, 30§/2022, 31§/2022, 32§/2022, 38§/2022, 40§/2022,

42§/2022, 1§/2023, Central Finland Biobank 1-2017, BB_2021-0161, BB_2021-0169, BB_2021-0179, BB_2021-0170, BB_2022-0256, BB_2022-0262, BB22-0067, Decision allowing to continue data processing until 31st Aug 2024 for projects: BB_2021-0179, BB22-0067,BB_2022-0262, BB_2021-0170, BB_2021-0164, BB_2021-0161, and BB_2021-0169, and Terveystalo Biobank STB 2018001 and amendment 25th Aug 2020, Finnish Hematological Registry and Clinical Biobank decision 18th June 2021, Arctic biobank P0844: ARC_2021_1001.

#### Proteome-wide association studies

##### Brain PWAS data preparation

Non-sex-stratified and sex-stratified brain variant protein weights were created, using FUSION, in human genome build hg19/GRCh37(cf. Wingo et al. 2023 for details)[53]. AD GWAS summary statistics were thus lifted from hg38/GRCh38) to hg19/GRCh37, using the UCSC LiftOver tool[54]. EUR AD GWAS summary statistics were intersected by genomic position and allele matching with an LD reference panel built from 1,000 Genomes Project (Phase 3)[37] EUR samples (10,871,685 variants; minor allele frequency [MAF] > 0.005). Admixed AFR GWAS were intersected with an LD reference panel from 1,000 Genomes Project (Phase 3)[37] EUR, AFR, and Amerindian (AMR) samples (18,421,947 variants; MAF > 0.005). GWAS summary statistics were than processed with mungestats, filtering variants to MAF > 0.1%, and used for PWAS in FUSION[55] with default settings.

##### CSF PWAS data preparation

CSF variant protein weights were created, using FUSION, in build hg38, with non-stratified data available from Western et al. 2024[56] and sex-stratified data generated from the same samples. AD GWAS summary statistics were intersected by genomic position and allele matching with LD reference panels derived directly from the CSF proteogenomic data[56]. GWAS summary statistics were than processed with mungestats, filtering variants to MAF > 0.1%, and used for PWAS in FUSION[55] with default settings.

#### SMR and HEIDI data preparation & analysis

Sex-stratified EUR GWAS summary statistics were processed to retain *cis* SNPs within ±1 Mb of PWAS genes' median base pair position (**Methods**) for summary-based Mendelian randomization (SMR) and heterogeneity in dependent instruments (HEIDI) analyses using the SMR software tool[57]. Brain and CSF pQTL data were converted from ESD to BESD format, and GWAS statistics to "ma" format. SMR tested the top pQTL per gene for causal associations between protein expression and AD risk. The pQTL threshold parameter was relaxed from the default 5e-8 to 1e-3 to enhance the feasibility of testing genes whose top cis-pQTLs did not reach genome-wide significance. The HEIDI test was applied to assess whether observed associations were due to true causal effects or LD-induced heterogeneity, using LD data from stage 1 ADGC GWAS data. HEIDI analysis included SNPs with *P*-values below the default threshold of $1.57 \times 10^{-3}$, LD-pruned ($R^2$ 0.05–0.9), requiring ≥ 3 *cis* SNPs and ≤ 20 top-ranked SNPs.

#### Colocalization data preparation & analysis

Variants were selected within ±1 Mb of PWAS genes' median base pair position or lead GWAS variants (**Methods**). Across GWAS and QTL data, non-matching variants were excluded using genomic position alignment and allele matching, while affect alleles were aligned and filtered to those displaying ≤ 10% frequency deviations across GWAS and QTL data. If respective QTL studies used smaller window sizes, then variants were inherently restricted to those overlapping AD GWAS and QTL data. Datasets released with *trans* QTL results (i.e. genome-wide associations with respective molecular traits) were reprocessed to cis QTLs within ±1 Mb of GWAS lead index variants or PWAS gene's median base pair position.

Colocalization analyses were performed as described in the main **Methods**, using coloc.abf and coloc.susie from the coloc v.4.2.1[58,59] R package to validate and prioritize causal genes. Coloc.abf used the p-value approach, excluding beta coefficients and standard errors to avoid potential issues with variants close to MAF 50%. Coloc.susie used the runsusie function with effect size, variance, MAF inputs, and an LD matrix from stage 1 ADGC GWAS data to identify credible sets of independent causal variants. SuSiE employs an iterative Bayesian stepwise selection method to identify credible sets of variants; if SuSiE failed to converge after 7 days runtime, the respective analysis was terminated and no results were reported.

#### Enrichment analyses

##### Sex biased cell type enrichment analyses details

Gene expression data for human endothelial cells, oligodendrocytes, mature astrocytes, neurons, and microglia/macrophages were obtained from Zhang et al. 2016[60], comprising RNA-seq data from human brain samples across multiple regions (normalized counts). Sex-specific gene lists (female: N=125; male: N=21; **Table-S27**) and the background gene list (N=19,117) were updated using checkGeneSymbols (HGNChelper package, species = "human", unmapped.as.na = TRUE, expand.ambiguous = FALSE). Unmapped or duplicate gene symbols were excluded. Genes without expression in at least one cell type were removed to ensure reliable cell type-specificity assignments.

Enrichment was tested using the hypergeometric test ('phyper' function, stats package, R v.4.4.1) with parameters: q = overlap - 1, m = cell type-specific genes, n = background genes - cell type-specific genes, k = sex-specific genes, lower.tail = FALSE. The background gene set comprised all genes with cell type-specificity assignments (N=19,117). *P*-values were adjusted using the Benjamini-Hochberg method ($P_{FDR} < 0.05$). Gene ratio (overlap/cell type-specific genes), fold enrichment ((overlap/sex-specific genes)/(cell type-specific genes/background) - 1), and percent overlap (overlap/sex-specific genes × 100) were calculated.

##### Drug enrichment and repurposing analyses

The main **Methods** section details gene-drug network and drug enrichment analyses, including subsequent filtering steps to filter to sex biased drugs. To identify drugs relevant to sex hormone biology, we leveraged ChatGPT (OpenAI, GPT-4o, accessed July 2025 and GPT-5o, accessed September 2025) and Grok (xAI, Version 3, accessed September 2025) to match drugs to 6 classes of sex hormone related drugs: Estrogenic, Selective Estrogen Receptor Modulator (SERM), Androgenic, Aromatase Inhibitor, Progestogenic, and GNRH Agonist/Antagonist. We then filtered all unique drugs across each of the 3 queries to those that had at least 3 genes contributing to their significant enrichment (to increase robustness), leading to a final set of 13 female biased drugs that were manually reviewed for relevance to sex hormone biology and drug classes. These 13 drugs and their related genes were used to build a gene-drug graph in Cytoscape[61], which included gene-gene connections from PPI network data. In men, only 1 sex hormone related (progestogenic) drug was identified with at least 3 related genes, Medroxyprogesterone Acetate, which was not further explored.

#### HP1 linkage analyses and imputation

To identify individuals carrying the HP1 deletion (chr16:72,057,133, DEL, –1,716 bp), we developed a supervised prediction model using support vector classification (SVC). The model was trained on genotype data from 431 individuals with paired long-read sequencing (LRS) and short-read sequencing (SRS) available from the ADRC and Stanford Aging & Memory Study (SAMS) cohorts. These cohorts included 258 healthy controls, 91 individuals with AD or mild cognitive impairment (MCI), 81 individuals with alpha-synucleinopathies (Parkinson's disease or Lewy body disease), and 1 individual with an unknown diagnosis. The sample comprised 205 males and 226 females.

The target variable was derived from LRS-based structural variant calling using Sniffles2 in population mode, as described in https://www.medrxiv.org/content/10.1101/2025.10.10.25337775v2. High-molecular-weight DNA was extracted from primary blood mononuclear cells (PBMCs); samples were sequenced on the Oxford Nanopore PromethION48 platform, and reads were aligned to GRCh38. The HP1 deletion genotype was extracted for each individual.

Predictor variables consisted of single-nucleotide variants (SNVs) located within ±1 Mb of the deletion site, genotyped via SRS, mapped to GRCh38, and extracted using PLINK v.1.9., as described in https://www.medrxiv.org/content/10.1101/2025.10.10.25337775v2.

LD between candidate SNVs and the HP1 deletion was evaluated using $R^2$ and D′ statistics. Genotypes from both LRS (via Clair3) and SRS were harmonized using identity-by-descent filtering with PLINK v.1.9. LD was computed using the cubic Hill equation and implemented via the CubeX formula implemented in Python, based on Gaunt TR et al. 2007[62]. Only SNVs with $R^2 > 0.1$ with the HP[1] deletion were retained for downstream model training.

To train the classifier, we used an SVC with a polynomial kernel (degree = 3). A total of 100 permutations were conducted to account for feature selection variability. For each permutation:

- The data were split (70% train / 30% test, stratified by HP1 status);
- Low-variance SNVs were removed using a variance threshold of 0.01;

- The top 20 most informative SNVs were selected using SelectKBest (ANOVA F-score);
- A polynomial SVC (C = 1.0) was trained on the selected features;
- Training and test accuracy were recorded, and selected SNVs were saved.

After 100 permutations, a consensus list of the 20 most frequently selected SNVs was compiled (cf. **Figure-S27**). From this list, 19 SNVs were found to be shared across all imputed cohorts and were used to retrain the final prediction model on the entire ADRC/SAMS dataset (cf. **Figure-S28**). This retrained model was then applied to impute the HP1 deletion genotype in external cohorts (TOPMed, EUR, ADSP, UK Biobank, and the f05 subcohort) (cf. **Table-S34**).

### Acknowledgements

Data for this study were prepared, archived, and distributed by the National Institute on Aging Alzheimer's Disease Data Storage Site (NIAGADS) at the University of Pennsylvania (U24-AG041689), funded by the National Institute on Aging. The contents of this article do not represent the views of the National Institutes of Health, the U.S. Department of Veterans Affairs, or the United States Government.

#### Acknowledgments for the use of ADSP data

The Alzheimer's Disease Sequencing Project (ADSP) is comprised of two Alzheimer's Disease (AD) genetics consortia and three National Human Genome Research Institute (NHGRI) funded Large Scale Sequencing and Analysis Centers (LSAC). The two AD genetics consortia are the Alzheimer's Disease Genetics Consortium (ADGC) funded by NIA (U01 AG032984), and the Cohorts for Heart and Aging Research in Genomic Epidemiology (CHARGE) funded by NIA (R01 AG033193), the National Heart, Lung, and Blood Institute (NHLBI), other National Institute of Health (NIH) institutes and other foreign governmental and non-governmental organizations. The Discovery Phase analysis of sequence data is supported through UF1AG047133 (to Drs. Schellenberg, Farrer, Pericak-Vance, Mayeux, and Haines); U01AG049505 to Dr. Seshadri; U01AG049506 to Dr. Boerwinkle; U01AG049507 to Dr. Wijsman; and U01AG049508 to Dr. Goate and the Discovery Extension Phase analysis is supported through U01AG052411 to Dr. Goate, U01AG052410 to Dr. Pericak-Vance and U01 AG052409 to Drs. Seshadri and Fornage.
Sequencing for the Follow Up Study (FUS) is supported through U01AG057659 (to Drs. PericakVance, Mayeux, and Vardarajan) and U01AG062943 (to Drs. Pericak-Vance and Mayeux). Data generation and harmonization in the Follow-up Phase is supported by U54AG052427 (to Drs. Schellenberg and Wang). The FUS Phase analysis of sequence data is supported through U01AG058589 (to Drs. Destefano, Boerwinkle, De Jager, Fornage, Seshadri, and Wijsman), U01AG058654 (to Drs. Haines, Bush, Farrer, Martin, and Pericak-Vance), U01AG058635 (to Dr. Goate), RF1AG058066 (to Drs. Haines, Pericak-Vance, and Scott), RF1AG057519 (to Drs. Farrer and Jun), R01AG048927 (to Dr. Farrer), and RF1AG054074 (to Drs. Pericak-Vance and Beecham).
The ADGC cohorts include: Adult Changes in Thought (ACT) (UO1 AG006781, UO1 HG004610, UO1 HG006375, U01 HG008657), the Alzheimer's Disease Centers (ADC) ( P30 AG019610, P30 AG013846, P50 AG008702, P50 AG025688, P50 AG047266, P30 AG010133, P50 AG005146, P50 AG005134, P50 AG016574, P50 AG005138, P30 AG008051, P30 AG013854, P30 AG008017, P30 AG010161, P50 AG047366, P30 AG010129, P50 AG016573, P50 AG016570, P50 AG005131, P50 AG023501, P30 AG035982, P30 AG028383, P30 AG010124, P50 AG005133, P50 AG005142, P30 AG012300, P50 AG005136, P50 AG033514, P50 AG005681, and P50 AG047270), the Chicago Health and Aging Project (CHAP) (R01 AG11101, RC4 AG039085, K23 AG030944), Indianapolis Ibadan (R01 AG009956, P30 AG010133), the Memory and Aging Project (MAP) ( R01 AG17917), Mayo Clinic (MAYO) (R01 AG032990, U01 AG046139, R01 NS080820, RF1 AG051504, P50 AG016574), Mayo Parkinson's Disease controls (NS039764, NS071674, 5RC2HG005605), University of Miami (R01 AG027944, R01 AG028786, R01 AG019085, IIRG09133827, A2011048), the Multi-Institutional Research in Alzheimer's Genetic Epidemiology Study (MIRAGE) (R01 AG09029, R01 AG025259), the

National Cell Repository for Alzheimer's Disease (NCRAD) (U24 AG21886), the National Institute on Aging Late Onset Alzheimer's Disease Family Study (NIA- LOAD) (R01 AG041797), the Religious Orders Study (ROS) (P30 AG10161, R01 AG15819), the Texas Alzheimer's Research and Care Consortium (TARCC) (funded by the Darrell K Royal Texas Alzheimer's Initiative), Vanderbilt University/Case Western Reserve University (VAN/CWRU) (R01 AG019757, R01 AG021547, R01 AG027944, R01 AG028786, P01 NS026630, and Alzheimer's Association), the Washington Heights-Inwood Columbia Aging Project (WHICAP) (RF1 AG054023), the University of Washington Families (VA Research Merit Grant, NIA: P50AG005136, R01AG041797, NINDS: R01NS069719), the Columbia University HispanicEstudio Familiar de Influencia Genetica de Alzheimer (EFIGA) (RF1 AG015473), the University of Toronto (UT) (funded by Wellcome Trust, Medical Research Council, Canadian Institutes of Health Research), and Genetic Differences (GD) (R01 AG007584). The CHARGE cohorts are supported in part by National Heart, Lung, and Blood Institute (NHLBI) infrastructure grant HL105756 (Psaty), RC2HL102419 (Boerwinkle) and the neurology working group is supported by the National Institute on Aging (NIA) R01 grant AG033193.

The CHARGE cohorts participating in the ADSP include the following: Austrian Stroke Prevention Study (ASPS), ASPS-Family study, and the Prospective Dementia Registry-Austria (ASPS/PRODEM-Aus), the Atherosclerosis Risk in Communities (ARIC) Study, the Cardiovascular Health Study (CHS), the Erasmus Rucphen Family Study (ERF), the Framingham Heart Study (FHS), and the Rotterdam Study (RS). ASPS is funded by the Austrian Science Fond (FWF) grant number P20545-P05 and P13180 and the Medical University of Graz. The ASPS-Fam is funded by the Austrian Science Fund (FWF) project I904),the EU Joint Programme - Neurodegenerative Disease Research (JPND) in frame of the BRIDGET project (Austria, Ministry of Science) and the Medical University of Graz and the Steiermärkische Krankenanstalten Gesellschaft. PRODEM-Austria is supported by the Austrian Research Promotion agency (FFG) (Project No. 827462) and by the Austrian National Bank (Anniversary Fund, project 15435. ARIC research is carried out as a collaborative study supported by NHLBI contracts (HHSN268201100005C, HHSN268201100006C, HHSN268201100007C, HHSN268201100008C, HHSN268201100009C, HHSN268201100010C, HHSN268201100011C, and HHSN268201100012C). Neurocognitive data in ARIC is collected by U01 2U01HL096812, 2U01HL096814, 2U01HL096899, 2U01HL096902, 2U01HL096917 from the NIH (NHLBI, NINDS, NIA and NIDCD), and with previous brain MRI examinations funded by R01-HL70825 from the NHLBI. CHS research was supported by contracts HHSN268201200036C, HHSN268200800007C, N01HC55222, N01HC85079, N01HC85080, N01HC85081, N01HC85082, N01HC85083, N01HC85086, and grants U01HL080295 and U01HL130114 from the NHLBI with additional contribution from the National Institute of Neurological Disorders and Stroke (NINDS). Additional support was provided by R01AG023629, R01AG15928, and R01AG20098 from the NIA. FHS research is supported by NHLBI contracts N01-HC-25195 and HHSN268201500001I. This study was also supported by additional grants from the NIA (R01s AG054076, AG049607 and AG033040 and NINDS (R01 NS017950). The ERF study as a part of EUROSPAN (European Special Populations Research Network) was supported by European Commission FP6 STRP grant number 018947 (LSHG-CT-2006-01947) and also received funding from the European Community's Seventh Framework Programme (FP7/2007-2013)/grant agreement HEALTH-F4- 2007-201413 by the European Commission under the programme "Quality of Life and Management of the Living Resources" of 5th Framework Programme (no. QLG2-CT-2002- 01254). High-throughput analysis of the ERF data was supported by a joint grant

from the Netherlands Organization for Scientific Research and the Russian Foundation for Basic Research (NWO-RFBR 047.017.043). The Rotterdam Study is funded by Erasmus Medical Center and Erasmus University, Rotterdam, the Netherlands Organization for Health Research and Development (ZonMw), the Research Institute for Diseases in the Elderly (RIDE), the Ministry of Education, Culture and Science, the Ministry for Health, Welfare and Sports, the European Commission (DG XII), and the municipality of Rotterdam. Genetic data sets are also supported by the Netherlands Organization of Scientific Research NWO Investments (175.010.2005.011, 911-03-012), the Genetic Laboratory of the Department of Internal Medicine, Erasmus MC, the Research Institute for Diseases in the Elderly (014-93-015; RIDE2), and the Netherlands Genomics Initiative (NGI)/Netherlands Organization for Scientific Research (NWO) Netherlands Consortium for Healthy Aging (NCHA), project 050-060-810. All studies are grateful to their participants, faculty and staff. The content of these manuscripts is solely the responsibility of the authors and does not necessarily represent the official views of the National Institutes of Health or the U.S. Department of Health and Human Services.
The FUS cohorts include: the Alzheimer's Disease Centers (ADC) ( P30 AG019610, P30 AG013846, P50 AG008702, P50 AG025688, P50 AG047266, P30 AG010133, P50 AG005146, P50 AG005134, P50 AG016574, P50 AG005138, P30 AG008051, P30 AG013854, P30 AG008017, P30 AG010161, P50 AG047366, P30 AG010129, P50 AG016573, P50 AG016570, P50 AG005131, P50 AG023501, P30 AG035982, P30 AG028383, P30 AG010124, P50 AG005133, P50 AG005142, P30 AG012300, P50 AG005136, P50 AG033514, P50 AG005681, and P50 AG047270), Alzheimer's Disease Neuroimaging Initiative (ADNI) (U19AG024904), Amish Protective Variant Study (RF1AG058066), Cache County Study (R01AG11380, R01AG031272, R01AG21136, RF1AG054052), Case Western Reserve University Brain Bank (CWRUBB) (P50AG008012), Case Western Reserve University Rapid Decline (CWRURD) (RF1AG058267, NU38CK000480), CubanAmerican Alzheimer's Disease Initiative (CuAADI) (3U01AG052410), Estudio Familiar de Influencia Genetica en Alzheimer (EFIGA) (5R37AG015473, RF1AG015473, R56AG051876), Genetic and Environmental Risk Factors for Alzheimer Disease Among African Americans Study (GenerAAtions) (2R01AG09029, R01AG025259, 2R01AG048927), Gwangju Alzheimer and Related Dementias Study (GARD) (U01AG062602), Hussman Institute for Human Genomics Brain Bank (HIHGBB) (R01AG027944, Alzheimer's Association "Identification of Rare Variants in Alzheimer Disease"), Ibadan Study of Aging (IBADAN) (5R01AG009956), Mexican Health and Aging Study (MHAS) (R01AG018016), Multi-Institutional Research in Alzheimer's Genetic Epidemiology (MIRAGE) (2R01AG09029, R01AG025259, 2R01AG048927), Northern Manhattan Study (NOMAS) (R01NS29993), Peru Alzheimer's Disease Initiative (PeADI) (RF1AG054074), Puerto Rican 1066 (PR1066) (Wellcome Trust (GR066133/GR080002), European Research Council (340755)), Puerto Rican Alzheimer Disease Initiative (PRADI) (RF1AG054074), Reasons for Geographic and Racial Differences in Stroke (REGARDS) (U01NS041588), Research in African American Alzheimer Disease Initiative (REAAADI) (U01AG052410), Rush Alzheimer's Disease Center (ROSMAP) (P30AG10161, R01AG15819, R01AG17919), University of Miami Brain Endowment Bank (MBB), and University of Miami/Case Western/North Carolina A&T African American (UM/CASE/NCAT) (U01AG052410, R01AG028786).
The four LSACs are: the Human Genome Sequencing Center at the Baylor College of Medicine (U54 HG003273), the Broad Institute Genome Center (U54HG003067), The American Genome Center at the Uniformed Services University of the Health Sciences (U01AG057659), and the Washington University Genome Institute (U54HG003079).

Biological samples and associated phenotypic data used in primary data analyses were stored at Study Investigators institutions, and at the National Cell Repository for Alzheimer's Disease (NCRAD, U24AG021886) at Indiana University funded by NIA. Associated Phenotypic Data used in primary and secondary data analyses were provided by Study Investigators, the NIA funded Alzheimer's Disease Centers (ADCs), and the National Alzheimer's Coordinating Center (NACC, U01AG016976) and the National Institute on Aging Genetics of Alzheimer's Disease Data Storage Site (NIAGADS, U24AG041689) at the University of Pennsylvania, funded by NIA This research was supported in part by the Intramural Research Program of the National Institutes of health, National Library of Medicine. Contributors to the Genetic Analysis Data included Study Investigators on projects that were individually funded by NIA, and other NIH institutes, and by private U.S. organizations, or foreign governmental or nongovernmental organizations.
An up to date acknowledgment statement can be found on the ADSP site: https://www.niagads.org/adsp/content/acknowledgement-statement.
Data collection and sharing for this project was funded by the Alzheimer's Disease Neuroimaging Initiative (ADNI) (National Institutes of Health Grant U01 AG024904) and DOD ADNI (Department of Defense award number W81XWH-12-2-0012). ADNI is funded by the National Institute on Aging, the National Institute of Biomedical Imaging and Bioengineering, and through generous contributions from the following: AbbVie, Alzheimer's Association; Alzheimer's Drug Discovery Foundation; Araclon Biotech; BioClinica, Inc.; Biogen; Bristol-Myers Squibb Company; CereSpir, Inc.; Cogstate; Eisai Inc.; Elan Pharmaceuticals, Inc.; Eli Lilly and Company; EuroImmun; F. Hoffmann-La Roche Ltd and its affiliated company Genentech, Inc.; Fujirebio; GE Healthcare; IXICO Ltd.; Janssen Alzheimer Immunotherapy Research & Development, LLC.; Johnson & Johnson Pharmaceutical Research & Development LLC.; Lumosity; Lundbeck; Merck & Co., Inc.; Meso Scale Diagnostics, LLC.; NeuroRx Research; Neurotrack Technologies; Novartis Pharmaceuticals Corporation; Pfizer Inc.; Piramal Imaging; Servier; Takeda Pharmaceutical Company; and Transition Therapeutics. The Canadian Institutes of Health Research is providing funds to support ADNI clinical sites in Canada. Private sector contributions are facilitated by the Foundation for the National Institutes of Health (www.fnih.org). The grantee organization is the Northern California Institute for Research and Education, and the study is coordinated by the Alzheimer's Therapeutic Research Institute at the University of Southern California. ADNI data are disseminated by the Laboratory for Neuro Imaging at the University of Southern California.
Additional information to include in an acknowledgment statement can be found on the LONI site: https://adni.loni.usc.edu/wp-content/uploads/how_to_apply/ADNI_Data_Use_Agreement.pdf.
The Alzheimer's Disease Genetics Consortium (ADGC) supported sample preparation, whole exome sequencing and data processing through NIA grant U01AG032984. Sequencing data generation and harmonization is supported by the Genome Center for Alzheimer's Disease, U54AG052427, and data sharing is supported by NIAGADS, U24AG041689. Samples from the National Centralized Repository for Alzheimer's Disease and Related Dementias (NCRAD), which receives government support under a cooperative agreement grant (U24 AG021886) awarded by the National Institute on Aging (NIA), were used in this study. We thank contributors who collected samples used in this study, as well as patients and their families, whose help and participation made this work possible. NIH grants supported enrollment and data collection for the individual studies including: GenerAAtions R01AG20688 (PI M. Daniele Fallin, PhD); Miami/Duke R01 AG027944, R01 AG028786 (PI Margaret A. Pericak-Vance, PhD); NC A&T

P20 MD000546, R01 AG28786-01A1 (PI Goldie S. Byrd, PhD); Case Western (PI Jonathan L. Haines, PhD); MIRAGE R01 AG009029 (PI Lindsay A. Farrer, PhD); ROS P30AG10161, R01AG15819, R01AG30146, TGen (PI David A. Bennett, MD); MAP R01AG17917, R01AG15819, TGen (PI David A. Bennett, MD). The NACC database is funded by NIA/NIH Grant U01 AG016976. NACC data are contributed by the NIA-funded ADCs: P30 AG019610 (PI Eric Reiman, MD), P30 AG013846 (PI Neil Kowall, MD), P30 AG062428-01 (PI James Leverenz, MD) P50 AG008702 (PI Scott Small, MD), P50 AG025688 (PI Allan Levey, MD, PhD), P50 AG047266 (PI Todd Golde, MD, PhD), P30 AG010133 (PI Andrew Saykin, PsyD), P50 AG005146 (PI Marilyn Albert, PhD), P30 AG062421-01 (PI Bradley Hyman, MD, PhD), P30 AG062422-01 (PI Ronald Petersen, MD, PhD), P50 AG005138 (PI Mary Sano, PhD), P30 AG008051 (PI Thomas Wisniewski, MD), P30 AG013854 (PI Robert Vassar, PhD), P30 AG008017 (PI Jeffrey Kaye, MD), P30 AG010161 (PI David Bennett, MD), P50 AG047366 (PI Victor Henderson, MD, MS), P30 AG010129 (PI Charles DeCarli, MD), P50 AG016573 (PI Frank LaFerla, PhD), P30 AG062429-01(PI James Brewer, MD, PhD), P50 AG023501 (PI Bruce Miller, MD), P30 AG035982 (PI Russell Swerdlow, MD), P30 AG028383 (PI Linda Van Eldik, PhD), P30 AG053760 (PI Henry Paulson, MD, PhD), P30 AG010124 (PI John Trojanowski, MD, PhD), P50 AG005133 (PI Oscar Lopez, MD), P50 AG005142 (PI Helena Chui, MD), P30 AG012300 (PI Roger Rosenberg, MD), P30 AG049638 (PI Suzanne Craft, PhD), P50 AG005136 (PI Thomas Grabowski, MD), P30 AG062715-01 (PI Sanjay Asthana, MD, FRCP), P50 AG005681 (PI John Morris, MD), P50 AG047270 (PI Stephen Strittmatter, MD, PhD).

This work was supported by grants from the National Institutes of Health (R01AG044546, P01AG003991, RF1AG053303, R01AG058501, U01AG058922, RF1AG058501 and R01AG057777). The recruitment and clinical characterization of research participants at Washington University were supported by NIH P50 AG05681, P01 AG03991, and P01 AG026276. This work was supported by access to equipment made possible by the Hope Center for Neurological Disorders, and the Departments of Neurology and Psychiatry at Washington University School of Medicine.

We thank the contributors who collected samples used in this study, as well as patients and their families, whose help and participation made this work possible. Members of the National Institute on Aging Late-Onset Alzheimer Disease/National Cell Repository for Alzheimer Disease (NIA-LOAD NCRAD) Family Study Group include the following: Richard Mayeux, MD, MSc; Martin Farlow, MD; Tatiana Foroud, PhD; Kelley Faber, MS; Bradley F. Boeve, MD; Neill R. Graff-Radford, MD; David A. Bennett, MD; Robert A. Sweet, MD; Roger Rosenberg, MD; Thomas D. Bird, MD; Carlos Cruchaga, PhD; and Jeremy M. Silverman, PhD.

This work was partially supported by grant funding from NIH R01 AG039700 and NIH P50 AG005136. Subjects and samples used here were originally collected with grant funding from NIH U24 AG026395, U24 AG021886, P50 AG008702, P01 AG007232, R37 AG015473, P30 AG028377, P50 AG05128, P50 AG16574, P30 AG010133, P50 AG005681, P01 AG003991, U01MH046281, U01 MH046290 and U01 MH046373. The funders had no role in study design, analysis or preparation of the manuscript. The authors declare no competing interests.

This work was supported by the National Institutes of Health (R01 AG027944, R01 AG028786 to MAPV, R01 AG019085 to JLH, P20 MD000546); a joint grant from the Alzheimer's Association (SG-14-312644) and the Fidelity Biosciences Research Initiative to MAPV; the BrightFocus Foundation (A2011048 to MAPV). NIA-LOAD Family-Based Study supported the collection of samples used in this study through NIH grants U24 AG026395 and R01 AG041797 and the MIRAGE cohort was supported through the NIH grants R01 AG025259 and R01 AG048927. We thank contributors, including the Alzheimer's disease Centers who collected samples used in this study, as well as patients and their families, whose help and participation made this work possible. Study design: HNC, BWK, JLH, MAPV; Sample collection: MLC, JMV, RMC, LAF, JLH, MAPV; Whole exome sequencing and Sanger sequencing: SR, PLW; Sequencing data analysis: HNC, BWK, KLHN, SR, MAK, JRG, ERM, GWB, MAPV; Statistical analysis: BWK, KLHN, JMJ, MAPV; Preparation of manuscript: HNC, BWK. The authors jointly discussed the experimental results throughout the duration of the study. All authors read and approved the final manuscript.

Data collection and sharing for this project was supported by the Washington Heights-Inwood Columbia Aging Project (WHICAP, PO1AG07232, R01AG037212, RF1AG054023) funded by the National Institute on Aging (NIA) and by the National Center for Advancing Translational Sciences, National Institutes of Health, through Grant Number UL1TR001873. This manuscript has been reviewed by WHICAP investigators for scientific content and consistency of data interpretation with previous WHICAP Study publications. We acknowledge the WHICAP study participants and the WHICAP research and support staff for their contributions to this study.

This work was supported by grants from the National Institutes of Health (R01AG044546, P01AG003991, RF1AG053303, R01AG058501, U01AG058922, RF1AG058501 and R01AG057777). The recruitment and clinical characterization of research participants at Washington University were supported by NIH P50 AG05681, P01 AG03991, and P01 AG026276. This work was supported by access to equipment made possible by the Hope Center for Neurological Disorders, and the Departments of Neurology and Psychiatry at Washington University School of Medicine.

We thank the contributors who collected samples used in this study, as well as patients and their families, whose help and participation made this work possible. Members of the National Institute on Aging Late-Onset Alzheimer Disease/National Cell Repository for Alzheimer Disease (NIA-LOAD NCRAD) Family Study Group include the following: Richard Mayeux, MD, MSc; Martin Farlow, MD; Tatiana Foroud, PhD; Kelley Faber, MS; Bradley F. Boeve, MD; Neill R. Graff-Radford, MD; David A. Bennett, MD; Robert A. Sweet, MD; Roger Rosenberg, MD; Thomas D. Bird, MD; Carlos Cruchaga, PhD; and Jeremy M. Silverman, PhD.

This work was supported by grants from the National Institutes of Health (R01AG044546, P01AG003991, RF1AG053303, R01AG058501, U01AG058922, RF1AG058501 and R01AG057777). The recruitment and clinical characterization of research participants at Washington University were supported by NIH P50 AG05681, P01 AG03991, and P01 AG026276. This work was supported by access to equipment made possible by the Hope Center for Neurological Disorders, and the Departments of Neurology and Psychiatry at Washington University School of Medicine.

We thank the contributors who collected samples used in this study, as well as patients and their families, whose help and participation made this work possible. Members of the National Institute on Aging Late-Onset Alzheimer Disease/National Cell Repository for Alzheimer Disease (NIA-LOAD NCRAD) Family Study Group include the following: Richard Mayeux, MD, MSc; Martin Farlow, MD; Tatiana Foroud, PhD; Kelley Faber, MS; Bradley F. Boeve, MD; Neill R. Graff-Radford, MD; David A. Bennett, MD; Robert A. Sweet, MD; Roger Rosenberg, MD; Thomas D. Bird, MD; Carlos Cruchaga, PhD; and Jeremy M. Silverman, PhD.

Mayo RNAseq Study- Study data were provided by the following sources: The Mayo Clinic Alzheimer's Disease Genetic Studies, led by Dr. Nilufer Ertekin-Taner and Dr. Steven G. Younkin, Mayo Clinic, Jacksonville, FL using samples from the Mayo Clinic Study of Aging, the Mayo

Clinic Alzheimer's Disease Research Center, and the Mayo Clinic Brain Bank. Data collection was supported through funding by NIA grants P50 AG016574, R01 AG032990, U01 AG046139, R01 AG018023, U01 AG006576, U01 AG006786, R01 AG025711, R01 AG017216, R01 AG003949, NINDS grant R01 NS080820, CurePSP Foundation, and support from Mayo Foundation. Study data includes samples collected through the Sun Health Research Institute Brain and Body Donation Program of Sun City, Arizona. The Brain and Body Donation Program is supported by the National Institute of Neurological Disorders and Stroke (U24 NS072026 National Brain and Tissue Resource for Parkinson's Disease and Related Disorders), the National Institute on Aging (P30 AG19610 Arizona Alzheimer's Disease Core Center), the Arizona Department of Health Services (contract 211002, Arizona Alzheimer's Research Center), the Arizona Biomedical Research Commission (contracts 4001, 0011, 05-901 and 1001 to the Arizona Parkinson's Disease Consortium) and the Michael J. Fox Foundation for Parkinson's Research

ROSMAP- We are grateful to the participants in the Religious Order Study, the Memory and Aging Project. This work is supported by the US National Institutes of Health [U01 AG046152, R01 AG043617, R01 AG042210, R01 AG036042, R01 AG036836, R01 AG032990, R01 AG18023, RC2 AG036547, P50 AG016574, U01 ES017155, KL2 RR024151, K25 AG041906-01, R01 AG30146, P30 AG10161, R01 AG17917, R01 AG15819, K08 AG034290, P30 AG10161 and R01 AG11101.

Mount Sinai Brain Bank (MSBB)- This work was supported by the grants R01AG046170, RF1AG054014, RF1AG057440 and R01AG057907 from the NIH/National Institute on Aging (NIA). R01AG046170 is a component of the AMP-AD Target Discovery and Preclinical Validation Project. Brain tissue collection and characterization was supported by NIH HHSN271201300031C.

This study was supported by the National Institute on Aging (NIA) grants AG030653, AG041718, AG064877 and P30-AG066468.

We would like to thank study participants, their families, and the sample collectors for their invaluable contributions. This research was supported in part by the National Institute on Aging grant U01AG049508 (PI Alison M. Goate). This research was supported in part by Genentech, Inc. (PI Alison M. Goate, Robert R. Graham).

The NACC database is funded by NIA/NIH Grant U01 AG016976. NACC data are contributed by these NIA-funded ADCs: P30 AG013846 (PI Neil Kowall, MD), P50 AG008702 (PI Scott Small, MD), P50 AG025688 (PI Allan Levey, MD, PhD), P30 AG010133 (PI Andrew Saykin, PsyD), P50 AG005146 (PI Marilyn Albert, PhD), P50 AG005134 (PI Bradley Hyman, MD, PhD), P50 AG016574 (PI Ronald Petersen, MD, PhD), P30 AG013854 (PI M. Marsel Mesulam, MD), P30 AG008017 (PI Jeffrey Kaye, MD), P30 AG010161 (PI David Bennett, MD), P30 AG010129 (PI Charles DeCarli, MD), P50 AG016573 (PI Frank LaFerla, PhD), P50 AG005131 (PI Douglas Galasko, MD), P30 AG028383 (PI Linda Van Eldik, PhD), P30 AG010124 (PI John Trojanowski, MD, PhD), P50 AG005142 (PI Helena Chui, MD), P30 AG012300 (PI Roger Rosenberg, MD), P50 AG005136 (PI Thomas Grabowski, MD), P50 AG005681 (PI John Morris, MD), P30 AG028377 (Kathleen Welsh-Bohmer, PhD), and P50 AG008671 (PI Henry Paulson, MD, PhD).

Samples from the National Cell Repository for Alzheimer’s Disease (NCRAD), which receives government support under a cooperative agreement grant (U24 AG21886) awarded by the National Institute on Aging (NIA), were used in this study. We thank contributors who collected samples used in this study, as well as patients and their families, whose help and participation made this work possible.

The Alzheimer's Disease Genetics Consortium supported the collection of samples used in this study through National Institute on Aging (NIA) grants U01AG032984 and RC2AG036528.
We acknowledge the generous contributions of the Cache County Memory Study participants. Sequencing for this study was funded by RF1AG054052 (PI: John S.K. Kauwe)

#### Acknowledgments for the use of GWAS data distributed by NIAGADS

The NIA Genetics of Alzheimer's Disease Data Storage Site (NIAGADS) is supported by a collaborative agreement from the National Institute on Aging, U24AG041689.
NG00047: The NIA supported this work through grants U01-AG032984, RC2-AG036528, U01-AG016976 (Dr Kukull); U24 AG026395, U24 AG026390, R01AG037212, R37 AG015473 (Dr Mayeux); K23AG034550 (Dr Reitz); U24-AG021886 (Dr Foroud); R01AG009956, RC2 AG036650 (Dr Hall); UO1 AG06781, UO1 HG004610 (Dr Larson); R01 AG009029 (Dr Farrer); 5R01AG20688 (Dr Fallin); P50 AG005133, AG030653 (Dr Kamboh); R01 AG019085 (Dr Haines); R01 AG1101, R01 AG030146, RC2 AG036650 (Dr Evans); P30AG10161, R01AG15819, R01AG30146, R01AG17917, R01AG15819 (Dr Bennett); R01AG028786 (Dr Manly); R01AG22018, P30AG10161 (Dr Barnes); P50AG16574 (Dr Ertekin-Taner, Dr Graff-Radford), R01 AG032990 (Dr Ertekin-Taner), KL2 RR024151 (Dr Ertekin-Taner); R01 AG027944, R01 AG028786 (Dr Pericak-Vance); P20 MD000546, R01 AG28786-01A1 (Dr Byrd); AG005138 (Dr Buxbaum); P50 AG05681, P01 AG03991, P01 AG026276 (Dr Goate); and P30AG019610, P30AG13846, U01-AG10483, R01CA129769, R01MH080295, R01AG017173, R01AG025259, R01AG33193, P50AG008702, P30AG028377, AG05128, AG025688, P30AG10133, P50AG005146, P50AG005134, P01AG002219, P30AG08051, MO1RR00096, UL1RR029893, P30AG013854, P30AG008017, R01AG026916, R01AG019085, P50AG016582, UL1RR02777, R01AG031581, P30AG010129, P50AG016573, P50AG016575, P50AG016576, P50AG016577, P50AG016570, P50AG005131, P50AG023501, P50AG019724, P30AG028383, P50AG008671, P30AG010124, P50AG005142, P30AG012300, AG010491, AG027944, AG021547, AG019757, P50AG005136 (Alzheimer Disease GeneticsConsortium [ADGC]). We thank Creighton Phelps, Stephen Synder, and Marilyn Miller from the NIA, who are ex-officio members of the ADGC. Support was also provided by the Alzheimer's Association (IIRG-08-89720 [Dr Farrer] and IIRG-05-14147 [Dr Pericak-Vance]), National Institute of Neurological Disorders and Stroke grant NS39764, National Institute of Mental Health grant MH60451, GlaxoSmithKline, and the Office of Research and Development, Biomedical Laboratory Research Program, US Department of Veterans Affairs Administration. For the ADGC, biological samples and associated phenotypic data used in primary data analyses were stored at principal investigators' institutions and at the National Cell Repository for Alzheimer's Disease (NCRAD) at Indiana University, funded by the NIA. Associated phenotypic data used in secondary data analyses were stored at the National Alzheimer's Coordinating Center and at the NIA Alzheimer's Disease Data Storage Site at the University of Pennsylvania, funded by the NIA. Contributors to the genetic analysis data included principal investigators on projects ndividually funded by the NIA, other NIH institutes, or private entities.

#### Acknowledgments for other GWAS and phenotype data

##### *NACC*

The NACC database is funded by NIA/NIH Grant U01 AG016976. NACC data are contributed by the NIA-funded ADCs: P30 AG019610 (PI Eric Reiman, MD), P30 AG013846 (PI Neil Kowall, MD), P30 AG062428-01 (PI James Leverenz, MD) P50 AG008702 (PI Scott Small, MD), P50 AG025688 (PI Allan Levey, MD, PhD), P50 AG047266 (PI Todd Golde, MD, PhD), P30 AG010133 (PI Andrew Saykin, PsyD), P50 AG005146 (PI Marilyn Albert, PhD), P30 AG062421-01 (PI Bradley Hyman, MD, PhD), P30 AG062422-01 (PI Ronald Petersen, MD, PhD), P50 AG005138 (PI Mary Sano, PhD), P30 AG008051 (PI Thomas Wisniewski, MD), P30 AG013854 (PI Robert Vassar, PhD), P30 AG008017 (PI Jeffrey Kaye, MD), P30 AG010161 (PI David Bennett, MD), P50 AG047366 (PI Victor Henderson, MD, MS), P30 AG010129 (PI Charles DeCarli, MD), P50 AG016573 (PI Frank LaFerla, PhD), P30 AG062429-01(PI James Brewer, MD, PhD), P50 AG023501 (PI Bruce Miller, MD), P30 AG035982 (PI Russell Swerdlow, MD), P30 AG028383 (PI Linda Van Eldik, PhD), P30 AG053760 (PI Henry Paulson, MD, PhD), P30 AG010124 (PI John Trojanowski, MD, PhD), P50 AG005133 (PI Oscar Lopez, MD), P50 AG005142 (PI Helena Chui, MD), P30 AG012300 (PI Roger Rosenberg, MD), P30 AG049638 (PI Suzanne Craft, PhD), P50 AG005136 (PI Thomas Grabowski, MD), P30 AG062715-01 (PI Sanjay Asthana, MD, FRCP), P50 AG005681 (PI John Morris, MD), P50 AG047270 (PI Stephen Strittmatter, MD, PhD).

##### *MARS & LATC*

We thank all Minority Aging Research Study and Latino Core participants and the Rush Alzheimer's Disease Center staff. This database was funded by the NIH/NIA grants R01AG22018 (MARS) and P30AG 072975 (ADC).

##### *GenADA*

The genotypic and associated phenotypic data used in the study "Multi-Site Collaborative Study for Genotype-Phenotype Associations in Alzheimer's Disease (GenADA)" were provided by the GlaxoSmithKline, R&D Limited.

##### *ROSMAP*

ROSMAP study data were provided by the Rush Alzheimer's Disease Center, Rush University Medical Center, Chicago. Data collection was supported through funding by NIA grants P30AG10161, R01AG15819, R01AG17917, R01AG30146, R01AG36836, U01AG32984, U01AG46152, the Illinois Department of Public Health, and the Translational Genomics Research Institute.

##### *AddNeuroMed*

The AddNeuroMed data are from a public-private partnership supported by EFPIA companies and SMEs as part of InnoMed (Innovative Medicines in Europe), an Integrated Project funded by the European Union of the Sixth Framework program priority FP6-2004-LIFESCIHEALTH-5. Clinical leads responsible for data collection are Iwona Kłoszewska (Lodz), Simon Lovestone (London), Patrizia Mecocci (Perugia), Hilkka Soininen (Kuopio), Magda Tsolaki (Thessaloniki), and Bruno Vellas (Toulouse), imaging leads are Andy Simmons (London), Lars-Olad Wahlund (Stockholm) and Christian Spenger (Zurich) and bioinformatics leads are Richard Dobson (London) and Stephen Newhouse (London).

##### *ADNI*

Data collection and sharing for this project was funded by the Alzheimer's Disease Neuroimaging Initiative (ADNI) (National Institutes of Health Grant U01 AG024904) and DOD ADNI (Department of Defense award number W81XWH-12-2-0012). ADNI is funded by the National Institute on Aging, the National Institute of Biomedical Imaging and Bioengineering and through generous contributions from the following: AbbVie. Alzheimer's Association; Alzheimer's Drug Discovery Foundation; Araclon Biotech; BioClinica. Inc.; Biogen; Bristol-Myers Squibb Company; CereSpir. Inc.; Cogstate; Eisai Inc.; Elan Pharmaceuticals. Inc.; Eli Lilly and Company; EuroImmun; F. Hoffmann-La Roche Ltd and its affiliated company Genentech. Inc.; Fujirebio; GE HealtControlsare; IXICO Ltd.; Janssen Alzheimer Immunotherapy Research & Development. LLC.; Johnson & Johnson Pharmaceutical Research & Development LLC.; Lumosity; Lundbeck; Merck & Co. Inc.; Meso Scale Diagnostics. LLC.; NeuroRx Research; Neurotrack Technologies; Novartis Pharmaceuticals Corporation; Pfizer Inc.; Piramal Imaging; Servier; Takeda Pharmaceutical Company; and Transition Therapeutics. The Canadian Institutes of Health Research is providing funds to support ADNI clinical sites in Canada. Private sector contributions are facilitated by the Foundation for the National Institutes of Health. The grantee organization is the Northern California Institute for Research and Education, and the study is coordinated by the Alzheimer's Therapeutic Research Institute at the University of Southern California. ADNI data are disseminated by the Laboratory for Neuro Imaging at the University of Southern California.

##### *NCRAD*

Biological samples used in this study were stored at study investigators' institutions and at the National Cell Repository for Alzheimer's Disease (NCRAD) at Indiana University, which receives government support under a cooperative agreement grant (U24 AG21886) awarded by the National Institute on Aging (NIA). We thank contributors who collected samples used in this study, as well as patients and their families, whose help and participation made this work possible.

##### *UK Biobank*

UK Biobank data were analyzed under Application Number 45420.

##### *FinnGen Study*

We want to acknowledge the participants and investigators of the FinnGen study. The FinnGen project is funded by two grants from Business Finland (HUS 4685/31/2016 and UH 4386/31/2016) and the following industry partners: AbbVie Inc., AstraZeneca UK Ltd, Biogen MA Inc., Bristol Myers Squibb Inc. (and Celgene Corporation & Celgene International II Sàrl), Genentech Inc., Merck Sharp & Dohme LCC, Pfizer Inc., GlaxoSmithKline Intellectual Property Development Ltd., Sanofi US Services Inc., Maze Therapeutics Inc., Johnson&Johnson Innovative Medicine Inc., Novartis AG, Boehringer Ingelheim International GmbH and Bayer AG. Following biobanks are acknowledged for delivering biobank samples to FinnGen: Auria Biobank (www.auria.fi/biopankki), THL Biobank (www.thl.fi/biobank), Helsinki Biobank (www.helsinginbiopankki.fi), Biobank Borealis of Northern Finland (https://www.ppshp.fi/Tutkimus-ja-opetus/Biopankki/Pages/Biobank-Borealis-briefly-in-English.aspx), Finnish Clinical Biobank Tampere (www.tays.fi/en-US/Research_and_development/Finnish_Clinical_Biobank_Tampere), Biobank of Eastern Finland (www.ita-suomenbiopankki.fi/en), Central Finland Biobank (www.ksshp.fi/fi-FI/Potilaalle/Biopankki), Finnish Red Cross Blood Service Biobank (www.veripalvelu.fi/verenluovutus/biopankkitoiminta), Terveystalo Biobank (www.terveystalo.com/fi/Yritystietoa/Terveystalo-Biopankki/Biopankki/) and Arctic Biobank (https://www.oulu.fi/en/university/faculties-and-units/faculty-medicine/northern-finland-birth-cohorts-and-arctic-biobank). All Finnish Biobanks are members of BBMRI.fi infrastructure (https://www.bbmri-eric.eu/national-nodes/finland/). Finnish Biobank Cooperative -FINBB (https://finbb.fi/) is the coordinator of BBMRI-ERIC operations in Finland. The Finnish biobank data can be accessed through the Fingenious® services (https://site.fingenious.fi/en/) managed by FINBB.

#### FinnGen ethics statement

Study subjects in FinnGen provided informed consent for biobank research, based on the Finnish Biobank Act. Alternatively, separate research cohorts, collected prior the Finnish Biobank Act came into effect (in September 2013) and start of FinnGen (August 2017), were collected based on study-specific consents and later transferred to the Finnish biobanks after approval by Fimea (Finnish Medicines Agency), the National Supervisory Authority for Welfare and Health. Recruitment protocols followed the biobank protocols approved by Fimea. The Coordinating Ethics Committee of the Hospital District of Helsinki and Uusimaa (HUS) statement number for the FinnGen study is Nr HUS/990/2017.

The FinnGen study is approved by Finnish Institute for Health and Welfare (permit numbers: THL/2031/6.02.00/2017, THL/1101/5.05.00/2017, THL/341/6.02.00/2018, THL/2222/6.02.00/2018, THL/283/6.02.00/2019, THL/1721/5.05.00/2019 and THL/1524/5.05.00/2020), Digital and population data service agency (permit numbers: VRK43431/2017-3, VRK/6909/2018-3, VRK/4415/2019-3), the Social Insurance Institution (permit numbers: KELA 58/522/2017, KELA 131/522/2018, KELA 70/522/2019, KELA 98/522/2019, KELA 134/522/2019, KELA 138/522/2019, KELA 2/522/2020, KELA 16/522/2020), Findata permit numbers THL/2364/14.02/2020, THL/4055/14.06.00/2020, THL/3433/14.06.00/2020, THL/4432/14.06/2020, THL/5189/14.06/2020, THL/5894/14.06.00/2020, THL/6619/14.06.00/2020, THL/209/14.06.00/2021, THL/688/14.06.00/2021, THL/1284/14.06.00/2021, THL/1965/14.06.00/2021, THL/5546/14.02.00/2020, THL/2658/14.06.00/2021, THL/4235/14.06.00/2021, Statistics Finland (permit numbers: TK-53-1041-17 and TK/143/07.03.00/2020 (earlier TK-53-90-20) TK/1735/07.03.00/2021, TK/3112/07.03.00/2021) and Finnish Registry for Kidney Diseases permission/extract from the meeting minutes on 4th July 2019.

The Biobank Access Decisions for FinnGen samples and data utilized in FinnGen Data Freeze 12 include: THL Biobank BB2017_55, BB2017_111, BB2018_19, BB_2018_34, BB_2018_67, BB2018_71, BB2019_7, BB2019_8, BB2019_26, BB2020_1, BB2021_65, Finnish Red Cross Blood Service Biobank 7.12.2017, Helsinki Biobank HUS/359/2017, HUS/248/2020, HUS/430/2021 §28, §29, HUS/150/2022 §12, §13, §14, §15, §16, §17, §18, §23, §58, §59, HUS/128/2023 §18, Auria Biobank AB17-5154 and amendment #1 (August 17 2020) and amendments BB_2021-0140, BB_2021-0156 (August 26 2021, Feb 2 2022), BB_2021-0169,

BB_2021-0179, BB_2021-0161, AB20-5926 and amendment #1 (April 23 2020) and it´s modifications (Sep 22 2021), BB_2022-0262, BB_2022-0256, Biobank Borealis of Northern Finland_2017_1013, 2021_5010, 2021_5010 Amendment, 2021_5018, 2021_5018 Amendment, 2021_5015, 2021_5015 Amendment, 2021_5015 Amendment_2, 2021_5023, 2021_5023 Amendment, 2021_5023 Amendment_2, 2021_5017, 2021_5017 Amendment, 2022_6001, 2022_6001 Amendment, 2022_6006 Amendment, 2022_6006 Amendment, 2022_6006 Amendment_2, BB22-0067, 2022_0262, 2022_0262 Amendment, Biobank of Eastern Finland 1186/2018 and amendment 22§/2020, 53§/2021, 13§/2022, 14§/2022, 15§/2022, 27§/2022, 28§/2022, 29§/2022, 33§/2022, 35§/2022, 36§/2022, 37§/2022, 39§/2022, 7§/2023, 32§/2023, 33§/2023, 34§/2023, 35§/2023, 36§/2023, 37§/2023, 38§/2023, 39§/2023, 40§/2023, 41§/2023, Finnish Clinical Biobank Tampere MH0004 and amendments (21.02.2020 & 06.10.2020), BB2021-0140 8§/2021, 9§/2021, §9/2022, §10/2022, §12/2022, 13§/2022, §20/2022, §21/2022, §22/2022, §23/2022, 28§/2022, 29§/2022, 30§/2022, 31§/2022, 32§/2022, 38§/2022, 40§/2022, 42§/2022, 1§/2023, Central Finland Biobank 1-2017, BB_2021-0161, BB_2021-0169, BB_2021-0179, BB_2021-0170, BB_2022-0256, BB_2022-0262, BB22-0067, Decision allowing to continue data processing until 31st Aug 2024 for projects: BB_2021-0179, BB22-0067,BB_2022-0262, BB_2021-0170, BB_2021-0164, BB_2021-0161, and BB_2021-0169, and Terveystalo Biobank STB 2018001 and amendment 25th Aug 2020, Finnish Hematological Registry and Clinical Biobank decision 18th June 2021, Arctic biobank P0844: ARC_2021_1001.

### Supplementary Figures

### GWAS

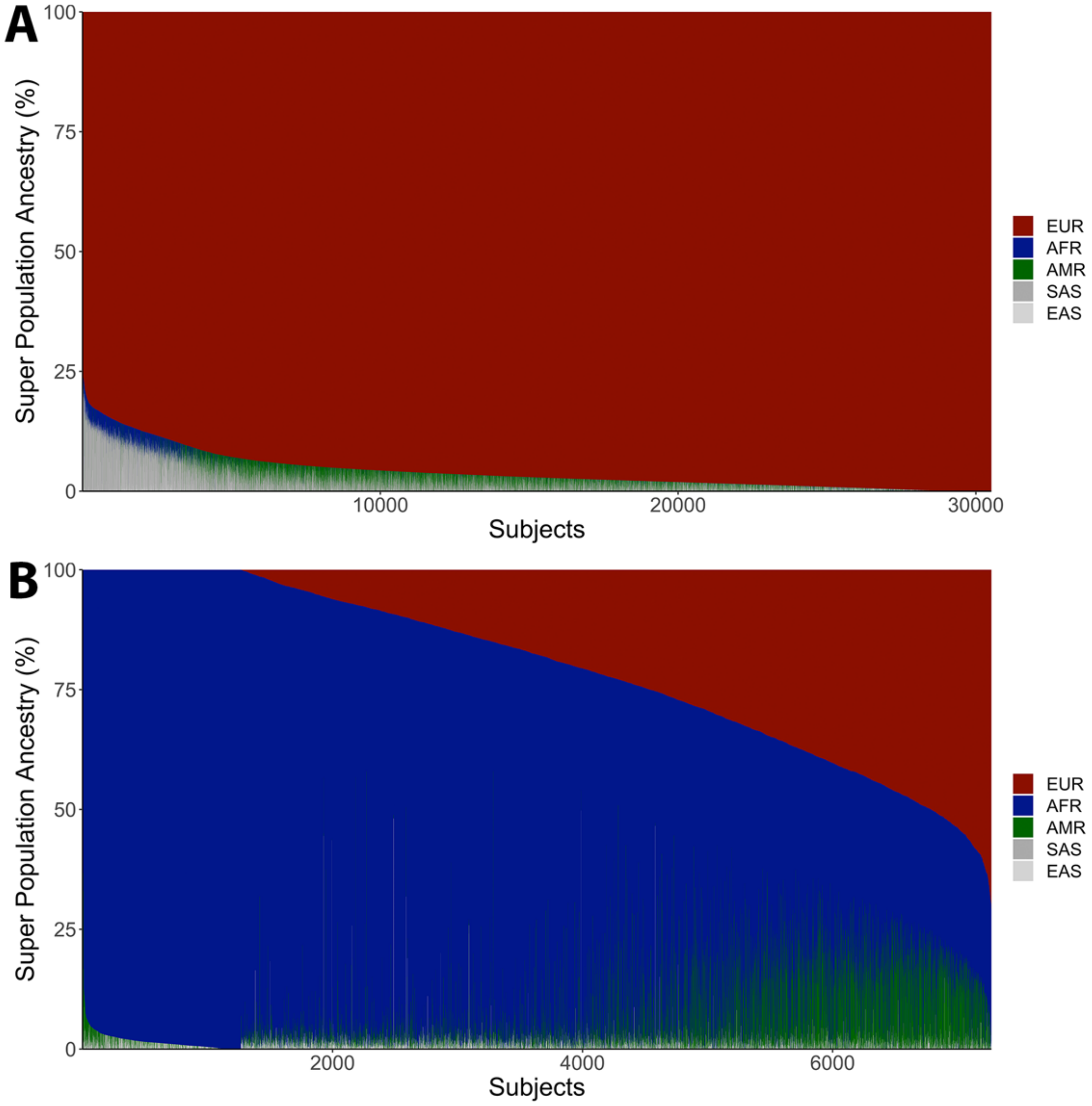

**Supplementary Figure 1. Admixture plot across the five major super populations, for case-control participants included in ADGC and ADSP. A)** Non-Hispanic White European ancestry. **B)** Non-Hispanic and Hispanic admixed African ancestry.

*Abbreviations: EUR, European; AFR, African; AMR; Amerindian; SAS, South Asian; EAS; East Asian.*

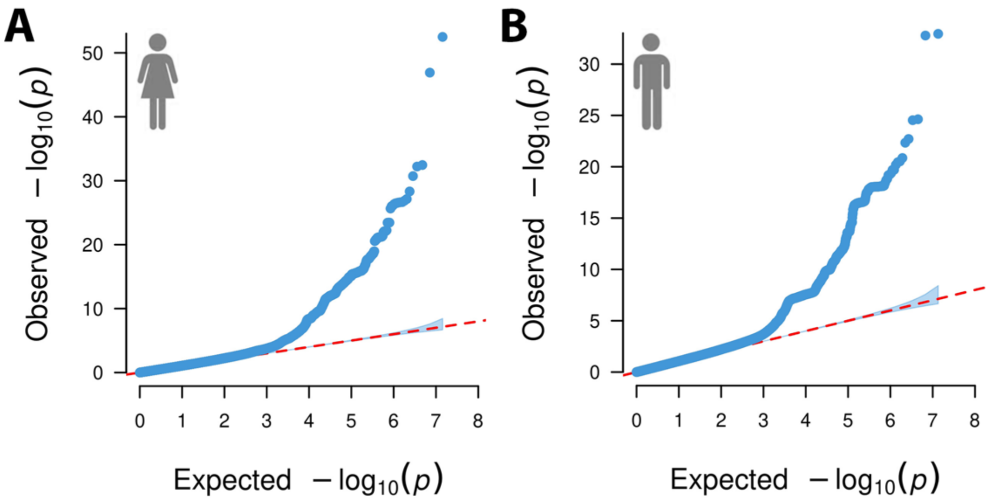

**Supplementary Figure 2. Quantile-Quantile (QQ) plots corresponding to primary sex-stratified GWAS of Alzheimer's disease. A)** Women. The inflation factor ($\lambda$=1.0978) and sample size-adjusted inflation factor ($\lambda_{1,000}$=1.0002) showed no sign of inflation. **B)** Men. The inflation factor ($\lambda$=1.1000) and sample size-adjusted inflation factor ($\lambda_{1,000}$=1.0002) showed no sign of inflation.

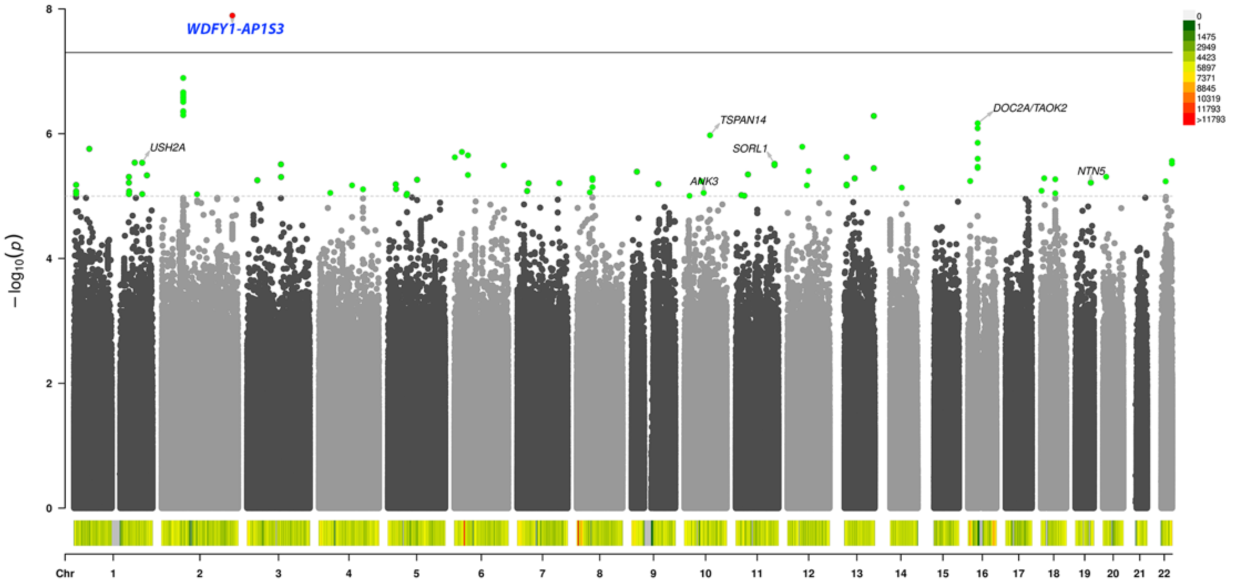

**Supplementary Figure 3. Sex heterogeneity GWAS of Alzheimer's disease.** The sole, novel genome-wide significant association (*P*<5e-8, red dots, solid black line) is marked in blue text, while signals passing suggestive significance *(P*<1e-5, green dots, dashed gray line) in known Alzheimer's disease loci are marked in black text. The color histogram at the bottom of the plot indicates variant density (cf. color scale legend).

## S4.1

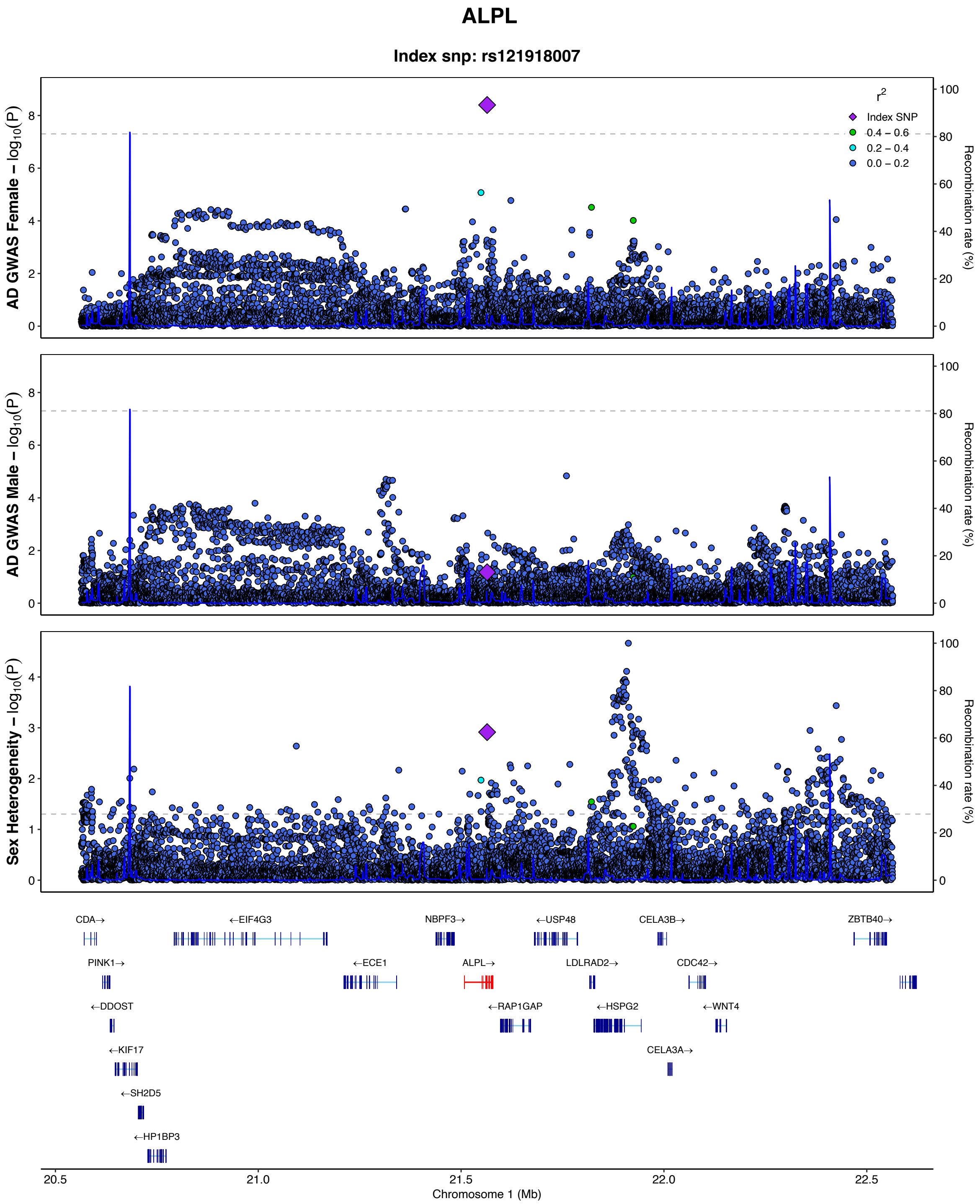

ALPL
Index snp: rs121918007
r2
Index SNP
0.4 – 0.6
0.2 – 0.4
0.0 – 0.2
AD GWAS Female – log10(P)
AD GWAS Male – log10(P)
Sex Heterogeneity – log10(P)
Recombination rate (%)
CDA→
←EIF4G3
NBPF3→
←USP48
CELA3B→
ZBTB40→
PINK1→
←ECE1
ALPL→
LDLRAD2→
CDC42→
←DDOST
←RAP1GAP
←HSPG2
←WNT4
←KIF17
CELA3A→
←SH2D5
←HP1BP3
20.5
21.0
21.5
22.0
22.5
Chromosome 1 (Mb)

## S4.2

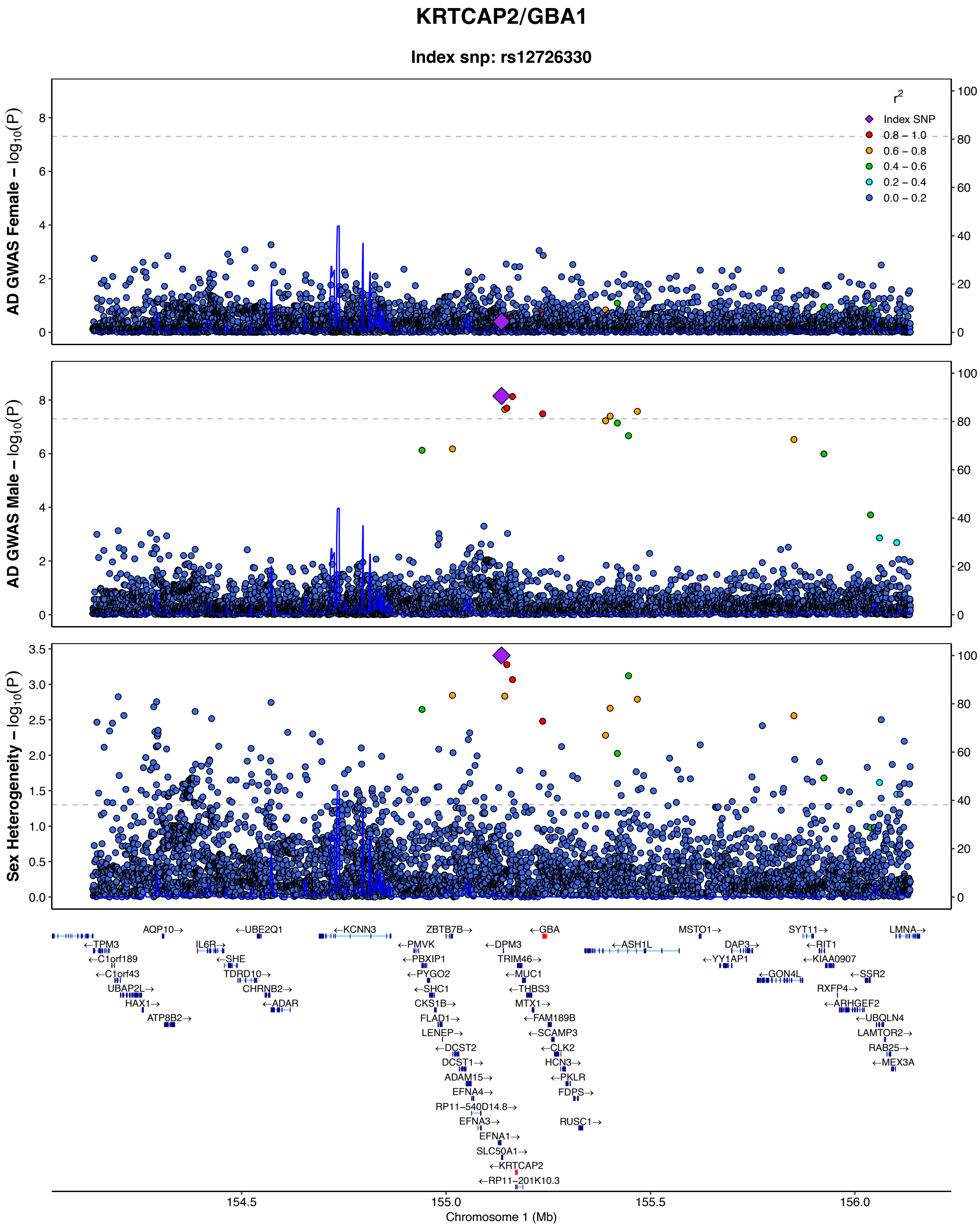
KRTCAP2/GBA1
Index snp: rs12726330
AD GWAS Female – log10(P)
AD GWAS Male – log10(P)
Sex Heterogeneity – log10(P)
Recombination rate (%)
r2
Index SNP
0.8 – 1.0
0.6 – 0.8
0.4 – 0.6
0.2 – 0.4
0.0 – 0.2
Chromosome 1 (Mb)
154.5
155.0
155.5
156.0

## S4.3

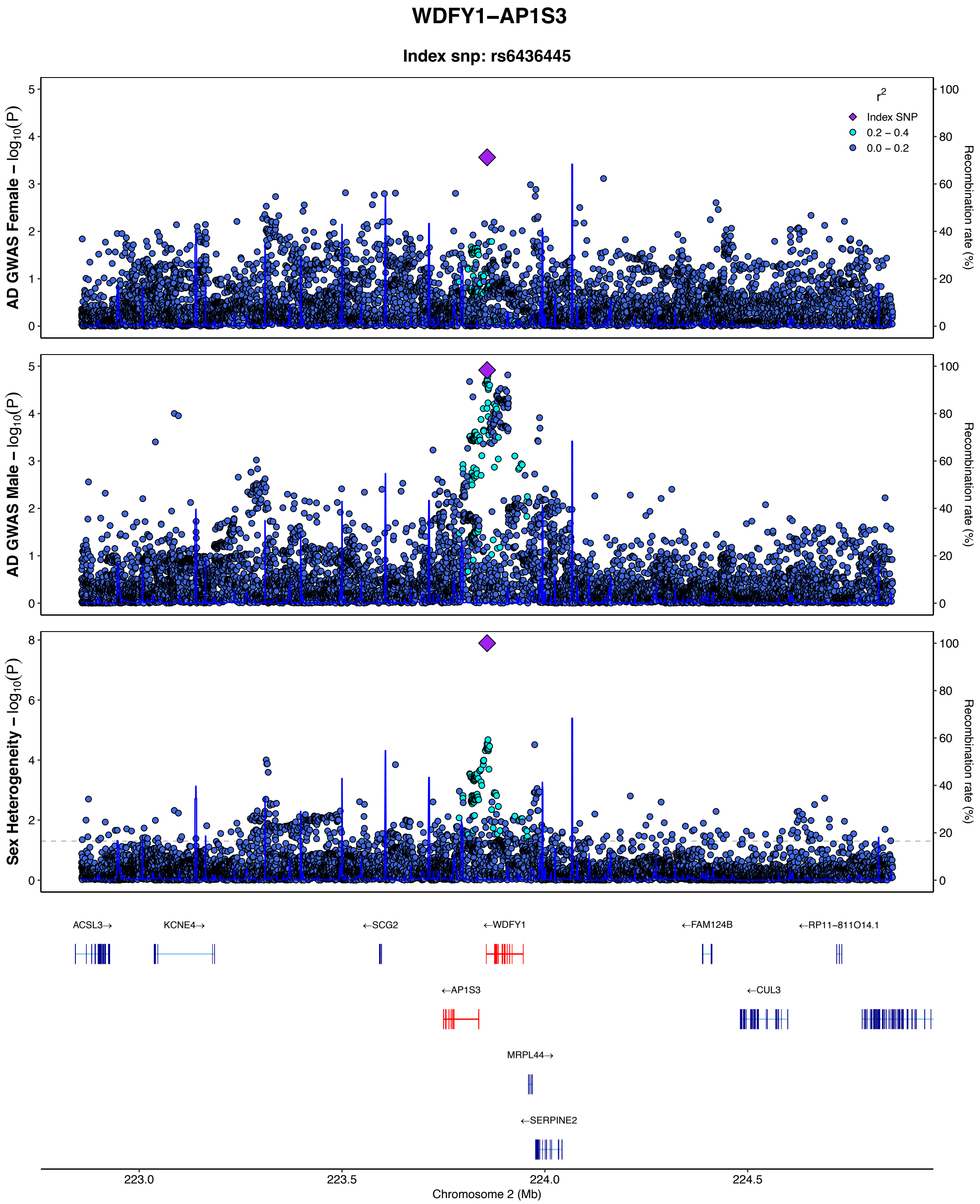

WDFY1−AP1S3
Index snp: rs6436445
AD GWAS Female − log10(P)
AD GWAS Male − log10(P)
Sex Heterogeneity − log10(P)
Recombination rate (%)
r2
Index SNP
0.2 − 0.4
0.0 − 0.2
ACSL3→
KCNE4→
←SCG2
←WDFY1
←FAM124B
←RP11−811O14.1
←AP1S3
←CUL3
MRPL44→
←SERPINE2
223.0
223.5
224.0
224.5
Chromosome 2 (Mb)

## S4.4

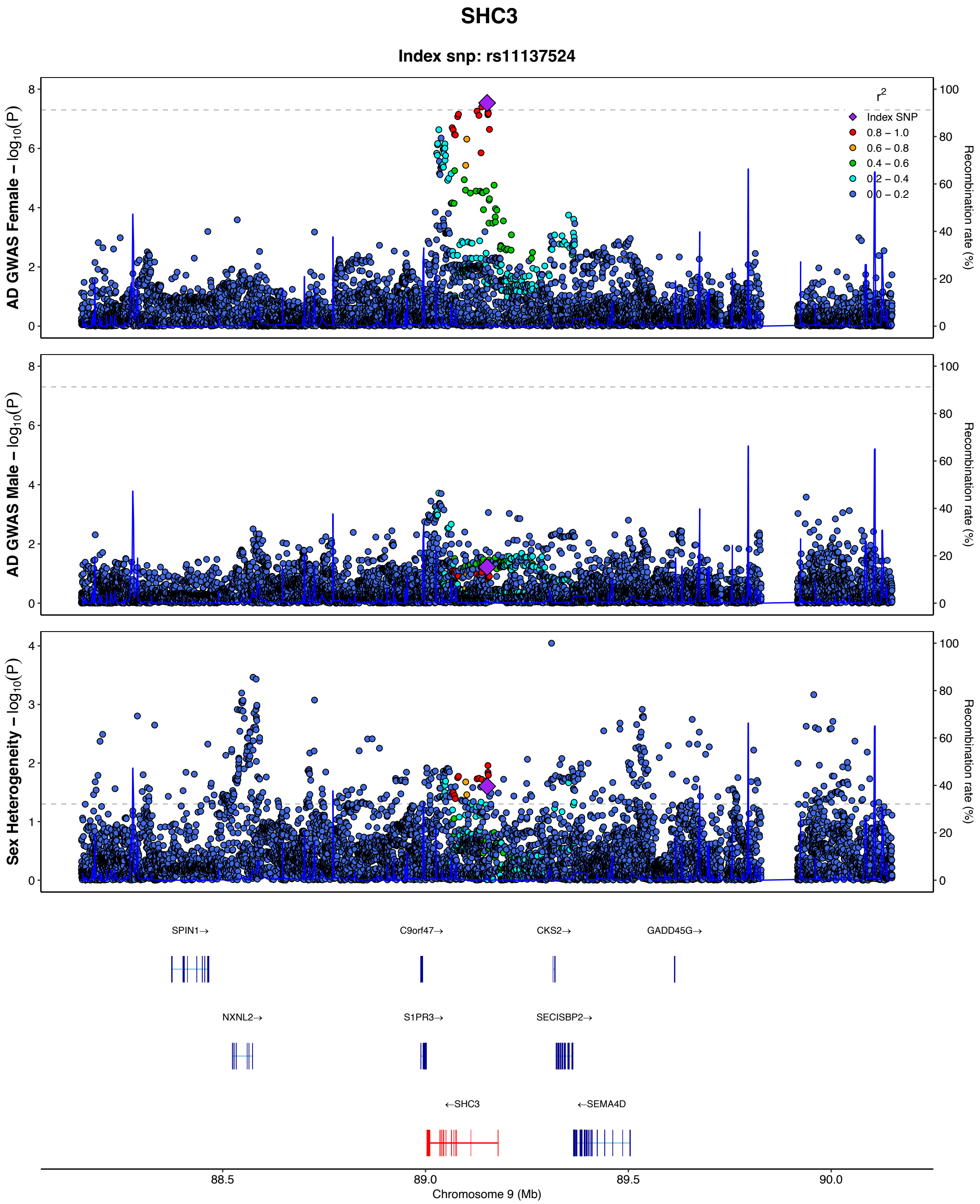
SHC3
Index snp: rs11137524
AD GWAS Female – log10(P)
AD GWAS Male – log10(P)
Sex Heterogeneity – log10(P)
Recombination rate (%)
r2
Index SNP
0.8 – 1.0
0.6 – 0.8
0.4 – 0.6
0.2 – 0.4
0.0 – 0.2
SPIN1→
C9orf47→
CKS2→
GADD45G→
NXNL2→
S1PR3→
SECISBP2→
←SHC3
←SEMA4D
88.5
89.0
89.5
90.0
Chromosome 9 (Mb)

## S4.5

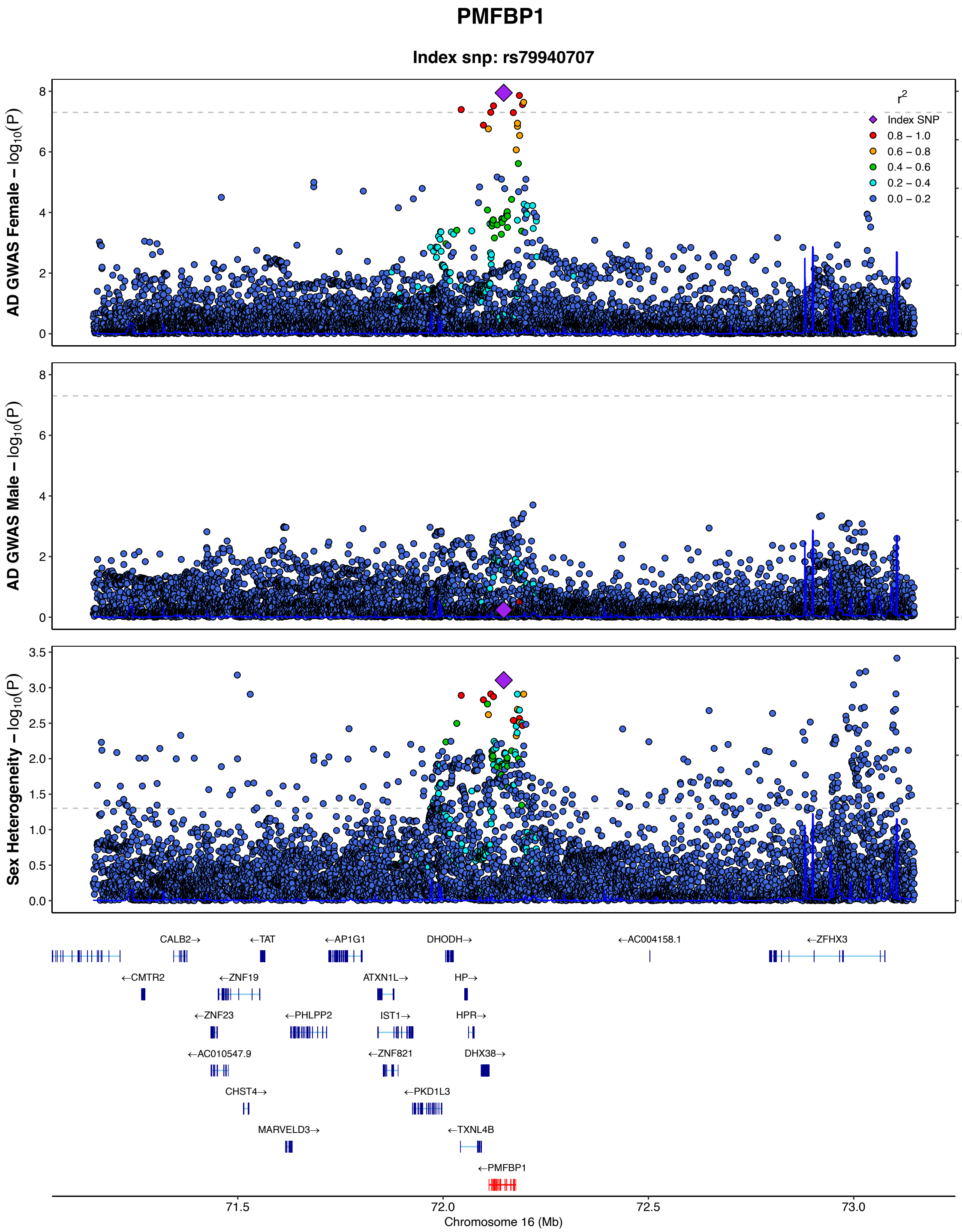

PMFBP1
Index snp: rs79940707
AD GWAS Female – log10(P)
AD GWAS Male – log10(P)
Sex Heterogeneity – log10(P)
Recombination rate (%)
r2
Index SNP
0.8 – 1.0
0.6 – 0.8
0.4 – 0.6
0.2 – 0.4
0.0 – 0.2
CALB2→
←TAT
←AP1G1
DHODH→
←AC004158.1
←ZFHX3
←CMTR2
←ZNF19
ATXN1L→
HP→
←ZNF23
←PHLPP2
IST1→
HPR→
←AC010547.9
←ZNF821
DHX38→
CHST4→
←PKD1L3
MARVELD3→
←TXNL4B
←PMFBP1
71.5
72.0
72.5
73.0
Chromosome 16 (Mb)

## S4.6

##### NCK2

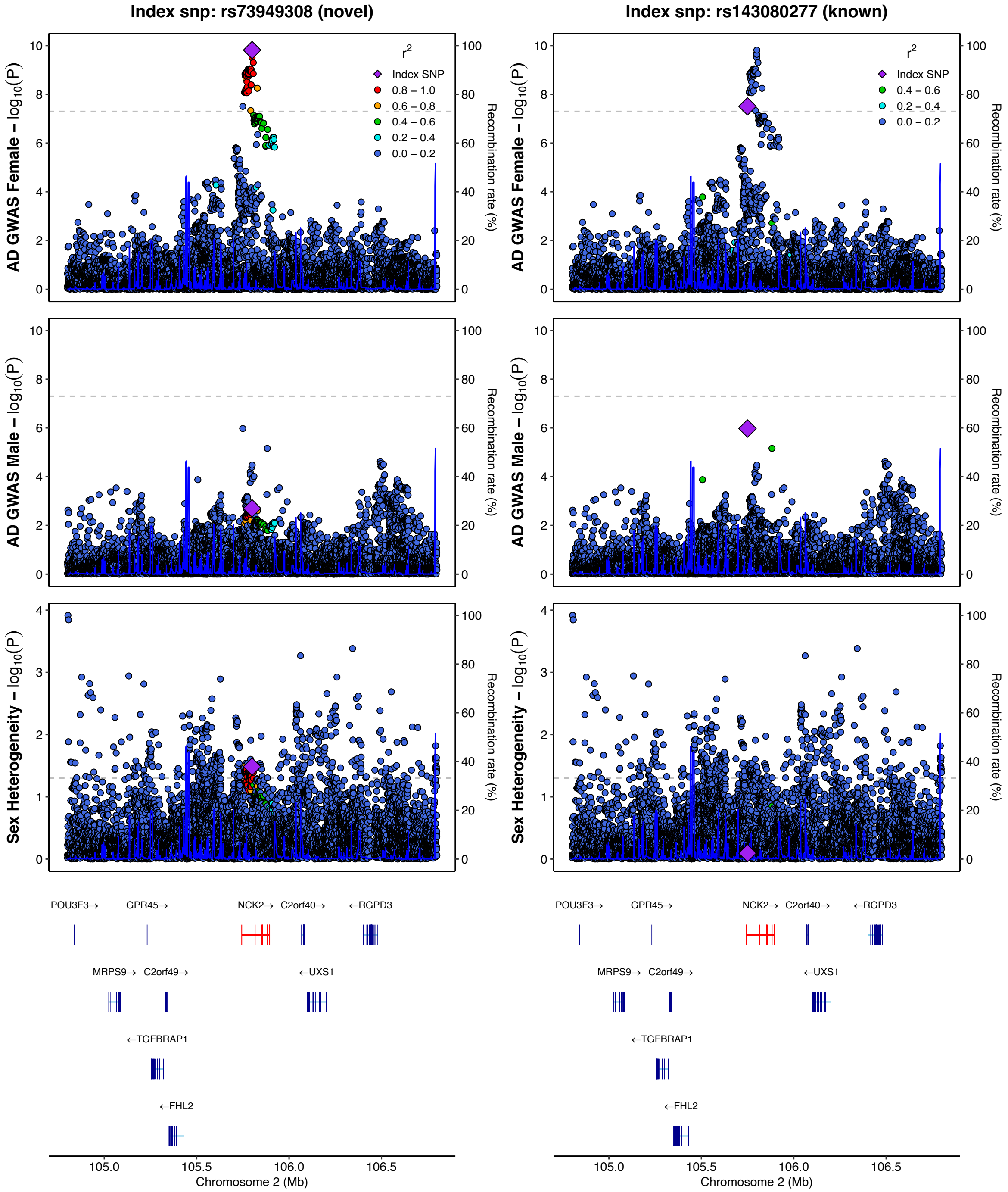

Index snp: rs73949308 (novel)
Index snp: rs143080277 (known)
AD GWAS Female – log10(P)
AD GWAS Male – log10(P)
Sex Heterogeneity – log10(P)
Recombination rate (%)
r2
Index SNP
0.8 – 1.0
0.6 – 0.8
0.4 – 0.6
0.2 – 0.4
0.0 – 0.2
POU3F3→
GPR45→
NCK2→
C2orf40→
←RGPD3
MRPS9→
C2orf49→
←UXS1
←TGFBRAP1
←FHL2
105.0
105.5
106.0
106.5
Chromosome 2 (Mb)

S4.7

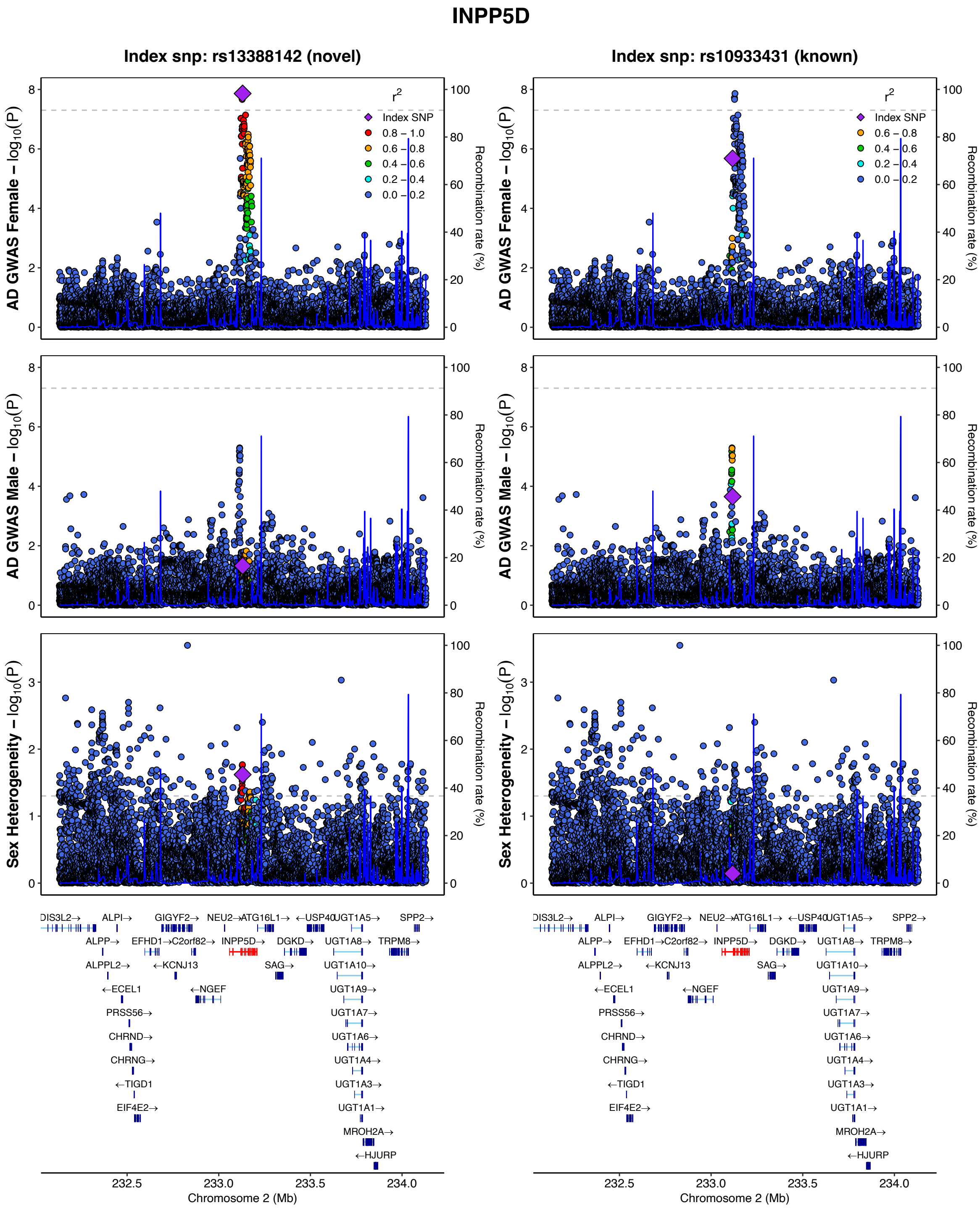
INPP5D
Index snp: rs13388142 (novel)
Index snp: rs10933431 (known)
AD GWAS Female – log10(P)
AD GWAS Male – log10(P)
Sex Heterogeneity – log10(P)
Recombination rate (%)
r2
Index SNP
0.8 – 1.0
0.6 – 0.8
0.4 – 0.6
0.2 – 0.4
0.0 – 0.2
Chromosome 2 (Mb)
232.5
233.0
233.5
234.0
ALPI→
GIGYF2→
NEU2→
ATG16L1→
←USP40
UGT1A5→
SPP2→
ALPP→
EFHD1→
C2orf82→
INPP5D→
DGKD→
UGT1A8→
TRPM8→
ALPPL2→
←KCNJ13
SAG→
UGT1A10→
←ECEL1
←NGEF
UGT1A9→
PRSS56→
UGT1A7→
CHRND→
UGT1A6→
CHRNG→
UGT1A4→
←TIGD1
UGT1A3→
EIF4E2→
UGT1A1→
MROH2A→
←HJURP

**S4.8**

#### RBCK1

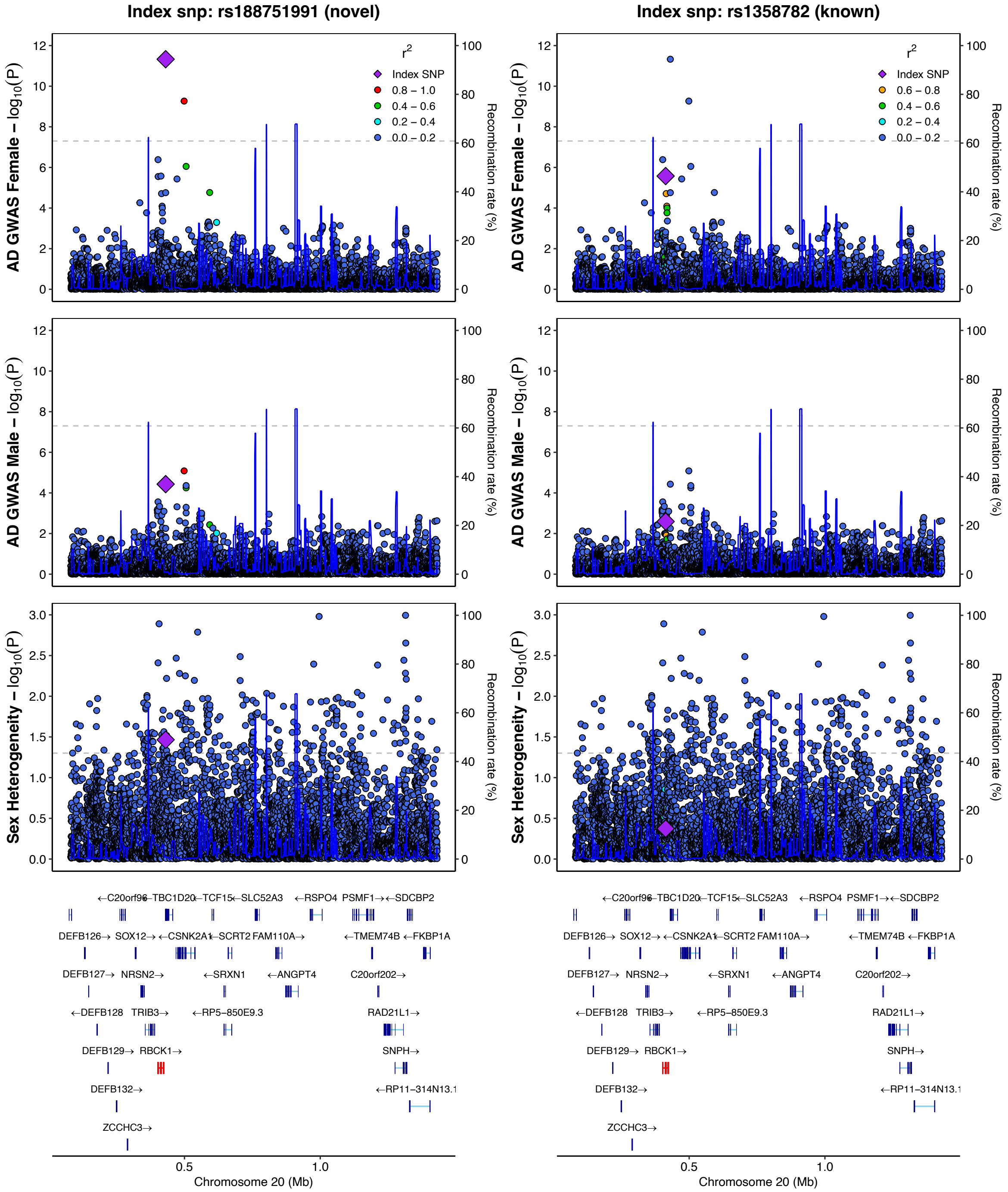

Index snp: rs188751991 (novel)
Index snp: rs1358782 (known)
AD GWAS Female – log10(P)
AD GWAS Male – log10(P)
Sex Heterogeneity – log10(P)
Recombination rate (%)
r2
Index SNP
0.8 – 1.0
0.6 – 0.8
0.4 – 0.6
0.2 – 0.4
0.0 – 0.2
←C20orf96 ←TBC1D20 ←TCF15 ←SLC52A3 ←RSPO4 PSMF1→ ←SDCBP2
DEFB126→ SOX12→ ←CSNK2A1 ←SCRT2 FAM110A→ ←TMEM74B ←FKBP1A
DEFB127→ NRSN2→ ←SRXN1 ←ANGPT4 C20orf202→
←DEFB128 TRIB3→ ←RP5−850E9.3 RAD21L1→
DEFB129→ RBCK1→ SNPH→
DEFB132→ ←RP11−314N13.1
ZCCHC3→
Chromosome 20 (Mb)

**S4.9**

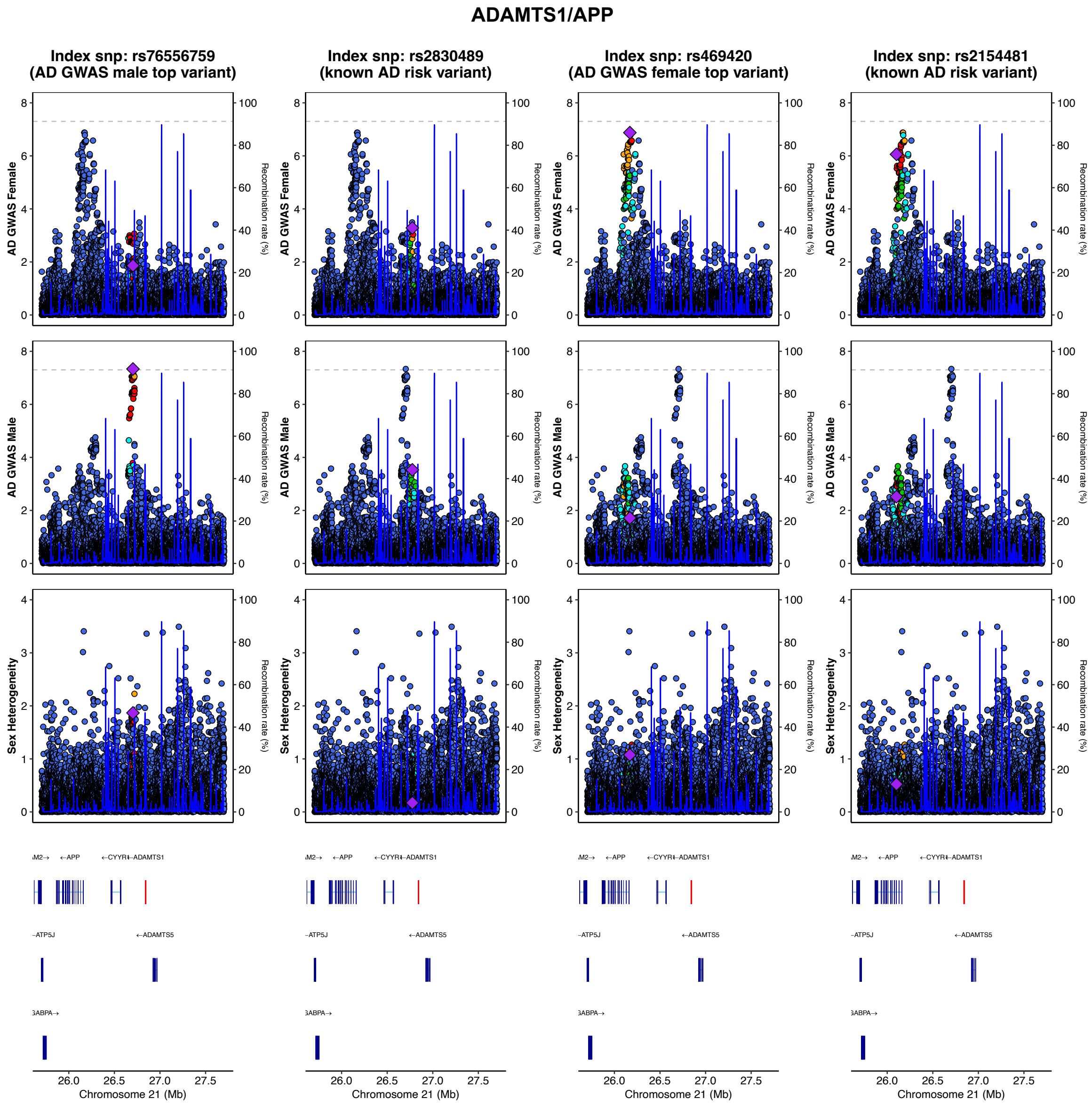

| | rs76556759 | | rs2830489 | | rs469420 | | rs2154481 | |
|---|---|---|---|---|---|---|---|---|
| | Beta | SE | Beta | SE | Beta | SE | Beta | SE |
| Female AD GWAS | 0.037 | 0.015 | -0.036 | 0.010 | 0.050 | 0.009 | 0.048 | 0.010 |
| Male AD GWAS | 0.094 | 0.017 | -0.043 | 0.012 | 0.025 | 0.011 | 0.032 | 0.011 |

**S4.10**

#### ABCA7

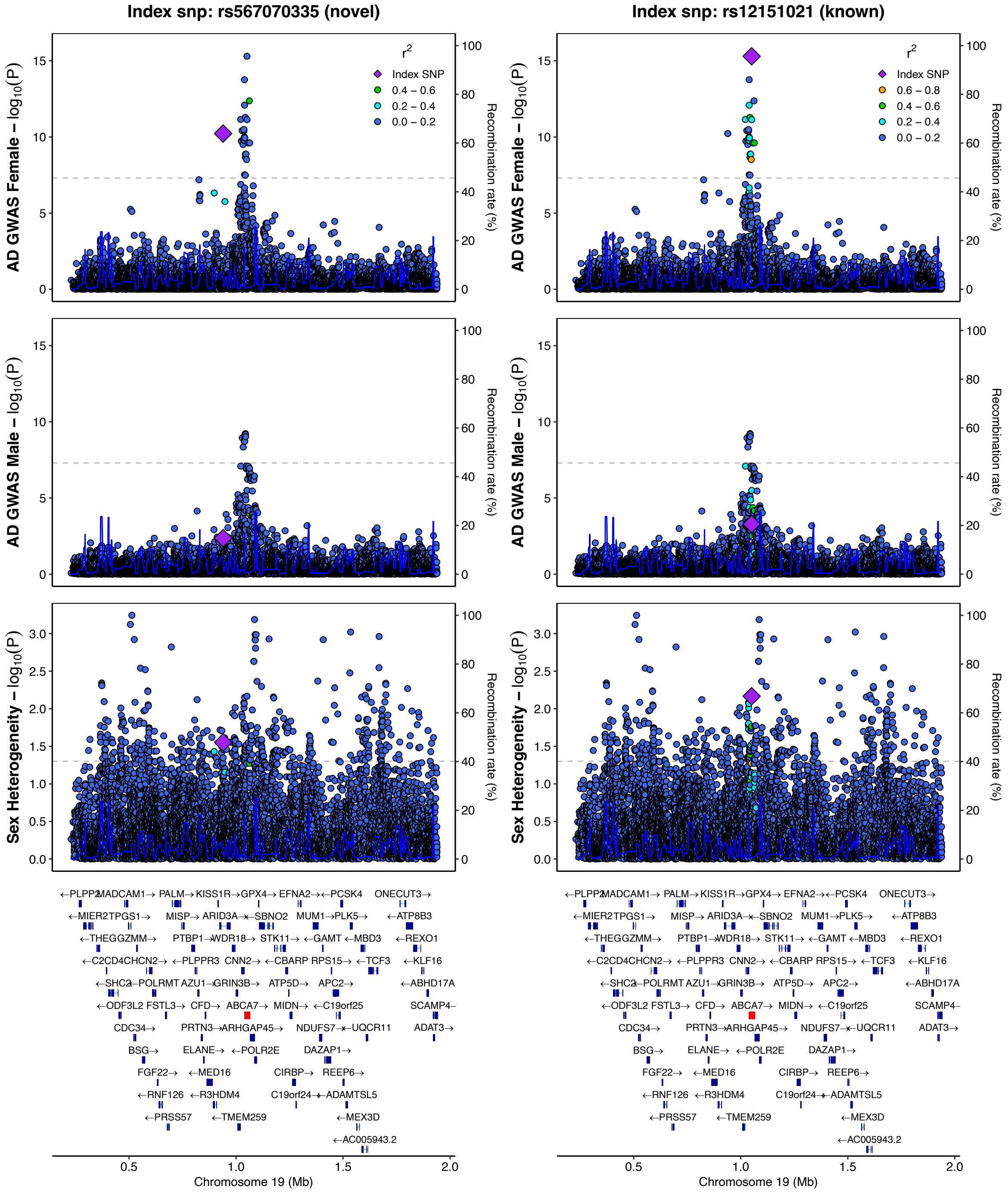

Index snp: rs567070335 (novel)
Index snp: rs12151021 (known)
AD GWAS Female – log10(P)
AD GWAS Male – log10(P)
Sex Heterogeneity – log10(P)
Recombination rate (%)
r2
Index SNP
0.6 – 0.8
0.4 – 0.6
0.2 – 0.4
0.0 – 0.2
Chromosome 19 (Mb)

## S4.11

##### CLU/PTK2B

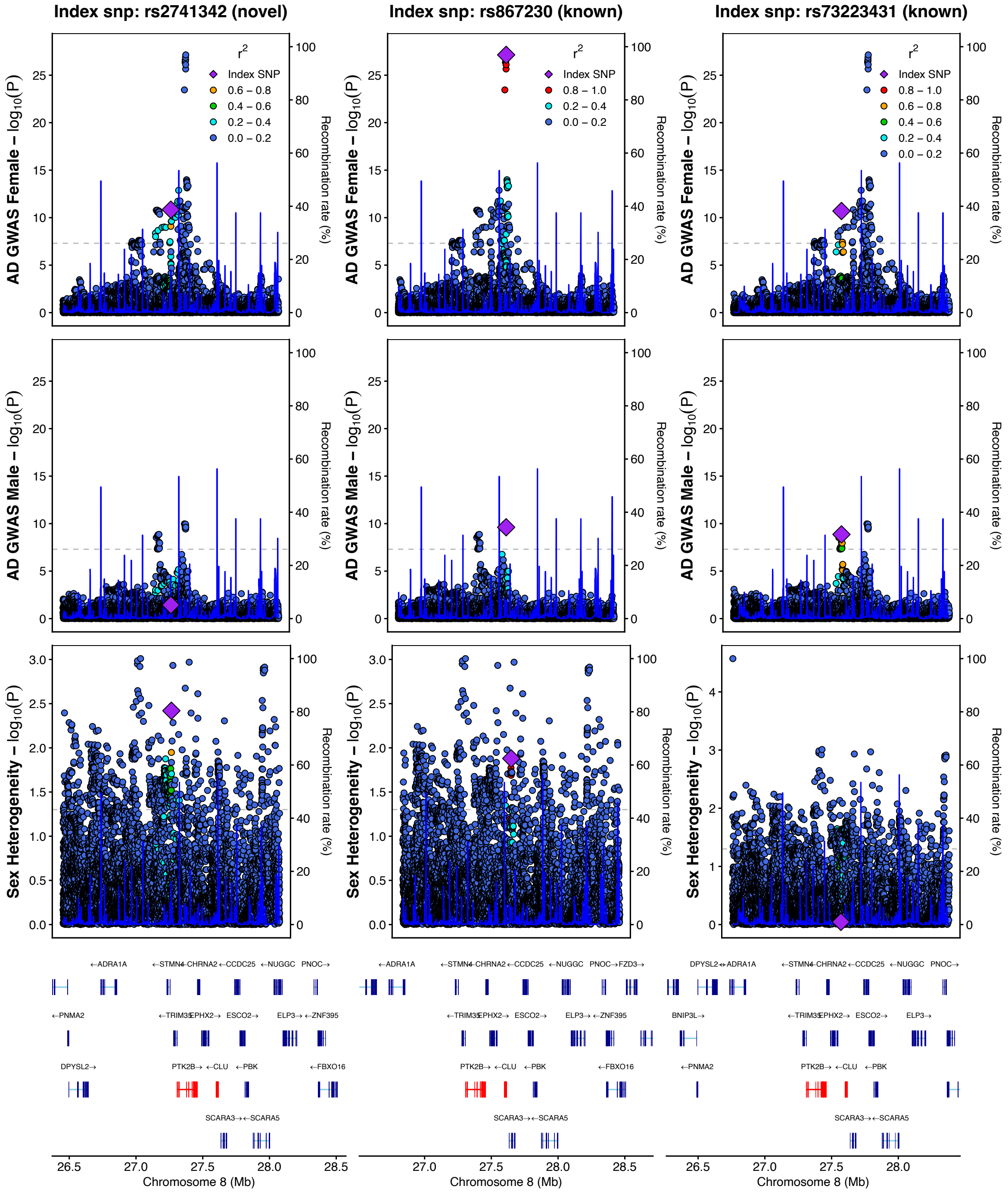

## S4.12

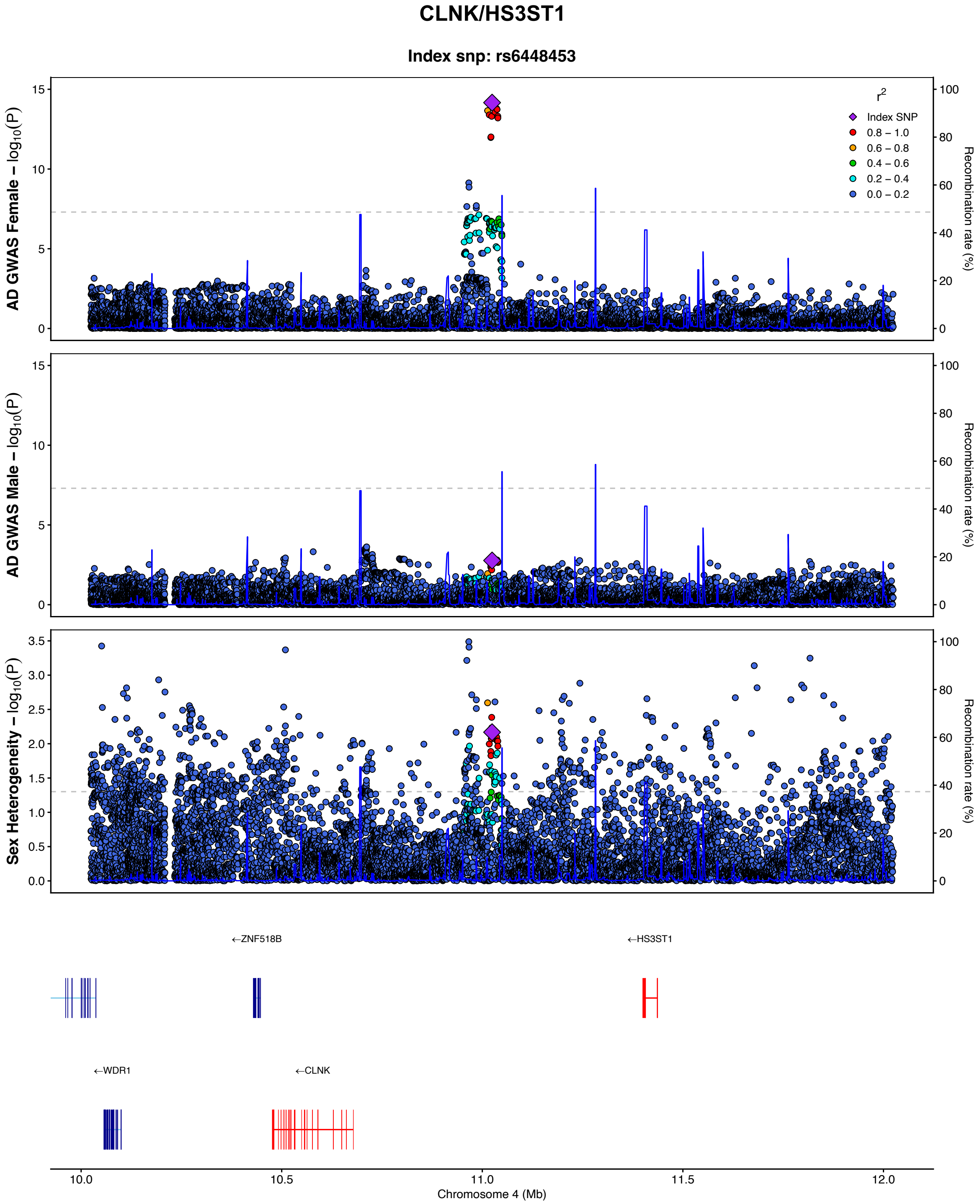

CLNK/HS3ST1
Index snp: rs6448453
AD GWAS Female – log10(P)
AD GWAS Male – log10(P)
Sex Heterogeneity – log10(P)
Recombination rate (%)
r2
Index SNP
0.8 – 1.0
0.6 – 0.8
0.4 – 0.6
0.2 – 0.4
0.0 – 0.2
←ZNF518B
←HS3ST1
←WDR1
←CLNK
10.0
10.5
11.0
11.5
12.0
Chromosome 4 (Mb)

## S4.13

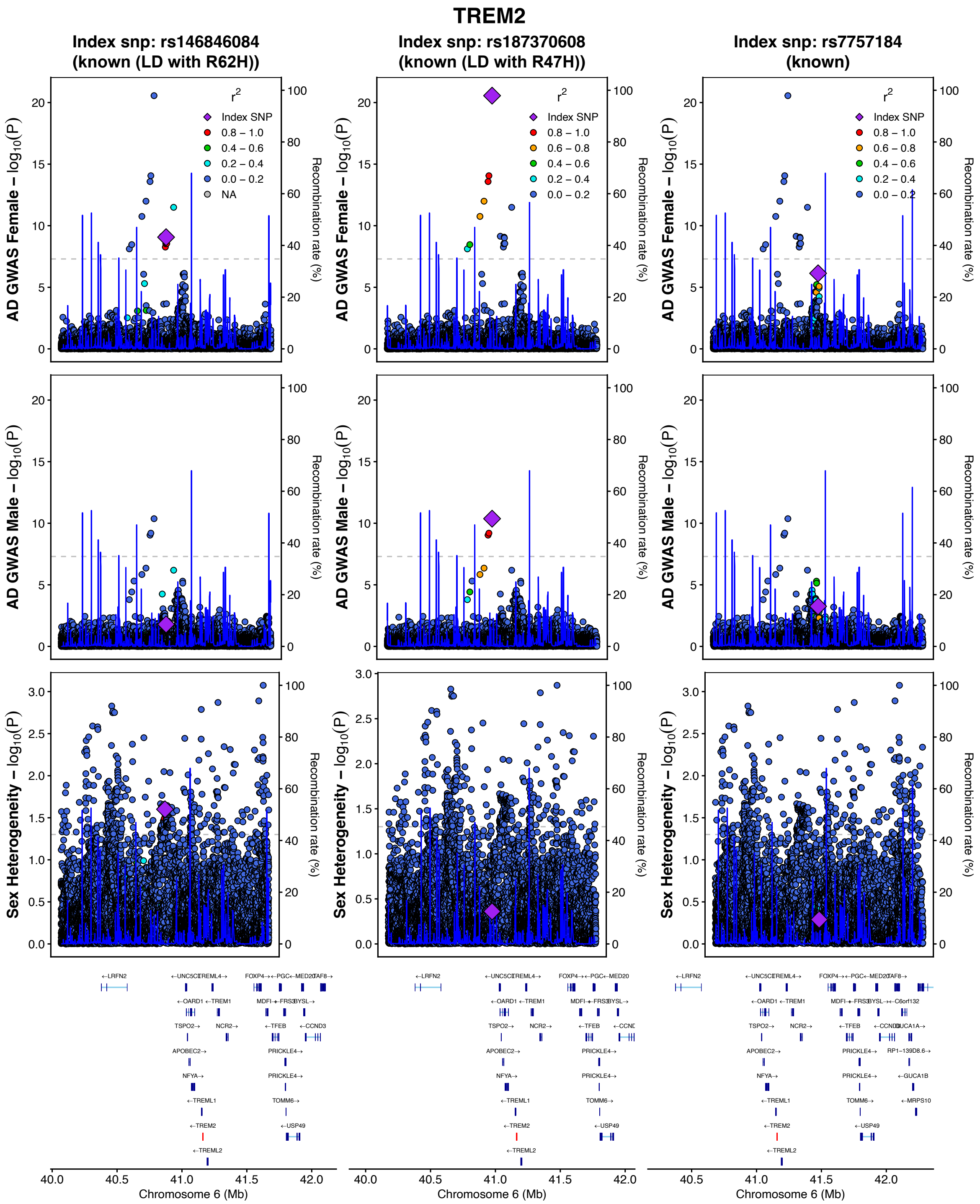
TREM2
Index snp: rs146846084
(known (LD with R62H))
Index snp: rs187370608
(known (LD with R47H))
Index snp: rs7757184
(known)
AD GWAS Female – log10(P)
AD GWAS Male – log10(P)
Sex Heterogeneity – log10(P)
Recombination rate (%)
Index SNP
0.8 – 1.0
0.6 – 0.8
0.4 – 0.6
0.2 – 0.4
0.0 – 0.2
NA
Chromosome 6 (Mb)

## S4.14

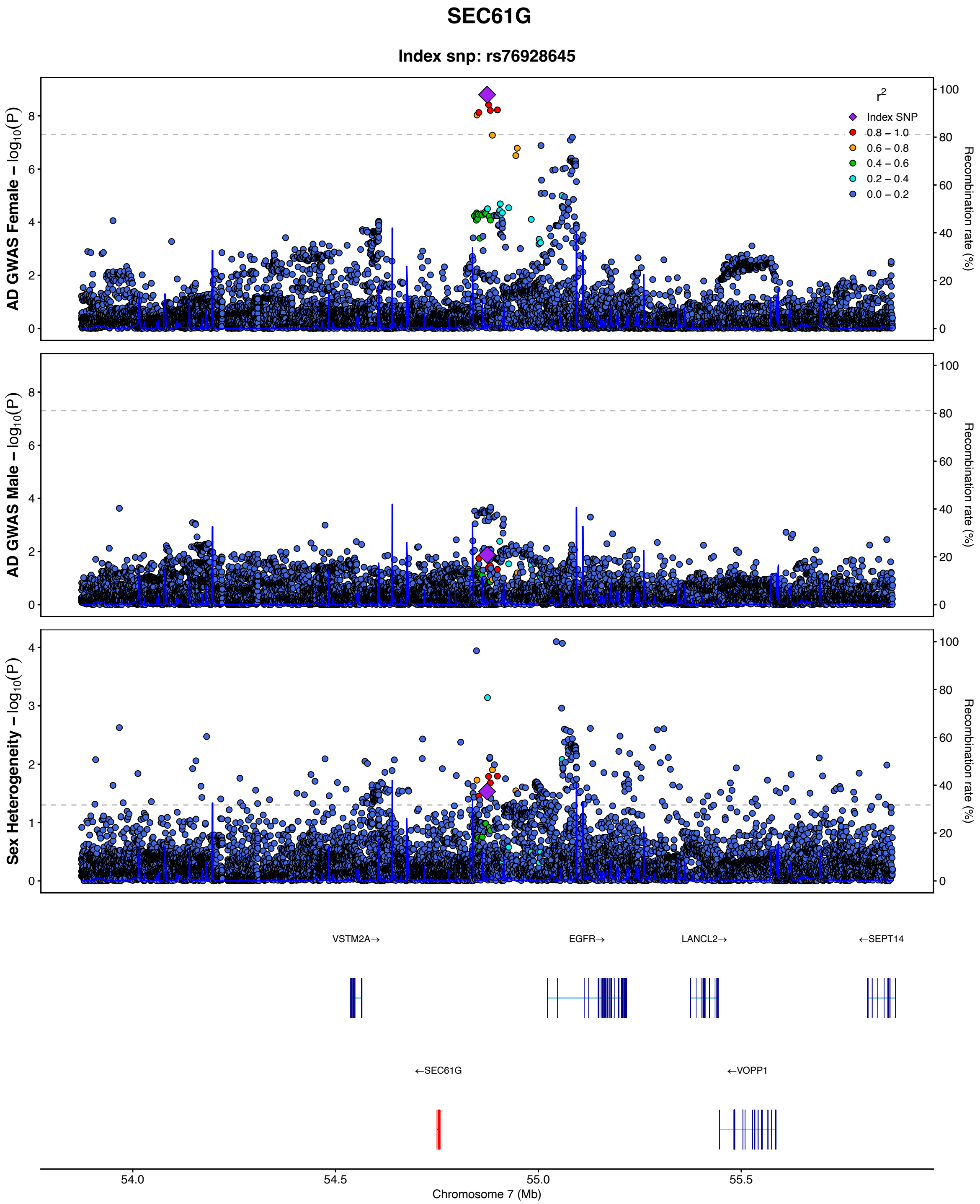
SEC61G
Index snp: rs76928645
AD GWAS Female – log10(P)
AD GWAS Male – log10(P)
Sex Heterogeneity – log10(P)
Recombination rate (%)
r2
Index SNP
0.8 – 1.0
0.6 – 0.8
0.4 – 0.6
0.2 – 0.4
0.0 – 0.2
VSTM2A→
EGFR→
LANCL2→
←SEPT14
←SEC61G
←VOPP1
54.0
54.5
55.0
55.5
Chromosome 7 (Mb)

## S4.15

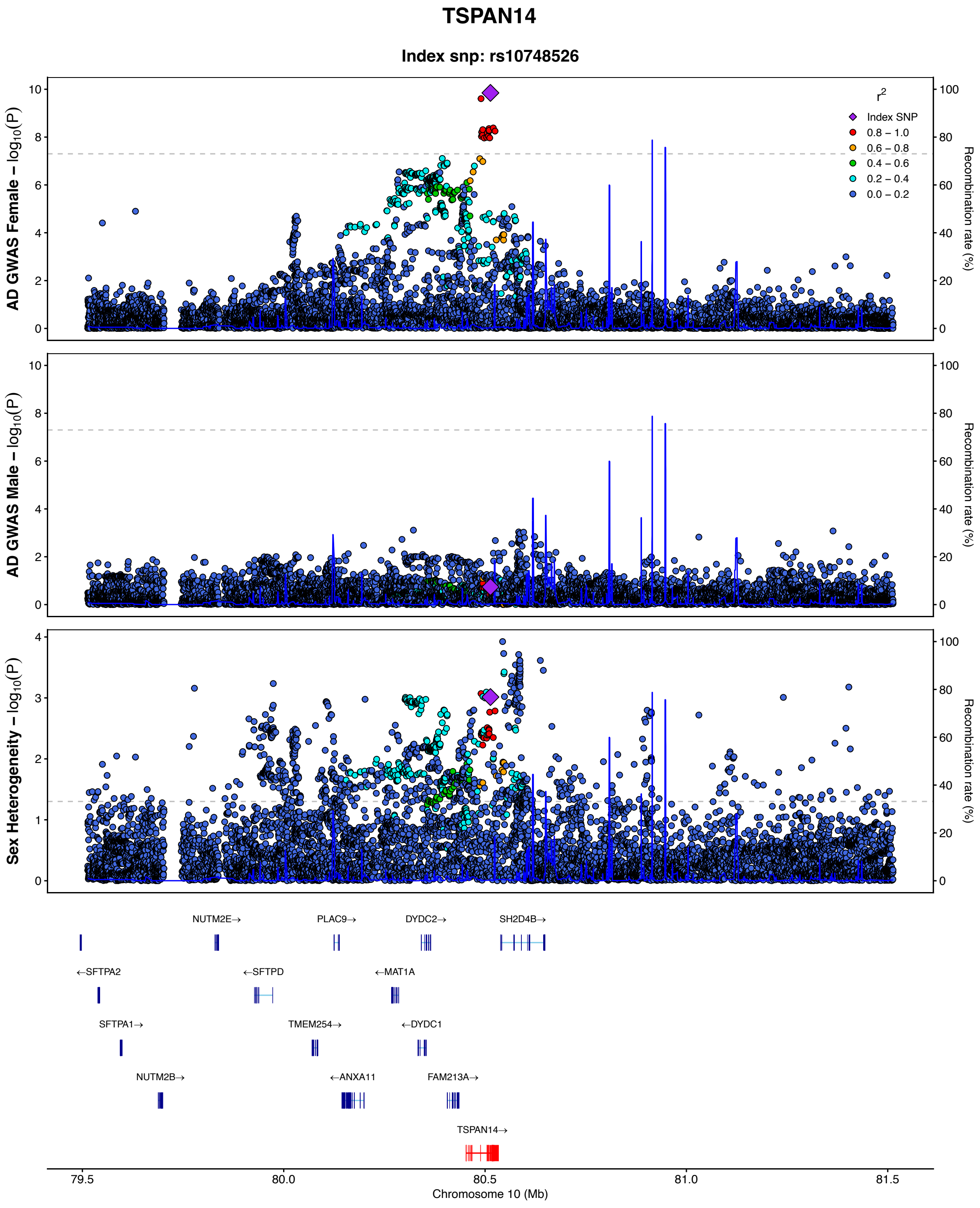
TSPAN14
Index snp: rs10748526
AD GWAS Female – log10(P)
AD GWAS Male – log10(P)
Sex Heterogeneity – log10(P)
Recombination rate (%)
r2
Index SNP
0.8 – 1.0
0.6 – 0.8
0.4 – 0.6
0.2 – 0.4
0.0 – 0.2
NUTM2E→
PLAC9→
DYDC2→
SH2D4B→
←SFTPA2
←SFTPD
←MAT1A
SFTPA1→
TMEM254→
←DYDC1
NUTM2B→
←ANXA11
FAM213A→
TSPAN14→
79.5
80.0
80.5
81.0
81.5
Chromosome 10 (Mb)

## S4.16

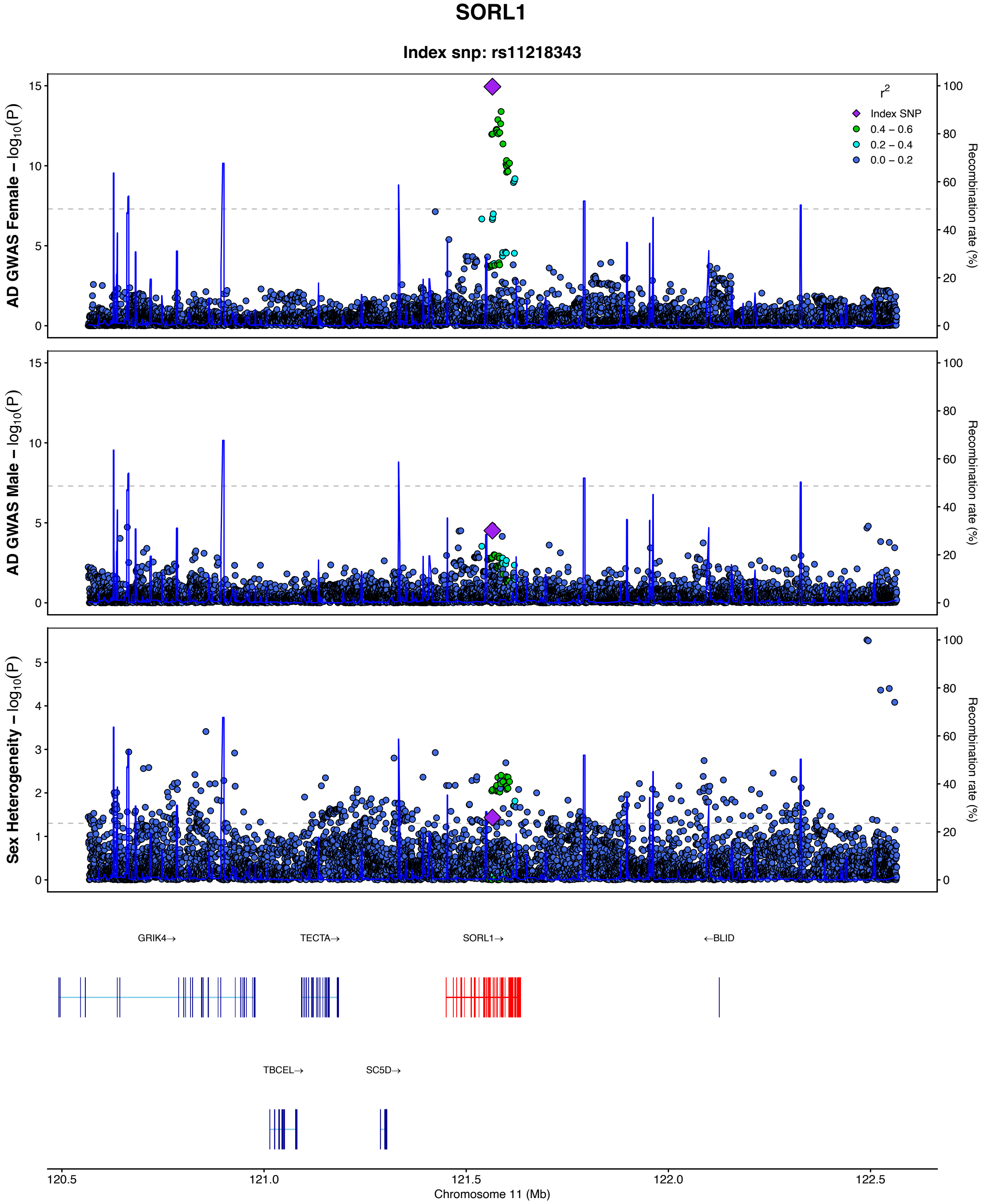

SORL1
Index snp: rs11218343
AD GWAS Female – log10(P)
AD GWAS Male – log10(P)
Sex Heterogeneity – log10(P)
Recombination rate (%)
r2
Index SNP
0.4 – 0.6
0.2 – 0.4
0.0 – 0.2
GRIK4→
TECTA→
SORL1→
←BLID
TBCEL→
SC5D→
120.5
121.0
121.5
122.0
122.5
Chromosome 11 (Mb)

## S4.17

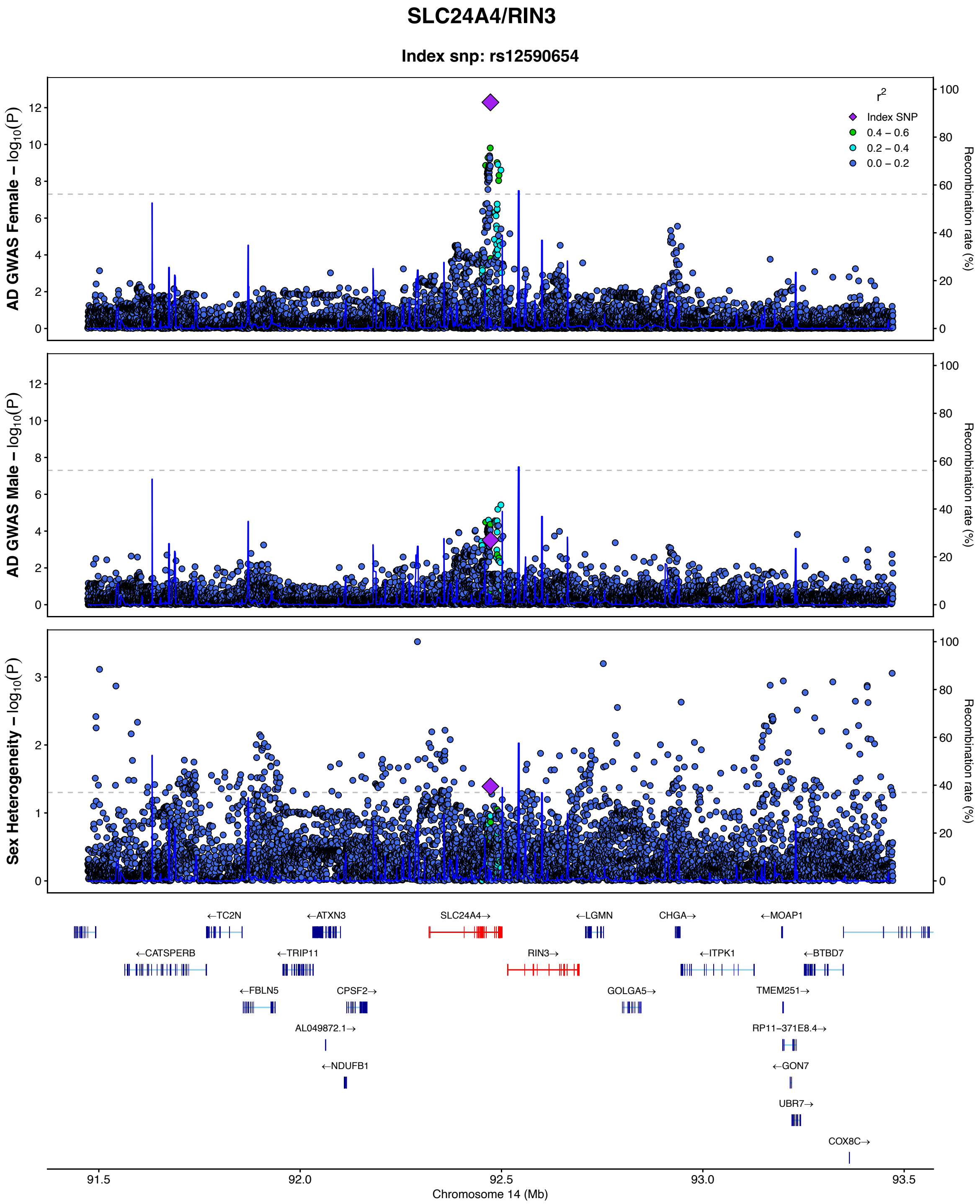
SLC24A4/RIN3
Index snp: rs12590654
AD GWAS Female – log10(P)
AD GWAS Male – log10(P)
Sex Heterogeneity – log10(P)
Recombination rate (%)
r2
Index SNP
0.4 – 0.6
0.2 – 0.4
0.0 – 0.2
←TC2N
←ATXN3
SLC24A4→
←LGMN
CHGA→
←MOAP1
←CATSPERB
←TRIP11
RIN3→
←ITPK1
←BTBD7
←FBLN5
CPSF2→
GOLGA5→
TMEM251→
AL049872.1→
RP11−371E8.4→
←NDUFB1
←GON7
UBR7→
COX8C→
91.5
92.0
92.5
93.0
93.5
Chromosome 14 (Mb)

## S4.18

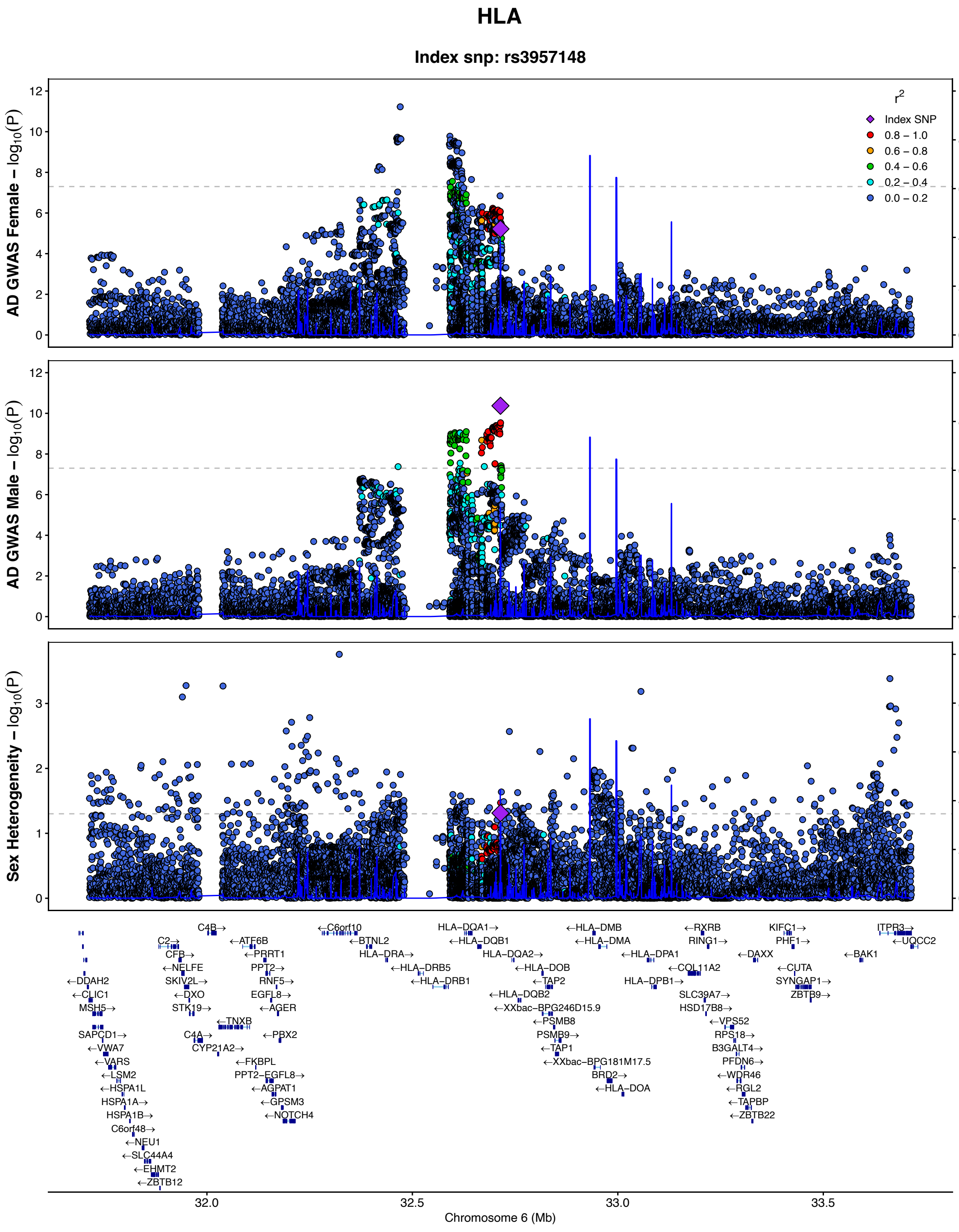

HLA
Index snp: rs3957148
AD GWAS Female – log10(P)
AD GWAS Male – log10(P)
Sex Heterogeneity – log10(P)
Recombination rate (%)
r2
Index SNP
0.8 – 1.0
0.6 – 0.8
0.4 – 0.6
0.2 – 0.4
0.0 – 0.2
Chromosome 6 (Mb)
32.0
32.5
33.0
33.5

## S4.19

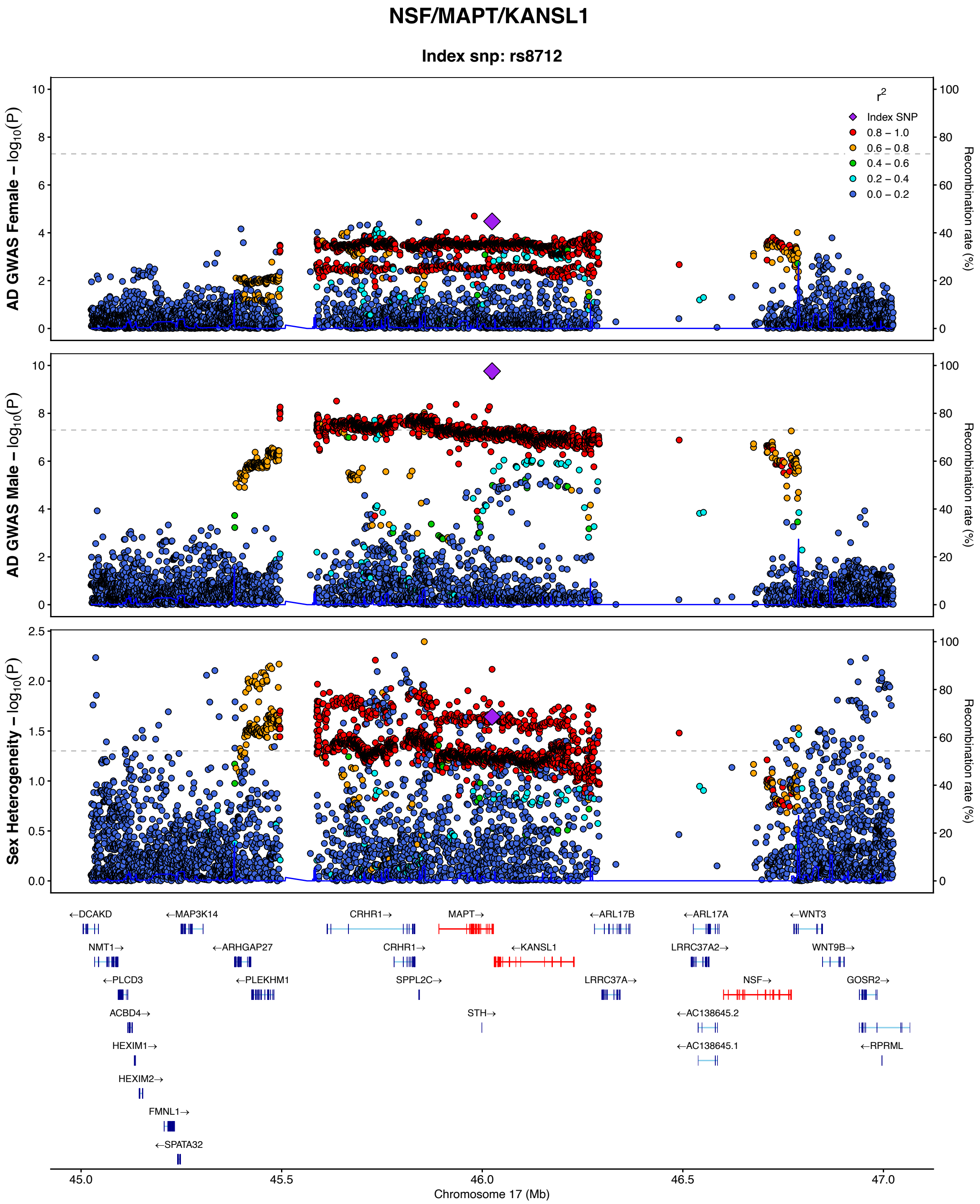

NSF/MAPT/KANSL1
Index snp: rs8712
AD GWAS Female – $\log_{10}(P)$
AD GWAS Male – $\log_{10}(P)$
Sex Heterogeneity – $\log_{10}(P)$
Recombination rate (%)
$r^2$
Index SNP
0.8 – 1.0
0.6 – 0.8
0.4 – 0.6
0.2 – 0.4
0.0 – 0.2
←DCAKD
←MAP3K14
CRHR1→
MAPT→
←ARL17B
←ARL17A
←WNT3
NMT1→
←ARHGAP27
CRHR1→
←KANSL1
LRRC37A2→
WNT9B→
←PLCD3
←PLEKHM1
SPPL2C→
LRRC37A→
NSF→
GOSR2→
ACBD4→
STH→
←AC138645.2
HEXIM1→
←AC138645.1
←RPRML
HEXIM2→
FMNL1→
←SPATA32
45.0
45.5
46.0
46.5
47.0
Chromosome 17 (Mb)

## S4.20

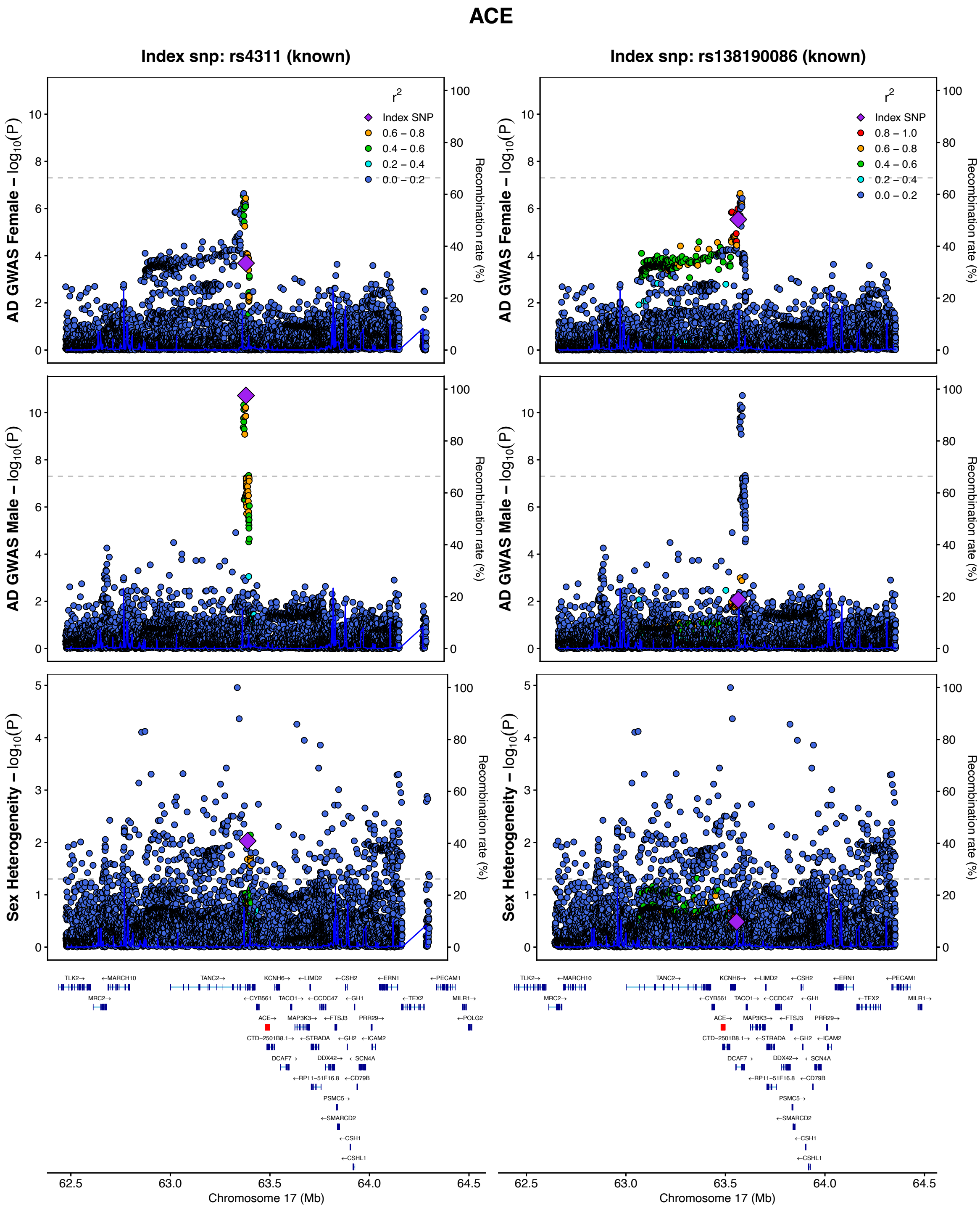
ACE
Index snp: rs4311 (known)
Index snp: rs138190086 (known)
r2
Index SNP
0.8 – 1.0
0.6 – 0.8
0.4 – 0.6
0.2 – 0.4
0.0 – 0.2
AD GWAS Female – log10(P)
AD GWAS Male – log10(P)
Sex Heterogeneity – log10(P)
Recombination rate (%)
Chromosome 17 (Mb)
TLK2 MARCH10 TANC2 KCNH6 LIMD2 CSH2 ERN1 PECAM1
MRC2 CYB561 TACO1 CCDC47 GH1 TEX2 MILR1
ACE MAP3K3 FTSJ3 PRR29 POLG2
CTD-2501B8.1 STRADA GH2 ICAM2
DCAF7 DDX42 SCN4A
RP11-51F16.8 CD79B
PSMC5
SMARCD2
CSH1
CSHL1

**Supplementary Figure 4. Locus zoom plot of sex-specific Alzheimer's disease GWAS lead variants that represented novel Alzheimer's disease loci or signals. (1-5)** Novel AD loci. **(6-8)** Novel independent signals in known AD loci (left column); the right column indicates the known AD variant as the LD tagging variant. **(9)** Novel independent signal in known AD locus APP (first column); the second column indicates a known AD variant as the LD tagging variant; the third column indicates a novel independent female variant just below genome wide significance; the fourth column indicates LD with another known AD variant, in LD with the female-biassed lead variant in column 3; These plots and appended table highlight evidence of both male and female biased heterogeneity, suggesting AD genetic signals near APP are not specifically male-biased. **(10-11**) 2 novel independent signals in known AD loci were not the top signal at the locus (i.e. the known AD variant was more significant) and not reported in Figure 1 and Table 1 but are shown here for completeness (discovery criteria required novel hits to be the most significant signal at the locus); the second (and third) column indicates the known AD variant as the LD tagging variant. **(12-20)** Known signals in known loci. For some loci across (6-20), we additionally displayed known AD variants. Dot color coding represents LD with tagging variant (purple diamond): red – R2: 0.8-1.0, orange – R2:0.6-0.8, green – R2:0.4-0.6, cyan – R2:0.2-0.4, blue – R2:0.0-0.2.

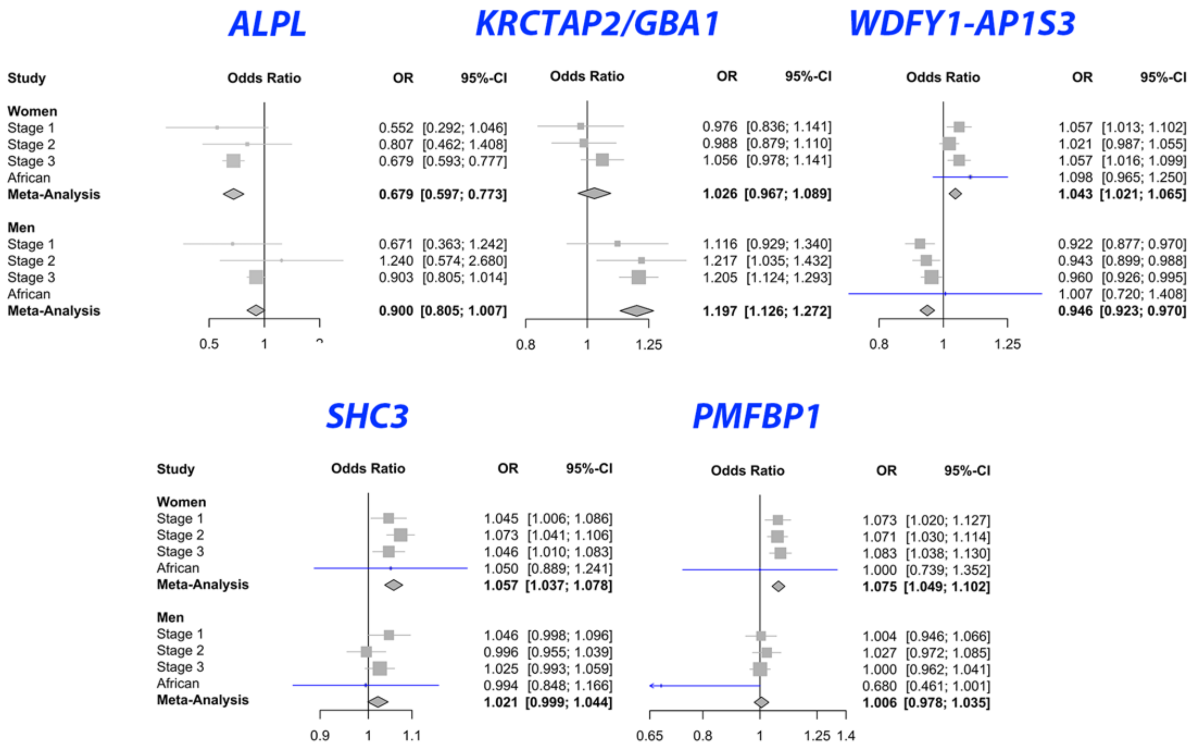

**Supplementary Figure 5. Forest plots for 5 sex-specific associated lead variants in novel Alzheimer's disease loci.** Plots illustrate consistent sex heterogeneity across study stages and the African ancestry dataset.

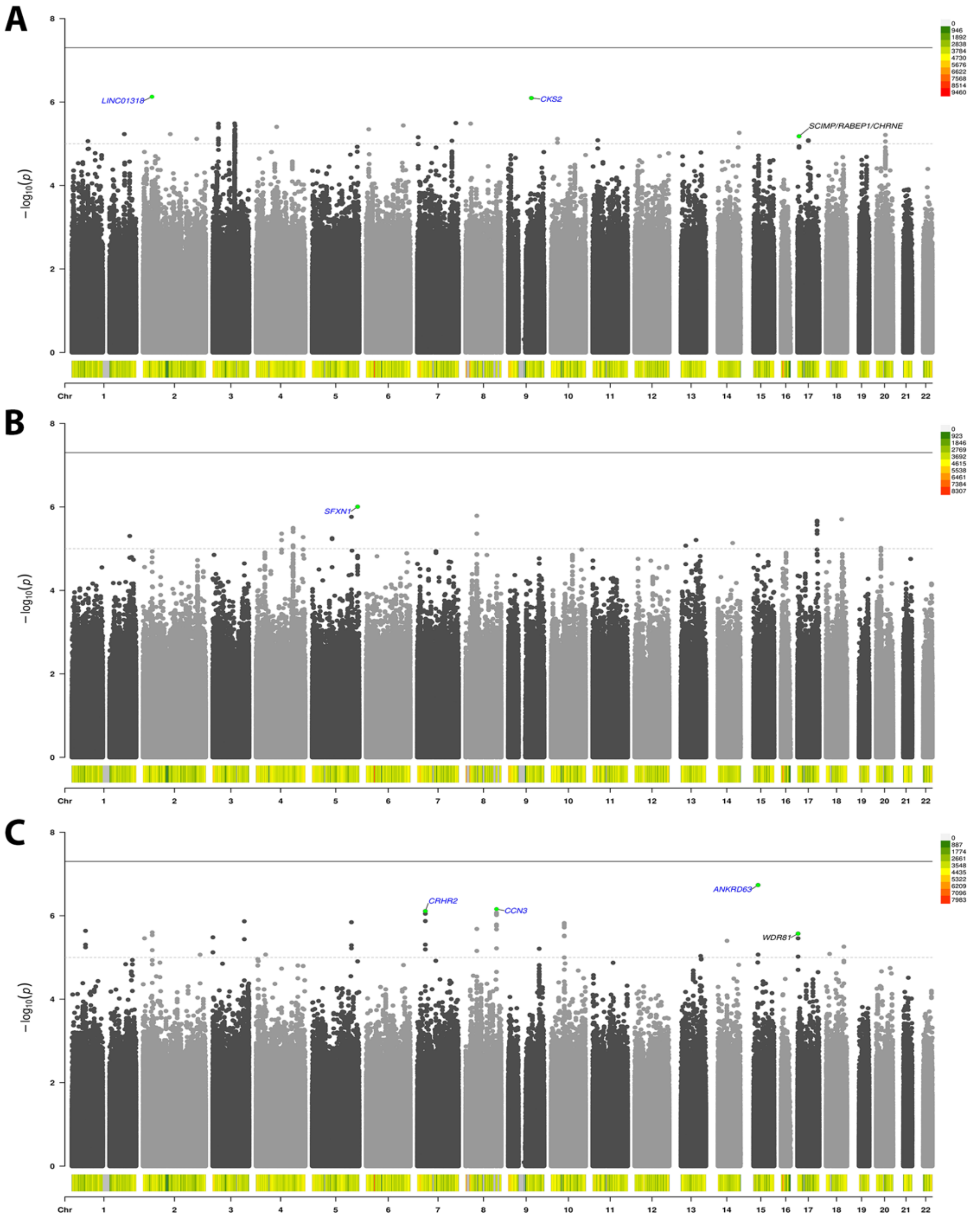

**Supplementary Figure 6. Sex stratified and heterogeneity GWAS of Alzheimer's disease in African ancestry data. A)** Women; **B)** Men; **C)** Sex heterogeneity. No novel genome-wide significant associations (*P*<5e-8, solid black line) were observed, while signals passing suggestive significance (*P*<1e-5, dashed gray line) in known Alzheimer's disease loci are marked in black text. For explorative/illustrative purposes, some lead variants in novel loci are marked with the nearest by gene for lead variant passing *P*<1e-6 (blue text). The color histograms at the bottom of the plots indicate variant density (cf. color scale legend).

**A**

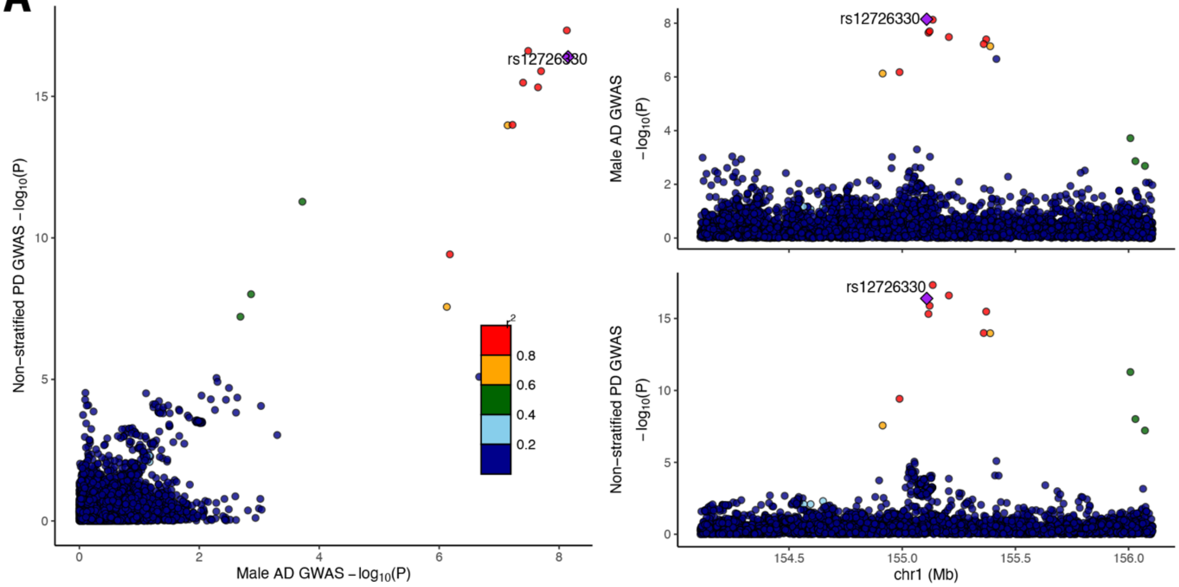

**B**

| | Male Stratum | | | Female Stratum | | |
|---|---|---|---|---|---|---|
| **rs12726330 associations with AD and AD-LB** | **OR [95% CI]** | **P-value** | **No.** | **OR [95% CI]** | **P-value** | **No.** |
| stage1 - all case-controls samples | 1.12 [0.93,1.34] | 0.24 | 12,337 | 0.98 [0.84,1.14] | 0.76 | 18,153 |
| stage1 - excluding pathology verified subjects | 1.29 [0.97,1.70] | 0.081 | 8,803 | 1.06 [0.84,1.35] | 0.60 | 13,391 |
| stage1 - pathology verified AD+/LB+ vs. AD+LB- | 1.24 [0.66,2.35] | 0.50 | 1,642 | 1.47 [0.81,2.69] | 0.21 | 1,898 |
| stage1 - pathology verified AD+ vs. AD- | 0.87 [0.51,1.49] | 0.61 | 3,545 | 0.70 [0.45,1.08] | 0.11 | 4,782 |
| stage2 | 1.22 [1.04,1.42] | 0.017 | 277,043 | 0.99 [0.88,1.11] | 0.84 | 335,917 |
| stage3 | 1.21 [1.12,1.29] | 1.90E-07 | 218,284 | 1.06 [0.98,1.14] | 0.16 | 282,064 |
| stage1+2+3 | 1.20 [1.13,1.28] | 7.09E-09 | 507,664 | 1.04 [0.97,1.10] | 0.39 | 636,134 |

| | Male Stratum | | | Female Stratum | | |
|---|---|---|---|---|---|---|
| **rs145330152 associations with PD and LBD** | **OR [95% CI]** | **P-value** | **No.** | **OR [95% CI]** | **P-value** | **No.** |
| Blauwendraat et al. 2021 - PD | 1.71 [1.48,1.97] | 1.50E-13 | 110,616 | 1.90 [1.64,2.19] | 5.20E-18 | 104,082 |
| Gibbons et al. 2022 - LBD | 3.27 [2.50,4.28] | 6.10E-18 | 11,709 | 1.92 [1.36,2.70] | 1.90E-04 | 10,652 |

**Supplementary Figure 7. Follow-up analyses at the *GBA1* locus. A)** The male Alzheimer's disease (AD) GWAS signal colocalizes with the non-sex-stratified GWAS signal for Parkinson's disease (PD) observed by Nalls et al. 2019 (PP4 = 1.00; PMID: 31701892). **B)** Top table shows association analyses for the top male AD GWAS variant (rs12726330) with AD across different stages and with AD with Lewy bodies (AD-LB) in stage 1. In stage 1, a subset of subjects had pathology verified diagnoses, with a further subset of those also having LB pathology data available. Three sensitivity analyses were conducted, (i) comparing AD cases versus controls using only subjects without pathology verified data; (ii) comparing subjects with AD pathology ($AD^+$) versus those without AD pathology ($AD^-$); and (iii) comparing subjects with both AD and LB pathology ($AD^+LB^+$) versus those with AD pathology but no LB pathology ($AD^+LB^-$). The bottom table shows sex-specific associations for rs145330152 (top PD variant, LD with rs12726330: $R^2$ = 0.86) with PD (Blauwendraat et al. 2021; PMID: 33901317) and Lewy Body Dementia (LBD; Gibbons et al. 2022; DOI: 10.1101/2022.11.22.22282597).
**We derive 3 primary conclusions**: **(1)** the smaller male effect size for rs12726330 in stage 1 relative to stages 2 and 3 AD analyses did not relate to potential biases with proxy and registry phenotypes, since the phase1 male effect size becoma larger when excluding pathology verified subjects; **(2)** the stage 1 $AD^+LB^-$ versus $AD^+LB^+$ analyses and LBD findings suggest rs12726330 contributes to increased risk of comorbid AD-LB pathology and AD-LBD mixed dementia, likely in a male-biased fashion (while the male bias appeared to be lost $AD^+LB^-$ versus $AD^+LB^+$, this may reflect power limitations in that subset of stage 1 data); **(3)** rs12726330 appears to have no effect or even a protective effect on AD pathology, but this observation may be affected by sample selection bias, since the subjects were first restricted to those with clinical AD case-control status, such that $AD^+$ pathology cases (which are almost all clinical AD cases) may be depleted for comorbid LB pathology and thus rs12726330 carriers.

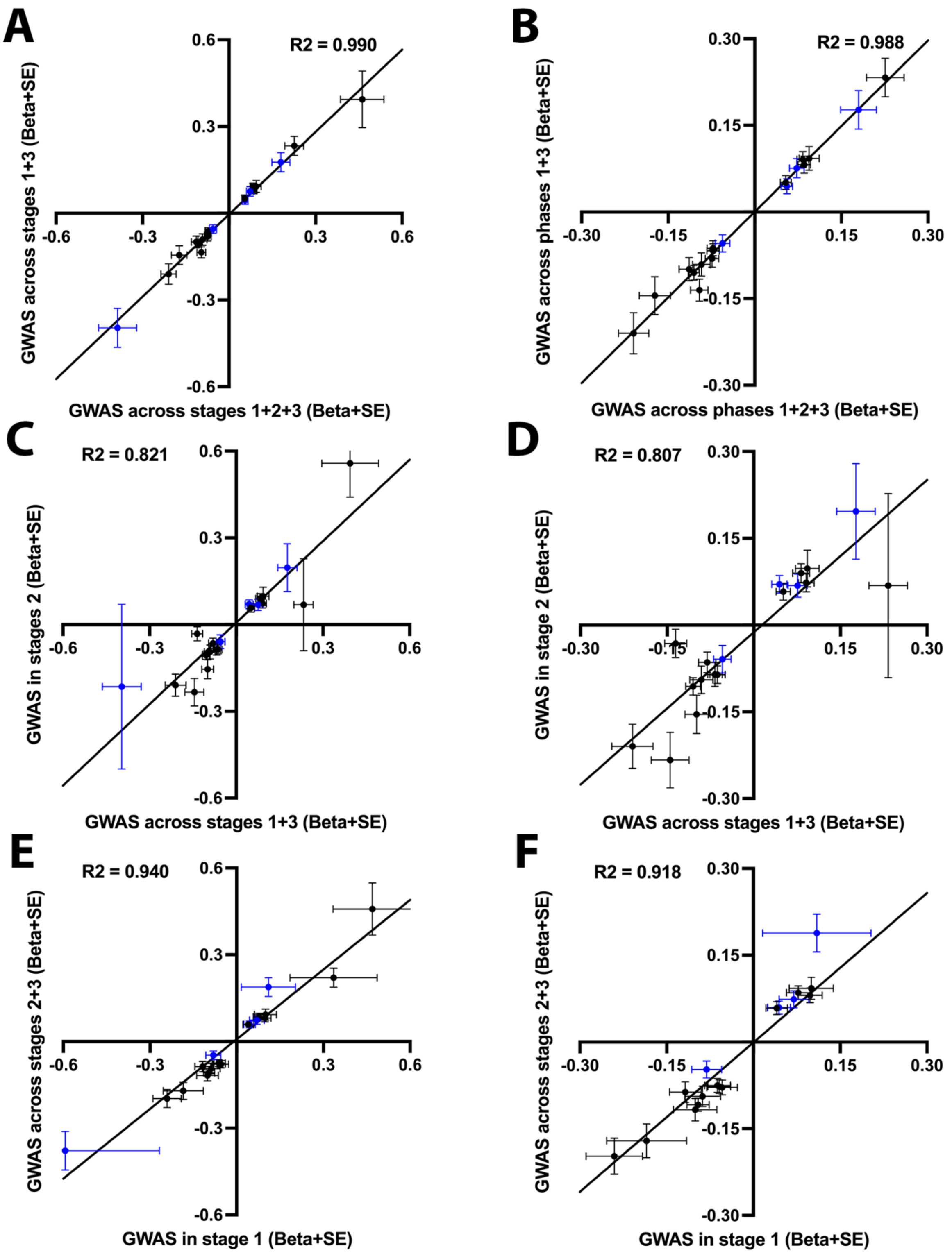
A
R2 = 0.990
GWAS across stages 1+3 (Beta+SE)
GWAS across stages 1+2+3 (Beta+SE)
0.6
0.3
-0.3
-0.6
-0.6
-0.3
0.3
0.6
B
R2 = 0.988
GWAS across phases 1+3 (Beta+SE)
GWAS across phases 1+2+3 (Beta+SE)
0.30
0.15
-0.15
-0.30
-0.30
-0.15
0.15
0.30
C
R2 = 0.821
GWAS in stages 2 (Beta+SE)
GWAS across stages 1+3 (Beta+SE)
D
R2 = 0.807
GWAS in stage 2 (Beta+SE)
GWAS across stages 1+3 (Beta+SE)
E
R2 = 0.940
GWAS across stages 2+3 (Beta+SE)
GWAS in stage 1 (Beta+SE)
F
R2 = 0.918
GWAS across stages 2+3 (Beta+SE)
GWAS in stage 1 (Beta+SE)

**Supplementary Figure 8. Consistency of sex-specific Alzheimer's disease lead variant associations across stages.** Given potential concerns with the use of Alzheimer's disease proxy (family history) or health registry phenotypes, we performed several comparisons of sex-specific lead variant effect sizes with meta-analyses variably excluding or including biobanks. Stage 1 corresponds to ADGC+ADSP ("clean" clinical and partly pathology verified diagnoses), stage 2 corresponds to UKB (mostly proxy phenotypes), and stage 3 corresponds to FinnGen (health registry phenotypes). Scatter plots illustrate high correlation for **A,C,E)** all sex-specific lead variants, and **B,D,F)** sex-specific variants with effect allele frequencies >1%. The 5 novel Alzheimer's disease risk loci are marked in blue. **A-B)** Comparison before and after adding UKB into the full meta-analysis. **C-D)** Comparison of UKB effect sizes versus effect sizes when excluding UKB. **E-F)** Comparison of effect sizes in clean case-control data (ADGC+ADSP) versus biobanks (UKB+FinnGen). In (F), the blue outlier in (top right corner) corresponds to *GBA1*, which displayed larger effect sizes in UKB (however cf. Figure S7). Further, one data point in (F) is omitted to allow for a sufficiently close-up view of all other data points.

### PWAS

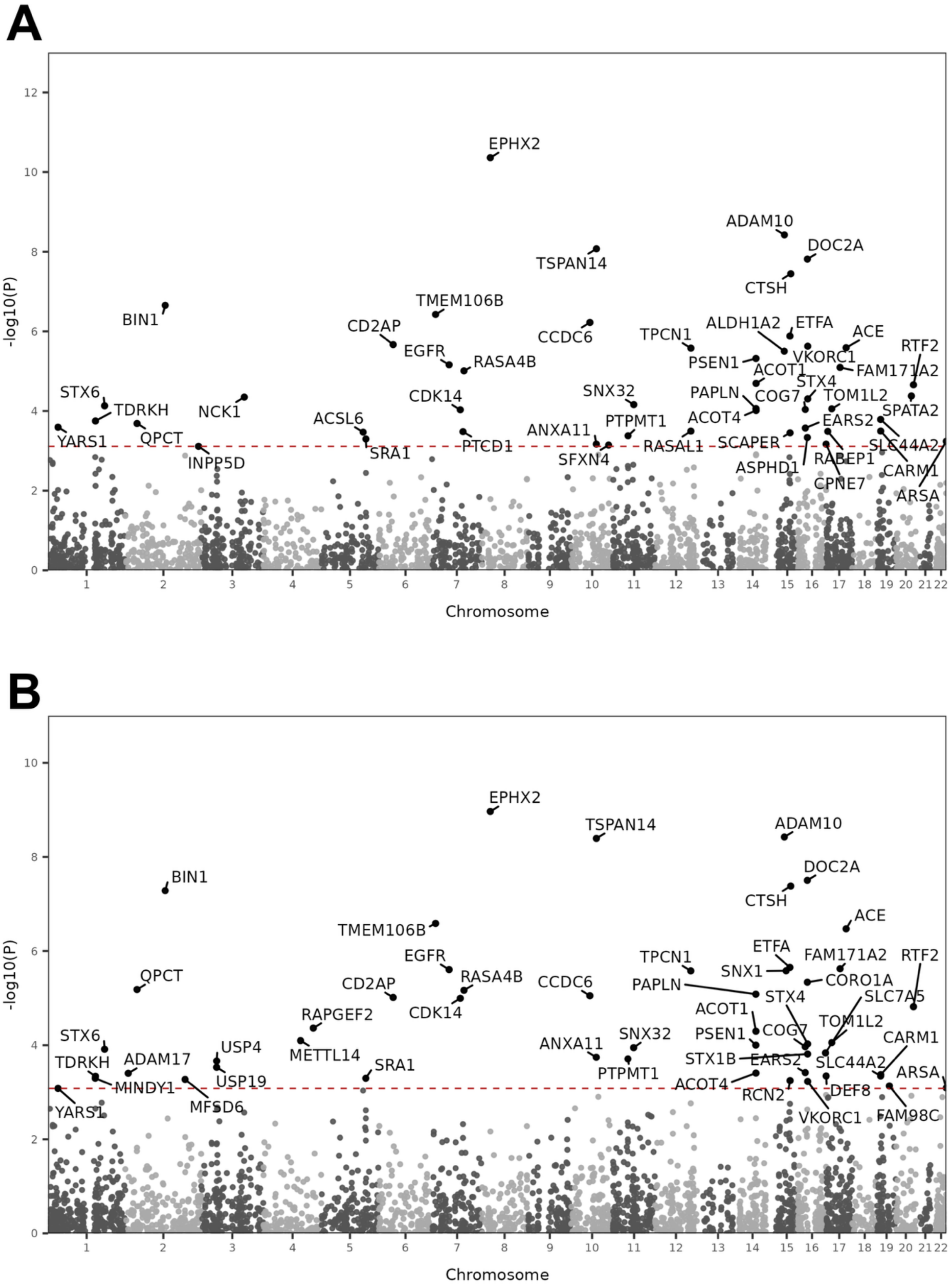
A
B
-log10(P)
Chromosome
EPHX2
ADAM10
DOC2A
TSPAN14
CTSH
BIN1
TMEM106B
CD2AP
CCDC6
TPCN1
ALDH1A2
ETFA
ACE
EGFR
RASA4B
PSEN1
VKORC1
RTF2
ACOT1
FAM171A2
STX6
CDK14
SNX32
PAPLN
STX4
COG7
TOM1L2
TDRKH
NCK1
ACSL6
PTPMT1
ACOT4
EARS2
SPATA2
ANXA11
YARS1
QPCT
PTCD1
RASAL1
SCAPER
SLC44A2
SRA1
SFXN4
RABEP1
INPP5D
ASPHD1
CARM1
CPNE7
ARSA
SNX1
CORO1A
SLC7A5
RAPGEF2
METTL14
USP4
ADAM17
MINDY1
USP19
MFSD6
STX1B
DEF8
RCN2
FAM98C

C

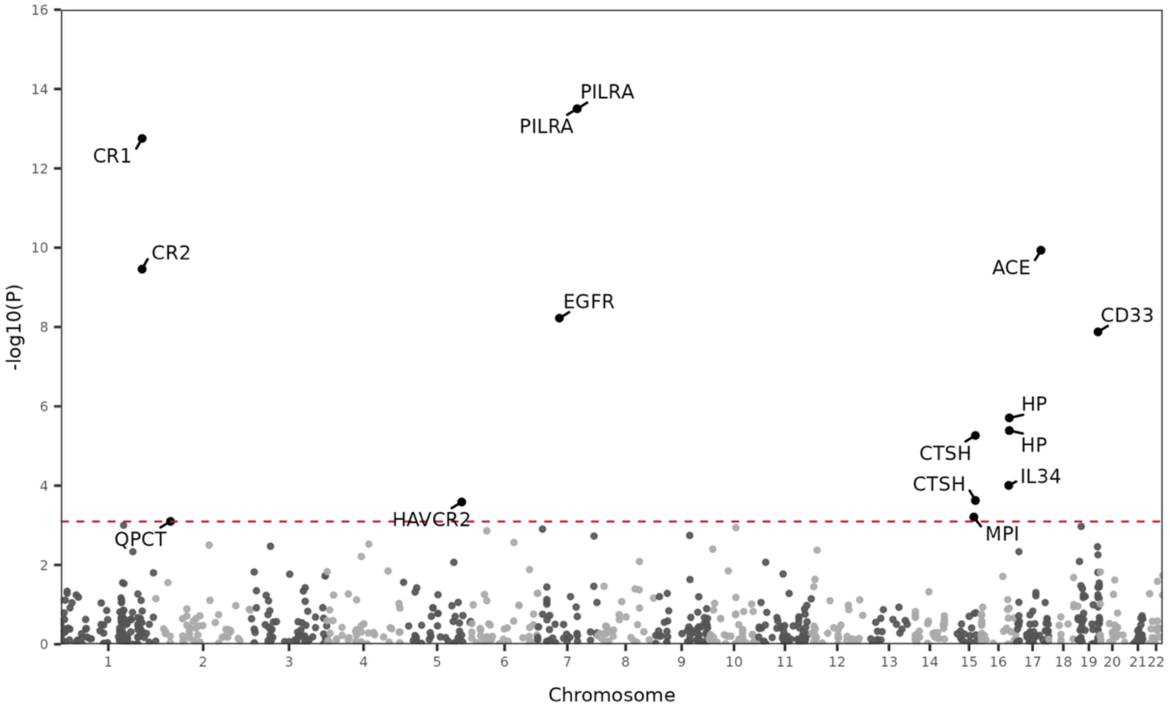
PILRA
PILRA
CR1
CR2
EGFR
ACE
CD33
HP
HP
CTSH
CTSH
IL34
HAVCR2
QPCT
MPI
-log10(P)
Chromosome

D

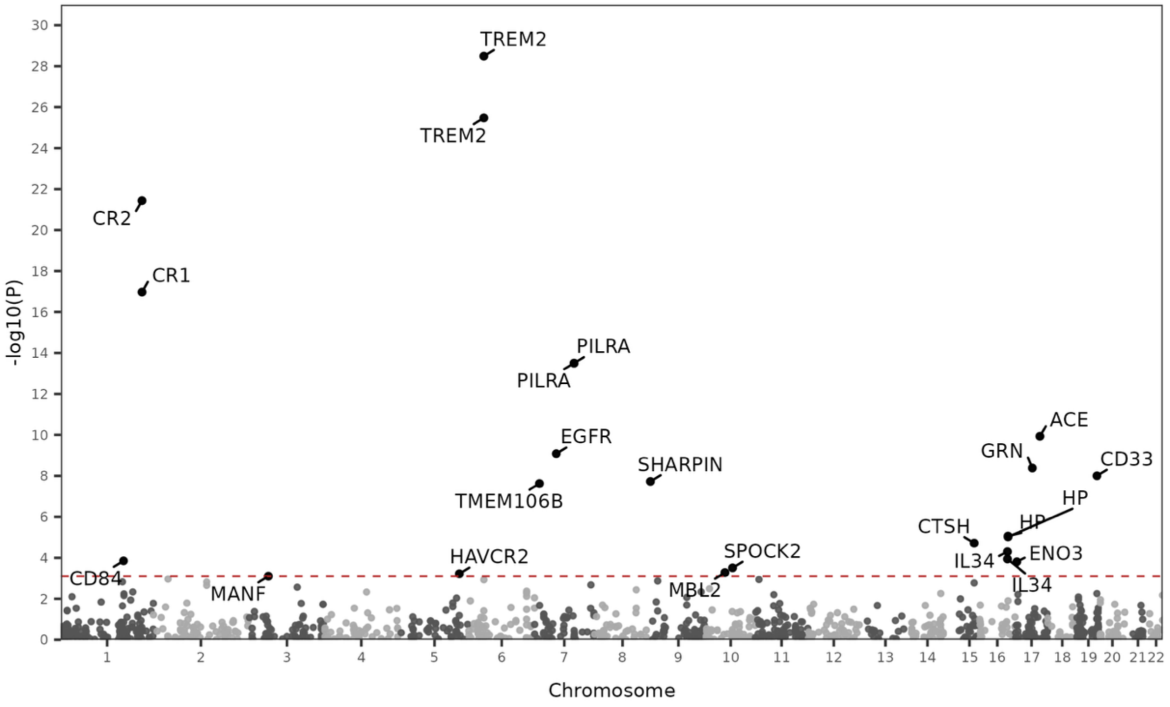
TREM2
TREM2
CR2
CR1
PILRA
PILRA
EGFR
SHARPIN
TMEM106B
ACE
GRN
CD33
HP
HP
CTSH
IL34
ENO3
IL34
SPOCK2
HAVCR2
MBL2
CD84
MANF
-log10(P)
Chromosome

E

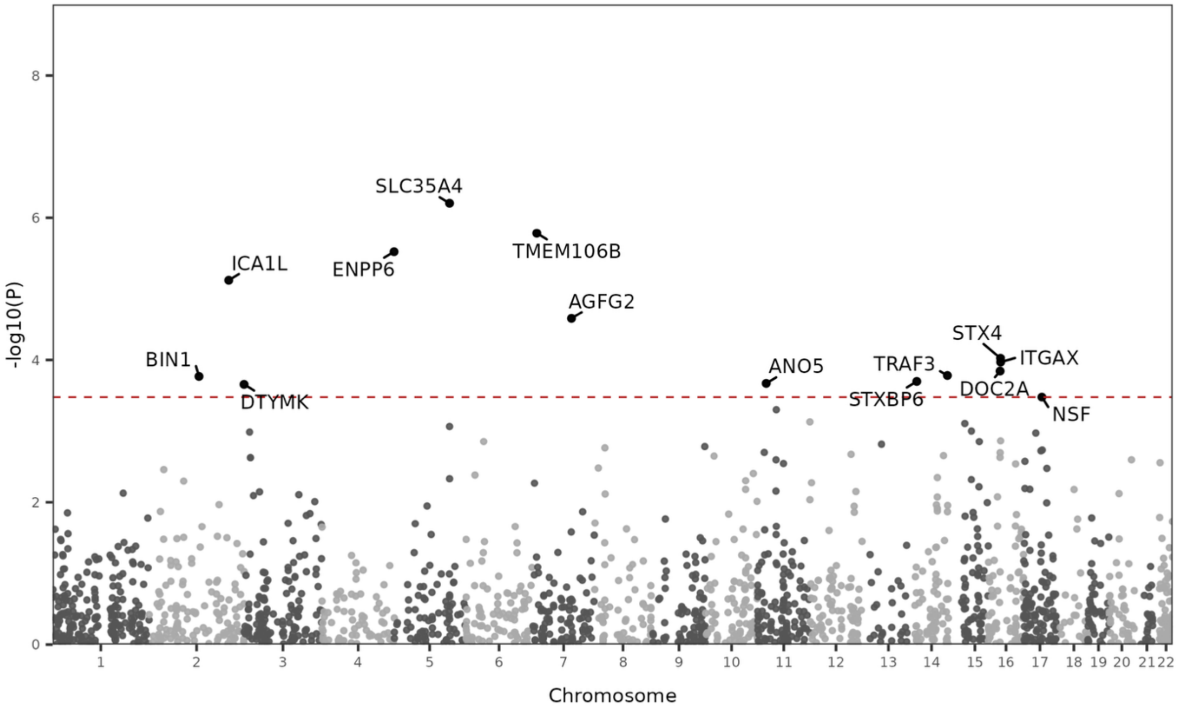

SLC35A4
TMEM106B
ICA1L
ENPP6
AGFG2
STX4
ITGAX
BIN1
ANO5
TRAF3
DOC2A
STXBP6
DTYMK
NSF
-log10(P)
Chromosome

F

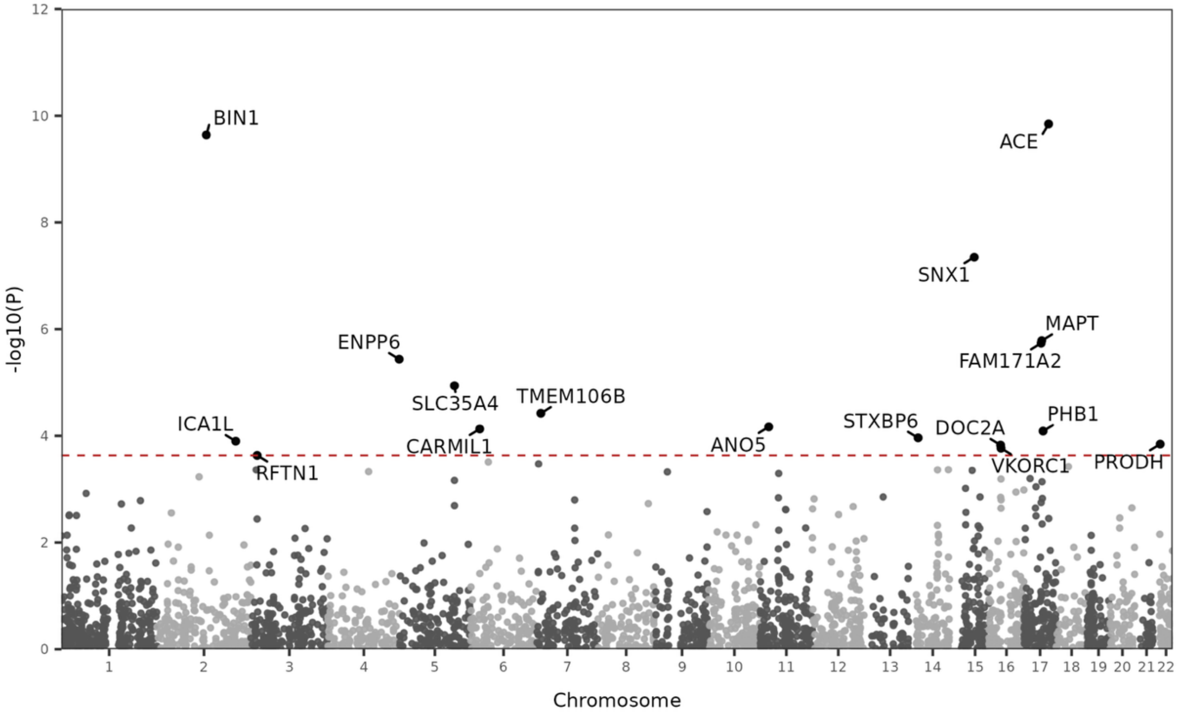

BIN1
ACE
SNX1
MAPT
FAM171A2
ENPP6
SLC35A4
TMEM106B
PHB1
ICA1L
STXBP6
DOC2A
CARMIL1
ANO5
VKORC1
PRODH
RFTN1
-log10(P)
Chromosome

G

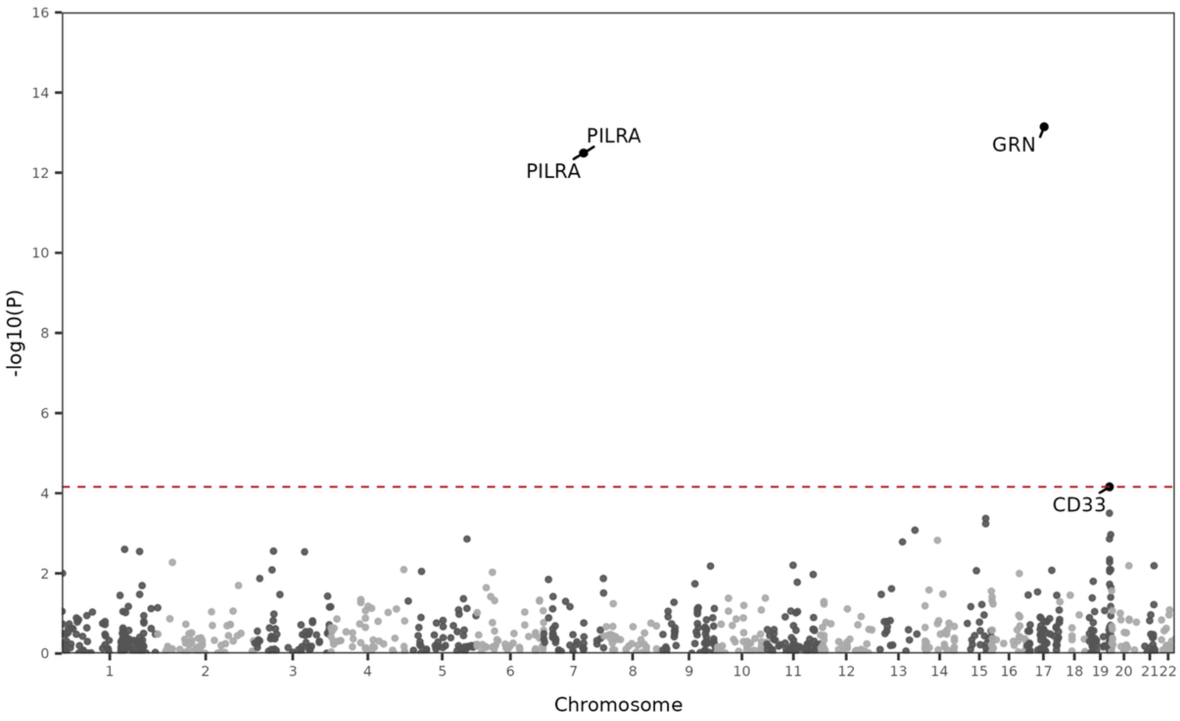
-log10(P)
Chromosome
PILRA
PILRA
GRN
CD33

H

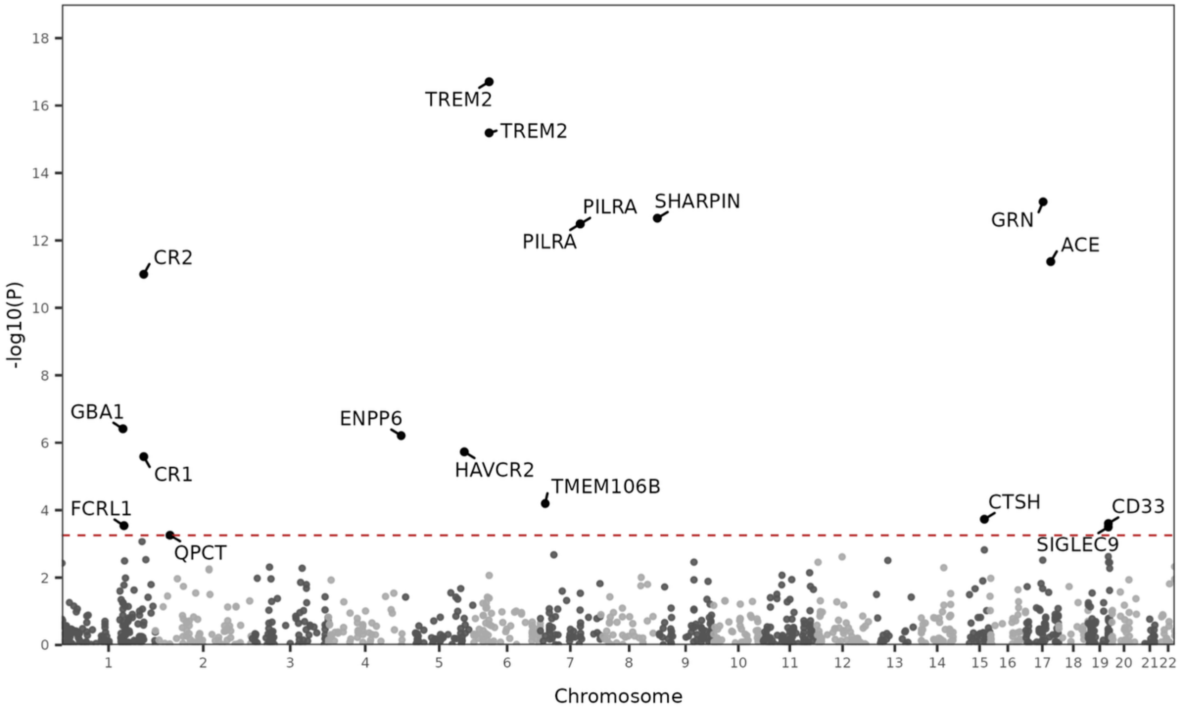
-log10(P)
Chromosome
TREM2
TREM2
PILRA
PILRA
SHARPIN
GRN
ACE
CR2
GBA1
ENPP6
CR1
HAVCR2
TMEM106B
FCRL1
QPCT
CTSH
CD33
SIGLEC9

**Supplementary Figure 9. Sex-stratified EUR Alzheimer's disease PWAS: All discoveries by sex and tissue. A)** Female primary brain discovery; **B)** Female secondary brain discovery; **C)** Female primary CSF discovery; **D)** Female secondary CSF discovery; **E)** Male primary brain discovery; **F)** Male secondary brain discovery; **G)** Male primary CSF discovery; **H)** Male secondary CSF discovery. Datapoints in the Manhattan plots represent association tests between AD and a given protein in brain or aptamer in the CSF proteogenomic data. Primary discoveries integrate the sex-stratified EUR GWAS with sex-matched protein weights, while secondary discoveries integrate the sex-stratified EUR GWAS with non-sex-stratified protein weights. X-axis denotes genomic positions, and y-axis represents -log10(*P*) from PWAS findings; the dashed red line denotes the proteome-wide significance threshold at $P_{FDR}$<0.05.

A

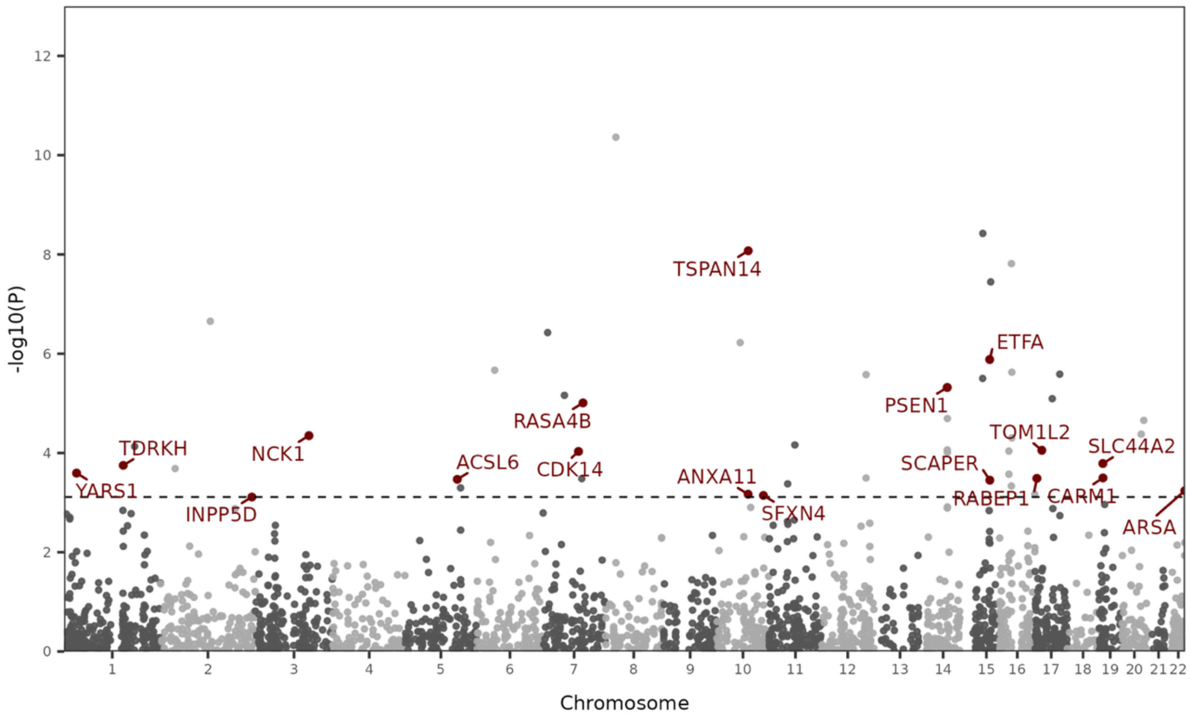
-log10(P)
Chromosome
TSPAN14
ETFA
PSEN1
RASA4B
TOM1L2
SLC44A2
TDRKH
NCK1
ACSL6
CDK14
SCAPER
ANXA11
YARS1
RABEP1
CARM1
INPP5D
SFXN4
ARSA

B

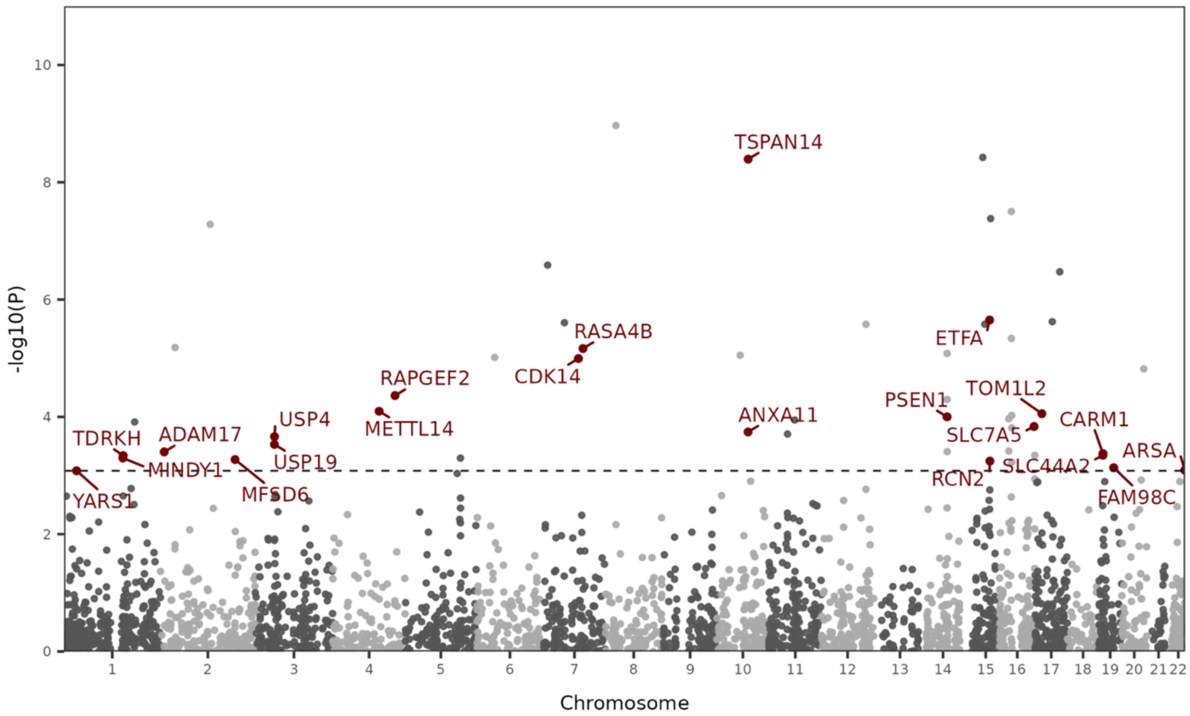
-log10(P)
Chromosome
TSPAN14
RASA4B
ETFA
RAPGEF2
CDK14
TOM1L2
PSEN1
ANXA11
USP4
METTL14
CARM1
TDRKH
ADAM17
SLC7A5
ARSA
USP19
MINDY1
SLC44A2
RCN2
YARS1
MFSD6
FAM98C

C

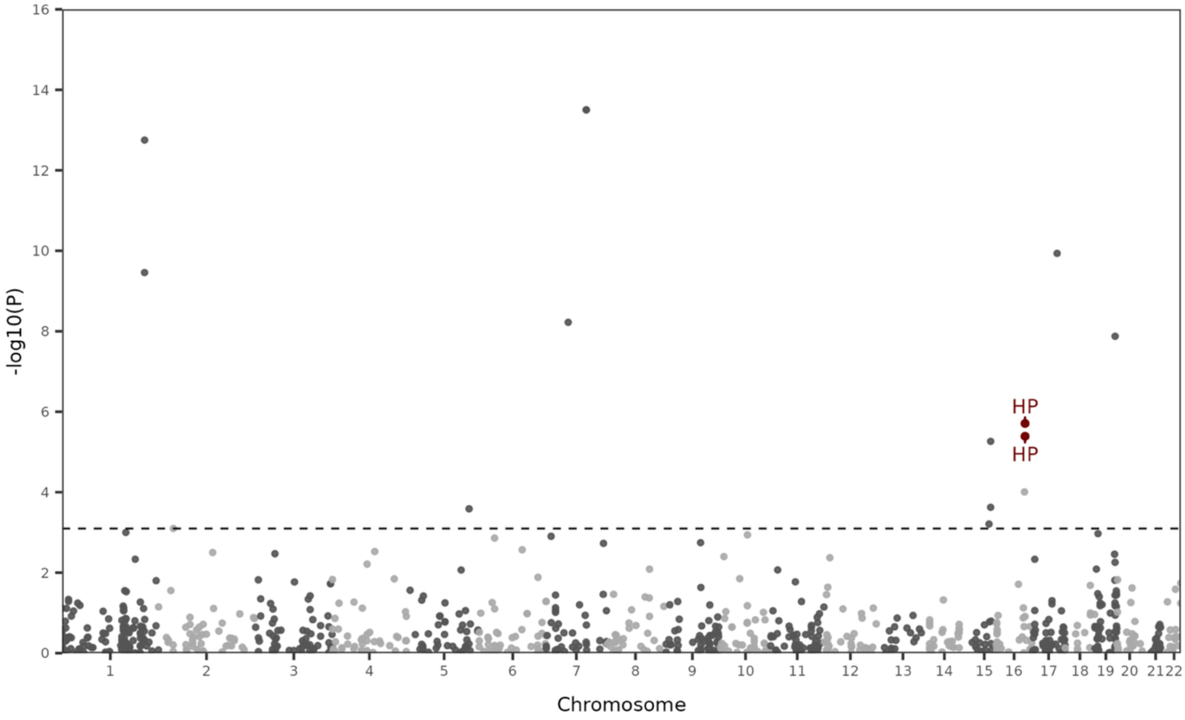

HP
HP
-log10(P)
Chromosome

D

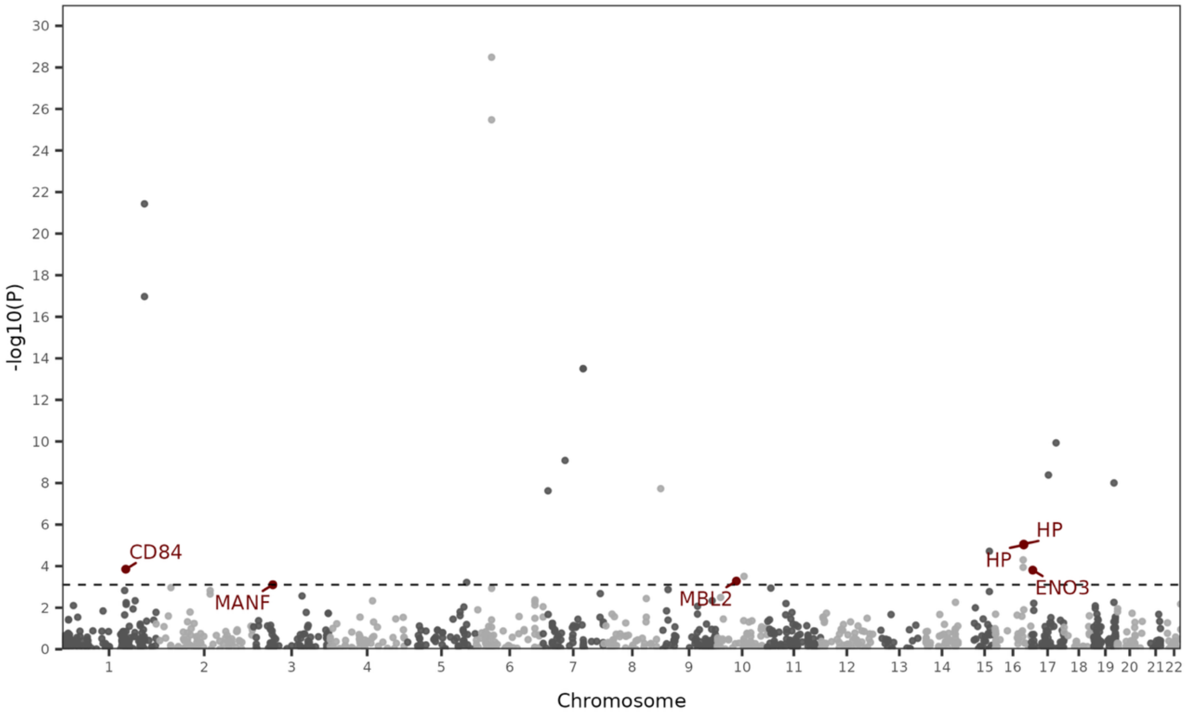

CD84
MANF
MBL2
HP
HP
ENO3
-log10(P)
Chromosome

**E**

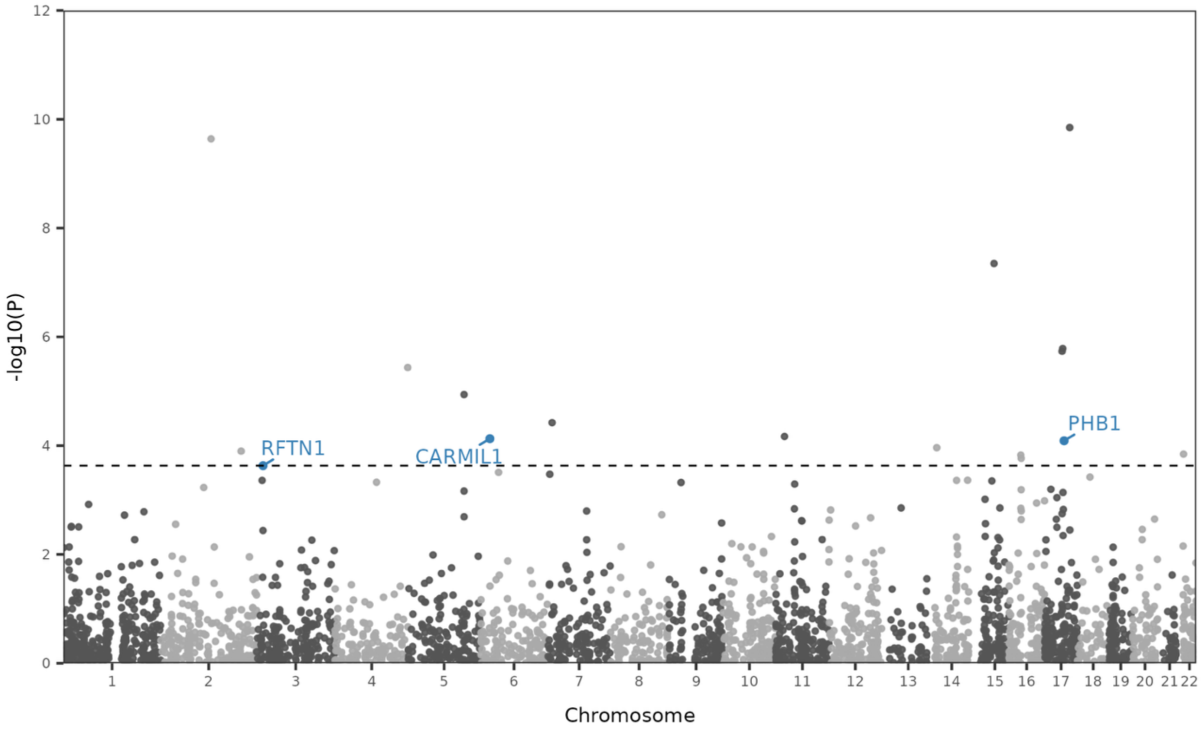

-log10(P)
Chromosome
RFTN1
CARMIL1
PHB1

**F**

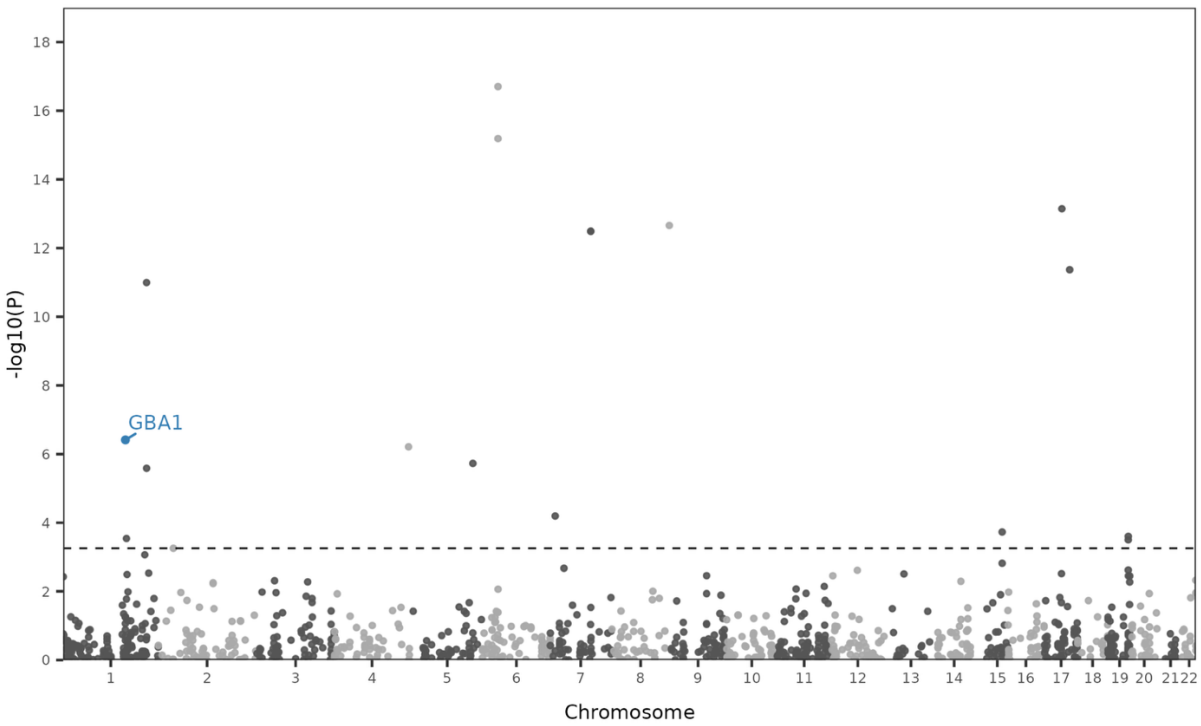

-log10(P)
Chromosome
GBA1

**Supplementary Figure 10. Sex-stratified EUR Alzheimer's disease PWAS: Sex-specific discoveries by sex and tissue. A)** Female primary brain discovery; **B)** Female secondary brain discovery; **C)** Female primary CSF discovery; **D)** Female secondary CSF discovery; **E)** Male secondary brain discovery; **F)** Male secondary CSF discovery. Datapoints in the Manhattan plots represent association tests between AD and a given protein in brain or aptamer in the CSF proteogenomic data. Only genes passing sex-heterogeneity filters are annotated and colored based on which sex the gene was prioritized in. Male primary discoveries in brain and CSF were excluded due to no sex-biased discoveries in the primary analyses. X-axis denotes genomic positions, and y-axis represents -log10($P$) from PWAS analyses; the dashed black line denotes the proteome-wide significance threshold at $P_{FDR}$<0.05.

#### Standard Discovery Filter (Female FDR.P < 0.05 & Male FDR.P < 0.05)

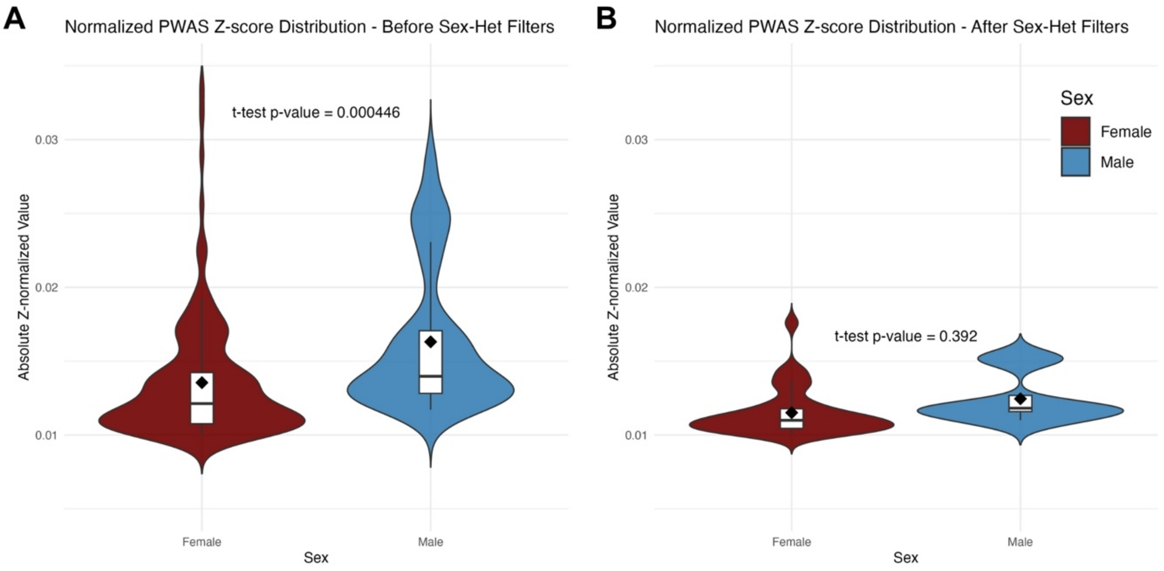

C

| Discovery | n_Genes (PWAS output) | n_Genes (P < 0.05) | n_Genes (FDR_P < 0.05) | n_Loci (FDR_P < 0.05) | n_Genes (sex-het) | n_Loci (sex-het) |
|---|---|---|---|---|---|---|
| Female Brain - Primary | 2418 | 276 | 49 | 39 | 18 | 15 |
| Male Brain - Primary | 1758 | 179 | 14 | 12 | 0 | 0 |
| Female Brain - Secondary | 2961 | 331 | 52 | 38 | 23 | 18 |
| Male Brain - Secondary | 2961 | 279 | 17 | 17 | 3 | 3 |
| Female CSF - Primary | 824 | 76 | 12 | 11 | 1 | 1 |
| Male CSF - Primary | 833 | 71 | 3 | 3 | 0 | 0 |
| Female CSF - Secondary | 1025 | 119 | 19 | 18 | 5 | 5 |
| Male CSF - Secondary | 1025 | 110 | 16 | 14 | 1 | 1 |

#### Lenient Male Discovery Filter (Female FDR.P < 0.05 & Male FDR.P < 0.10)

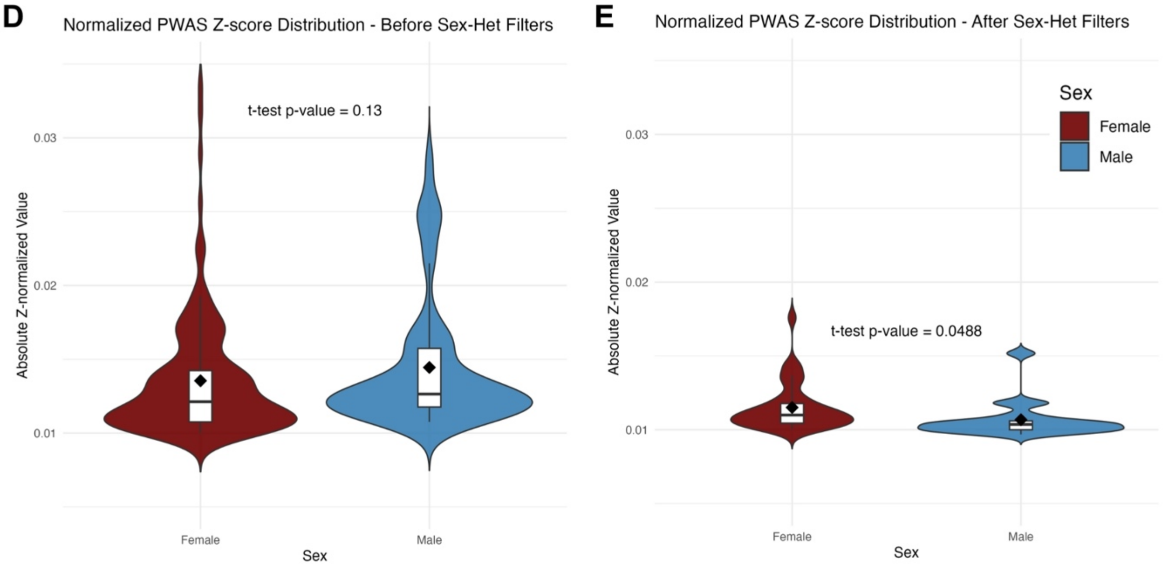

F

| Discovery | n_Genes (PWAS output) | n_Genes (P < 0.05) | n_Genes (Female FDR_P < 0.05)<br>n_Genes (Male FDR_P < 0.1) | n_Loci (Female FDR_P < 0.05)<br>n_Loci (Male FDR_P < 0.1) | n_Genes (sex-het) | n_Loci (sex-het) |
|---|---|---|---|---|---|---|
| Female Brain - Primary | 2418 | 276 | 49 | 39 | 18 | 15 |
| Male Brain - Primary | 1758 | 179 | 21 | 18 | 3 | 3 |
| Female Brain - Secondary | 2961 | 331 | 52 | 38 | 23 | 18 |
| Male Brain - Secondary | 2961 | 279 | 37 | 33 | 13 | 13 |
| Female CSF - Primary | 824 | 76 | 12 | 11 | 1 | 1 |
| Male CSF - Primary | 833 | 71 | 6 | 5 | 1 | 1 |
| Female CSF - Secondary | 1025 | 119 | 19 | 18 | 5 | 5 |
| Male CSF - Secondary | 1025 | 110 | 17 | 15 | 2 | 2 |

**Supplementary Figure 11. Comparison of normalized PWAS z-scores across sex-specific Alzheimer's disease discovery filters.** To assess potential power bias for increased female PWAS discoveries, we normalized PWAS Z-scores by the square root of female and male effective sample sizes respectively. **A)** Comparison of female and male normalized z-scores before sex-heterogeneity filters in standard discovery PWAS (Female $P_{FDR}$<0.05 , Male $P_{FDR}$<0.05), **B)** comparison after applying sex-het filters, and **C)** table summarizing the gene counts and significant loci from A and B. **D)** Comparison of female and male normalized z-scores before sex-heterogeneity filters in lenient male discovery PWAS (Female $P_{FDR}$<0.05, Male $P_{FDR}$<0.1), **E)** comparison after sex-het filters, and **F)** table summarizing the gene counts and significant loci from D and E. **Conclusion**: Female PWAS normalized Z-scores tended to be smaller suggesting more power in female analyses, but even when making discoveries in men more lenient and balancing normalized Z-scores across sexes, more discoveries were still made in women compared to men.

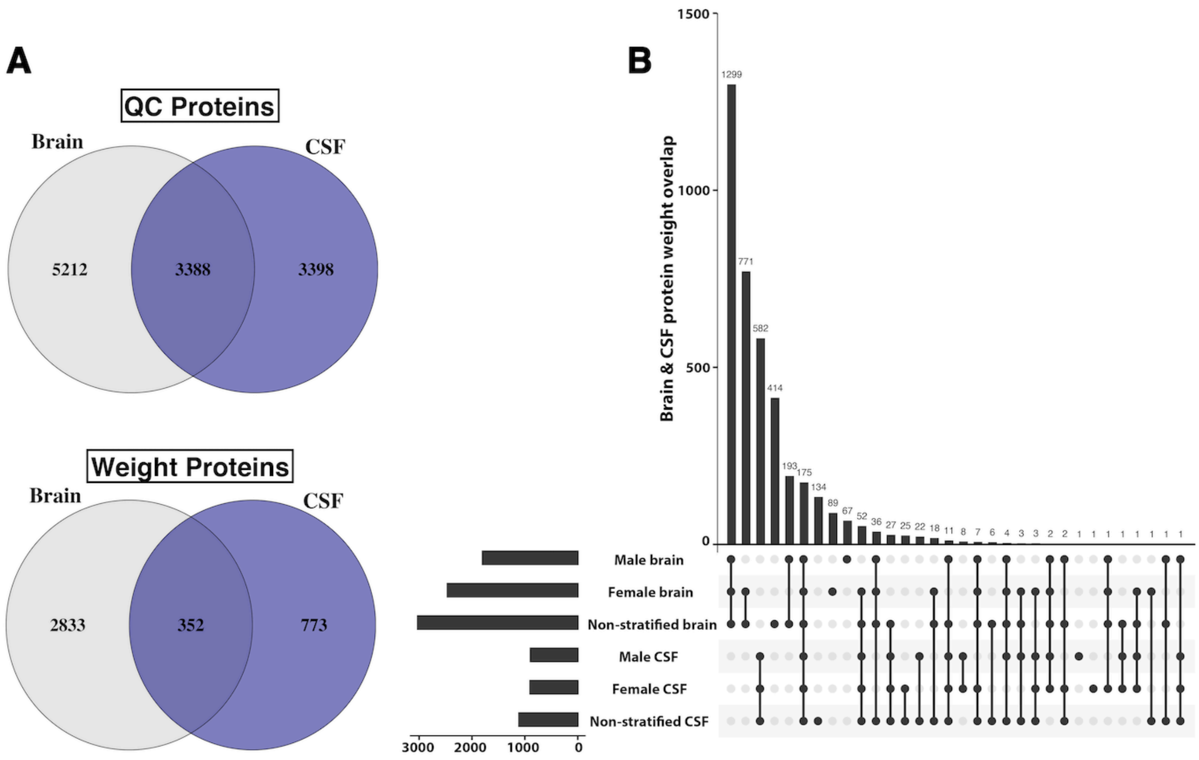

**Supplementary Figure 12. EUR AD PWAS: Protein overlap in brain versus CSF. A)** Top Venn-diagram shows the overlap between brain and CSF proteins that passed quality control (QC) and were available for calculating protein weights for *cis* genetic variants. 28.2% of all proteins passing QC overlapped between brain and CSF proteomic data. Bottom Venn-diagram illustrates the overlap between brain and CSF protein for which weights could be generated. 9% of all proteins with weights were shared between brain and CSF. **B)** UpSet plot of overlap between sex-stratified and non-sex-stratified brain and CSF proteins that had protein weights generated for *cis* genetic variants.

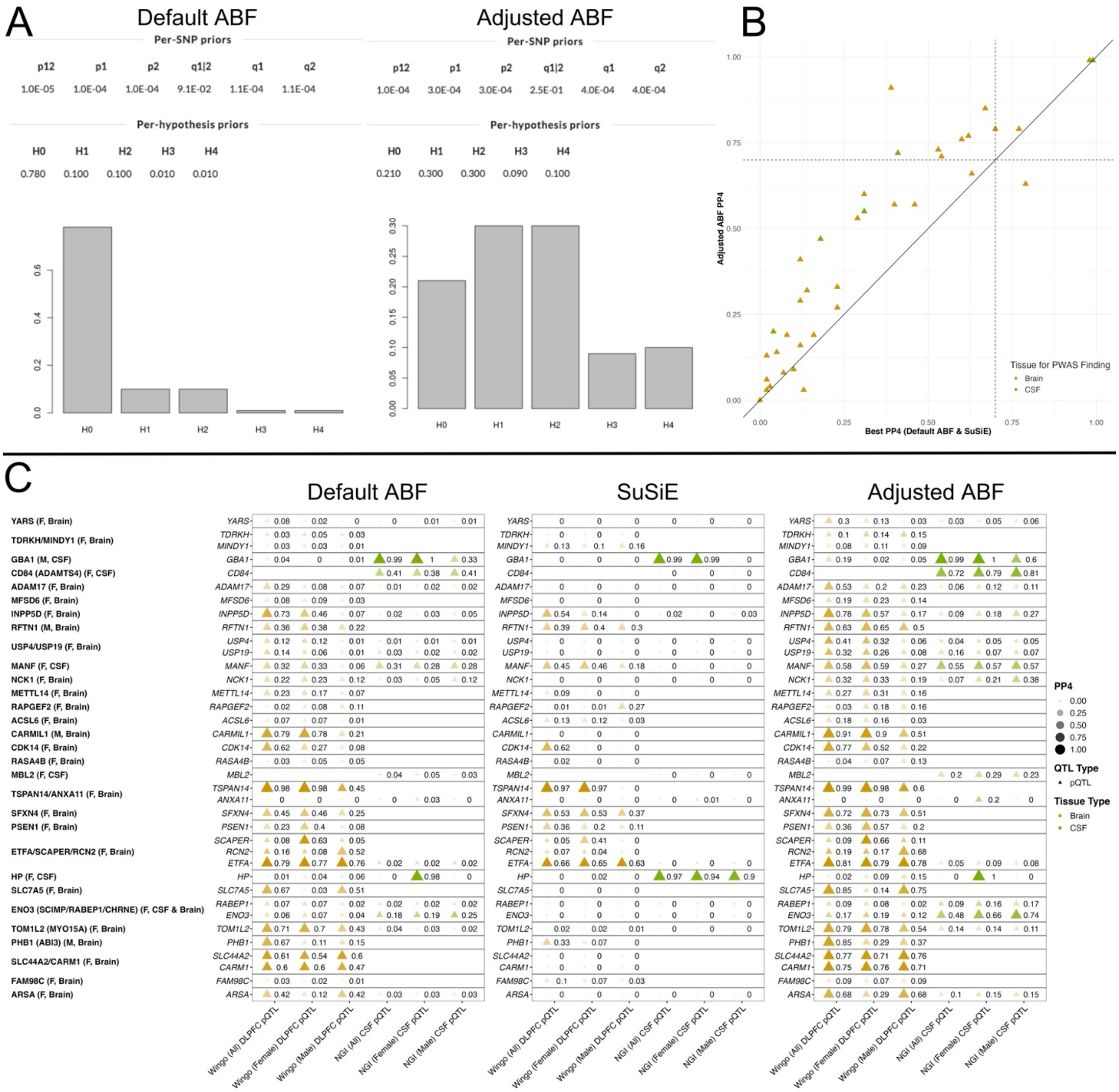
A
Default ABF
Per-SNP priors
p12 p1 p2 q1|2 q1 q2
1.0E-05 1.0E-04 1.0E-04 9.1E-02 1.1E-04 1.1E-04
Per-hypothesis priors
H0 H1 H2 H3 H4
0.780 0.100 0.100 0.010 0.010
Adjusted ABF
Per-SNP priors
p12 p1 p2 q1|2 q1 q2
1.0E-04 3.0E-04 3.0E-04 2.5E-01 4.0E-04 4.0E-04
Per-hypothesis priors
H0 H1 H2 H3 H4
0.210 0.300 0.300 0.090 0.100
B
Adjusted ABF PP4
Best PP4 (Default ABF & SuSiE)
Tissue for PWAS Finding
Brain
CSF
C
Default ABF
SuSiE
Adjusted ABF
YARS (F, Brain)
TDRKH/MINDY1 (F, Brain)
GBA1 (M, CSF)
CD84 (ADAMTS4) (F, CSF)
ADAM17 (F, Brain)
MFSD6 (F, Brain)
INPP5D (F, Brain)
RFTN1 (M, Brain)
USP4/USP19 (F, Brain)
MANF (F, CSF)
NCK1 (F, Brain)
METTL14 (F, Brain)
RAPGEF2 (F, Brain)
ACSL6 (F, Brain)
CARMIL1 (M, Brain)
CDK14 (F, Brain)
RASA4B (F, Brain)
MBL2 (F, CSF)
TSPAN14/ANXA11 (F, Brain)
SFXN4 (F, Brain)
PSEN1 (F, Brain)
ETFA/SCAPER/RCN2 (F, Brain)
HP (F, CSF)
SLC7A5 (F, Brain)
ENO3 (SCIMP/RABEP1/CHRNE) (F, CSF & Brain)
TOM1L2 (MYO15A) (F, Brain)
PHB1 (ABI3) (M, Brain)
SLC44A2/CARM1 (F, Brain)
FAM98C (F, Brain)
ARSA (F, Brain)
Wingo (All) DLPFC pQTL
Wingo (Female) DLPFC pQTL
Wingo (Male) DLPFC pQTL
NGI (All) CSF pQTL
NGI (Female) CSF pQTL
NGI (Male) CSF pQTL
PP4
0.00
0.25
0.50
0.75
1.00
QTL Type
pQTL
Tissue Type
Brain
CSF

**Supplementary Figure 13. Brain and CSF pQTL colocalization analyses matching to sex-specific Alzheimer's disease PWAS discoveries.** Beyond standard ABF and SuSiE colocalization analyses, we also performed "adjusted" ABF analyses with more lenient priors to increase the assumption that signals exist in both the AD GWAS and PWAS-matched pQTL data. This is merited by the fact that significant PWAS findings require there to be some minimal signal in both the AD GWAS and pQTL loci, as well as the fact that our PWAS approach sought to leverage subthreshold AD GWAS signals ($P$>5e-8) which could reduce sensitivity of ABF colocalization analyses with standard priors. **A)** Illustration of the default versus adjusted ABF priors. **B)** Comparison of the best PP4 values from standard ABF or SuSiE colocalization analyses versus adjusted ABF colocalization analyses, for the AD GWAS and pQTL matches. **C)** All PP4 values for different colocalization approaches for significant sex-specific PWAS genes across all strata in brain and CSF pQTL data.

## S14.1

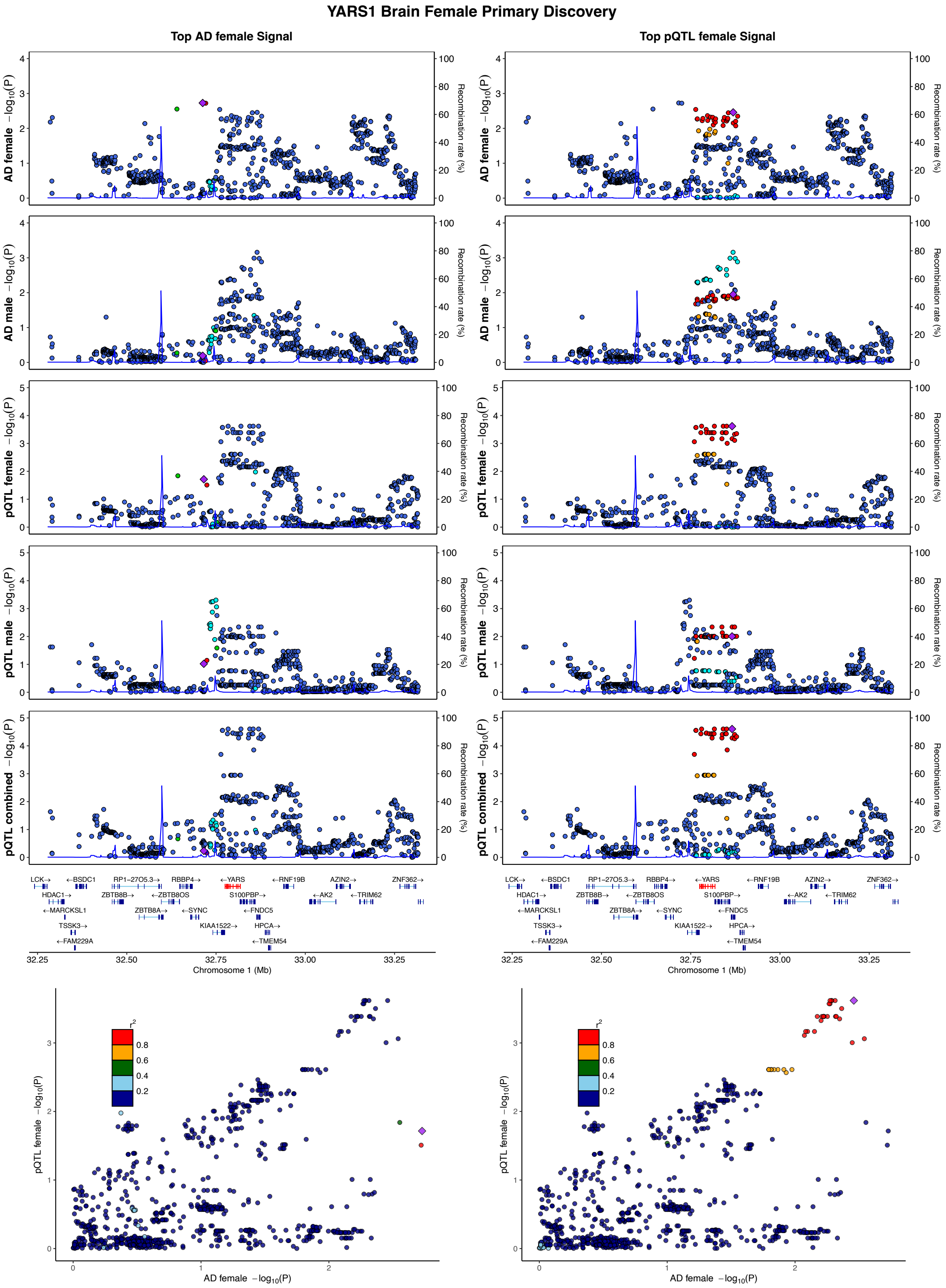

YARS1 Brain Female Primary Discovery
Top AD female Signal
Top pQTL female Signal
AD female −log10(P)
AD male −log10(P)
pQTL female −log10(P)
pQTL male −log10(P)
pQTL combined −log10(P)
Recombination rate (%)
LCK→
←BSDC1
RP1−27O5.3→
RBBP4→
←YARS
←RNF19B
AZIN2→
ZNF362→
HDAC1→
ZBTB8B→
←ZBTB8OS
S100PBP→
←AK2
←TRIM62
←MARCKSL1
ZBTB8A→
←SYNC
←FNDC5
TSSK3→
KIAA1522→
HPCA→
←FAM229A
←TMEM54
Chromosome 1 (Mb)
r2
pQTL female −log10(P)
AD female −log10(P)

## S14.2

##### MINDY1 Brain Female Secondary Discovery

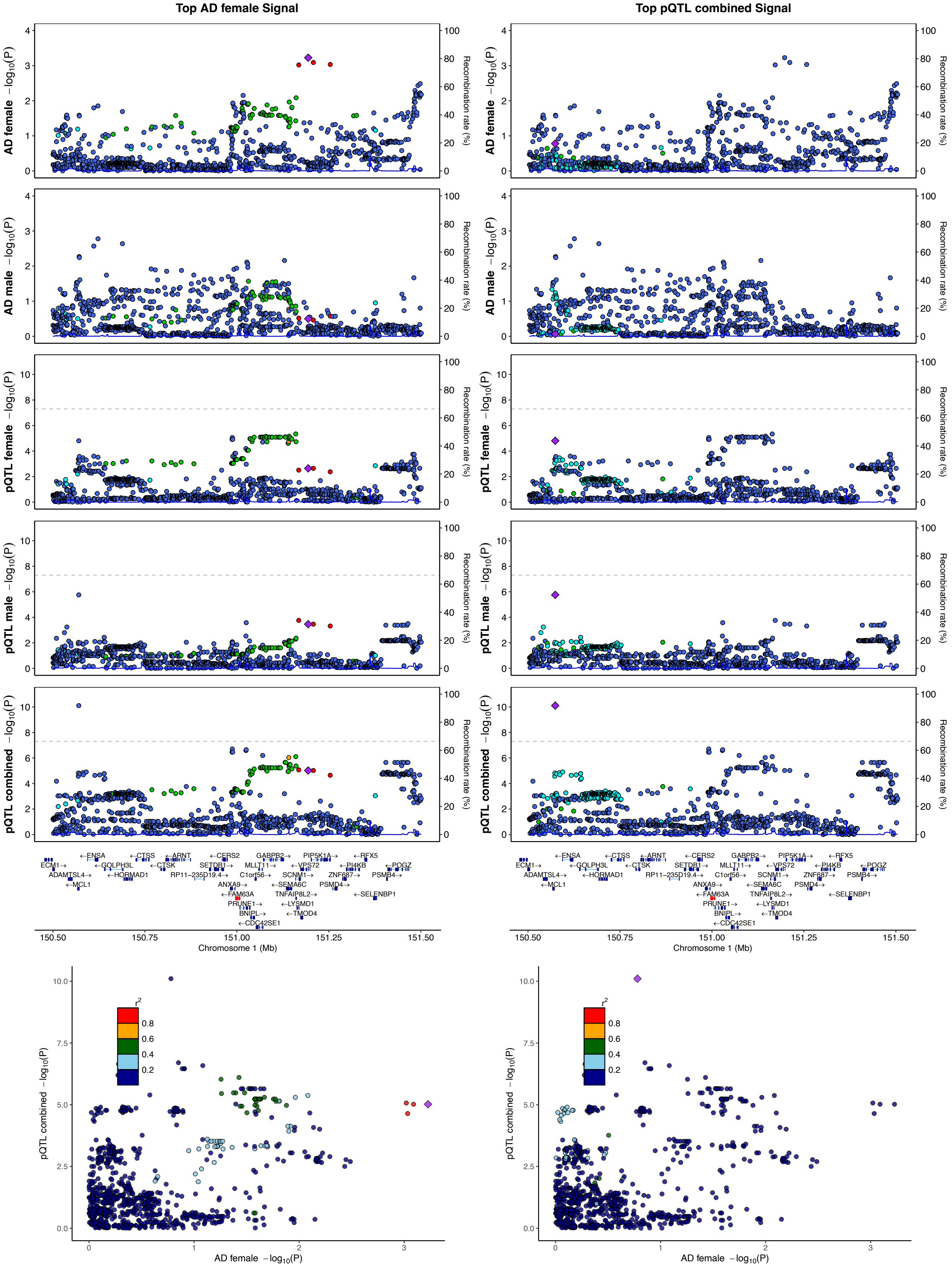

## S14.3

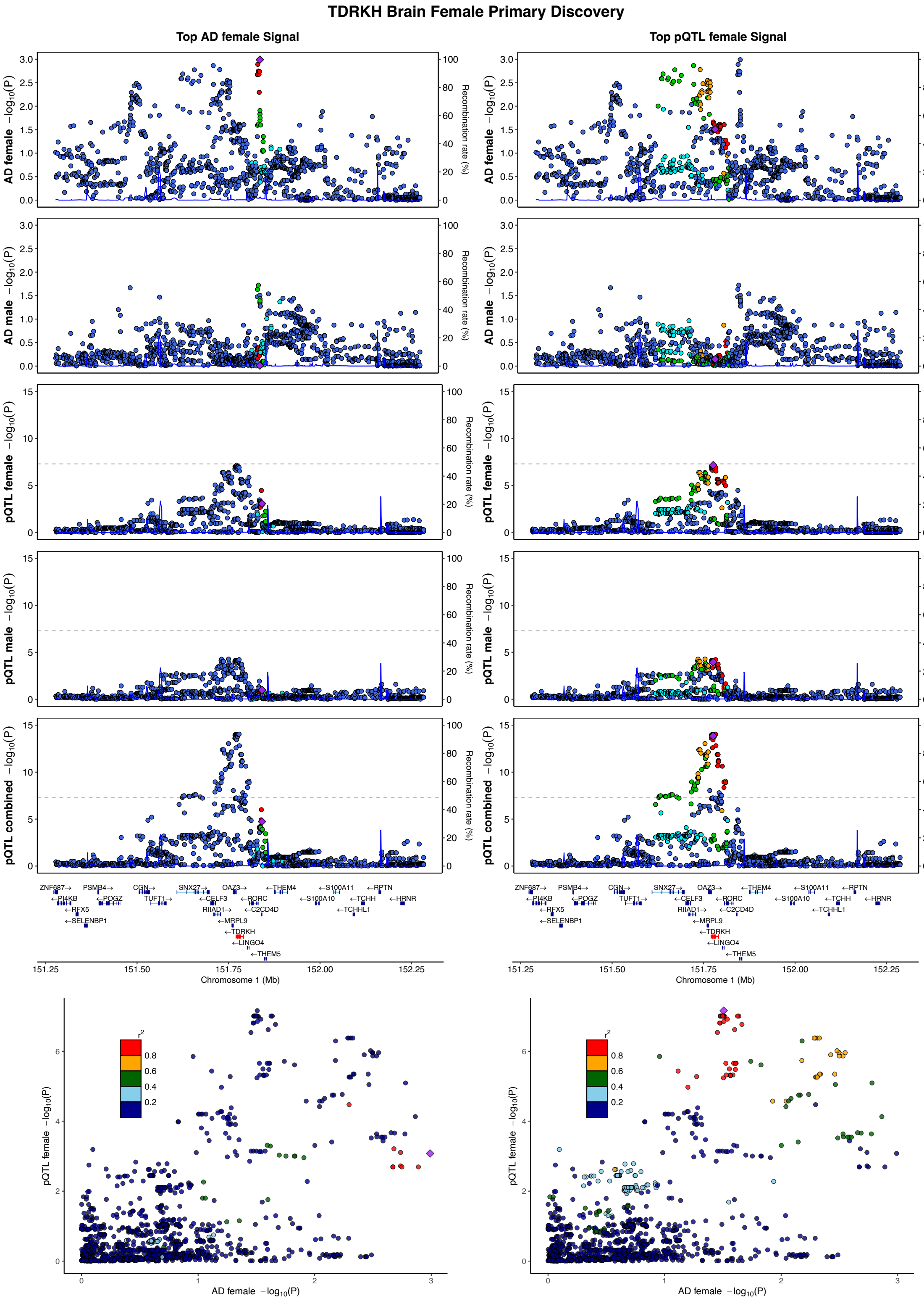
TDRKH Brain Female Primary Discovery
Top AD female Signal
Top pQTL female Signal
AD female −log10(P)
AD male −log10(P)
pQTL female −log10(P)
pQTL male −log10(P)
pQTL combined −log10(P)
Recombination rate (%)
Chromosome 1 (Mb)
AD female −log10(P)
pQTL female −log10(P)
r2
ZNF687→
PSMB4→
CGN→
SNX27→
OAZ3→
←THEM4
←S100A11
←RPTN
←PI4KB
←POGZ
TUFT1→
←CELF3
←RORC
←S100A10
←TCHH
←HRNR
←RFX5
RIIAD1→
←C2CD4D
←TCHHL1
←SELENBP1
←MRPL9
←TDRKH
←LINGO4
←THEM5

## S14.4

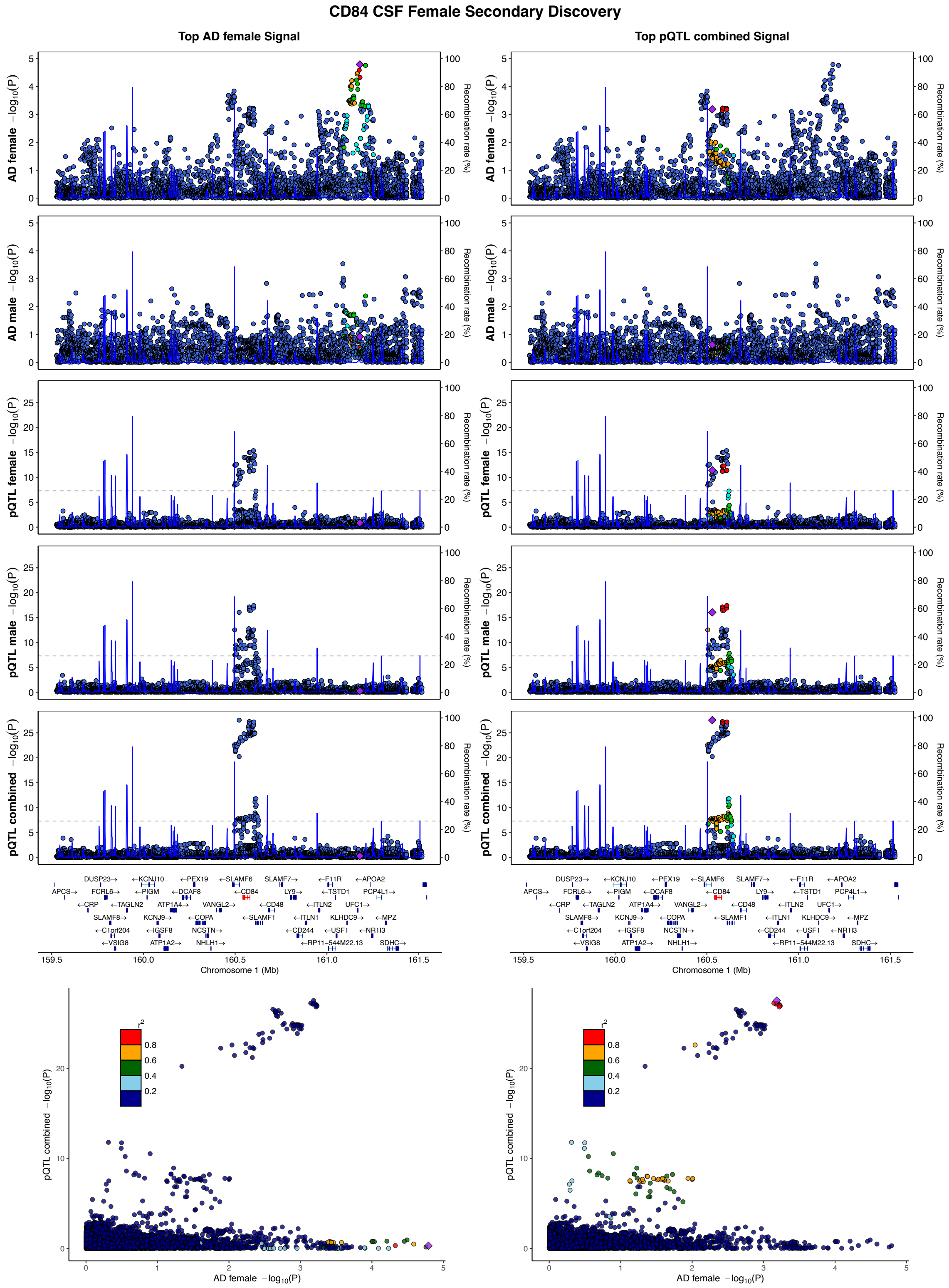
CD84 CSF Female Secondary Discovery
Top AD female Signal
Top pQTL combined Signal
AD female −log10(P)
AD male −log10(P)
pQTL female −log10(P)
pQTL male −log10(P)
pQTL combined −log10(P)
Recombination rate (%)
Chromosome 1 (Mb)
AD female −log10(P)
r2

## S14.5

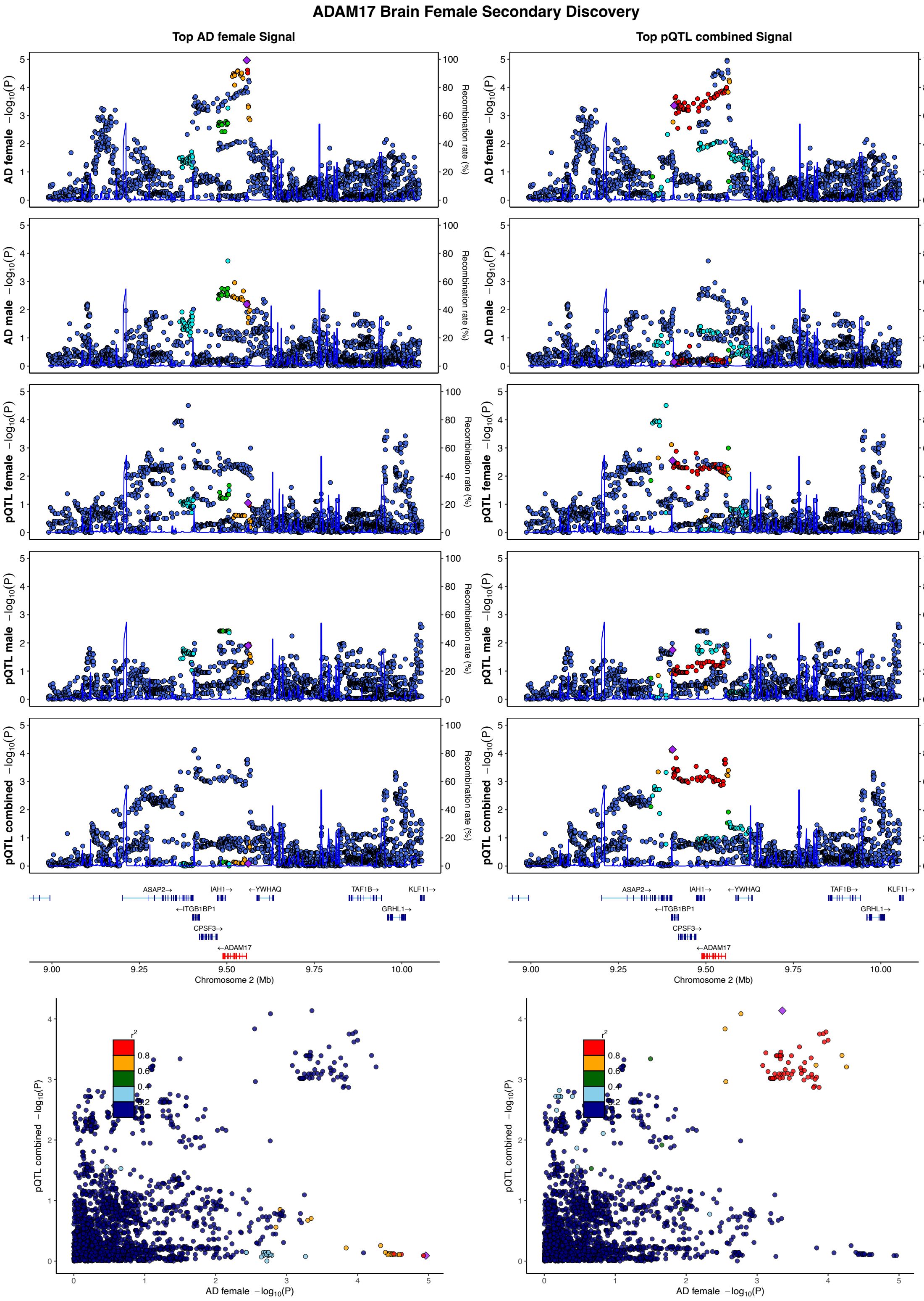
ADAM17 Brain Female Secondary Discovery
Top AD female Signal
Top pQTL combined Signal
AD female −log10(P)
AD male −log10(P)
pQTL female −log10(P)
pQTL male −log10(P)
pQTL combined −log10(P)
Recombination rate (%)
ASAP2→
IAH1→
←YWHAQ
TAF1B→
KLF11→
←ITGB1BP1
GRHL1→
CPSF3→
←ADAM17
Chromosome 2 (Mb)
AD female −log10(P)
r2

## S14.6

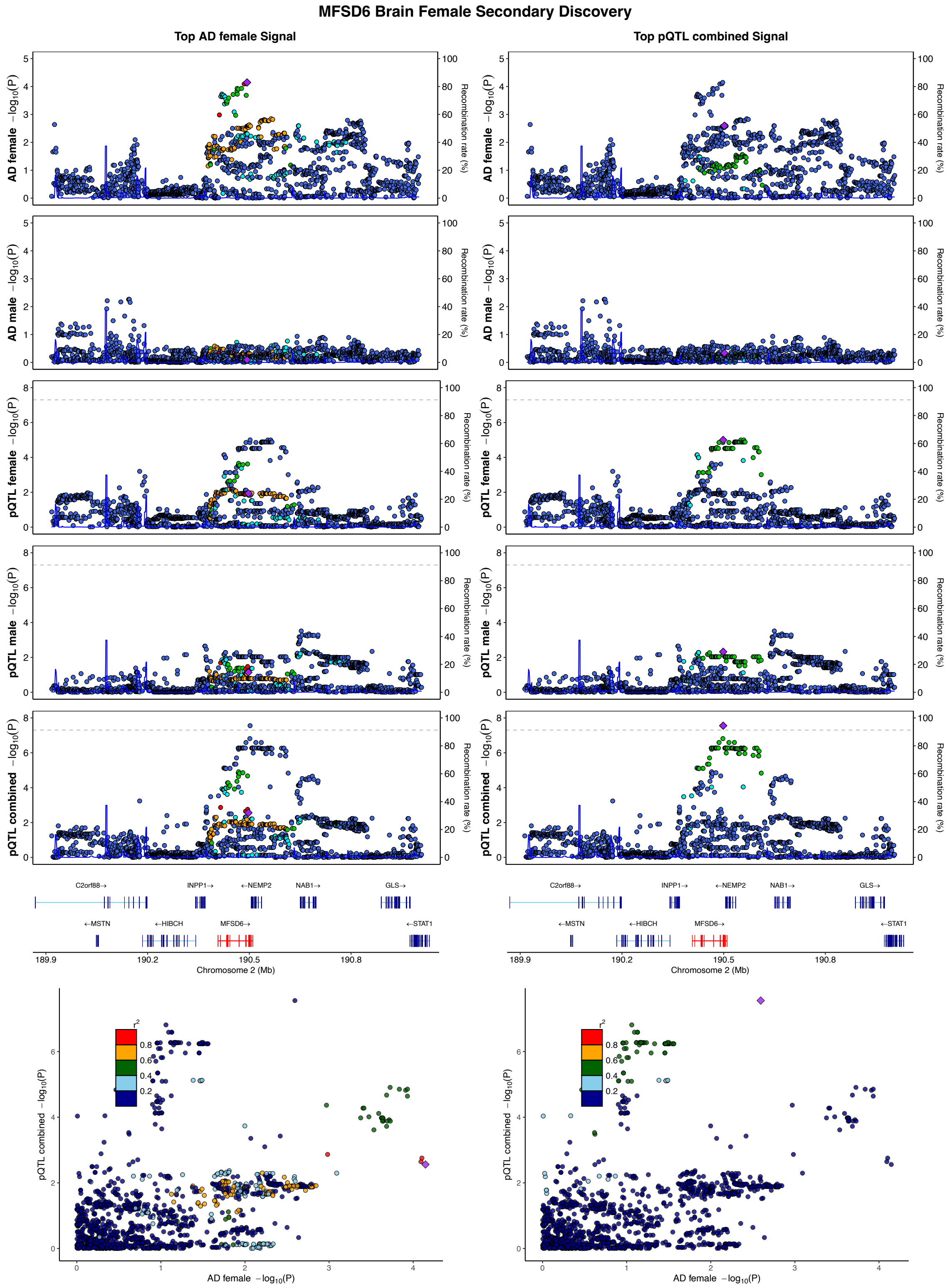

MFSD6 Brain Female Secondary Discovery
Top AD female Signal
Top pQTL combined Signal
AD female −log10(P)
AD male −log10(P)
pQTL female −log10(P)
pQTL male −log10(P)
pQTL combined −log10(P)
Recombination rate (%)
C2orf88→
INPP1→
←NEMP2
NAB1→
GLS→
←MSTN
←HIBCH
MFSD6→
←STAT1
Chromosome 2 (Mb)
AD female −log10(P)
r2

## S14.7

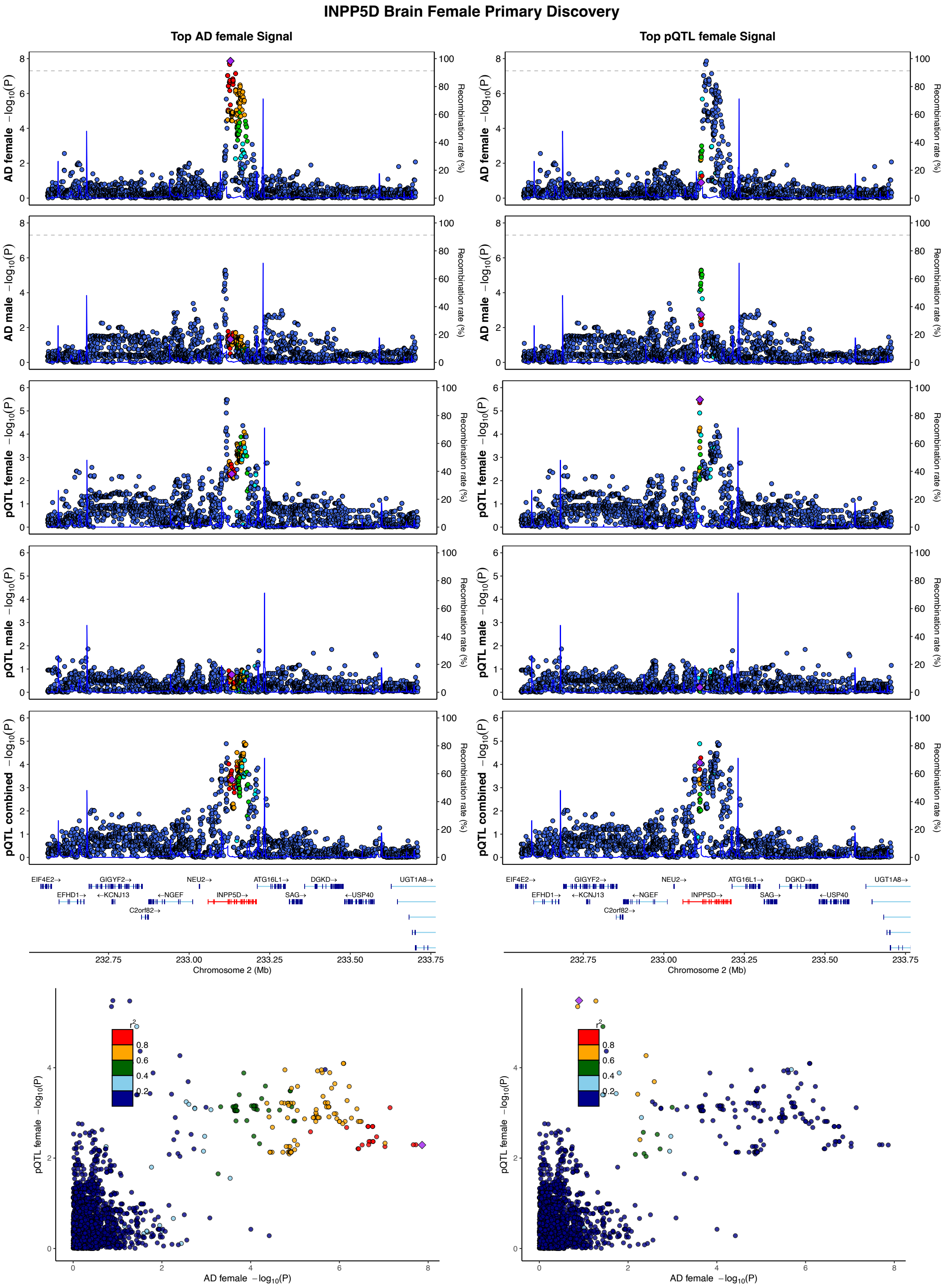
INPP5D Brain Female Primary Discovery
Top AD female Signal
Top pQTL female Signal
AD female −log10(P)
AD male −log10(P)
pQTL female −log10(P)
pQTL male −log10(P)
pQTL combined −log10(P)
Recombination rate (%)
EIF4E2→
GIGYF2→
NEU2→
ATG16L1→
DGKD→
UGT1A8→
EFHD1→
←KCNJ13
←NGEF
INPP5D→
SAG→
←USP40
C2orf82→
Chromosome 2 (Mb)
AD female −log10(P)
pQTL female −log10(P)
r2

## S14.8

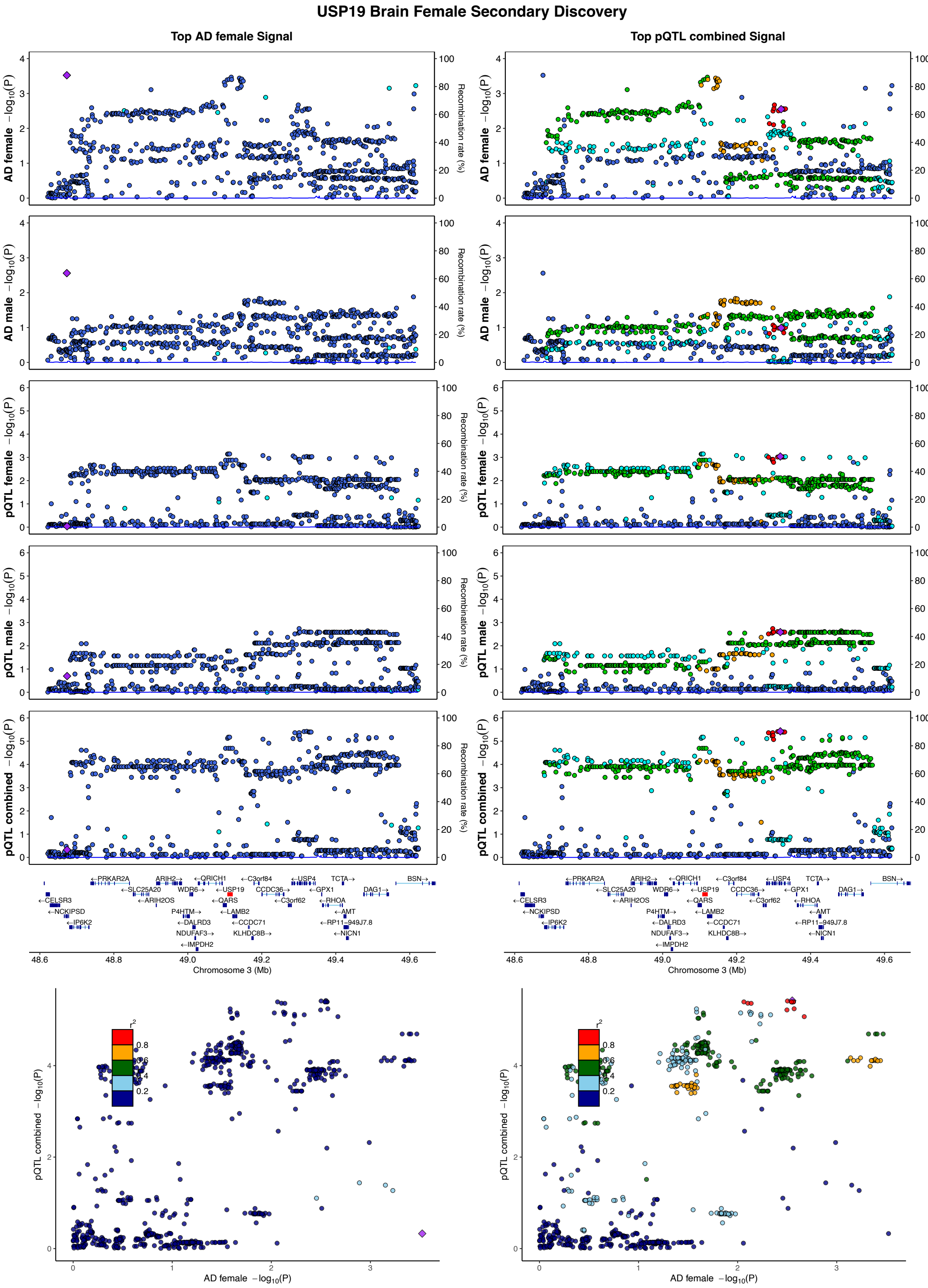

USP19 Brain Female Secondary Discovery
Top AD female Signal
Top pQTL combined Signal
AD female −log10(P)
AD male −log10(P)
pQTL female −log10(P)
pQTL male −log10(P)
pQTL combined −log10(P)
Recombination rate (%)
Chromosome 3 (Mb)
AD female −log10(P)
r2

## S14.9

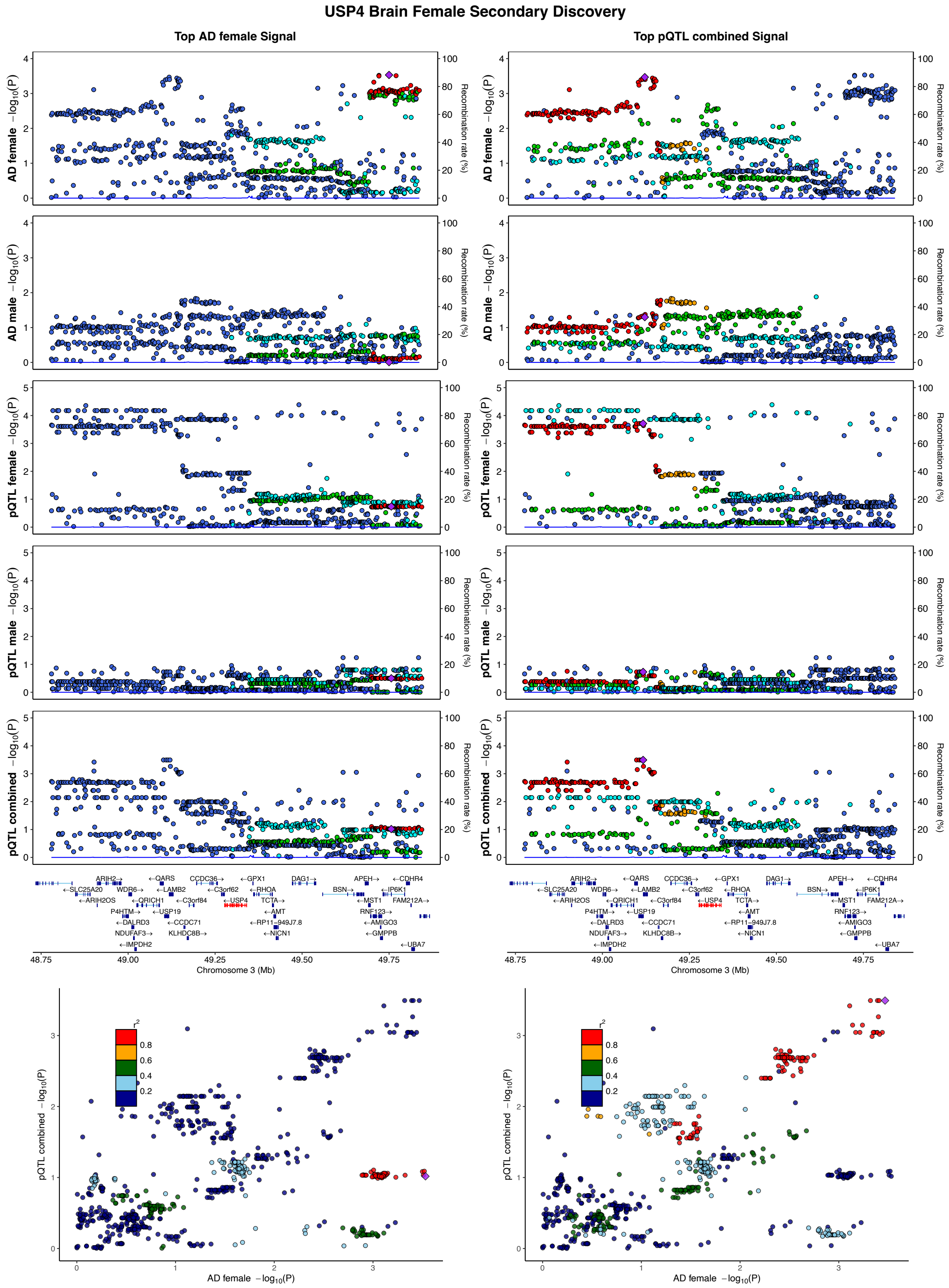

USP4 Brain Female Secondary Discovery
Top AD female Signal
Top pQTL combined Signal
AD female −log10(P)
AD male −log10(P)
pQTL female −log10(P)
pQTL male −log10(P)
pQTL combined −log10(P)
Recombination rate (%)
Chromosome 3 (Mb)
AD female −log10(P)
r2

## S14.10

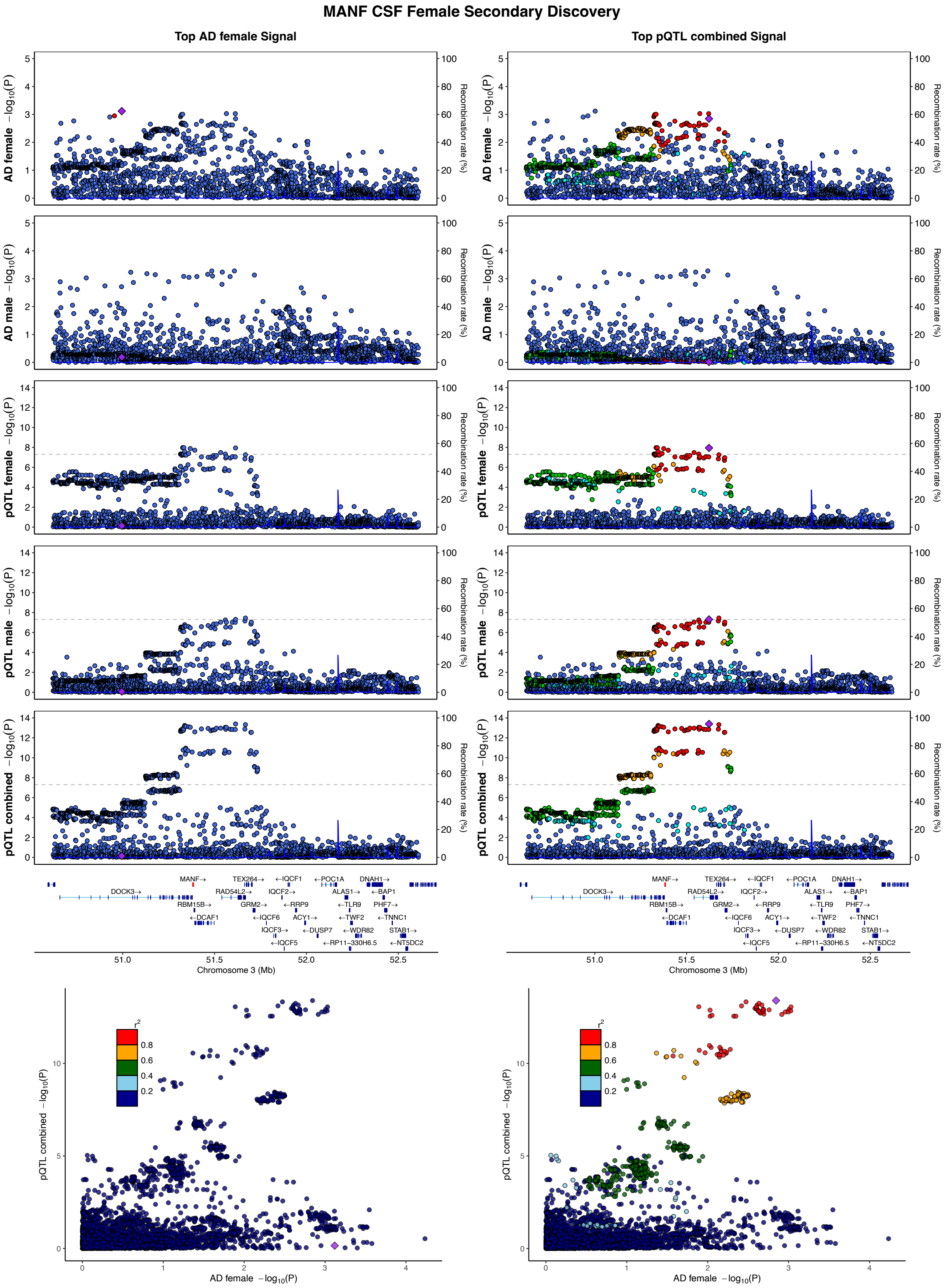

MANF CSF Female Secondary Discovery
Top AD female Signal
Top pQTL combined Signal
AD female −log10(P)
AD male −log10(P)
pQTL female −log10(P)
pQTL male −log10(P)
pQTL combined −log10(P)
Recombination rate (%)
Chromosome 3 (Mb)
AD female −log10(P)
pQTL combined −log10(P)
r2

## S14.11

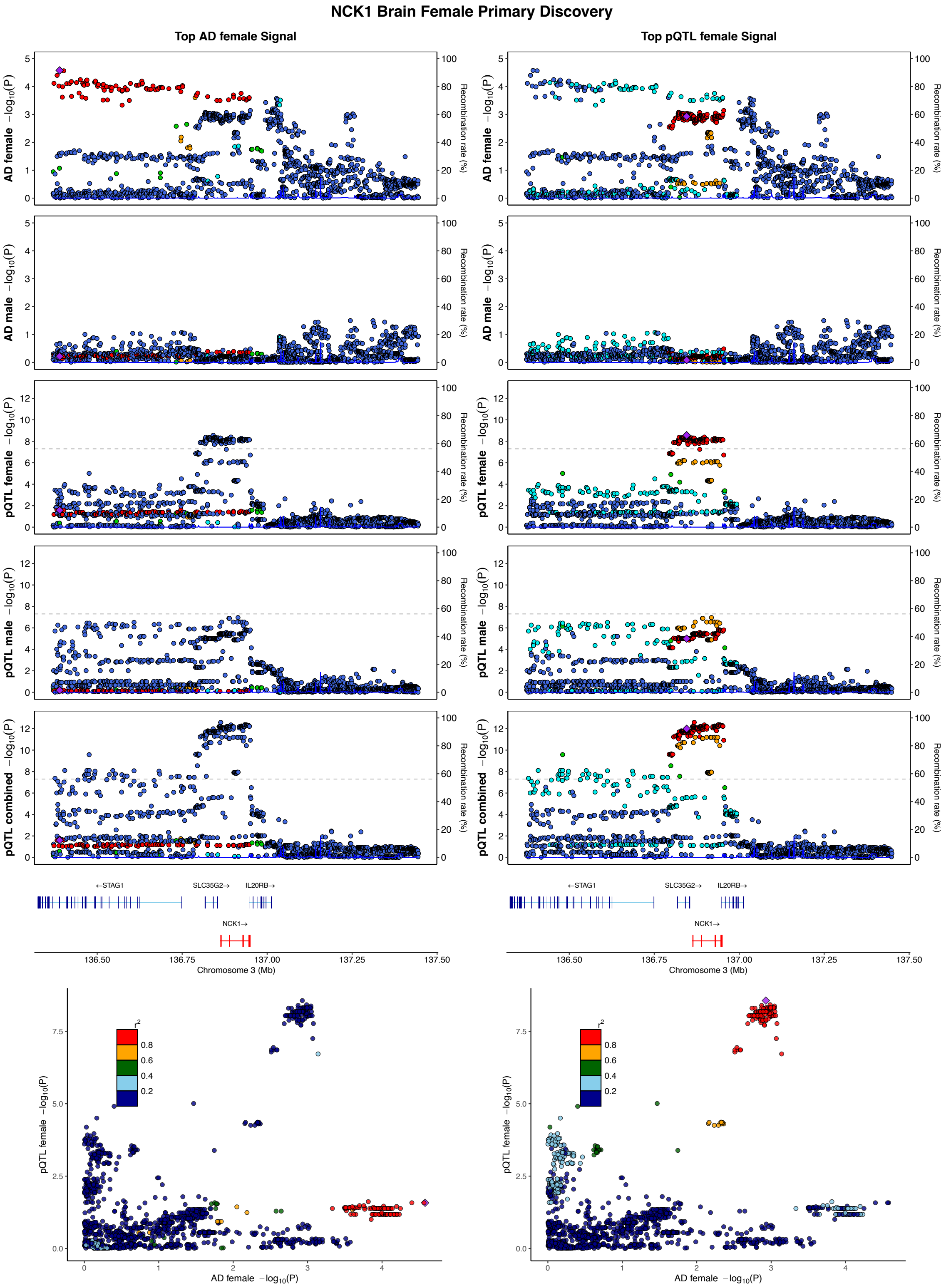
NCK1 Brain Female Primary Discovery
Top AD female Signal
Top pQTL female Signal
AD female −log10(P)
AD male −log10(P)
pQTL female −log10(P)
pQTL male −log10(P)
pQTL combined −log10(P)
Recombination rate (%)
←STAG1
SLC35G2→
IL20RB→
NCK1→
Chromosome 3 (Mb)
136.50
136.75
137.00
137.25
137.50
pQTL female −log10(P)
AD female −log10(P)
r2
0.8
0.6
0.4
0.2

## S14.12

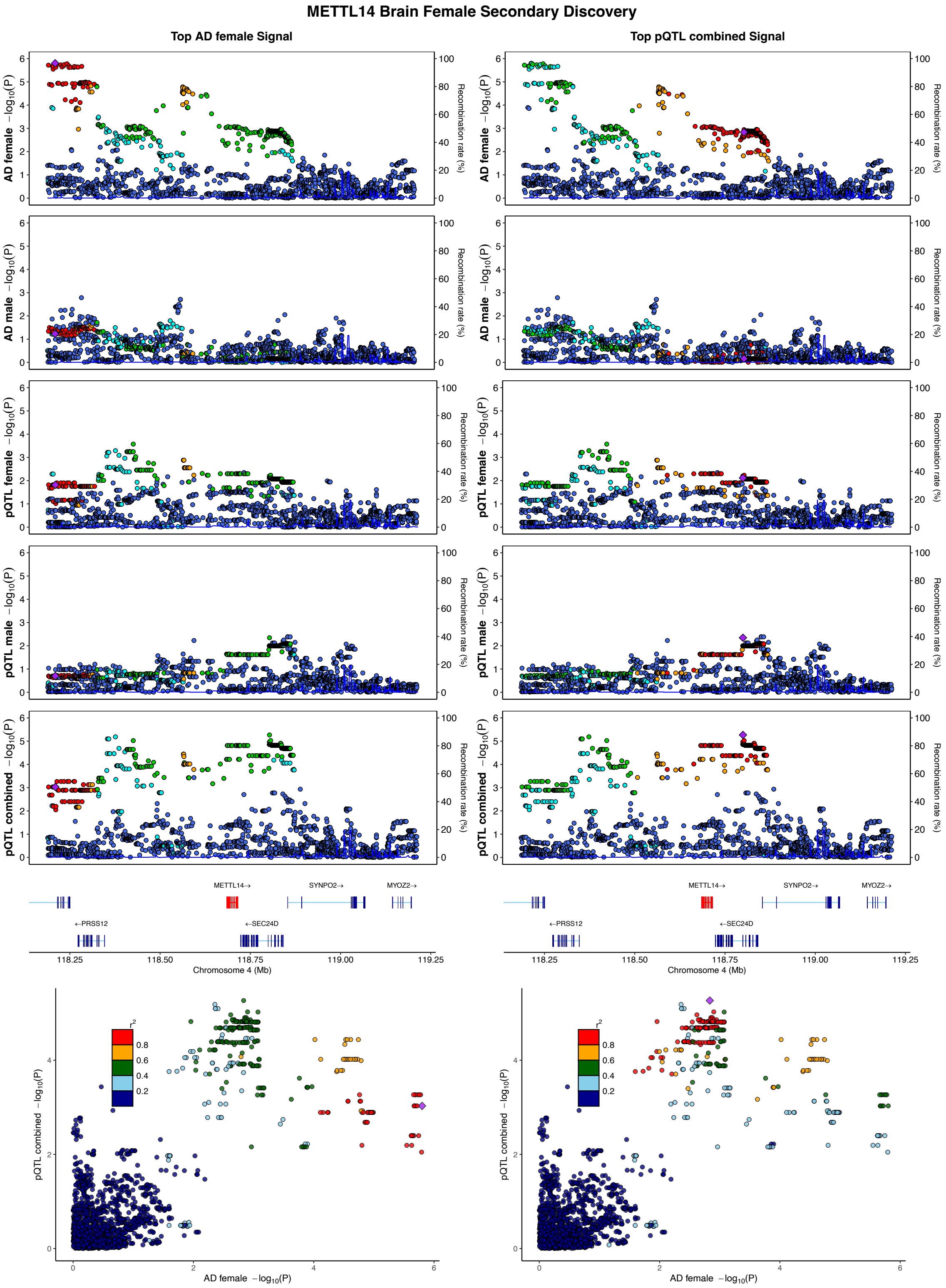
METTL14 Brain Female Secondary Discovery
Top AD female Signal
Top pQTL combined Signal
AD female −log10(P)
AD male −log10(P)
pQTL female −log10(P)
pQTL male −log10(P)
pQTL combined −log10(P)
Recombination rate (%)
METTL14→
SYNPO2→
MYOZ2→
←PRSS12
←SEC24D
Chromosome 4 (Mb)
118.25
118.50
118.75
119.00
119.25
r2
0.8
0.6
0.4
0.2
AD female −log10(P)

## S14.13

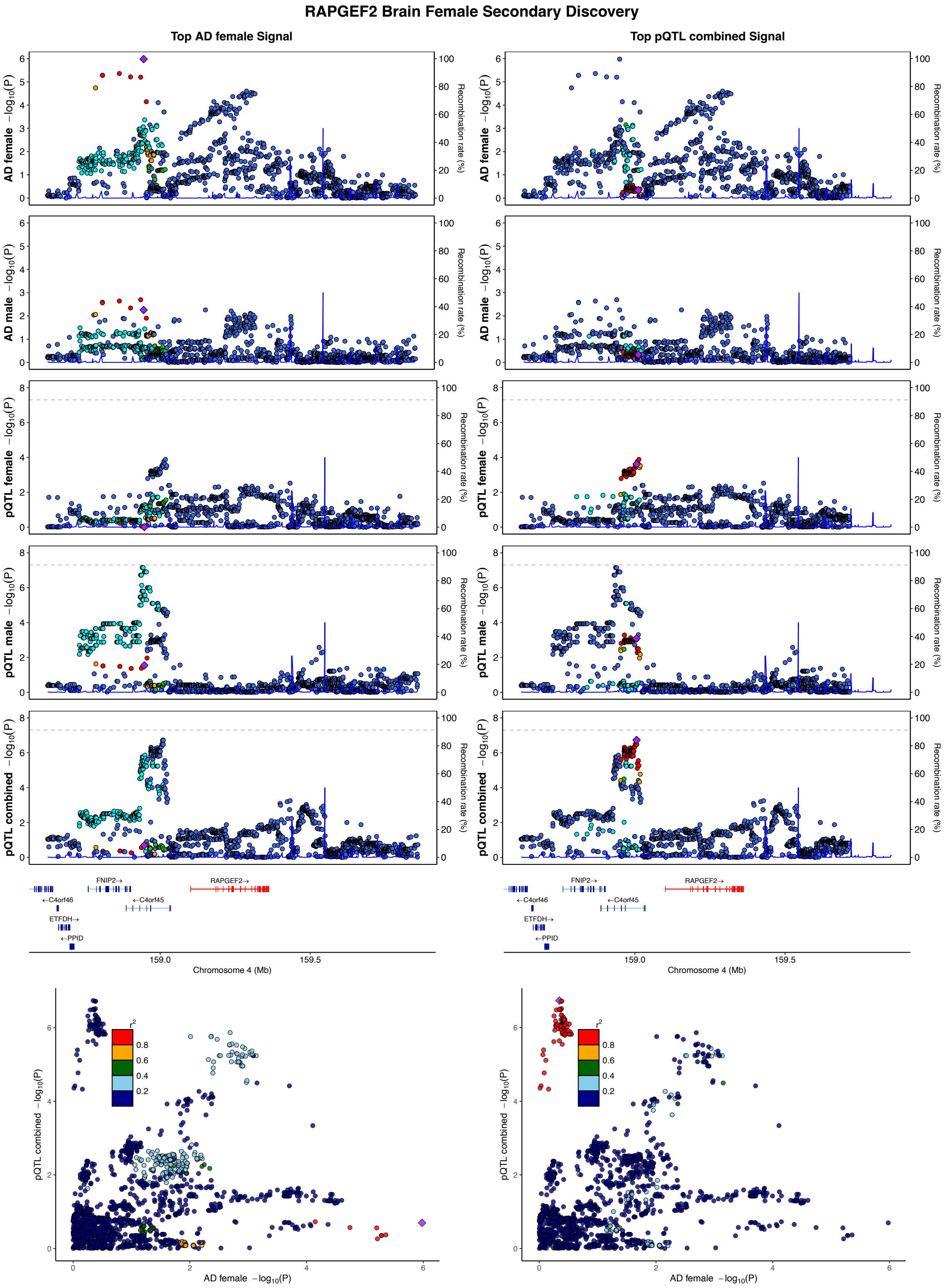
RAPGEF2 Brain Female Secondary Discovery
Top AD female Signal
Top pQTL combined Signal
AD female −log10(P)
AD male −log10(P)
pQTL female −log10(P)
pQTL male −log10(P)
pQTL combined −log10(P)
Recombination rate (%)
FNIP2→
RAPGEF2→
←C4orf46
←C4orf45
ETFDH→
←PPID
159.0
159.5
Chromosome 4 (Mb)
r2
0.8
0.6
0.4
0.2
pQTL combined −log10(P)
AD female −log10(P)

## S14.14

##### ACSL6 Brain Female Primary Discovery

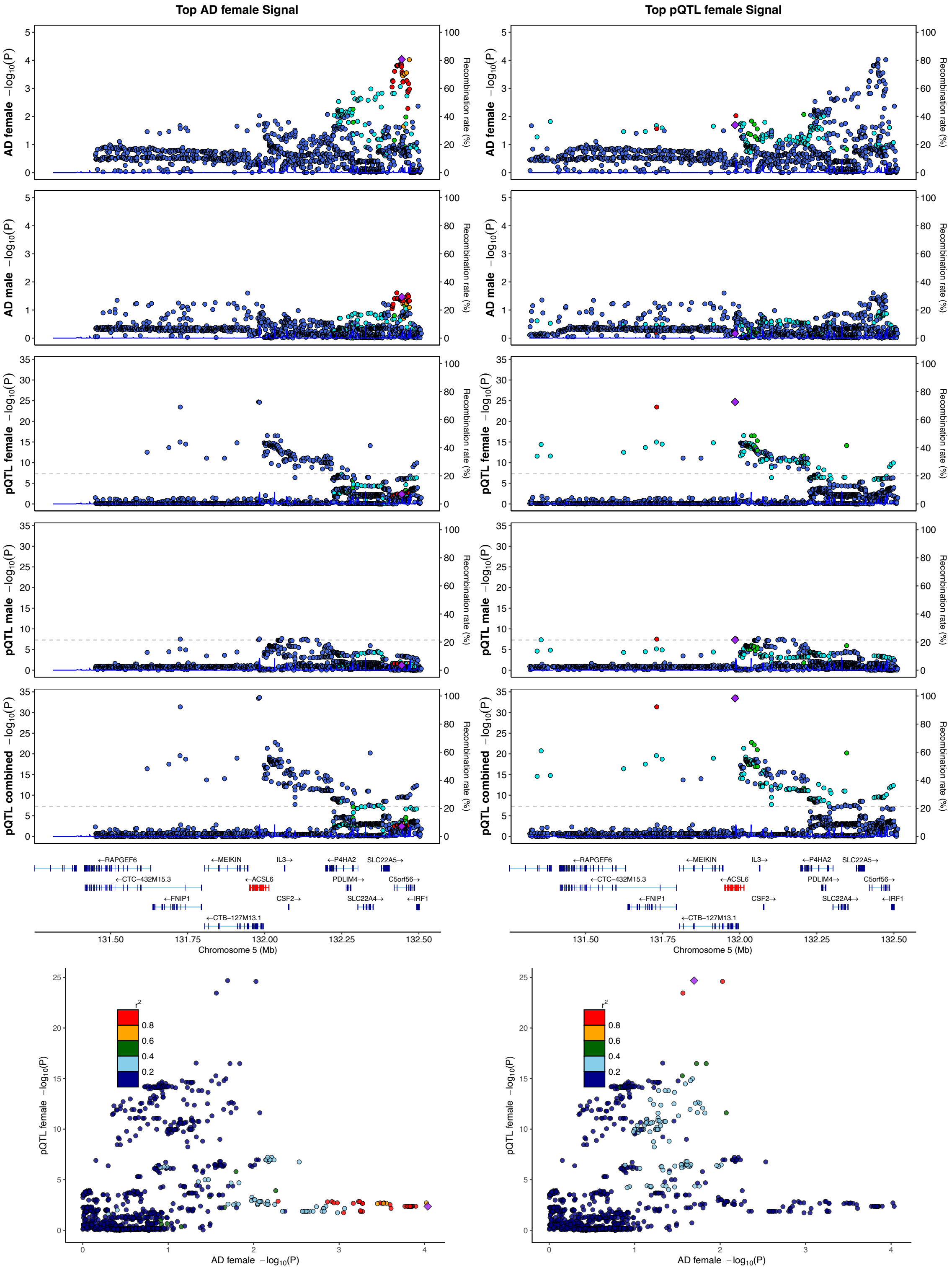

## S14.15

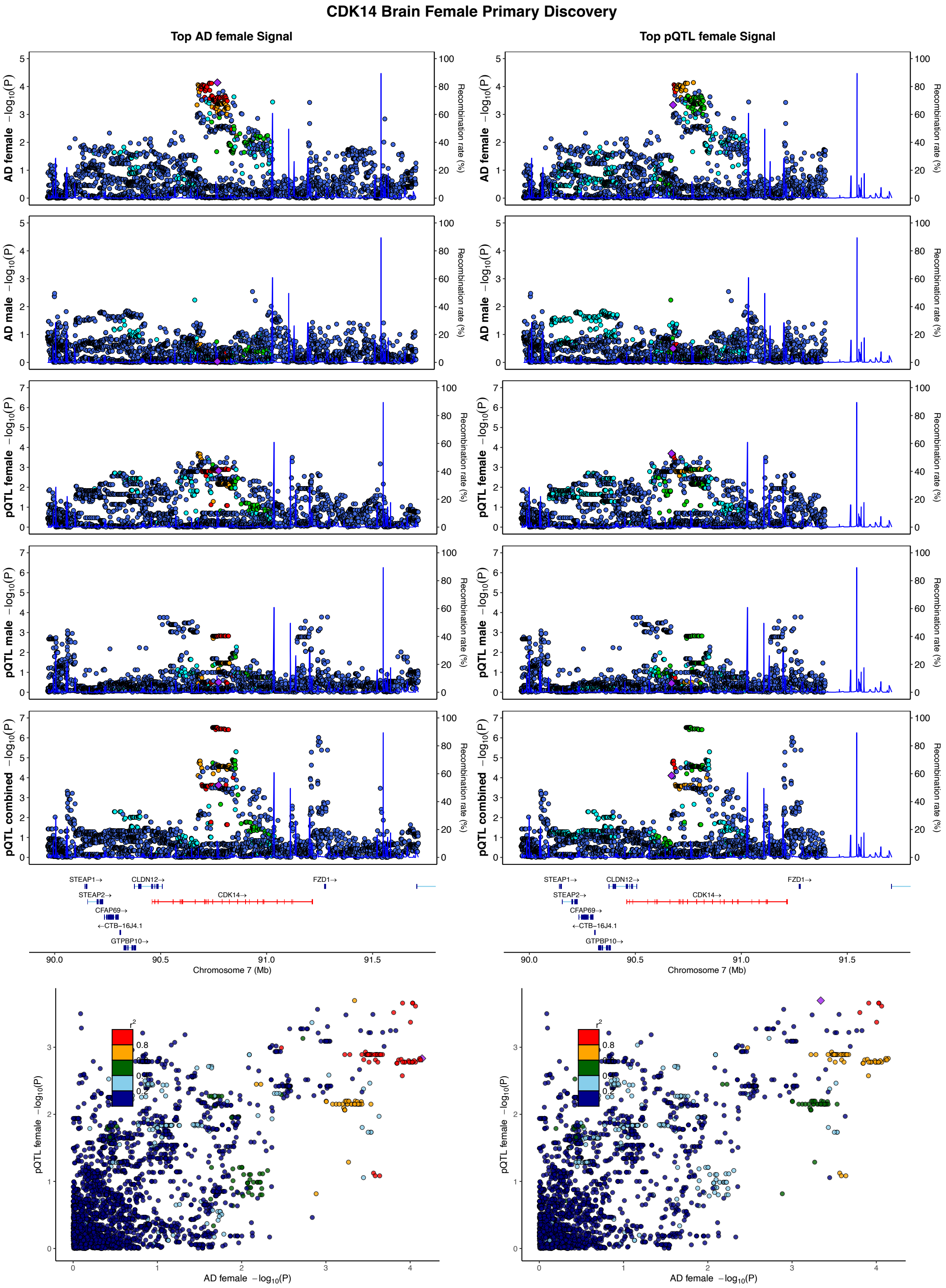

CDK14 Brain Female Primary Discovery
Top AD female Signal
Top pQTL female Signal
AD female −log10(P)
AD male −log10(P)
pQTL female −log10(P)
pQTL male −log10(P)
pQTL combined −log10(P)
Recombination rate (%)
STEAP1→
STEAP2→
CFAP69→
←CTB−16J4.1
GTPBP10→
CLDN12→
CDK14→
FZD1→
Chromosome 7 (Mb)
AD female −log10(P)
r2

## S14.16

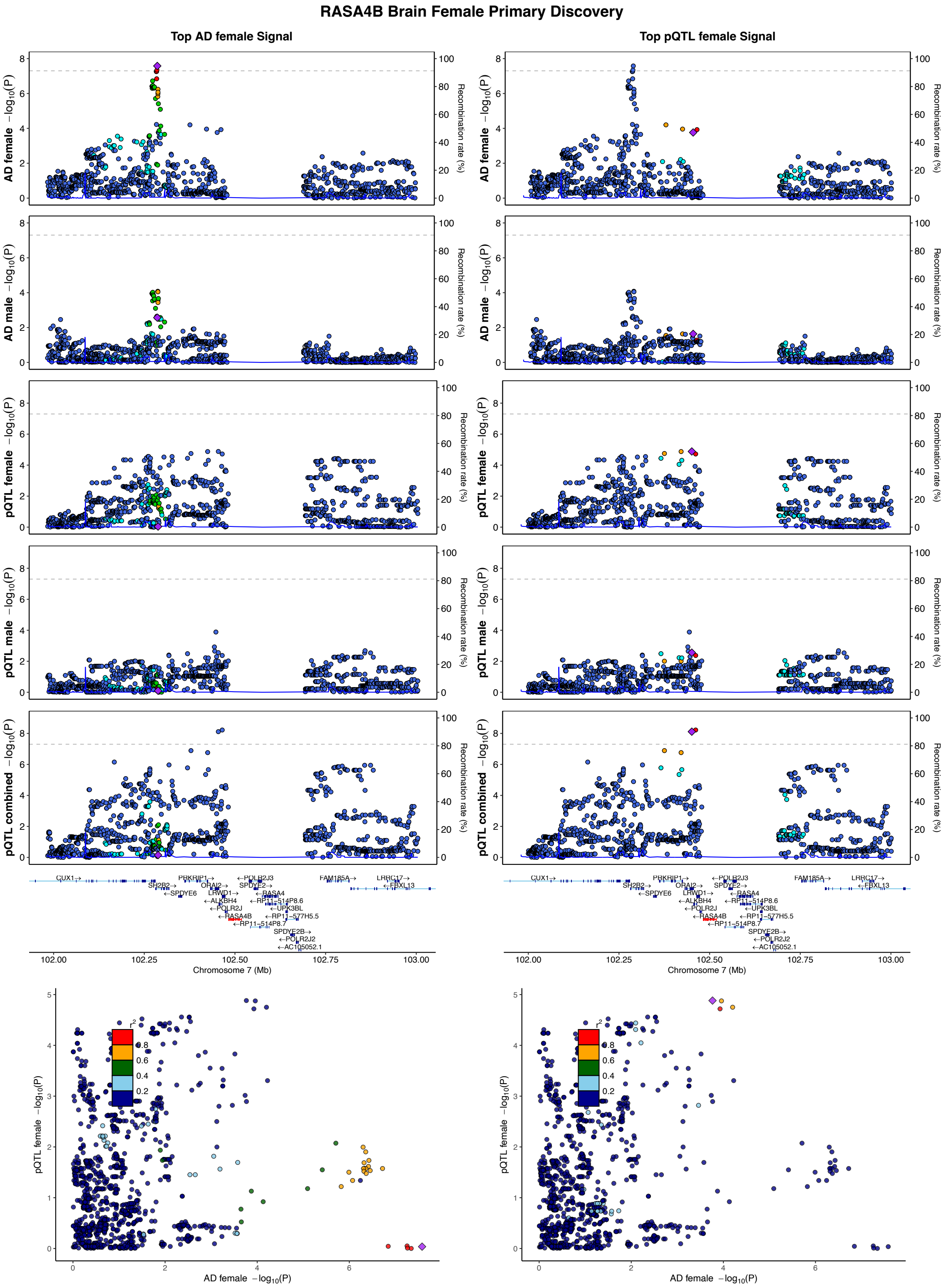

RASA4B Brain Female Primary Discovery
Top AD female Signal
Top pQTL female Signal
AD female −log10(P)
AD male −log10(P)
pQTL female −log10(P)
pQTL male −log10(P)
pQTL combined −log10(P)
Recombination rate (%)
Chromosome 7 (Mb)
CUX1
SH2B2
SPDYE6
PRKRIP1
ORAI2
LHFPL3
ALKBH4
POLR2J
RASA4B
RP11-514P8.7
POLR2J3
SPDYE2
RASA4
RP11-514P8.6
UPK3BL
RP11-577H5.5
SPDYE2B
POLR2J2
AC105052.1
FAM185A
LRRC17
FBXL13
AD female −log10(P)
pQTL female −log10(P)
r2

## S14.17

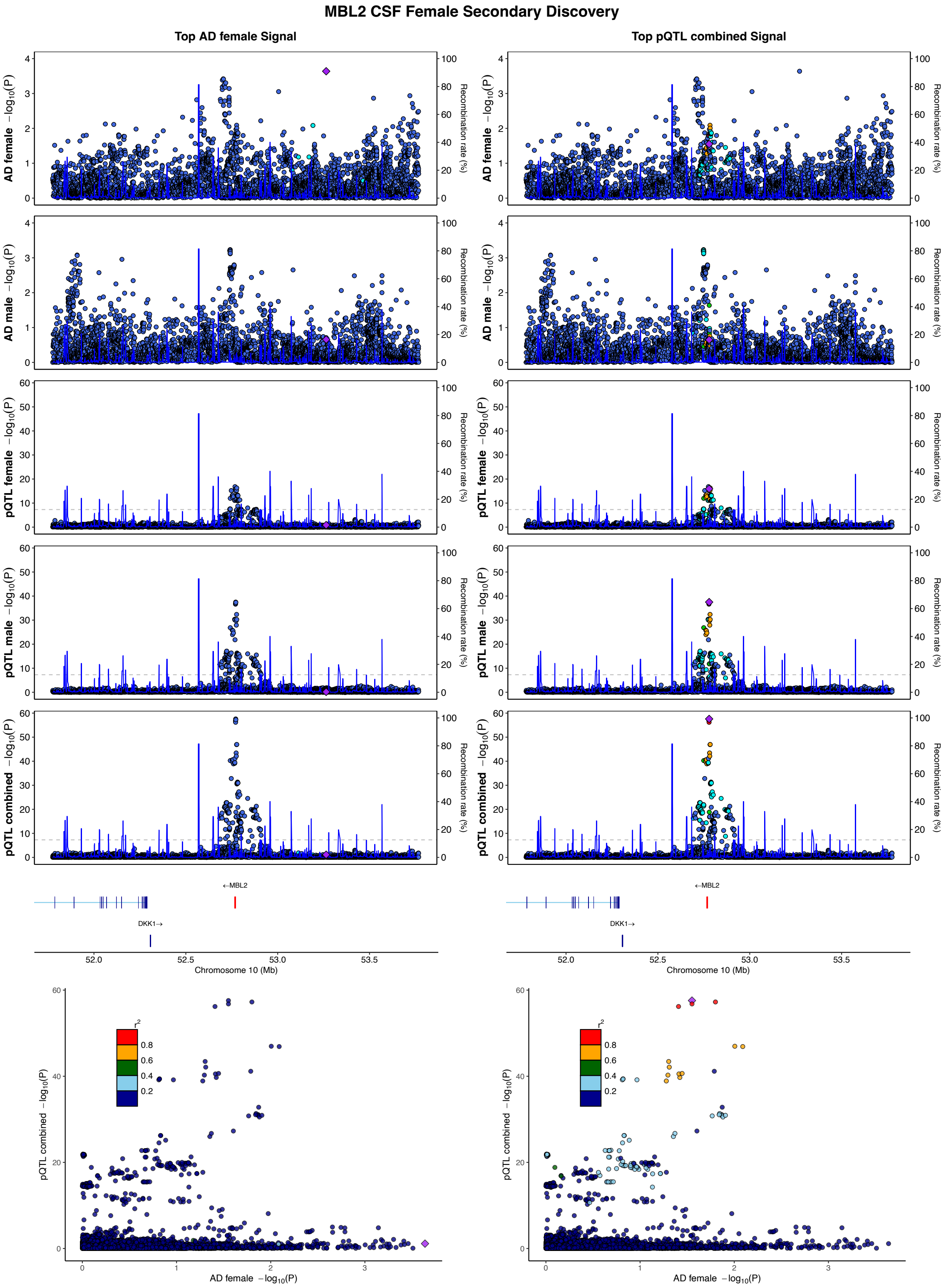

MBL2 CSF Female Secondary Discovery
Top AD female Signal
Top pQTL combined Signal
AD female −log10(P)
AD male −log10(P)
pQTL female −log10(P)
pQTL male −log10(P)
pQTL combined −log10(P)
Recombination rate (%)
←MBL2
DKK1→
Chromosome 10 (Mb)
r2
AD female −log10(P)

## S14.18

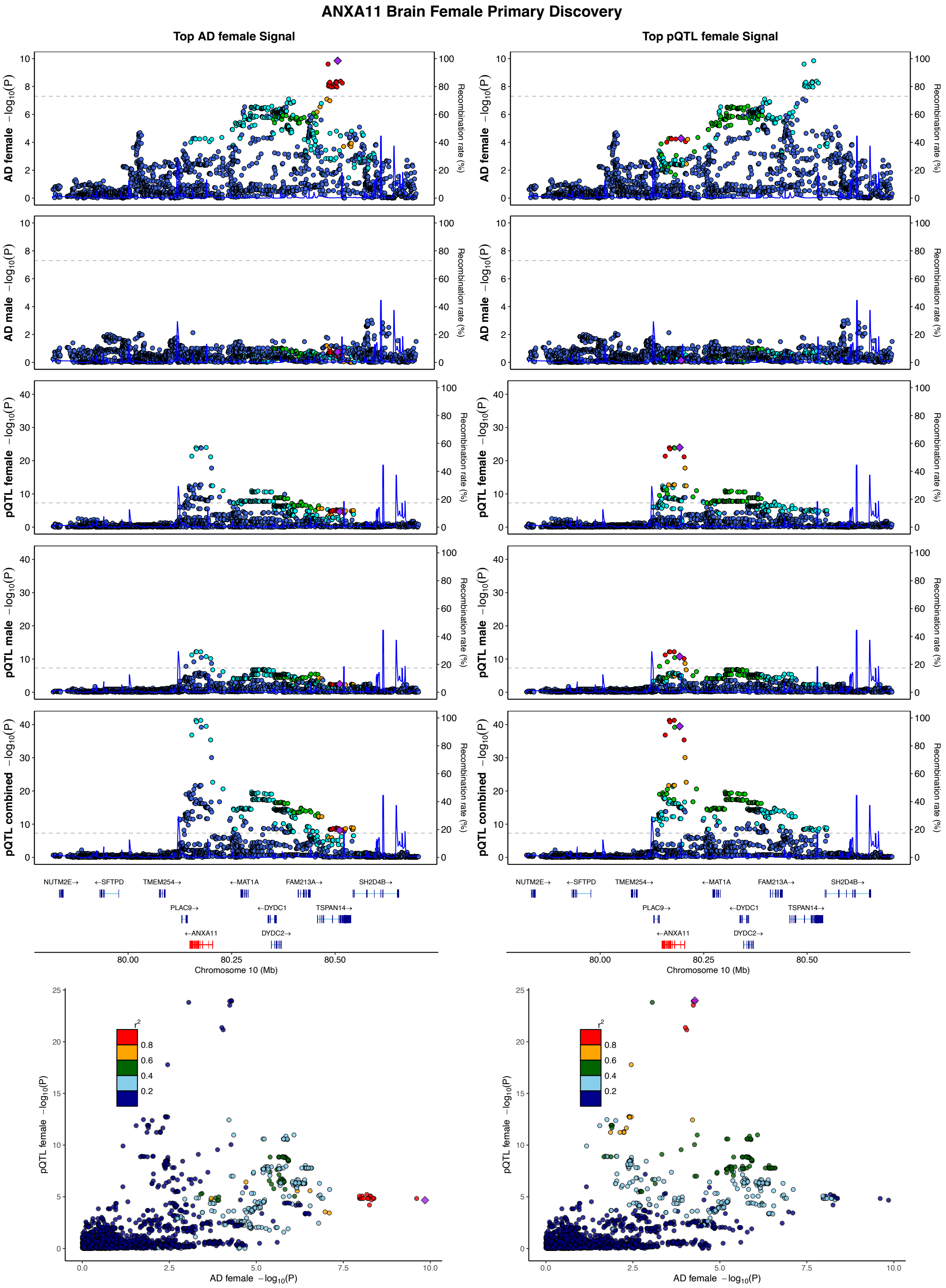
ANXA11 Brain Female Primary Discovery
Top AD female Signal
Top pQTL female Signal
AD female −log10(P)
AD male −log10(P)
pQTL female −log10(P)
pQTL male −log10(P)
pQTL combined −log10(P)
Recombination rate (%)
NUTM2E→
←SFTPD
TMEM254→
←MAT1A
FAM213A→
SH2D4B→
PLAC9→
←DYDC1
TSPAN14→
←ANXA11
DYDC2→
Chromosome 10 (Mb)
AD female −log10(P)
r2
0.8
0.6
0.4
0.2

## S14.19

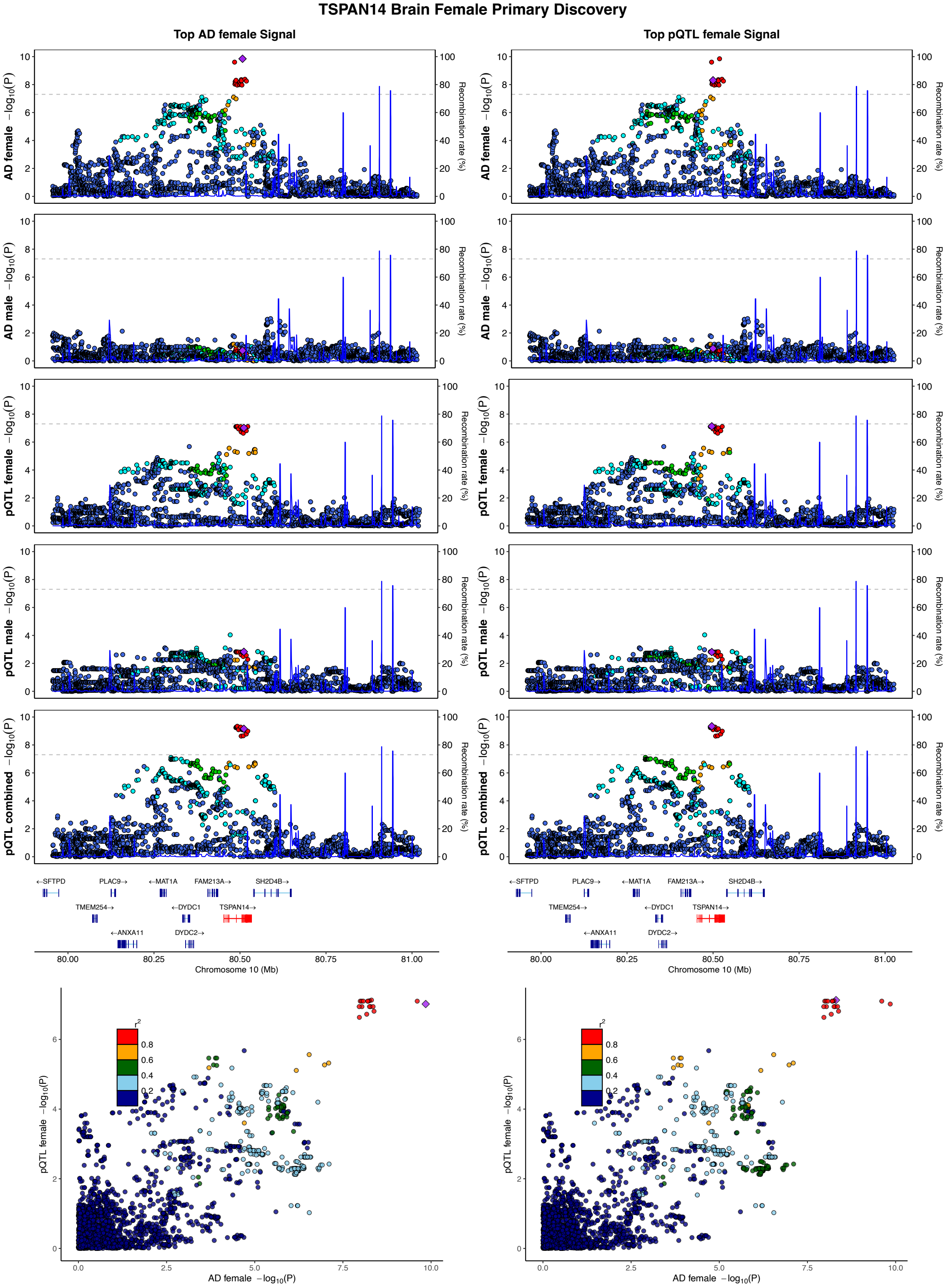
TSPAN14 Brain Female Primary Discovery
Top AD female Signal
Top pQTL female Signal
AD female −log10(P)
AD male −log10(P)
pQTL female −log10(P)
pQTL male −log10(P)
pQTL combined −log10(P)
Recombination rate (%)
←SFTPD
PLAC9→
←MAT1A
FAM213A→
SH2D4B→
TMEM254→
←DYDC1
TSPAN14→
←ANXA11
DYDC2→
80.00
80.25
80.50
80.75
81.00
Chromosome 10 (Mb)
r2
0.8
0.6
0.4
0.2
pQTL female −log10(P)
AD female −log10(P)

## S14.20

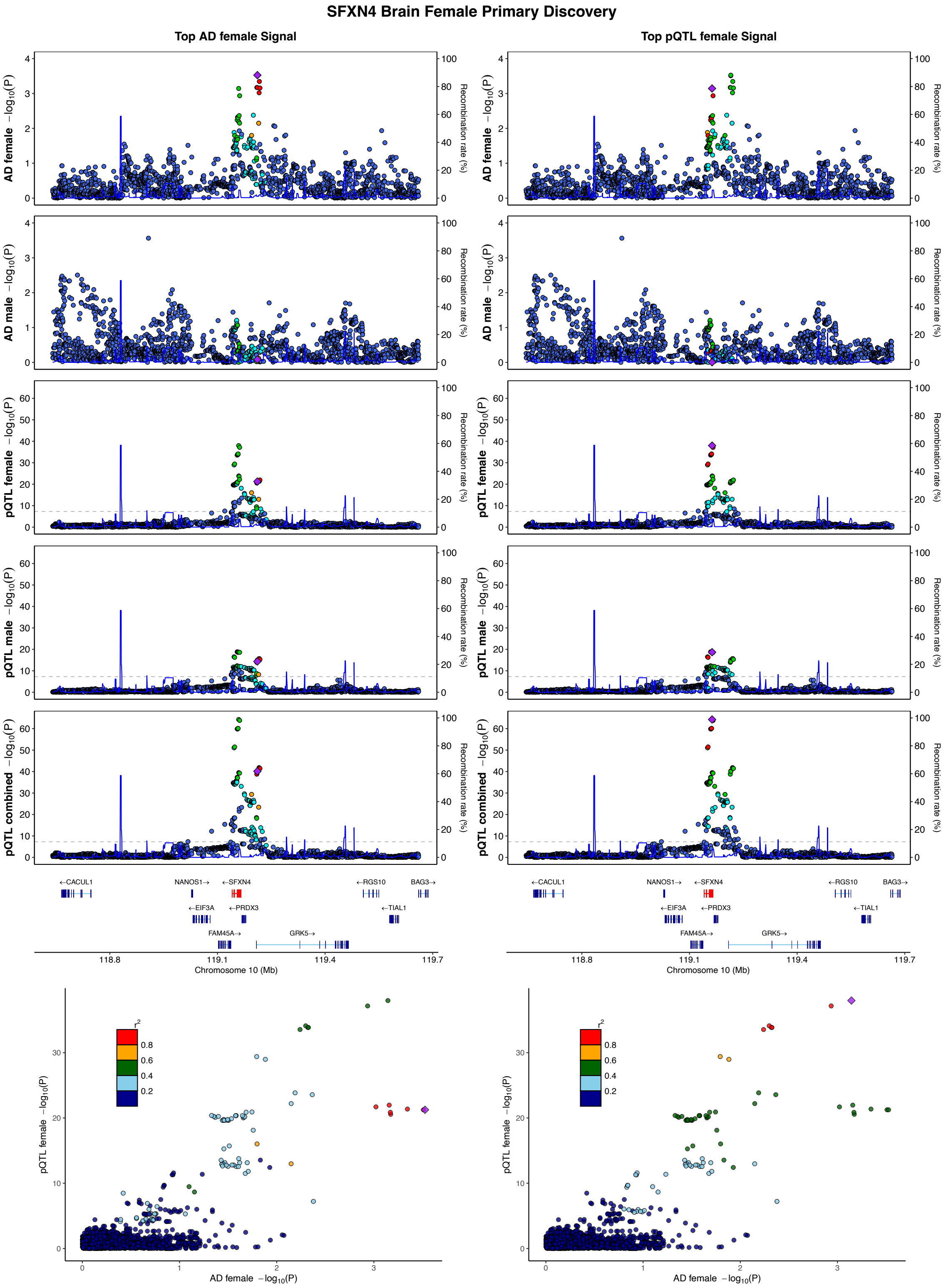

SFXN4 Brain Female Primary Discovery
Top AD female Signal
Top pQTL female Signal
AD female −log10(P)
AD male −log10(P)
pQTL female −log10(P)
pQTL male −log10(P)
pQTL combined −log10(P)
Recombination rate (%)
←CACUL1
NANOS1→
←SFXN4
←RGS10
BAG3→
←EIF3A
←PRDX3
←TIAL1
FAM45A→
GRK5→
118.8
119.1
119.4
119.7
Chromosome 10 (Mb)
r2
0.8
0.6
0.4
0.2
pQTL female −log10(P)
AD female −log10(P)

## S14.21

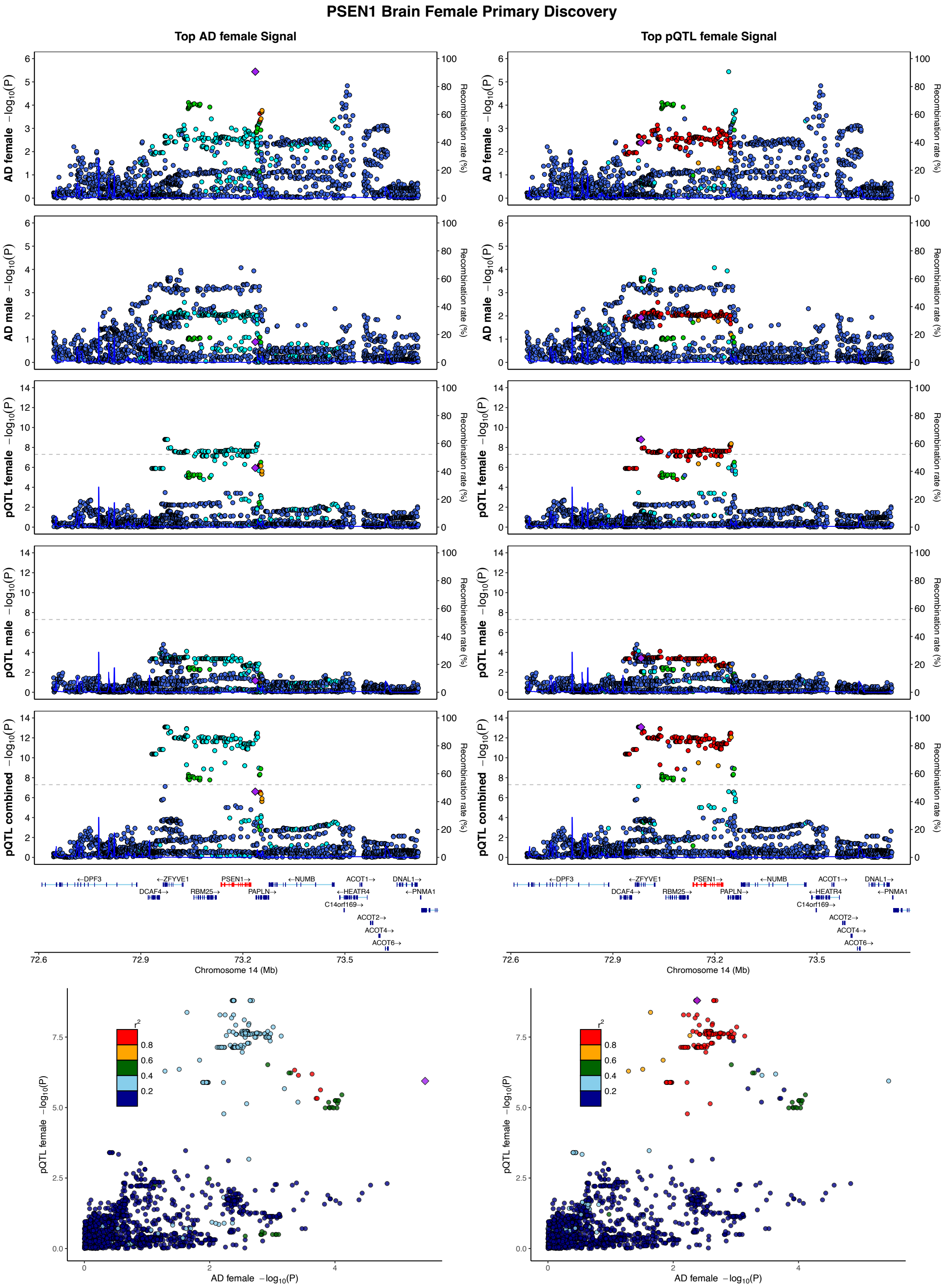

PSEN1 Brain Female Primary Discovery
Top AD female Signal
Top pQTL female Signal
AD female −log10(P)
AD male −log10(P)
pQTL female −log10(P)
pQTL male −log10(P)
pQTL combined −log10(P)
Recombination rate (%)
DPF3
ZFYVE1
PSEN1
NUMB
ACOT1
DNAL1
DCAF4
RBM25
PAPLN
HEATR4
PNMA1
C14orf169
ACOT2
ACOT4
ACOT6
Chromosome 14 (Mb)
r2
pQTL female −log10(P)
AD female −log10(P)

## S14.22

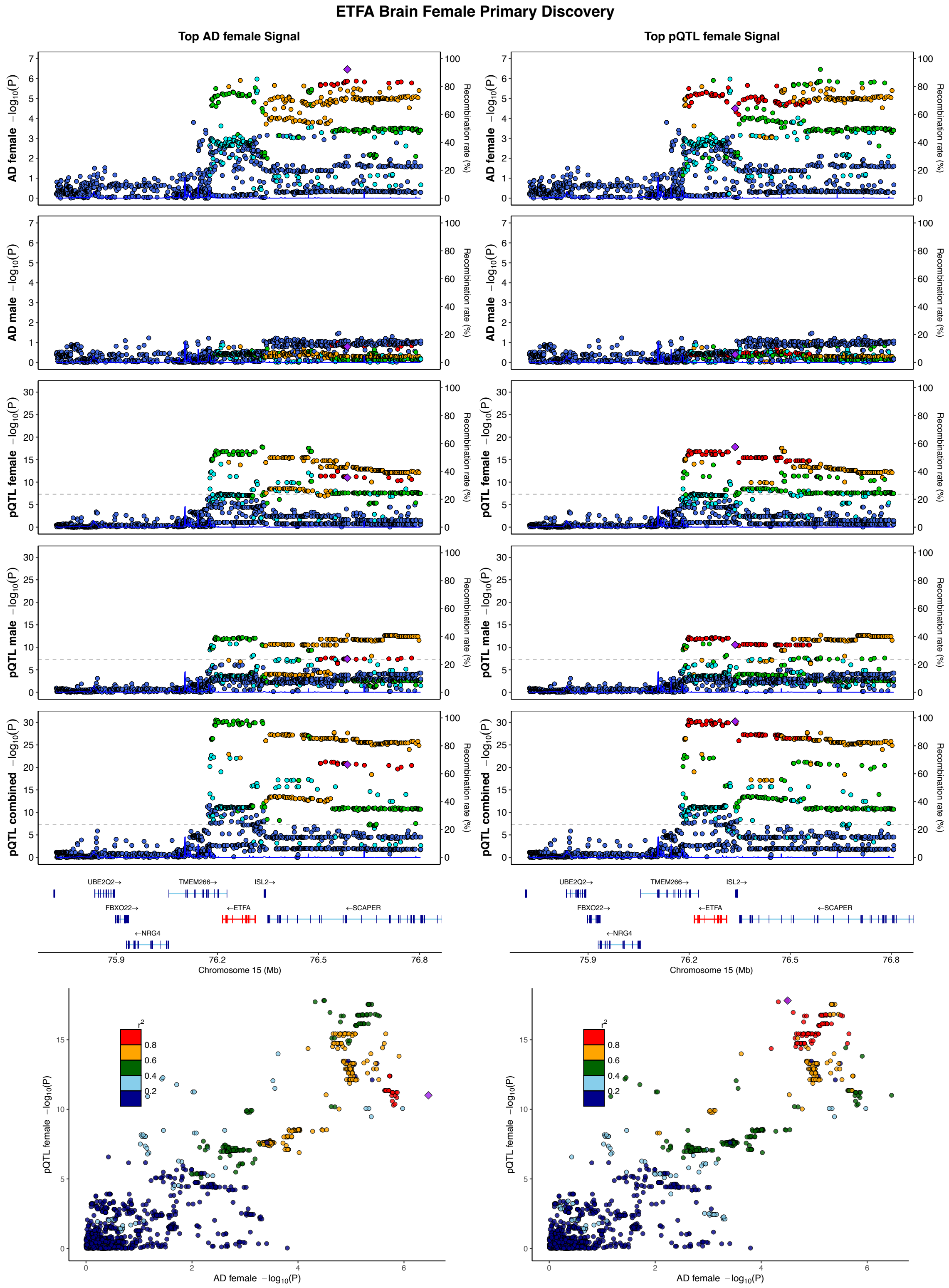

ETFA Brain Female Primary Discovery
Top AD female Signal
Top pQTL female Signal
AD female −log10(P)
AD male −log10(P)
pQTL female −log10(P)
pQTL male −log10(P)
pQTL combined −log10(P)
Recombination rate (%)
UBE2Q2→
TMEM266→
ISL2→
FBXO22→
←ETFA
←SCAPER
←NRG4
75.9
76.2
76.5
76.8
Chromosome 15 (Mb)
r2
0.8
0.6
0.4
0.2
pQTL female −log10(P)
AD female −log10(P)

S14.23

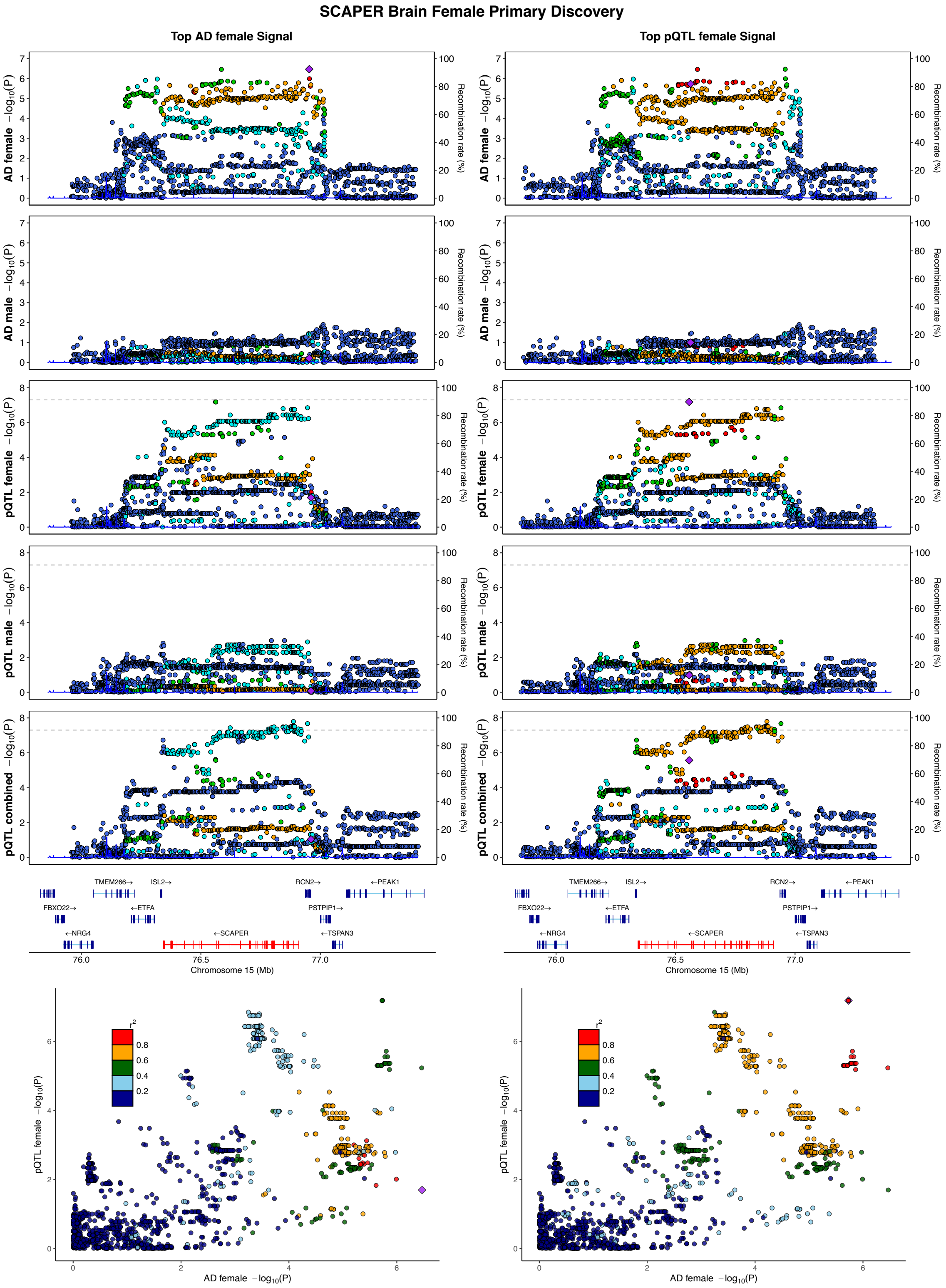
SCAPER Brain Female Primary Discovery
Top AD female Signal
Top pQTL female Signal
AD female −log10(P)
AD male −log10(P)
pQTL female −log10(P)
pQTL male −log10(P)
pQTL combined −log10(P)
Recombination rate (%)
TMEM266→
ISL2→
RCN2→
←PEAK1
FBXO22→
←ETFA
PSTPIP1→
←NRG4
←SCAPER
←TSPAN3
Chromosome 15 (Mb)
r2
AD female −log10(P)

## S14.24

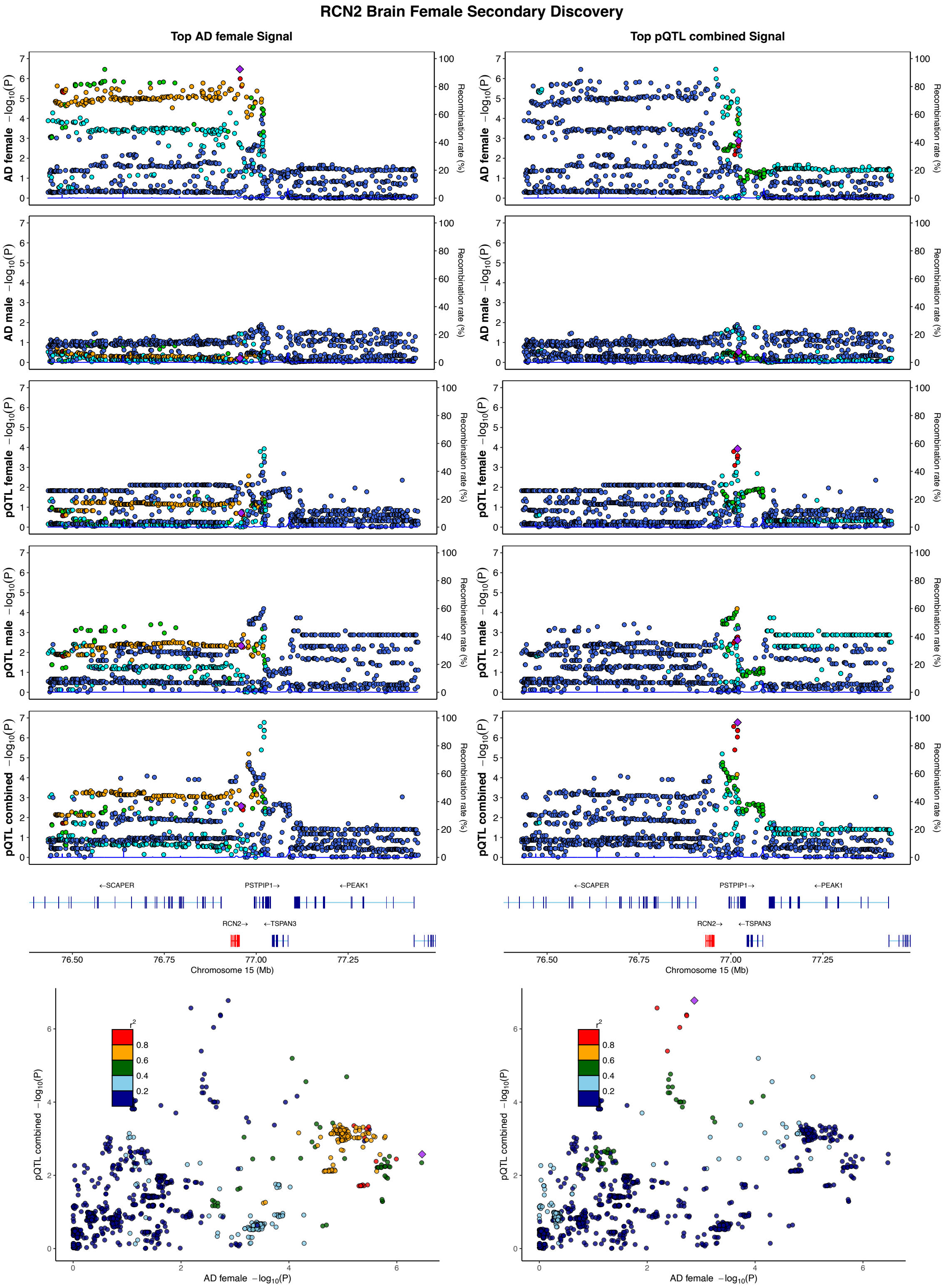
RCN2 Brain Female Secondary Discovery
Top AD female Signal
Top pQTL combined Signal
AD female −log10(P)
AD male −log10(P)
pQTL female −log10(P)
pQTL male −log10(P)
pQTL combined −log10(P)
Recombination rate (%)
←SCAPER
PSTPIP1→
←PEAK1
RCN2→
←TSPAN3
76.50
76.75
77.00
77.25
Chromosome 15 (Mb)
r2
0.8
0.6
0.4
0.2
AD female −log10(P)
pQTL combined −log10(P)

## S14.25

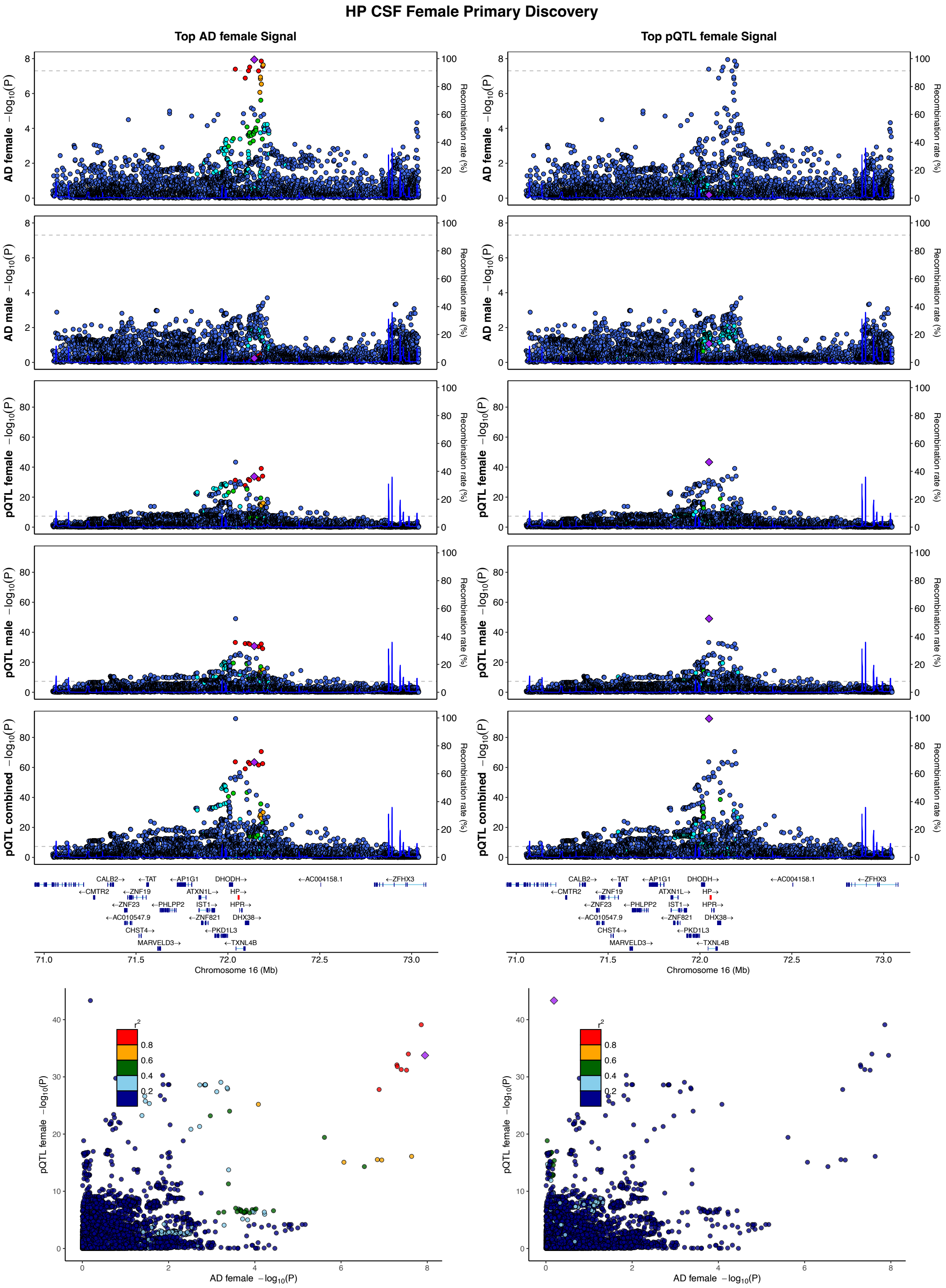

HP CSF Female Primary Discovery
Top AD female Signal
Top pQTL female Signal
AD female −log10(P)
AD male −log10(P)
pQTL female −log10(P)
pQTL male −log10(P)
pQTL combined −log10(P)
Recombination rate (%)
CALB2→
←TAT
←AP1G1
DHODH→
←AC004158.1
←ZFHX3
←CMTR2
←ZNF19
ATXN1L→
HP→
←ZNF23
←PHLPP2
IST1→
HPR→
←AC010547.9
←ZNF821
DHX38→
CHST4→
←PKD1L3
MARVELD3→
←TXNL4B
Chromosome 16 (Mb)
pQTL female −log10(P)
AD female −log10(P)
r2

## S14.26

##### SLC7A5 Brain Female Secondary Discovery

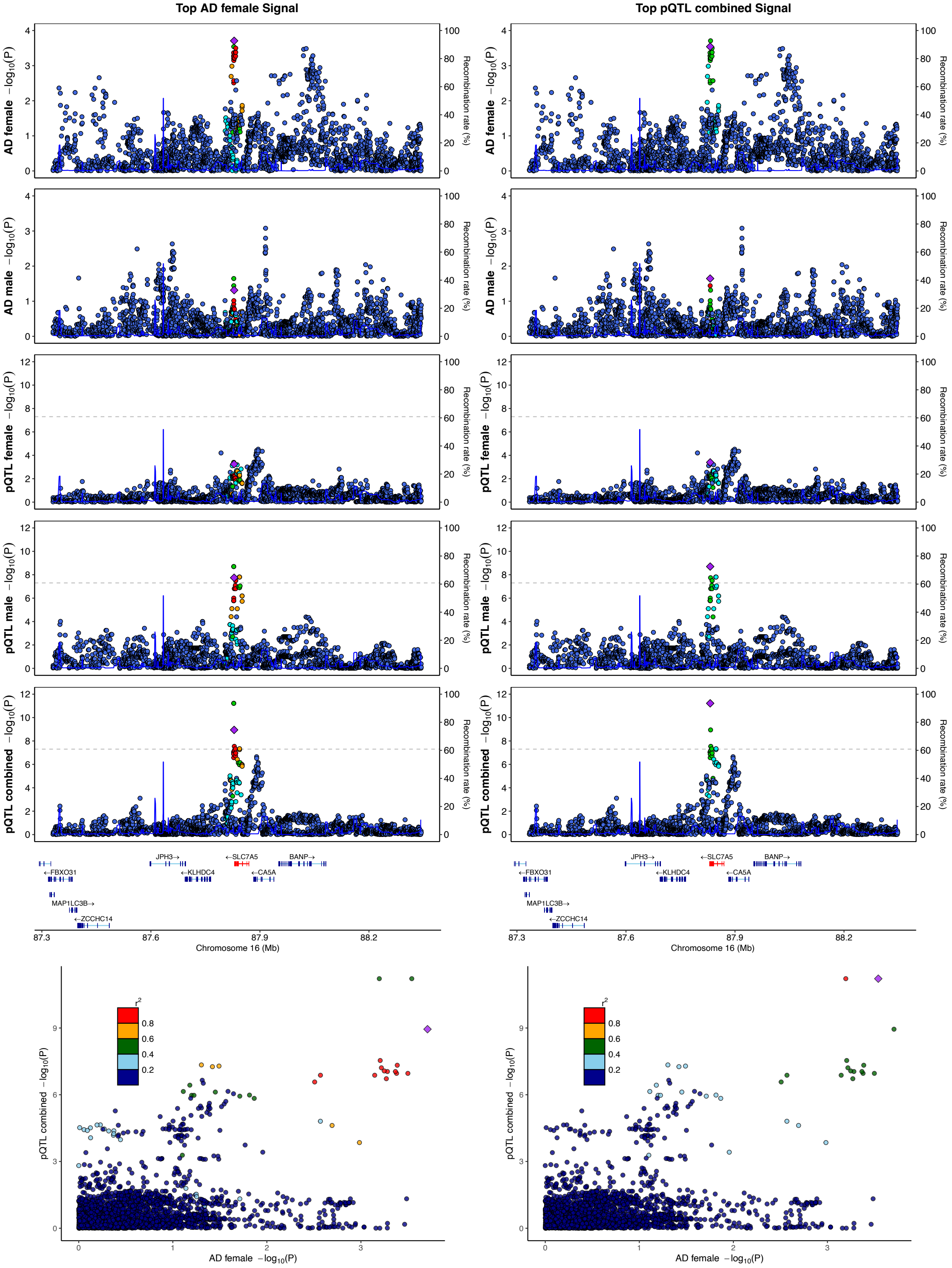

## S14.27

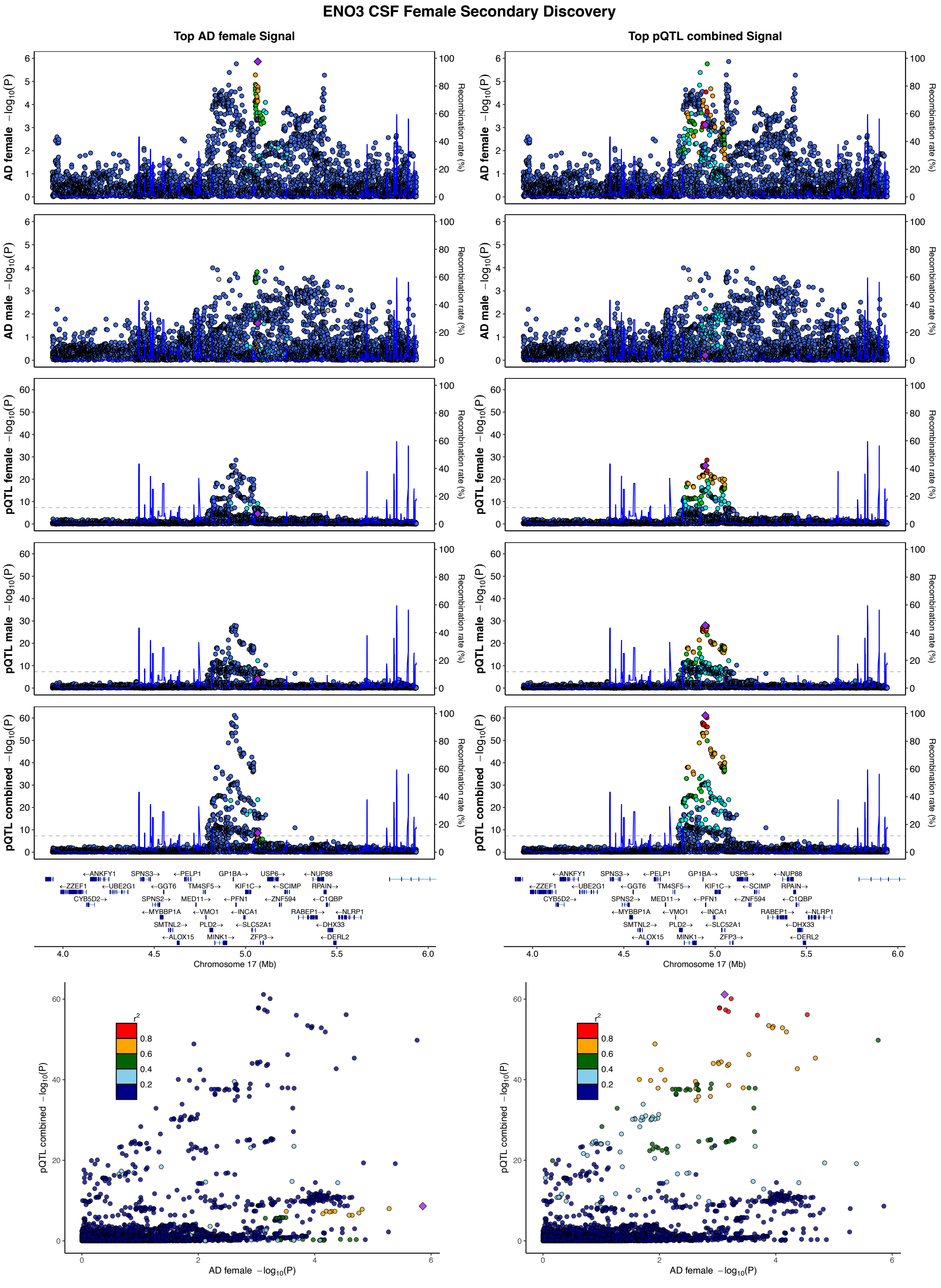
ENO3 CSF Female Secondary Discovery
Top AD female Signal
Top pQTL combined Signal
AD female −log10(P)
AD male −log10(P)
pQTL female −log10(P)
pQTL male −log10(P)
pQTL combined −log10(P)
Recombination rate (%)
Chromosome 17 (Mb)
AD female −log10(P)
r2

## S14.28

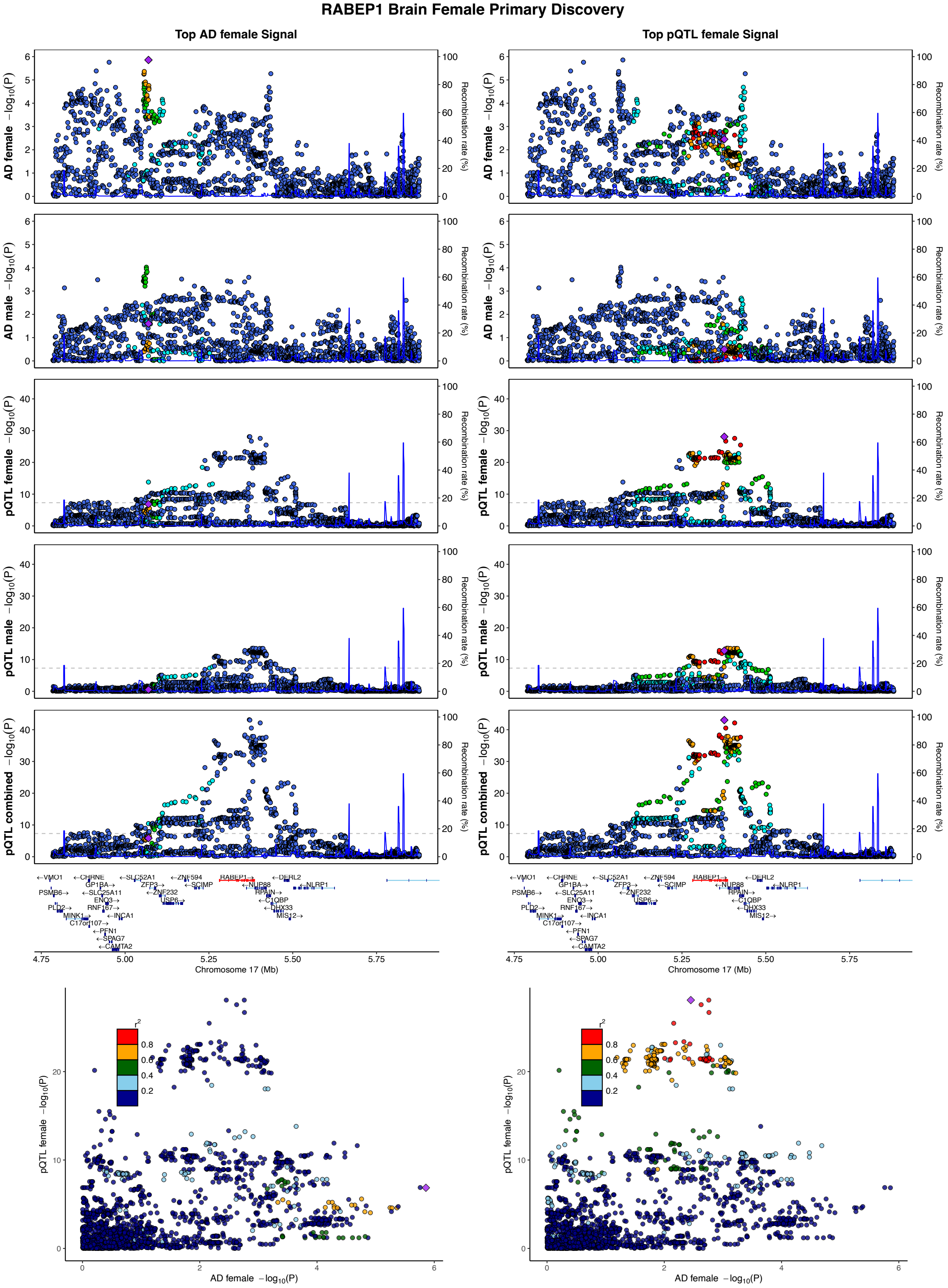

RABEP1 Brain Female Primary Discovery
Top AD female Signal
Top pQTL female Signal
AD female −log10(P)
AD male −log10(P)
pQTL female −log10(P)
pQTL male −log10(P)
pQTL combined −log10(P)
Recombination rate (%)
Chromosome 17 (Mb)
AD female −log10(P)
pQTL female −log10(P)

## S14.29

##### TOM1L2 Brain Female Primary Discovery

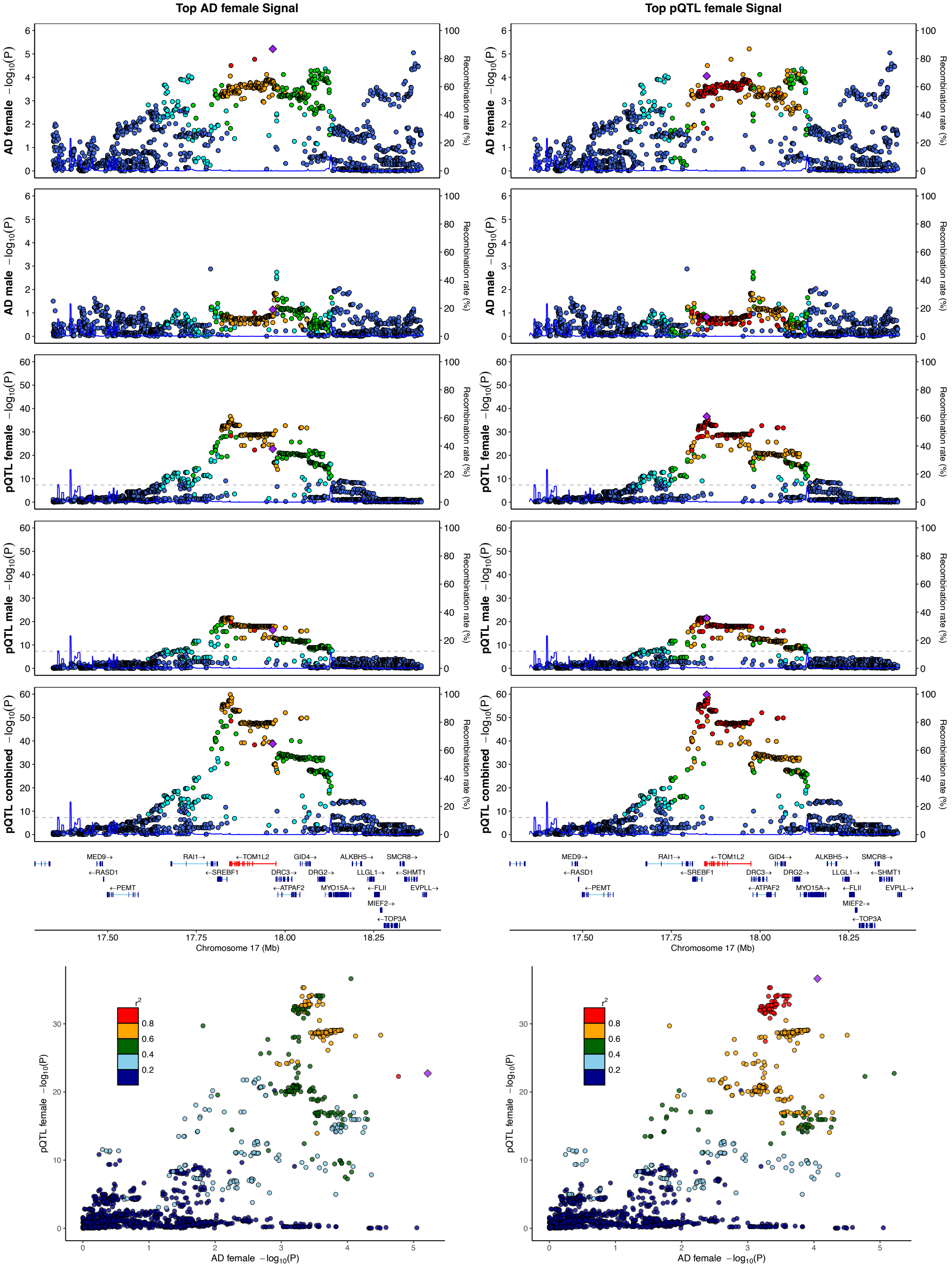

## S14.30

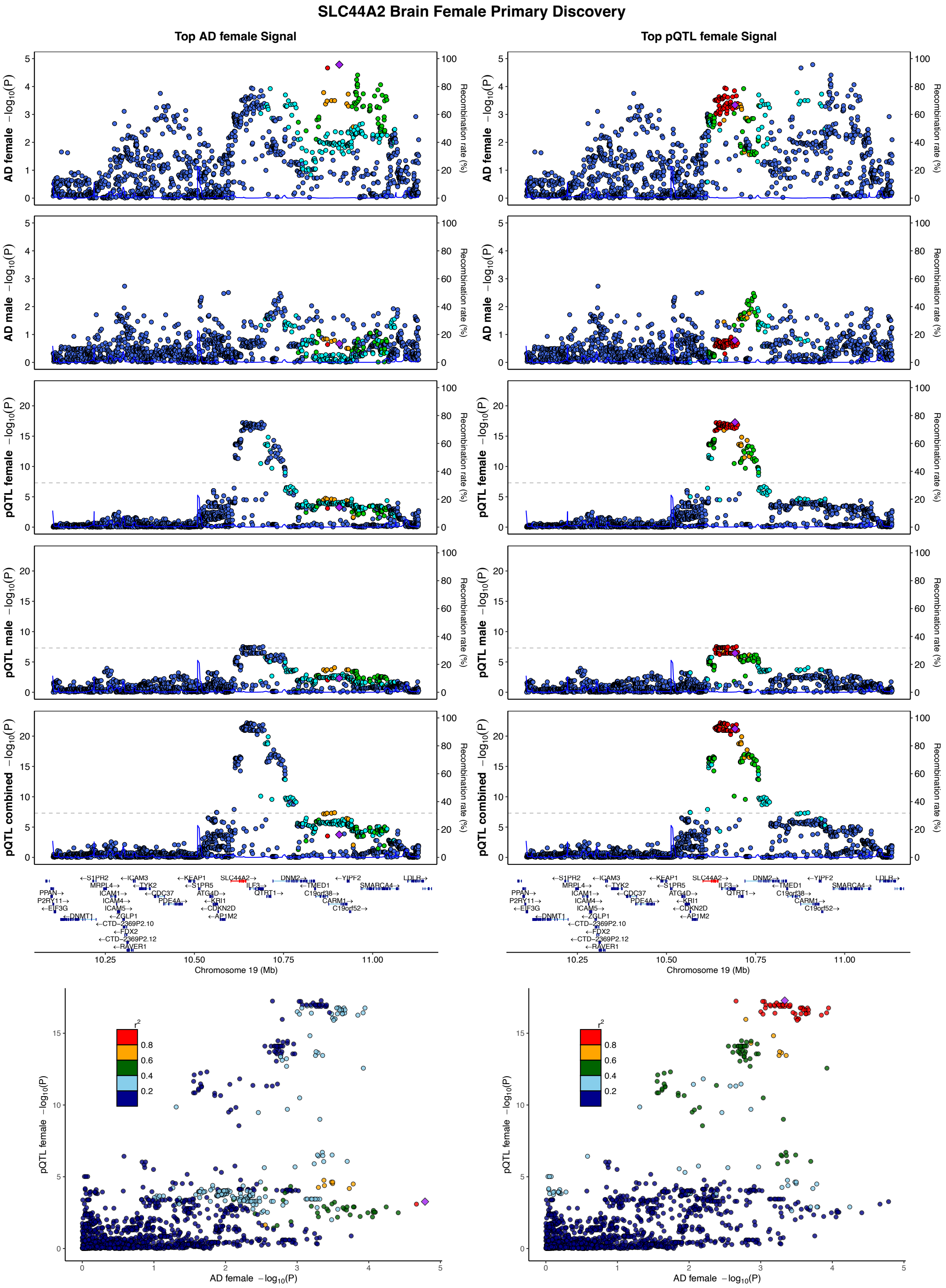
SLC44A2 Brain Female Primary Discovery
Top AD female Signal
Top pQTL female Signal
AD female −log10(P)
AD male −log10(P)
pQTL female −log10(P)
pQTL male −log10(P)
pQTL combined −log10(P)
Recombination rate (%)
Chromosome 19 (Mb)
AD female −log10(P)
r2

## S14.31

##### CARM1 Brain Female Primary Discovery

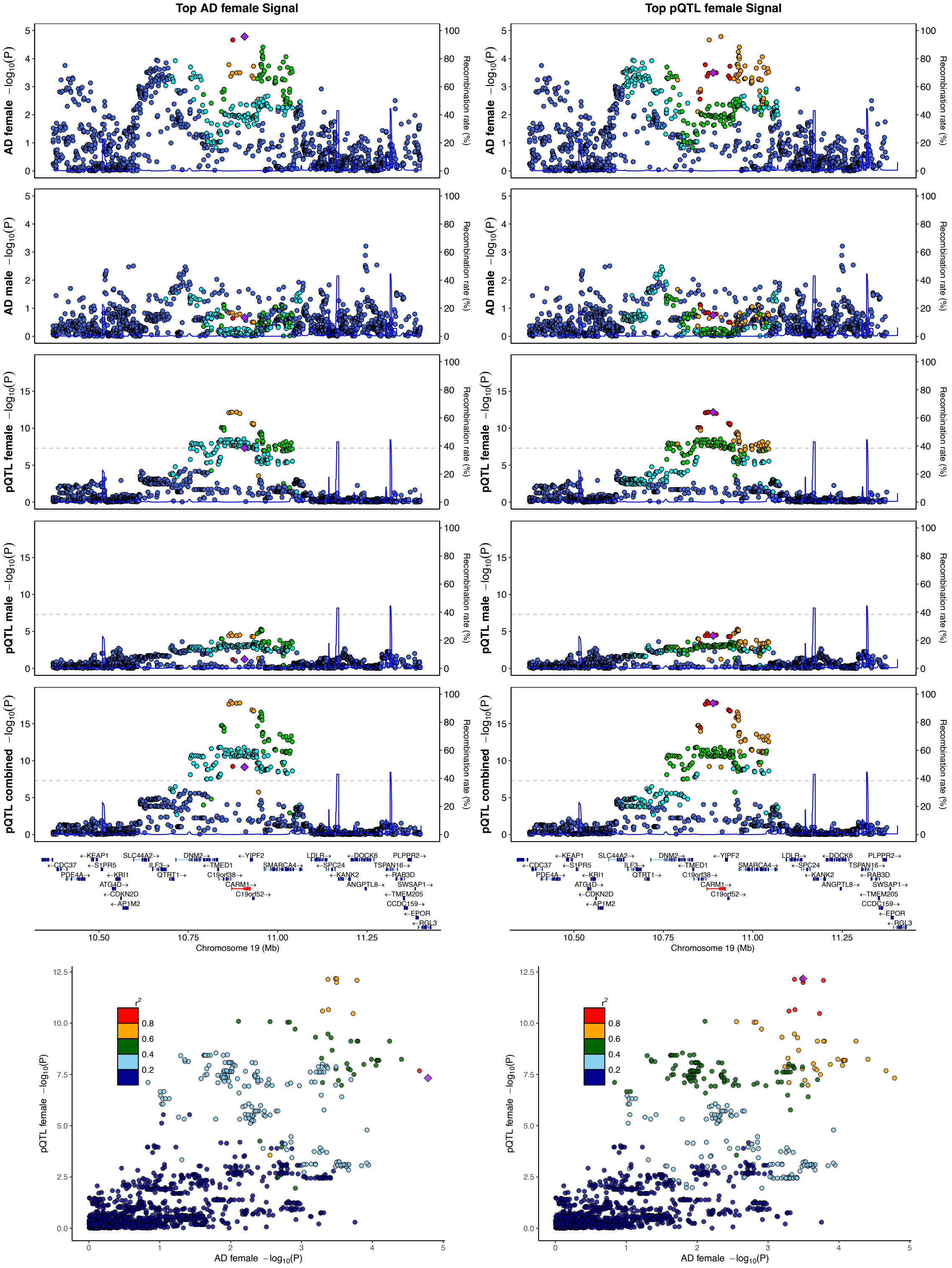

## S14.32

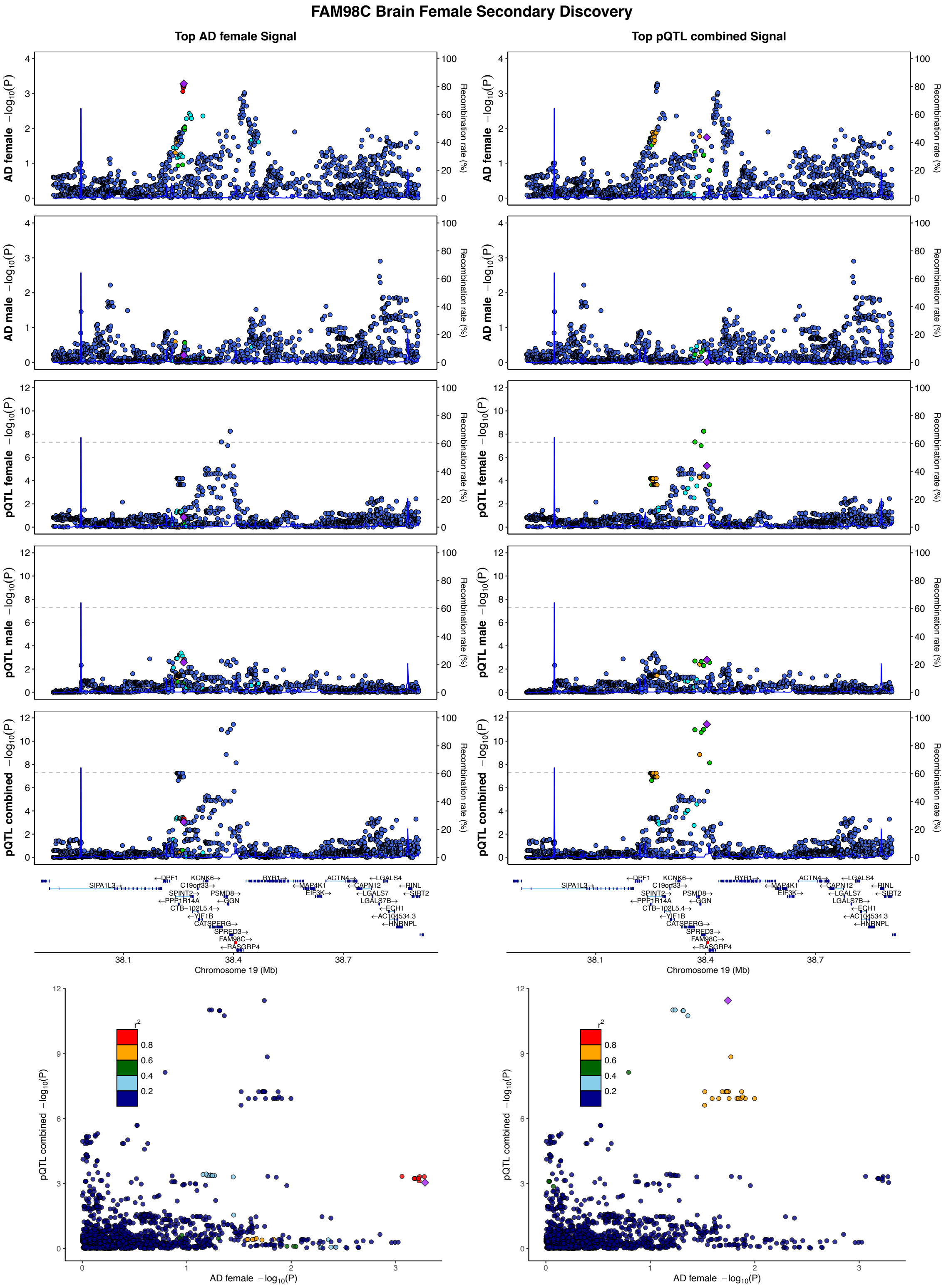
FAM98C Brain Female Secondary Discovery
Top AD female Signal
Top pQTL combined Signal
AD female −log10(P)
AD male −log10(P)
pQTL female −log10(P)
pQTL male −log10(P)
pQTL combined −log10(P)
Recombination rate (%)
Chromosome 19 (Mb)
AD female −log10(P)
pQTL combined −log10(P)
r2

## S14.33

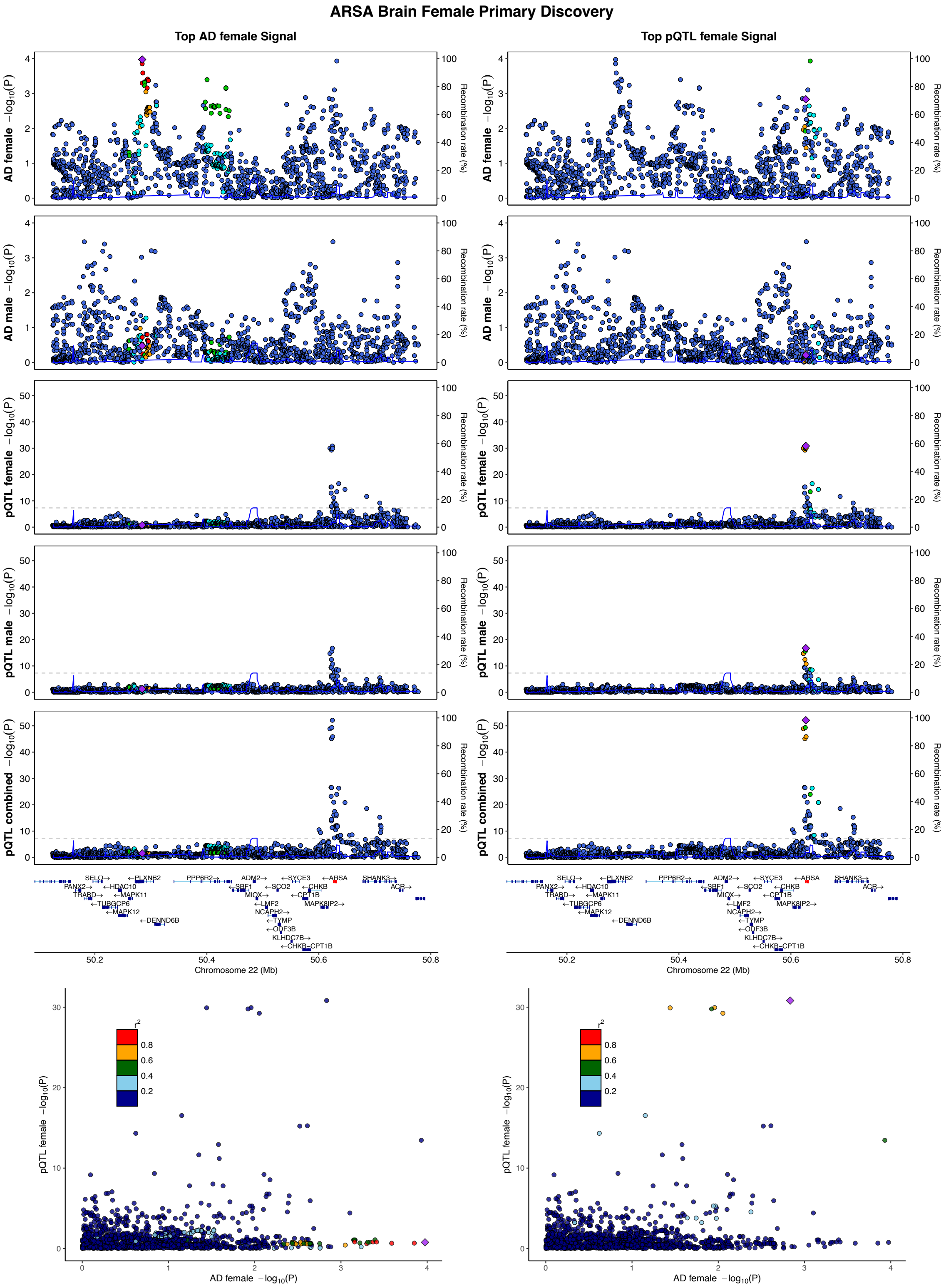
ARSA Brain Female Primary Discovery
Top AD female Signal
Top pQTL female Signal
AD female −log10(P)
AD male −log10(P)
pQTL female −log10(P)
pQTL male −log10(P)
pQTL combined −log10(P)
Recombination rate (%)
Chromosome 22 (Mb)
AD female −log10(P)
r2

## S14.34

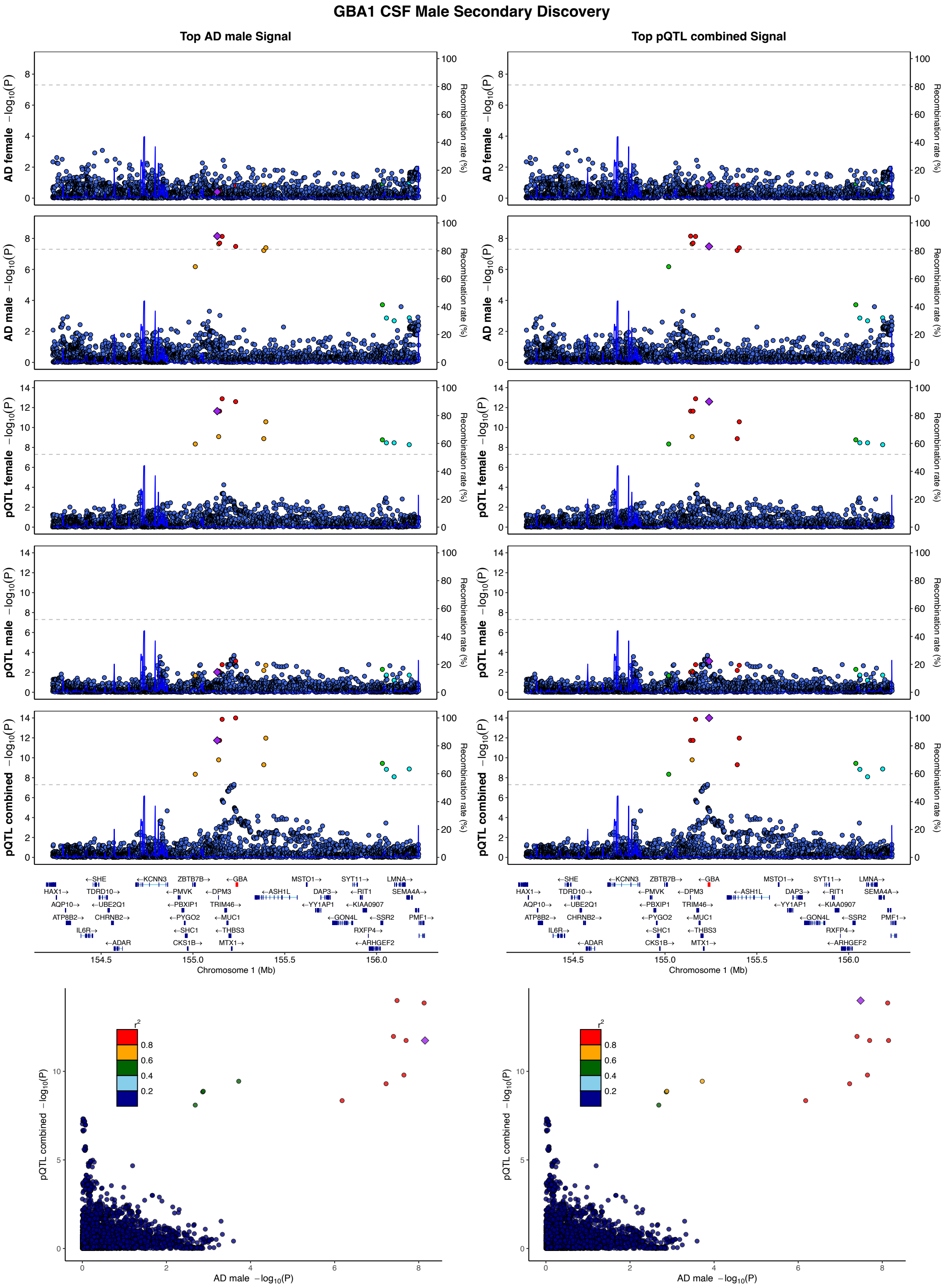

GBA1 CSF Male Secondary Discovery
Top AD male Signal
Top pQTL combined Signal
AD female −log10(P)
AD male −log10(P)
pQTL female −log10(P)
pQTL male −log10(P)
pQTL combined −log10(P)
Recombination rate (%)
Chromosome 1 (Mb)
154.5
155.0
155.5
156.0
←SHE
←KCNN3
ZBTB7B→
←GBA
MSTO1→
SYT11→
LMNA→
HAX1→
TDRD10→
←PMVK
←DPM3
←ASH1L
DAP3→
←RIT1
SEMA4A→
AQP10→
←UBE2Q1
←PBXIP1
TRIM46→
←YY1AP1
←KIAA0907
ATP8B2→
CHRNB2→
←PYGO2
←MUC1
←GON4L
←SSR2
PMF1→
IL6R→
←SHC1
←THBS3
RXFP4→
←ADAR
CKS1B→
MTX1→
←ARHGEF2
r2
0.8
0.6
0.4
0.2
pQTL combined −log10(P)
AD male −log10(P)

## S14.35

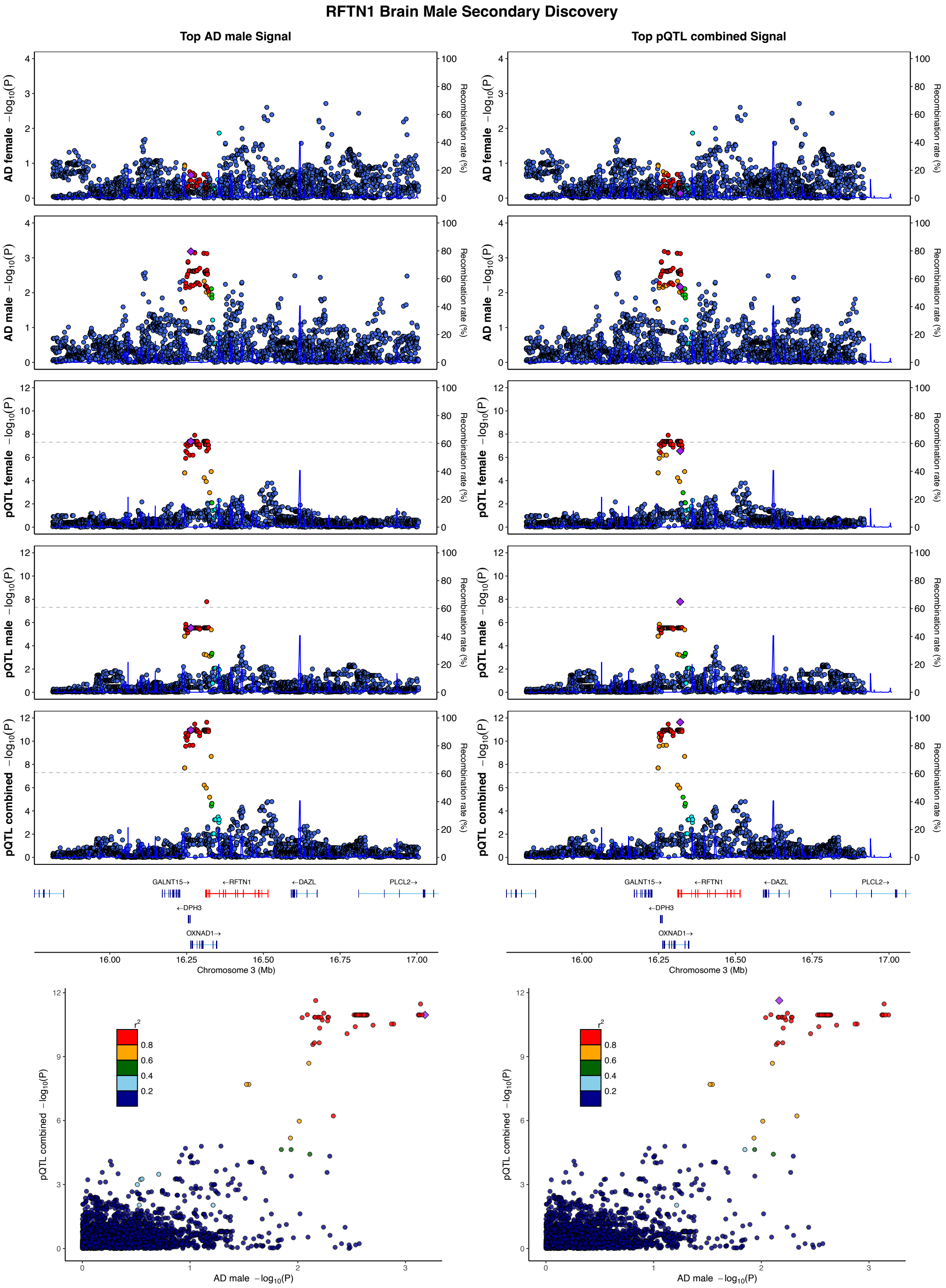
RFTN1 Brain Male Secondary Discovery
Top AD male Signal
Top pQTL combined Signal
AD female −log10(P)
AD male −log10(P)
pQTL female −log10(P)
pQTL male −log10(P)
pQTL combined −log10(P)
Recombination rate (%)
GALNT15→
←RFTN1
←DAZL
PLCL2→
←DPH3
OXNAD1→
Chromosome 3 (Mb)
AD male −log10(P)
r2
0.8
0.6
0.4
0.2

## S14.36

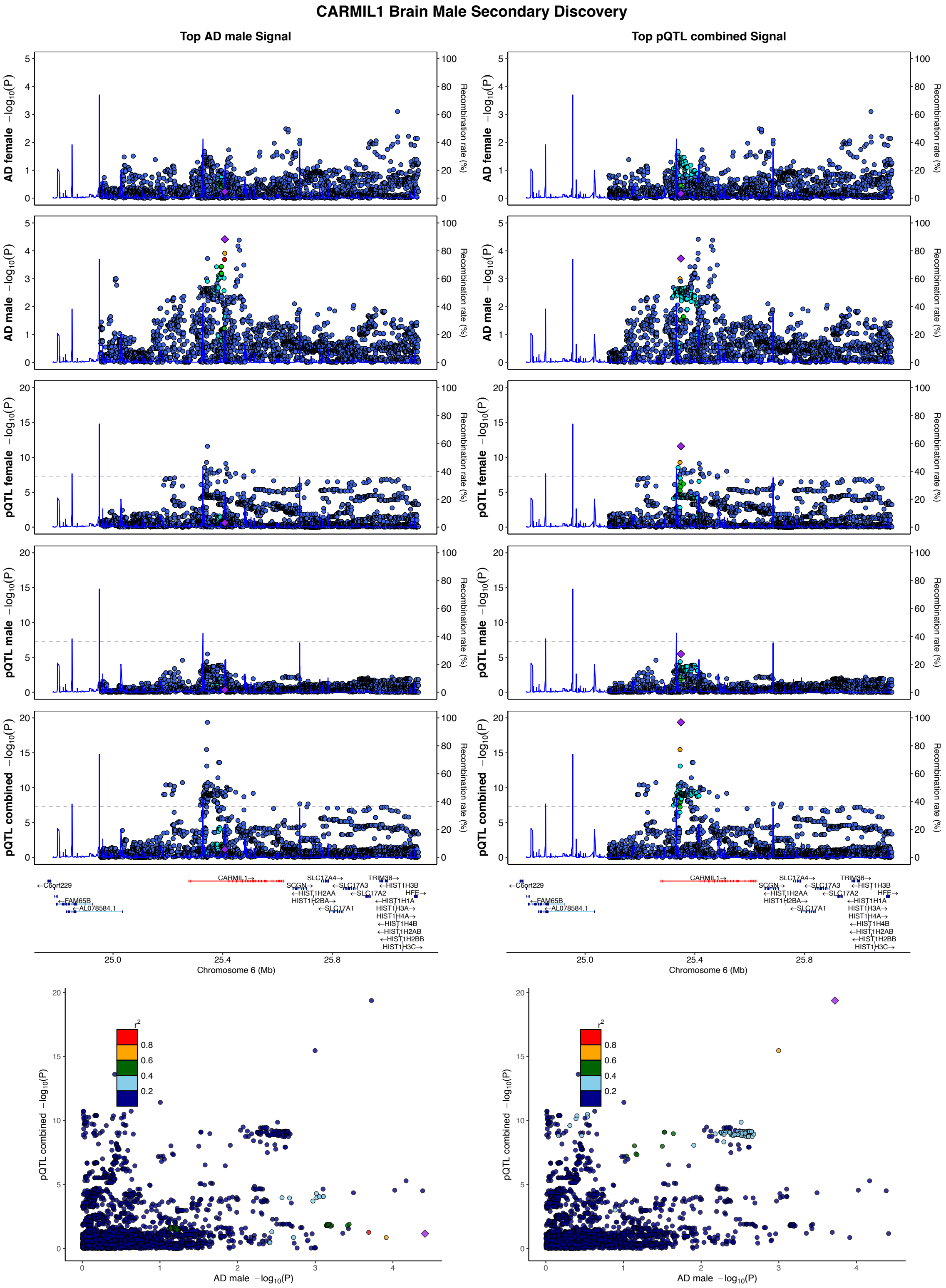

CARMIL1 Brain Male Secondary Discovery
Top AD male Signal
Top pQTL combined Signal
AD female −log10(P)
AD male −log10(P)
pQTL female −log10(P)
pQTL male −log10(P)
pQTL combined −log10(P)
Recombination rate (%)
Chromosome 6 (Mb)
AD male −log10(P)
r2

## S14.37

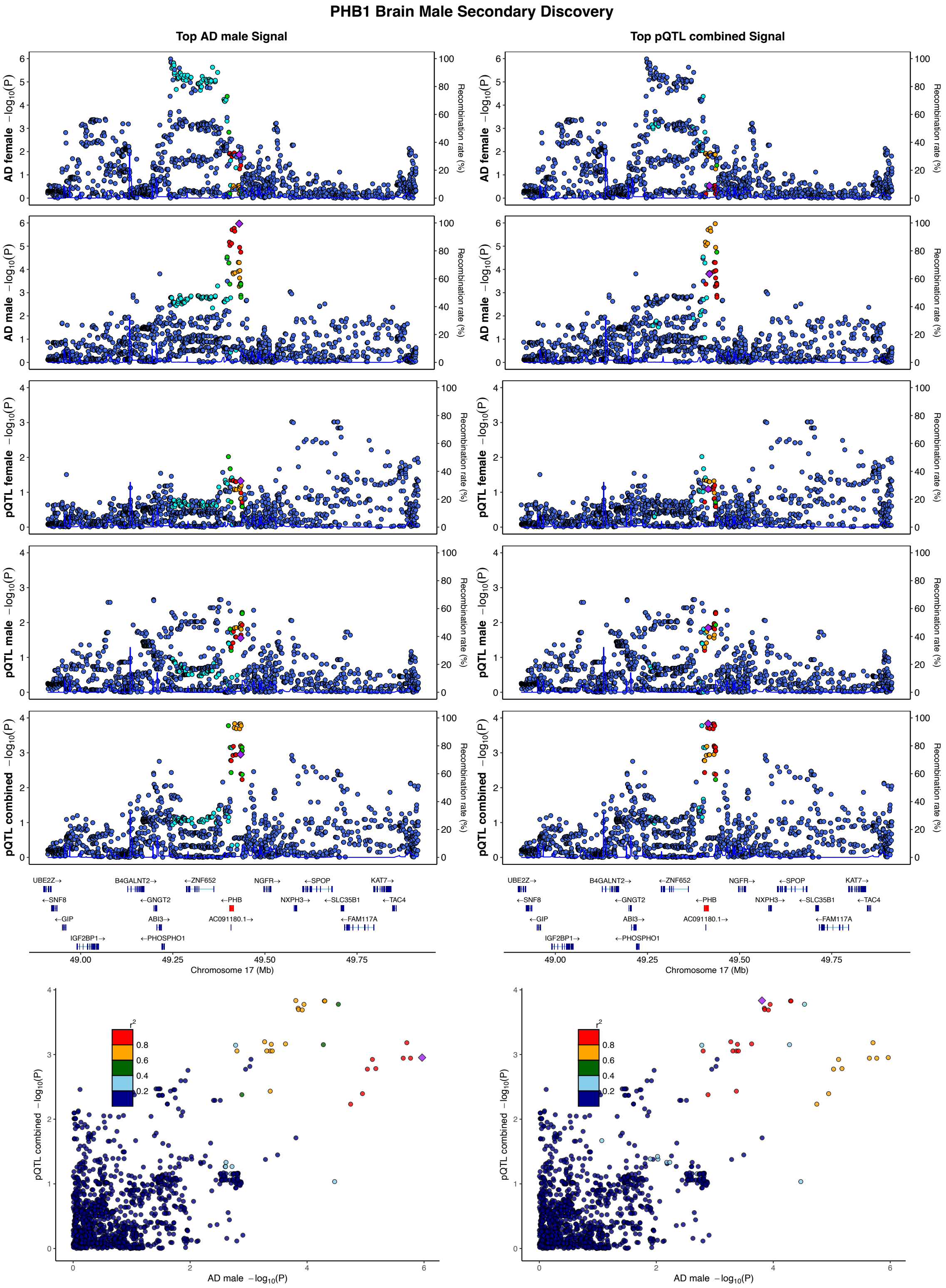
PHB1 Brain Male Secondary Discovery
Top AD male Signal
Top pQTL combined Signal
AD female −log10(P)
AD male −log10(P)
pQTL female −log10(P)
pQTL male −log10(P)
pQTL combined −log10(P)
Recombination rate (%)
UBE2Z→
←SNF8
←GIP
IGF2BP1→
B4GALNT2→
←GNGT2
ABI3→
←PHOSPHO1
←ZNF652
←PHB
AC091180.1→
NGFR→
NXPH3→
←SPOP
←SLC35B1
←FAM117A
KAT7→
←TAC4
49.00
49.25
49.50
49.75
Chromosome 17 (Mb)
r2
0.8
0.6
0.4
0.2
pQTL combined −log10(P)
AD male −log10(P)

**Supplementary Figure 14. Locus zoom and compare plots of GWAS and pQTL signals matching sex-specific Alzheimer's disease PWAS discoveries. (1-33)** Female-biased and **(34-37)** Male-biased gene discoveries ordered based on chromosome and base-pair position. The first column (left side) tags the top AD variant for the respective sex-specific discovery, while the second column (right side) tags the top variant from the female/male or combined pQTL. The locus zoom plots are arranged by the order of AD female, AD male, pQTL female, pQTL male, pQTL combined. In the last row, locus compare plots present AD GWAS and pQTL data corresponding to the sex-specificity and discovery type (secondary/primary) of the respective PWAS finding. Dot color coding represents LD with tagging variant (purple diamond): red – $R^2$: 0.8-1.0, orange – $R^2$:0.6-0.8, green – $R^2$:0.4-0.6, cyan – $R^2$:0.2-0.4, blue – $R^2$:0.0-0.2.

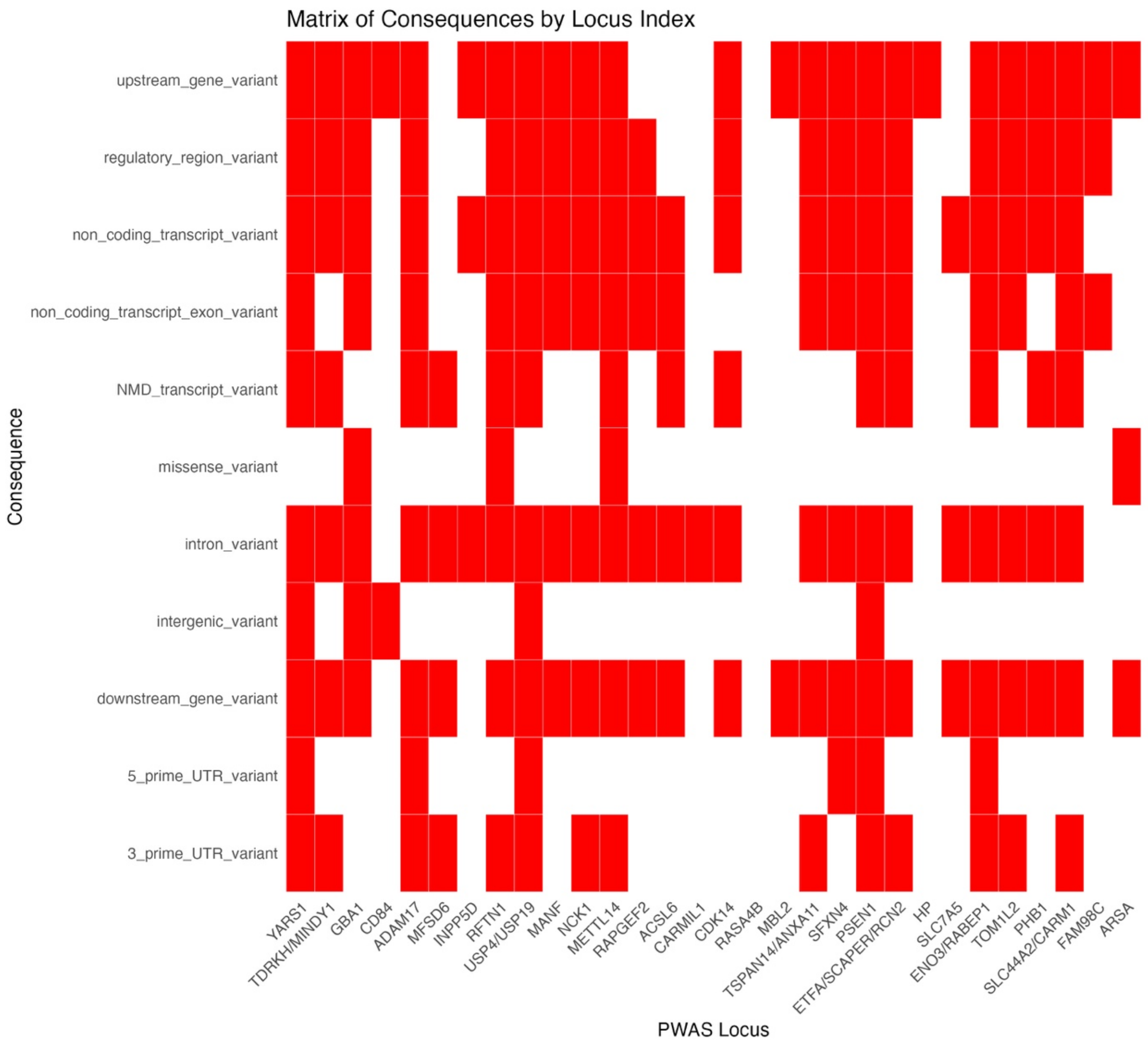

**Supplementary Figure 15. Matrix of variant consequence predictions across prioritized PWAS loci.** Matrix summarizing the variant effect predictor (VEP) consequences (y-axis) for lead pQTLs, and any variants in LD ($R^2$>0.8) , from sex-specific PWAS discoveries (x-axis). Red cells indicate the annotations.

A

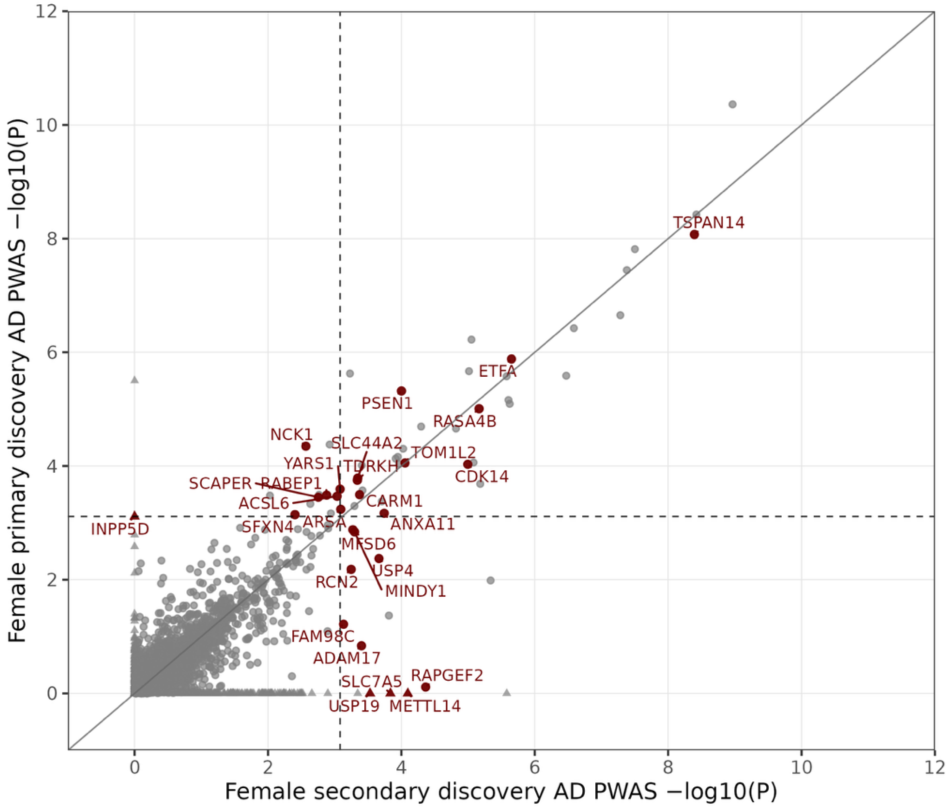
Female primary discovery AD PWAS −log10(P)
Female secondary discovery AD PWAS −log10(P)
TSPAN14
ETFA
PSEN1
RASA4B
NCK1
SLC44A2
TOM1L2
YARS1
ITDRKH
CDK14
SCAPER
RABEP1
ACSL6
CARM1
INPP5D
SFXN4
ARSA
ANXA11
MFSD6
USP4
RCN2
MINDY1
FAM98C
ADAM17
SLC7A5
RAPGEF2
USP19
METTL14

B

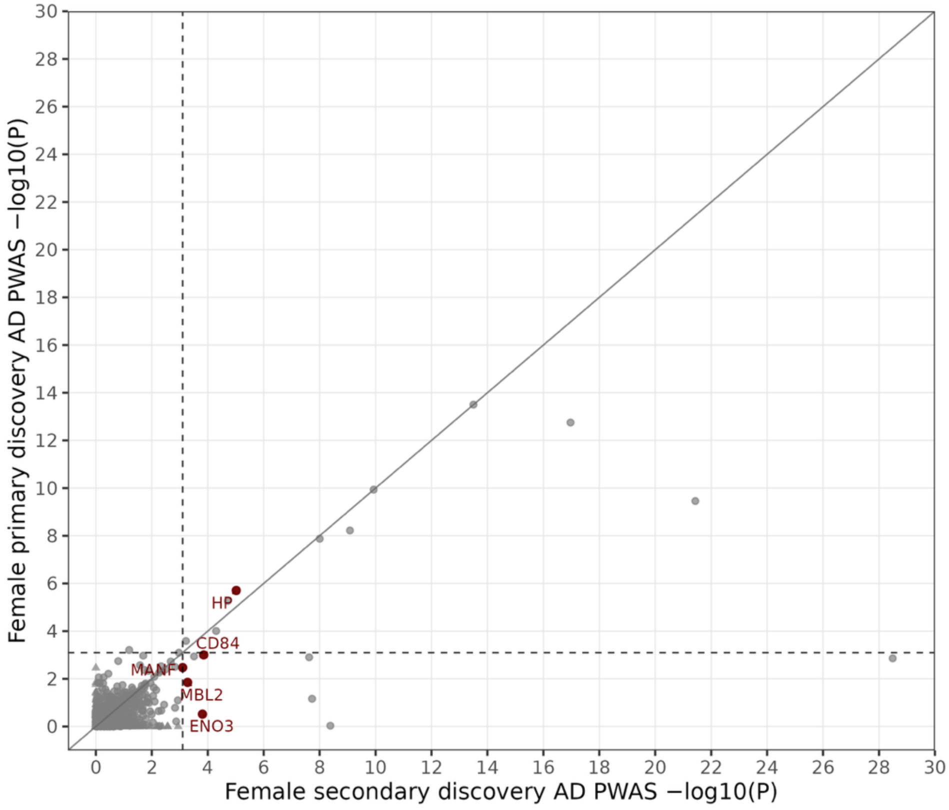
Female primary discovery AD PWAS −log10(P)
Female secondary discovery AD PWAS −log10(P)
HP
CD84
MANF
MBL2
ENO3

C

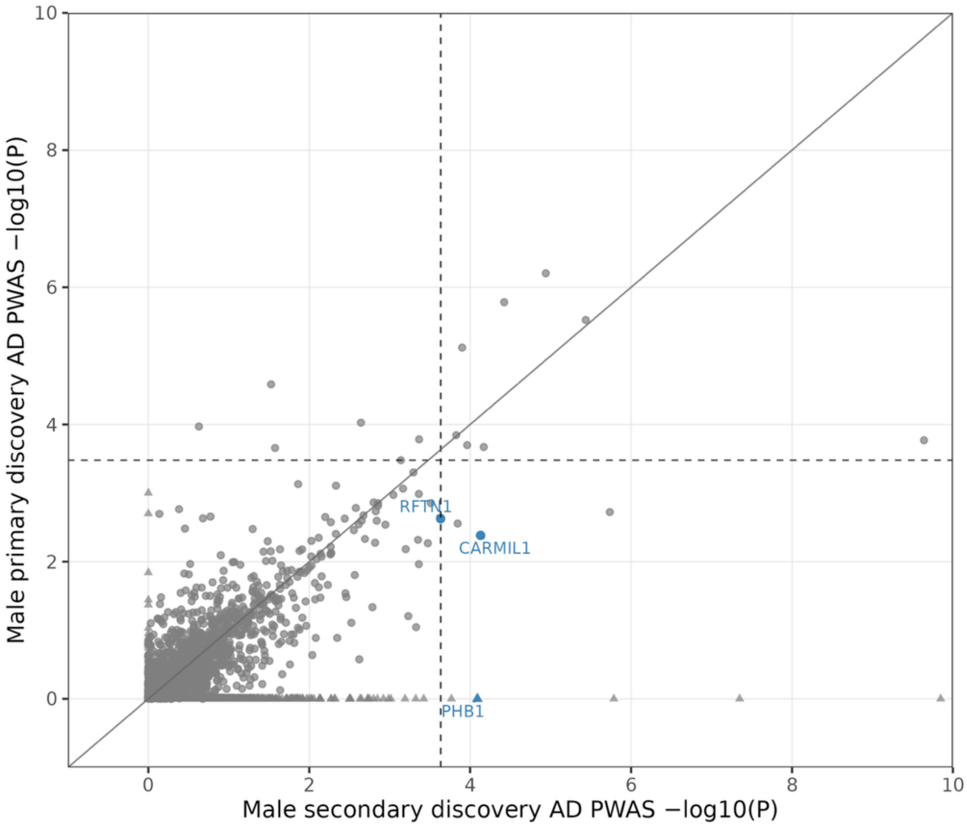
Male primary discovery AD PWAS −log10(P)
Male secondary discovery AD PWAS −log10(P)
RFTN1
CARMIL1
PHB1

D

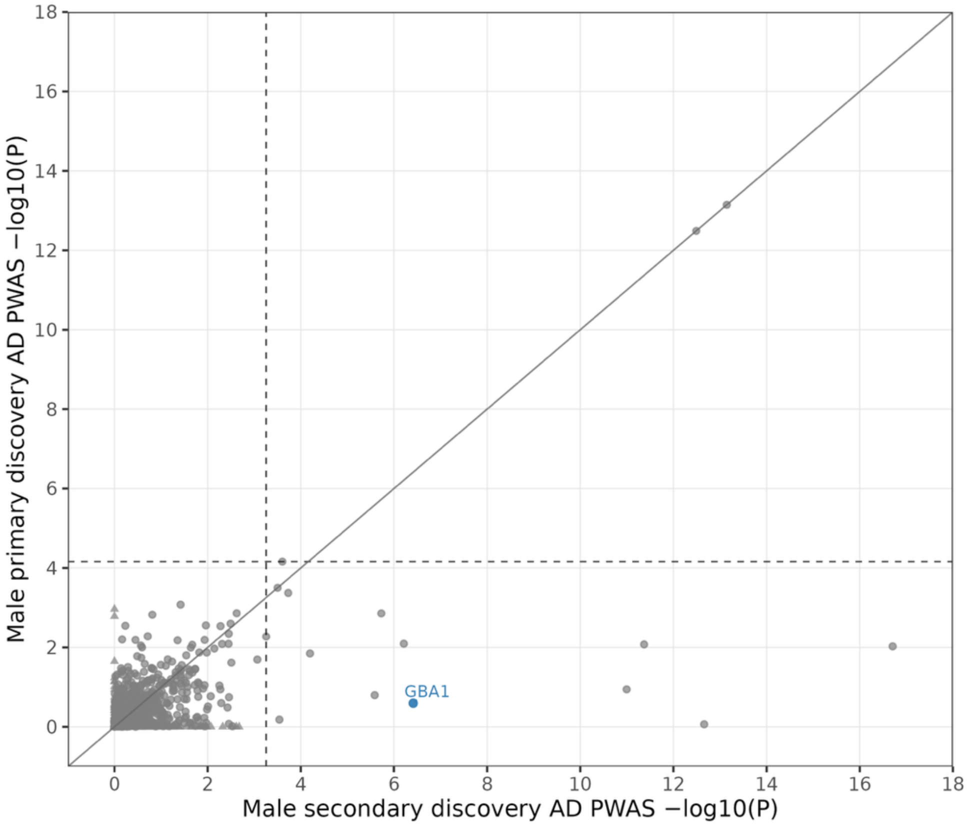
Male primary discovery AD PWAS −log10(P)
Male secondary discovery AD PWAS −log10(P)
GBA1

**Supplementary Figure 16. Comparison of primary and secondary EUR AD PWAS discoveries. A)** Female brain; **B)** Female CSF; **C)** Male brain; **D)** Male CSF. Datapoints in the scatter plots represent PWAS associations between AD and a given protein in brain or aptamer in the CSF for primary (y-axis) and secondary discoveries (x-axis). Only genes passing sex-heterogeneity filters are annotated and colored based on sex discovery. The dashed gray lines denote the proteome-wide significance thresholds at $P_{FDR}$<0.05 (lowest across primary and secondary discoveries). Association tests not observed in one of the discoveries (due to insufficient protein heritability in respective strata) were given a -log10(P) of 0 and are shown as triangles for visualization purposes.

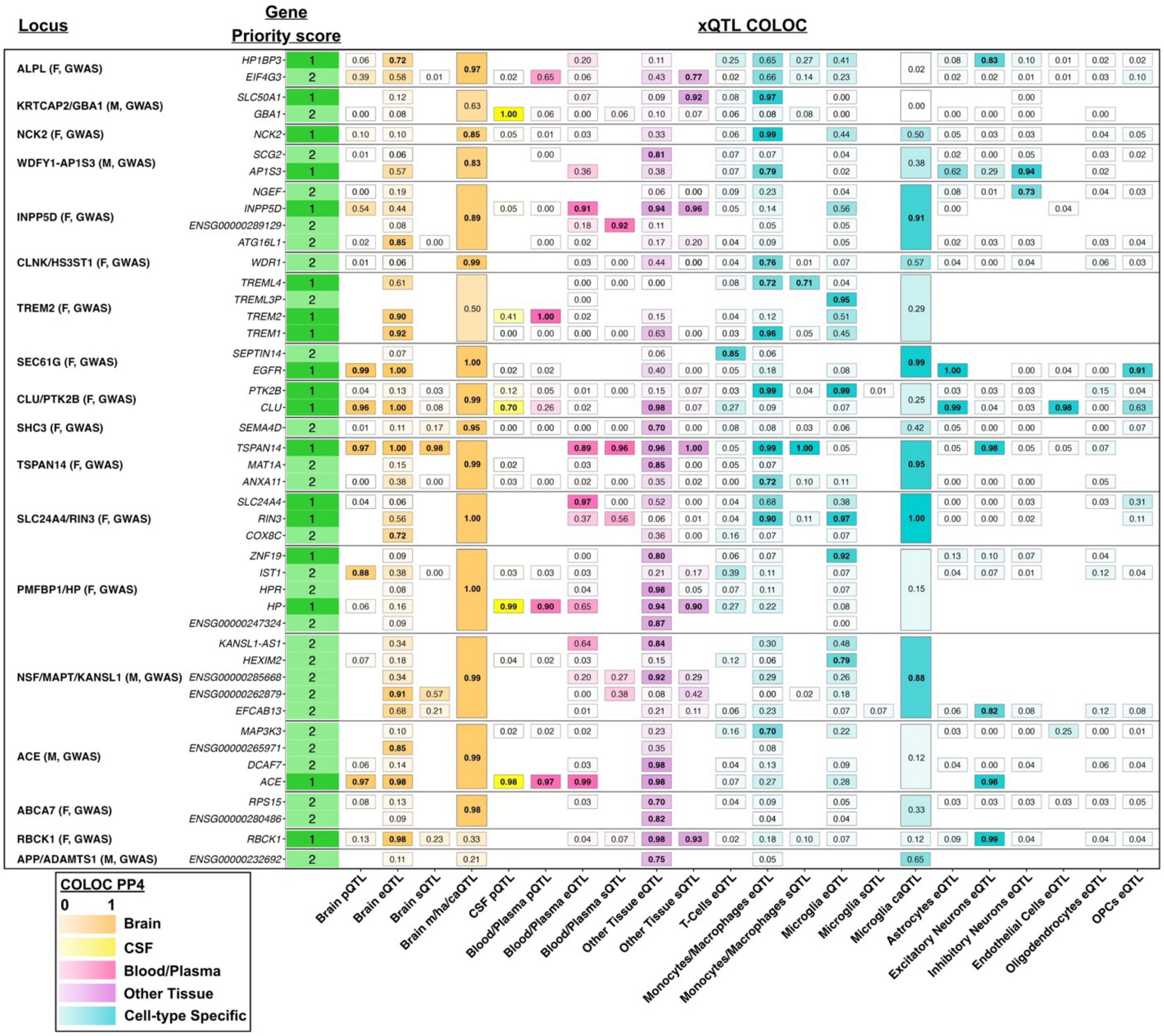

**Supplementary Figure 17. Gene prioritization at sex-specific loci identified in Alzheimer's disease GWAS.** To identify potential causal genes at sex-specific AD GWAS loci, genetic colocalization analyses were performed with various QTL types across different tissues and cell populations. Indicated values represent best posterior probability (PP4) findings out of COLOC.abf or COLOC.SuSiE, and out of multiple tissues or conditions in case of collapsed entries (e.g. 'Other Tissue' shows the best result out of 35 investigated tissues in GTEx v8). Datasets or genes where PP4 was never > 0.7 were not visualized (cf. methods for details regarding gene priority scoring). This figure is an extension of Figure 1, showing all GWAS loci and genes with prioritization support.

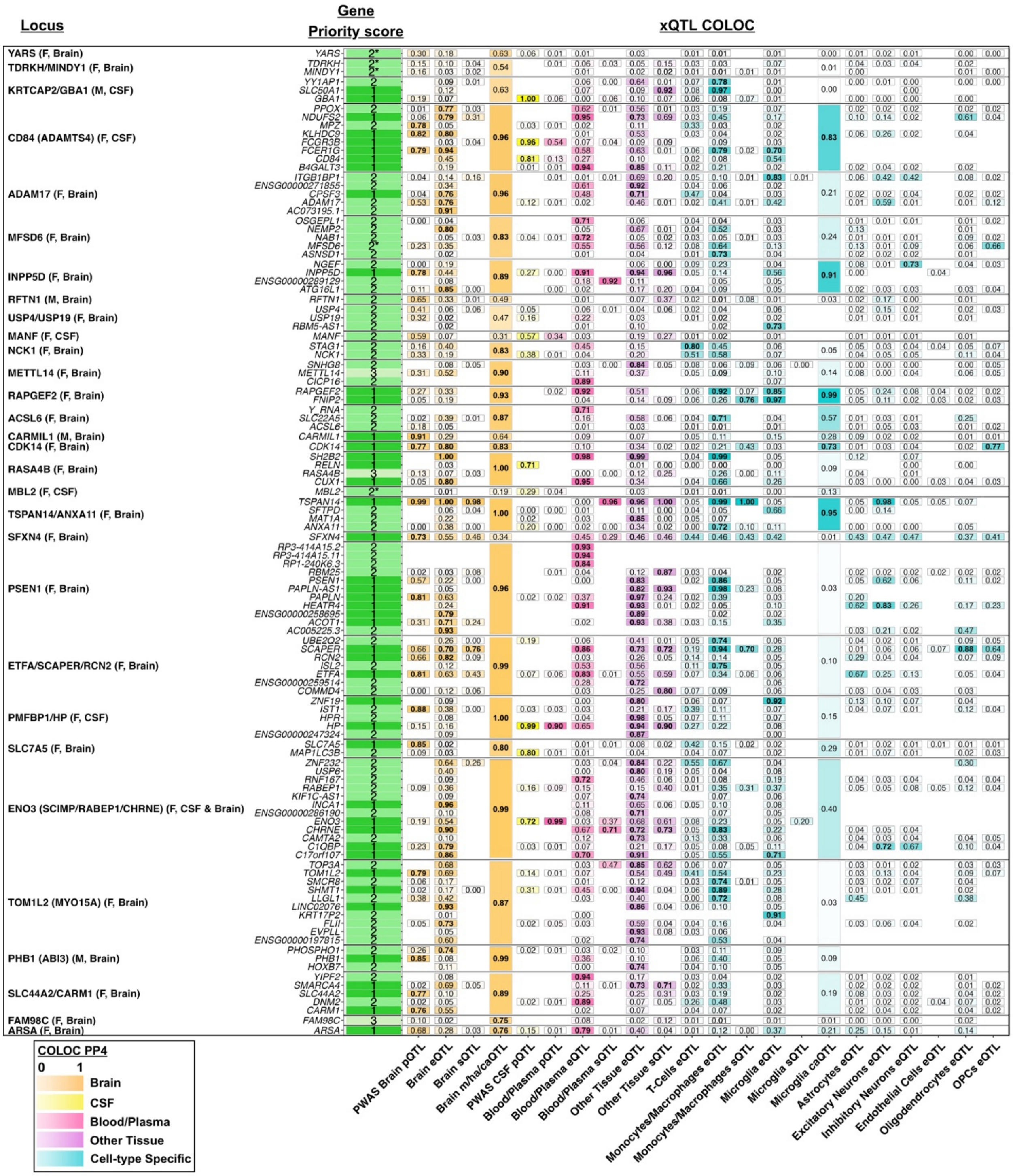

**Supplementary Figure 18. Gene prioritization at sex-specific loci identified in Alzheimer's disease PWAS.** To identify potential causal genes at sex-specific AD PWAS loci, genetic colocalization analyses were performed with various QTL types across different tissues and cell populations. Indicated values represent best posterior probability (PP4) findings out of COLOC.abf or COLOC.SuSiE, and out of multiple tissues or conditions in case of collapsed entries (e.g. ‘Other Tissue’ shows the best result out of 35 investigated tissues in GTEx v8) (cf. methods for details regarding gene priority scoring). This figure is an extension of Figure 2B, showing all PWAS loci and genes with prioritization support.

*Genes with priority score “2*” indicate genes with no xQTL COLOC support, but sex-stratified differential abundance (DAL) support.

S19.1

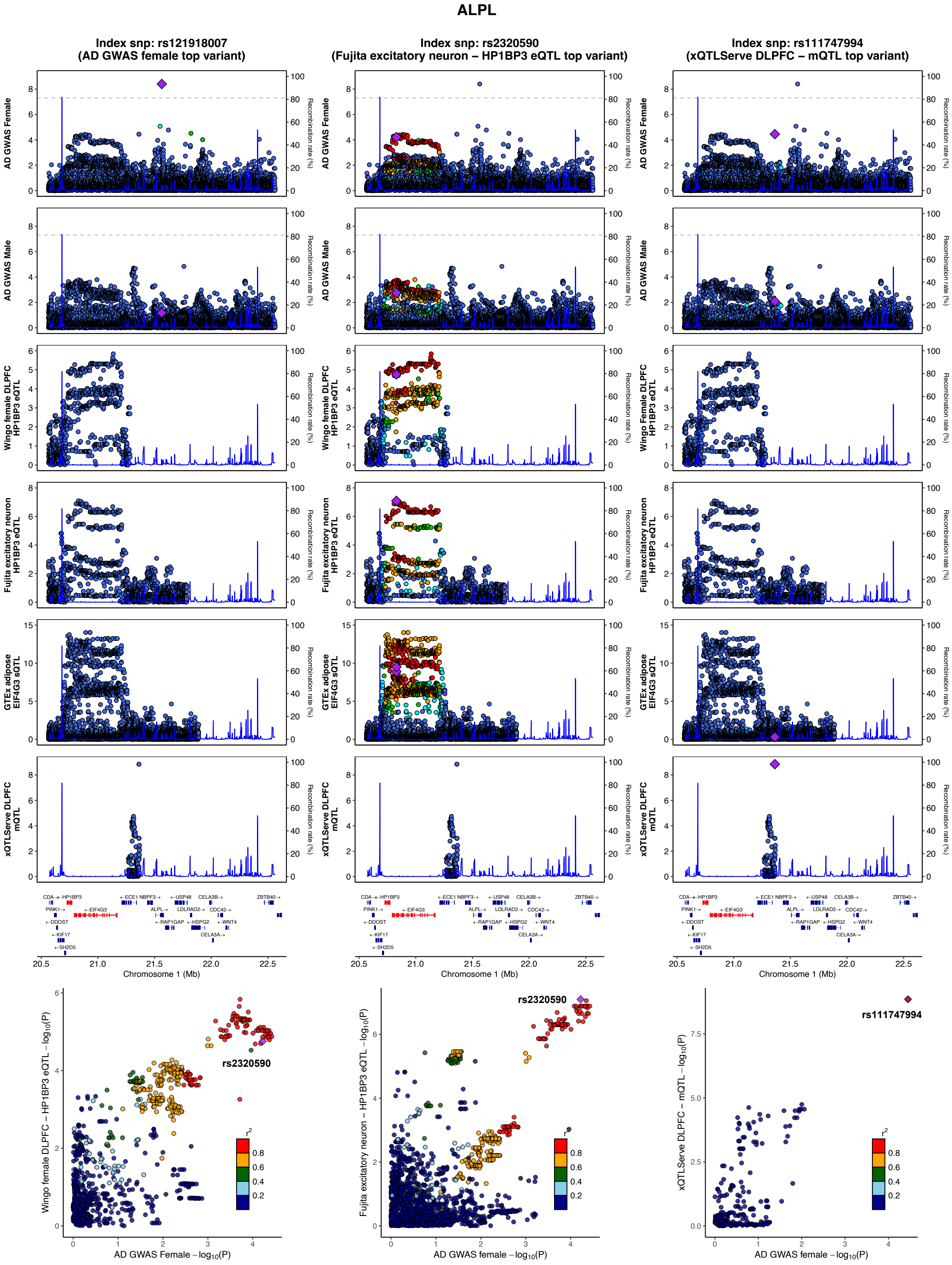

ALPL
Index snp: rs121918007
(AD GWAS female top variant)
Index snp: rs2320590
(Fujita excitatory neuron – HP1BP3 eQTL top variant)
Index snp: rs111747994
(xQTLServe DLPFC – mQTL top variant)
AD GWAS Female
AD GWAS Male
Wingo female DLPFC HP1BP3 eQTL
Fujita excitatory neuron HP1BP3 eQTL
GTEx adipose EIF4G3 sQTL
xQTLServe DLPFC mQTL
Recombination rate (%)
Chromosome 1 (Mb)
rs2320590
rs111747994
Wingo female DLPFC – HP1BP3 eQTL – log10(P)
Fujita excitatory neuron – HP1BP3 eQTL – log10(P)
xQTLServe DLPFC – mQTL – log10(P)
AD GWAS Female – log10(P)
AD GWAS female – log10(P)

**S19.2**

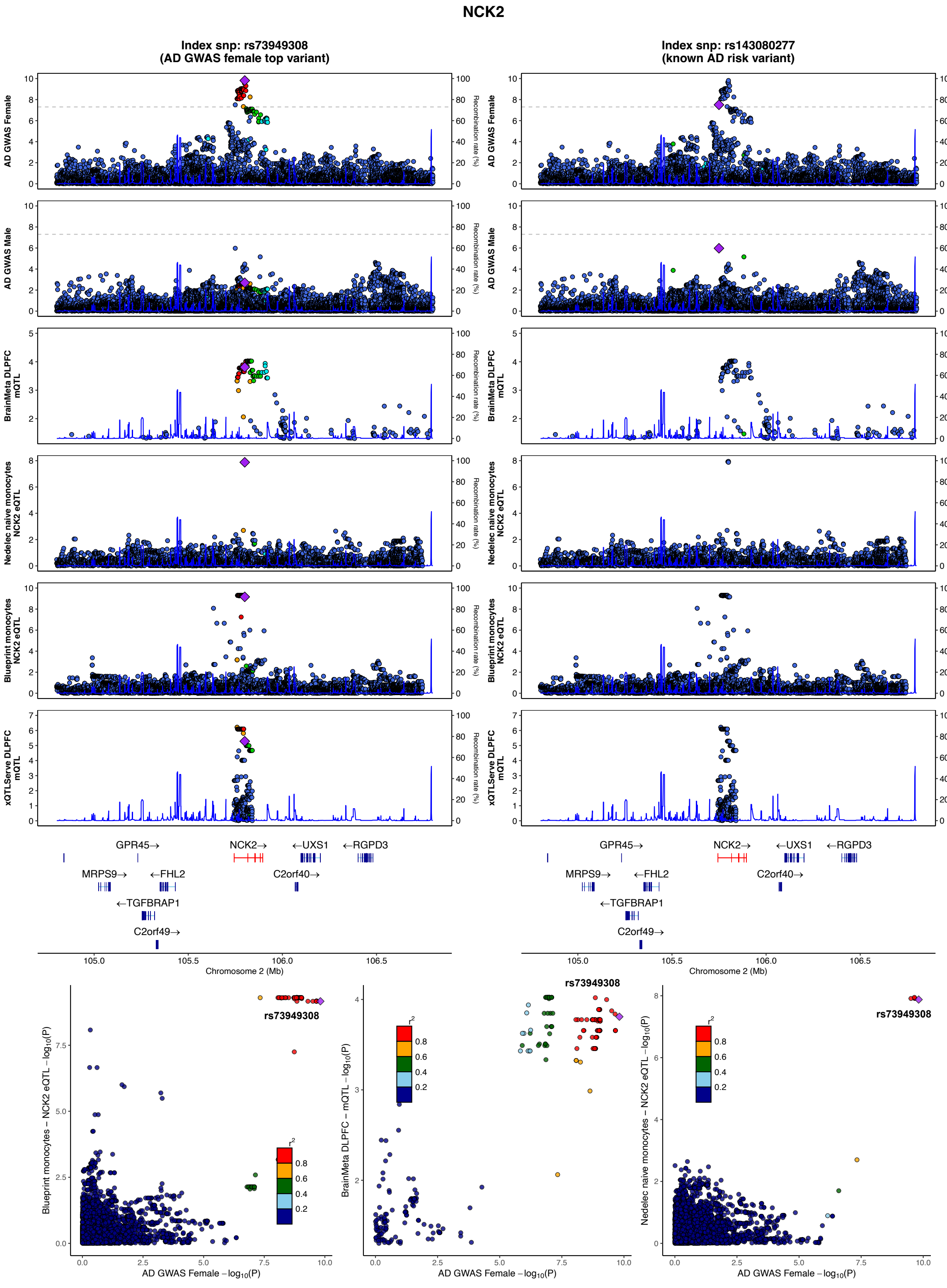
NCK2
Index snp: rs73949308
(AD GWAS female top variant)
Index snp: rs143080277
(known AD risk variant)
AD GWAS Female
AD GWAS Male
BrainMeta DLPFC mQTL
Nedelec naive monocytes NCK2 eQTL
Blueprint monocytes NCK2 eQTL
xQTLServe DLPFC mQTL
Recombination rate (%)
GPR45→
NCK2→
←UXS1
←RGPD3
MRPS9→
←FHL2
C2orf40→
←TGFBRAP1
C2orf49→
Chromosome 2 (Mb)
Blueprint monocytes – NCK2 eQTL – $\log_{10}(P)$
BrainMeta DLPFC – mQTL – $\log_{10}(P)$
Nedelec naive monocytes – NCK2 eQTL – $\log_{10}(P)$
AD GWAS Female – $\log_{10}(P)$
rs73949308
$r^2$

## S19.3

### PMFBP1

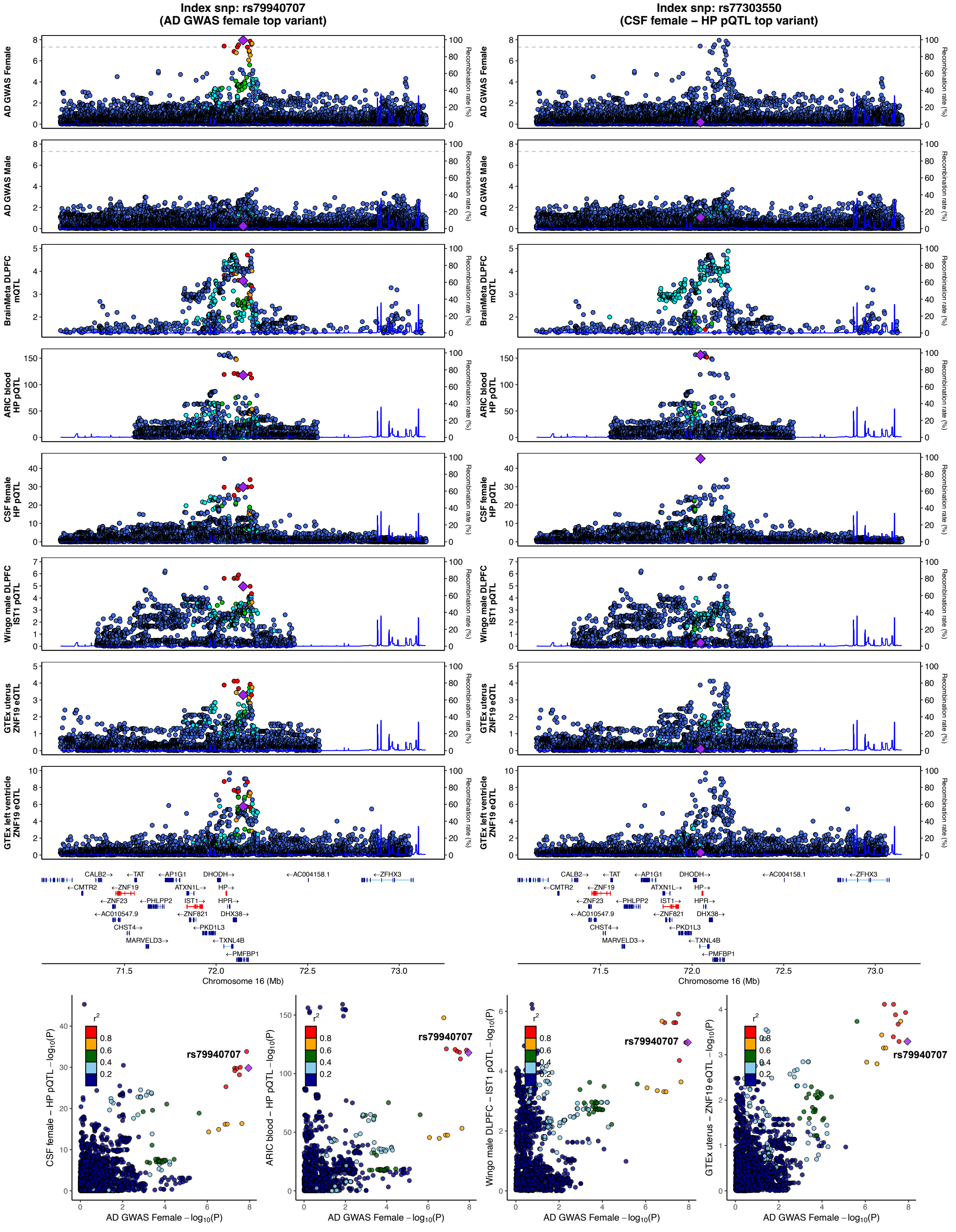

Index snp: rs79940707
(AD GWAS female top variant)
Index snp: rs77303550
(CSF female – HP pQTL top variant)
AD GWAS Female
AD GWAS Male
BrainMeta DLPFC mQTL
ARIC blood HP pQTL
CSF female HP pQTL
Wingo male DLPFC IST1 pQTL
GTEx uterus ZNF19 eQTL
GTEx left ventricle ZNF19 eQTL
Recombination rate (%)
CALB2→
←TAT
←AP1G1
DHODH→
←AC004158.1
←ZFHX3
←CMTR2
←ZNF19
ATXN1L→
HP→
←ZNF23
←PHLPP2
IST1→
HPR→
←AC010547.9
←ZNF821
DHX38→
CHST4→
←PKD1L3
MARVELD3→
←TXNL4B
←PMFBP1
Chromosome 16 (Mb)
rs79940707
r²
CSF female – HP pQTL –log10(P)
ARIC blood – HP pQTL –log10(P)
Wingo male DLPFC – IST1 pQTL –log10(P)
GTEx uterus – ZNF19 eQTL –log10(P)
AD GWAS Female –log10(P)

**S19.4**

**GBA**

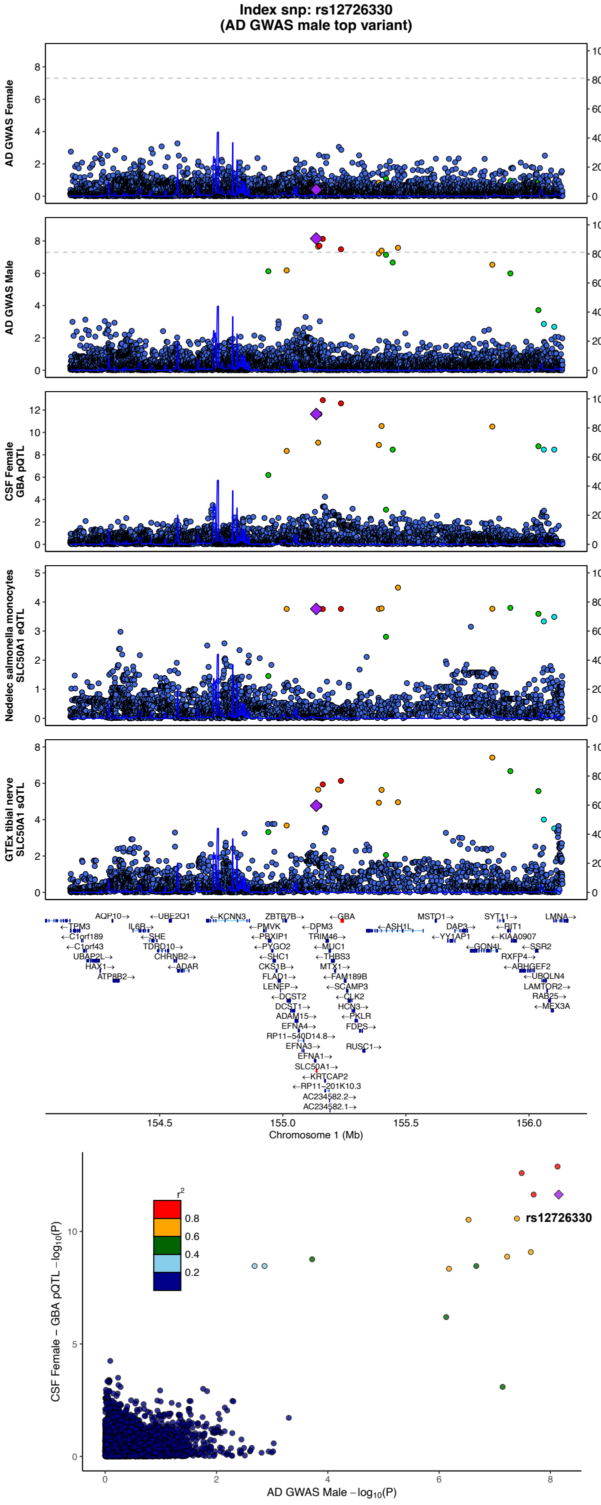

Index snp: rs12726330
(AD GWAS male top variant)
AD GWAS Female
AD GWAS Male
CSF Female
GBA pQTL
Nedelec salmonella monocytes
SLC50A1 eQTL
GTEx tibial nerve
SLC50A1 sQTL
Recombination rate (%)
Chromosome 1 (Mb)
CSF Female – GBA pQTL –log10(P)
AD GWAS Male –log10(P)
rs12726330
r2
0.8
0.6
0.4
0.2

**S19.5**

**SHC3**

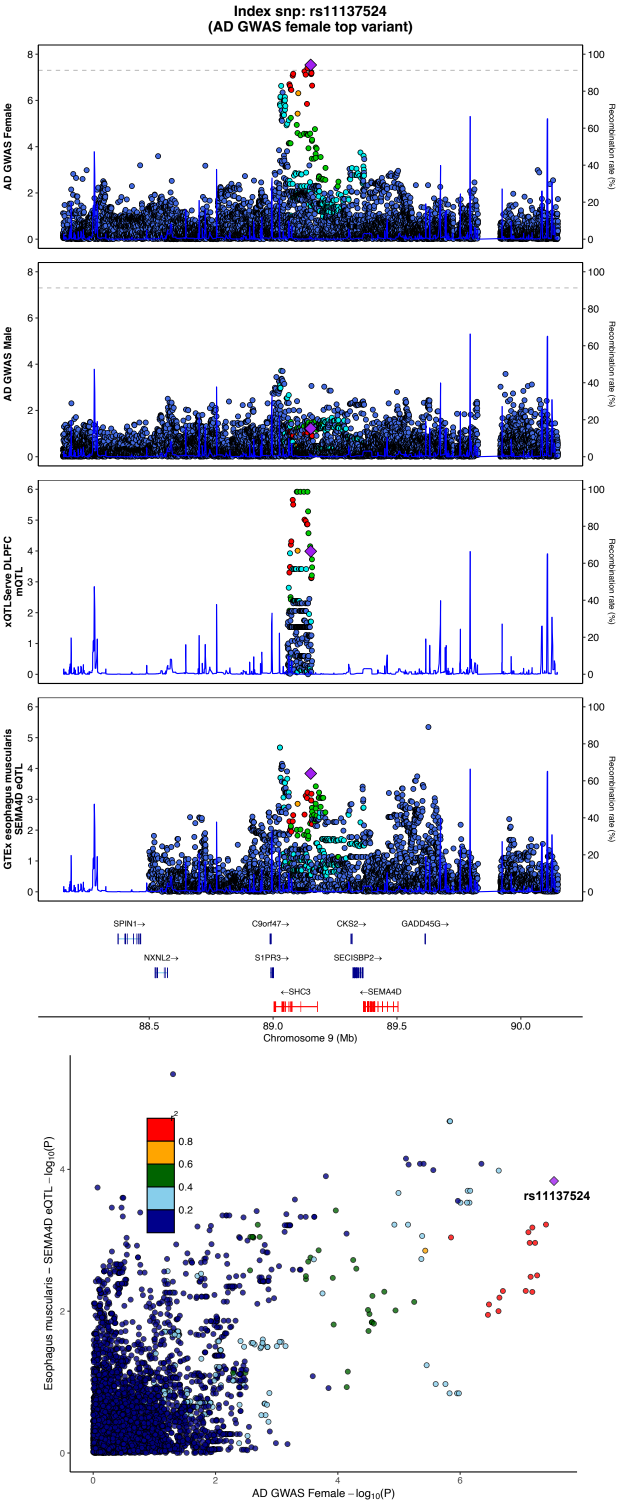

Index snp: rs11137524
(AD GWAS female top variant)
AD GWAS Female
AD GWAS Male
xQTLServe DLPFC mQTL
GTEx esophagus muscularis SEMA4D eQTL
Recombination rate (%)
SPIN1→
C9orf47→
CKS2→
GADD45G→
NXNL2→
S1PR3→
SECISBP2→
←SHC3
←SEMA4D
Chromosome 9 (Mb)
88.5
89.0
89.5
90.0
Esophagus muscularis − SEMA4D eQTL −log10(P)
AD GWAS Female −log10(P)
rs11137524
0.8
0.6
0.4
0.2

**S19.6**

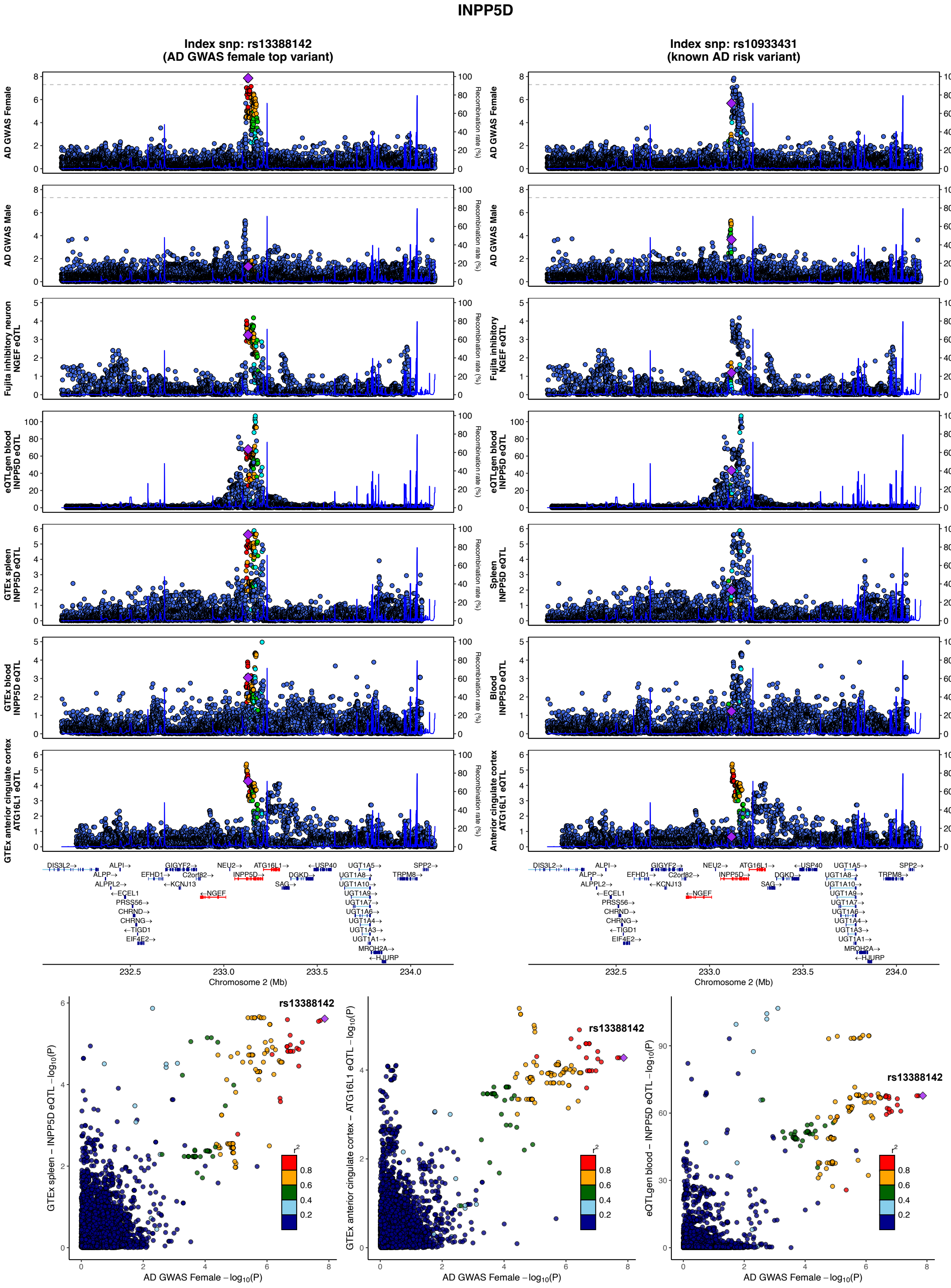

INPP5D
Index snp: rs13388142
(AD GWAS female top variant)
Index snp: rs10933431
(known AD risk variant)
AD GWAS Female
AD GWAS Male
Fujita inhibitory neuron NGEF eQTL
eQTLgen blood INPP5D eQTL
GTEx spleen INPP5D eQTL
GTEx blood INPP5D eQTL
GTEx anterior cingulate cortex ATG16L1 eQTL
Recombination rate (%)
Chromosome 2 (Mb)
rs13388142
AD GWAS Female −log10(P)
GTEx spleen − INPP5D eQTL −log10(P)
GTEx anterior cingulate cortex − ATG16L1 eQTL −log10(P)
eQTLgen blood − INPP5D eQTL −log10(P)

## S19.7

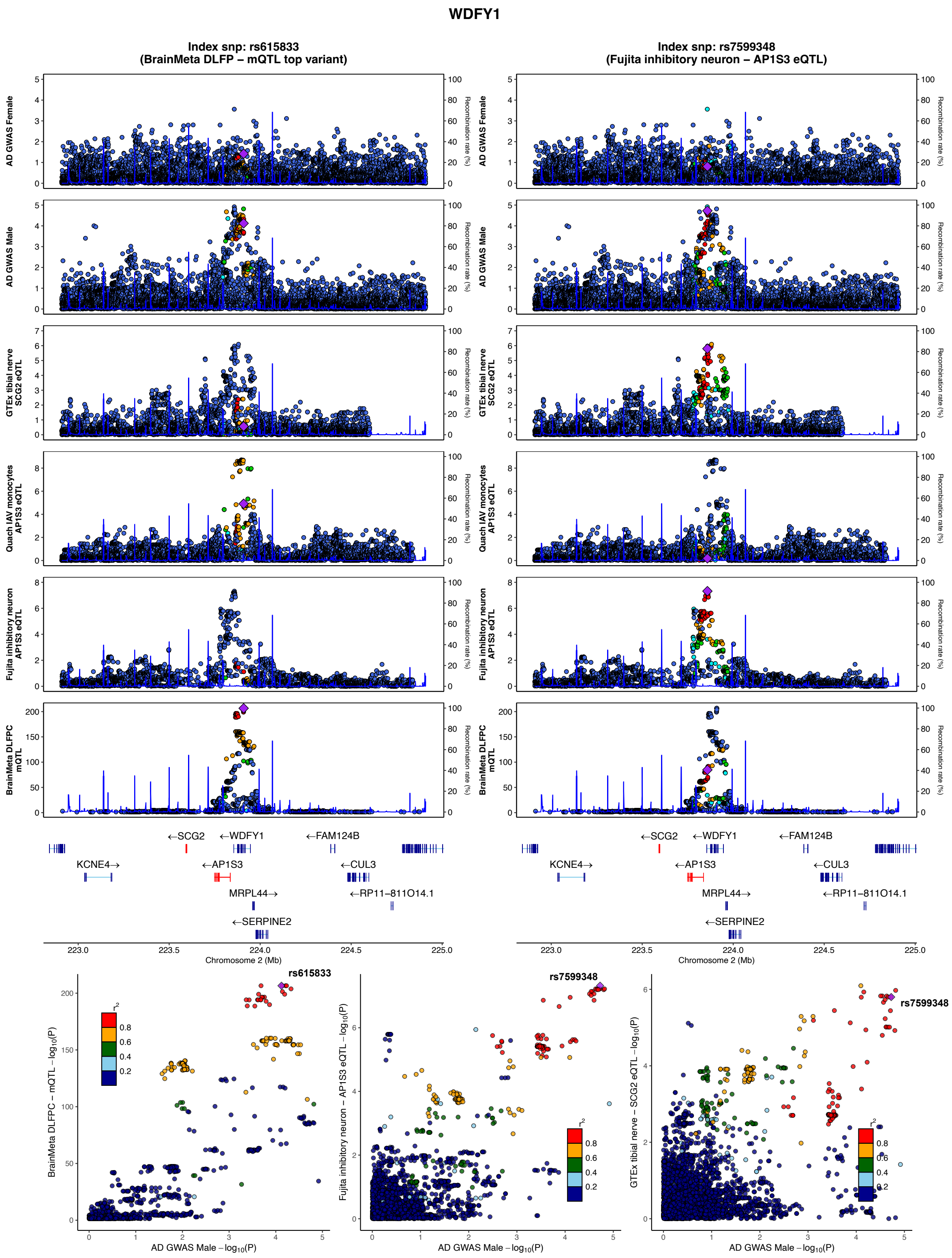

WDFY1
Index snp: rs615833
(BrainMeta DLFP – mQTL top variant)
Index snp: rs7599348
(Fujita inhibitory neuron – AP1S3 eQTL)
AD GWAS Female
AD GWAS Male
GTEx tibial nerve SCG2 eQTL
Quach IAV monocytes AP1S3 eQTL
Fujita inhibitory neuron AP1S3 eQTL
BrainMeta DLFPC mQTL
Recombination rate (%)
←SCG2
←WDFY1
←FAM124B
KCNE4→
←AP1S3
←CUL3
MRPL44→
←RP11-811O14.1
←SERPINE2
Chromosome 2 (Mb)
rs615833
rs7599348
BrainMeta DLFPC – mQTL –log10(P)
Fujita inhibitory neuron – AP1S3 eQTL –log10(P)
GTEx tibial nerve – SCG2 eQTL –log10(P)
AD GWAS Male –log10(P)
r2

**S19.8**

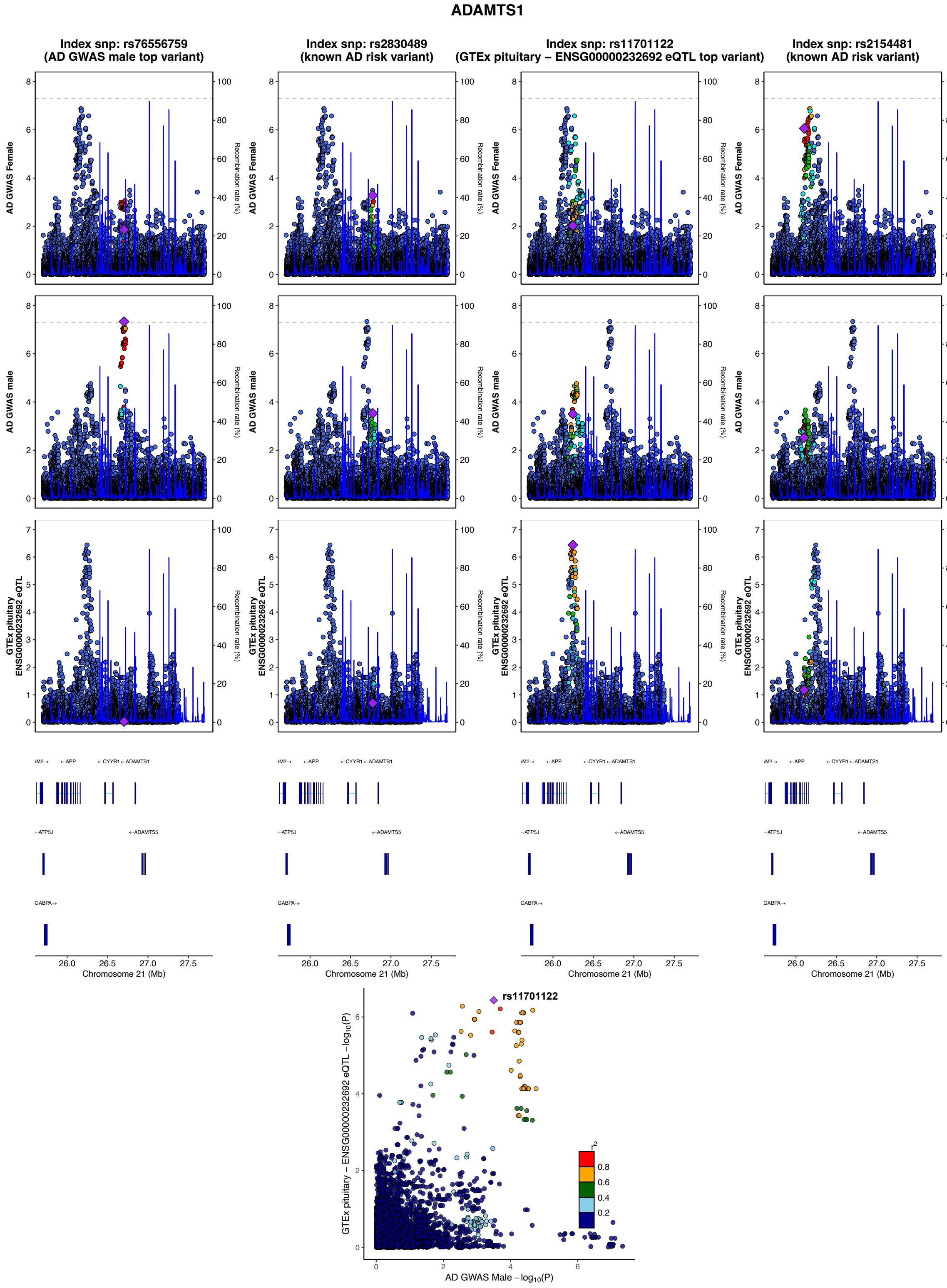

ADAMTS1
Index snp: rs76556759
(AD GWAS male top variant)
Index snp: rs2830489
(known AD risk variant)
Index snp: rs11701122
(GTEx pituitary – ENSG00000232692 eQTL top variant)
Index snp: rs2154481
(known AD risk variant)
AD GWAS Female
AD GWAS male
GTEx pituitary ENSG00000232692 eQTL
Recombination rate (%)
←APP
←CYYR1 ←ADAMTS1
←ATP5J
←ADAMTS5
GABPA→
Chromosome 21 (Mb)
rs11701122
GTEx pituitary – ENSG00000232692 eQTL – log10(P)
AD GWAS Male – log10(P)
r2

**S19.9**

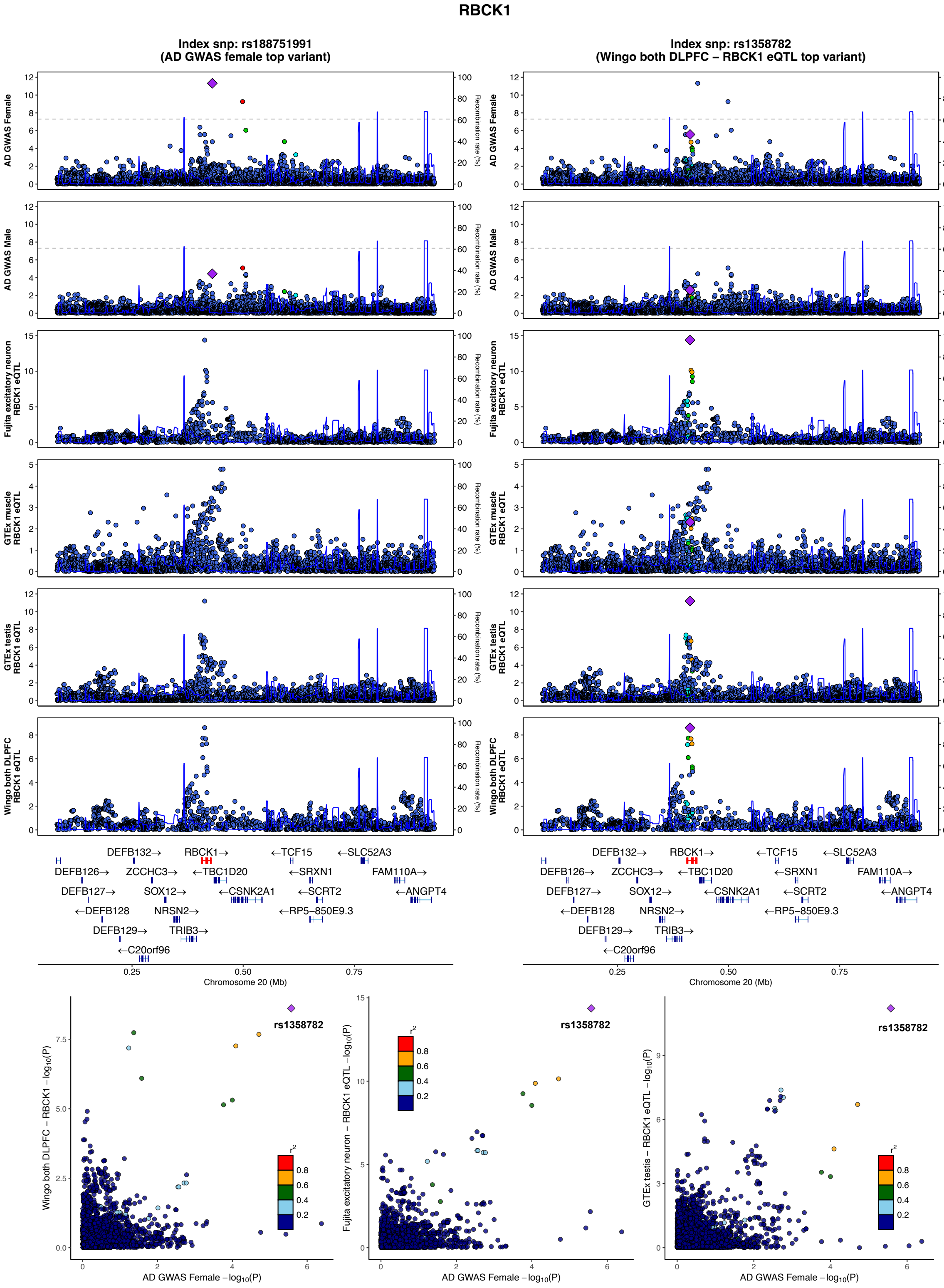
RBCK1
Index snp: rs188751991
(AD GWAS female top variant)
Index snp: rs1358782
(Wingo both DLPFC – RBCK1 eQTL top variant)
AD GWAS Female
AD GWAS Male
Fujita excitatory neuron RBCK1 eQTL
GTEx muscle RBCK1 eQTL
GTEx testis RBCK1 eQTL
Wingo both DLPFC RBCK1 eQTL
Recombination rate (%)
DEFB132→
RBCK1→
←TCF15
←SLC52A3
DEFB126→
ZCCHC3→
←TBC1D20
←SRXN1
FAM110A→
DEFB127→
SOX12→
←CSNK2A1
←SCRT2
←ANGPT4
←DEFB128
NRSN2→
←RP5−850E9.3
DEFB129→
TRIB3→
←C20orf96
Chromosome 20 (Mb)
rs1358782
Wingo both DLPFC – RBCK1 −log10(P)
Fujita excitatory neuron – RBCK1 eQTL −log10(P)
GTEx testis – RBCK1 eQTL −log10(P)
AD GWAS Female −log10(P)
r2

**Supplementary Figure 19. Illustrative AD GWAS locus zoom plots highlighting xQTL colocalizations.** The illustrative GWAS hits included are **1)** *ALPL*; **2)** *NCK2*; **3)** *PMFBP1*; **4)** *GBA1*; **5)** *SHC3*; **6)** *INPP5D*; **7)** *WDFY1*; **8)** *ADAMTS1*; **9)** *RBCK1*. Figures with more than one LocusZoom column indicate the presence of more than one independent signal at the locus contributing to QTL colocalizations. Dot color coding represents LD with tagging variant (purple diamond): red – $R^2$: 0.8-1.0, orange – $R^2$:0.6-0.8, green – $R^2$:0.4-0.6, cyan – $R^2$:0.2-0.4, blue – $R^2$:0.0-0.2.

S20.1

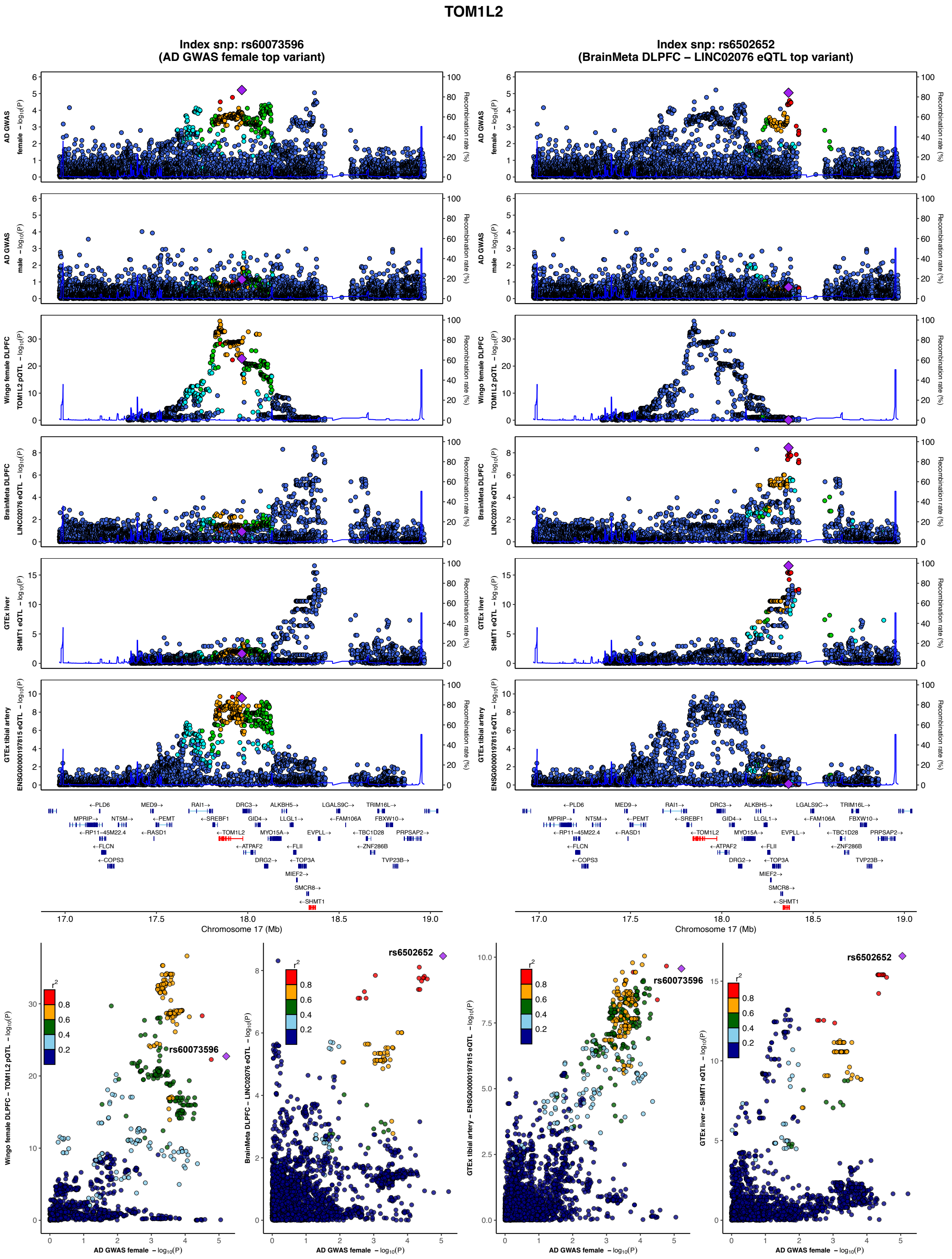
TOM1L2
Index snp: rs60073596
(AD GWAS female top variant)
Index snp: rs6502652
(BrainMeta DLPFC – LINC02076 eQTL top variant)
AD GWAS female – log10(P)
AD GWAS male – log10(P)
Wingo female DLPFC TOM1L2 pQTL – log10(P)
BrainMeta DLPFC LINC02076 eQTL – log10(P)
GTEx liver SHMT1 eQTL – log10(P)
GTEx tibial artery ENSG00000197815 eQTL – log10(P)
Recombination rate (%)
Chromosome 17 (Mb)
rs60073596
rs6502652
r2
0.8
0.6
0.4
0.2
AD GWAS female – log10(P)

**S20.2**

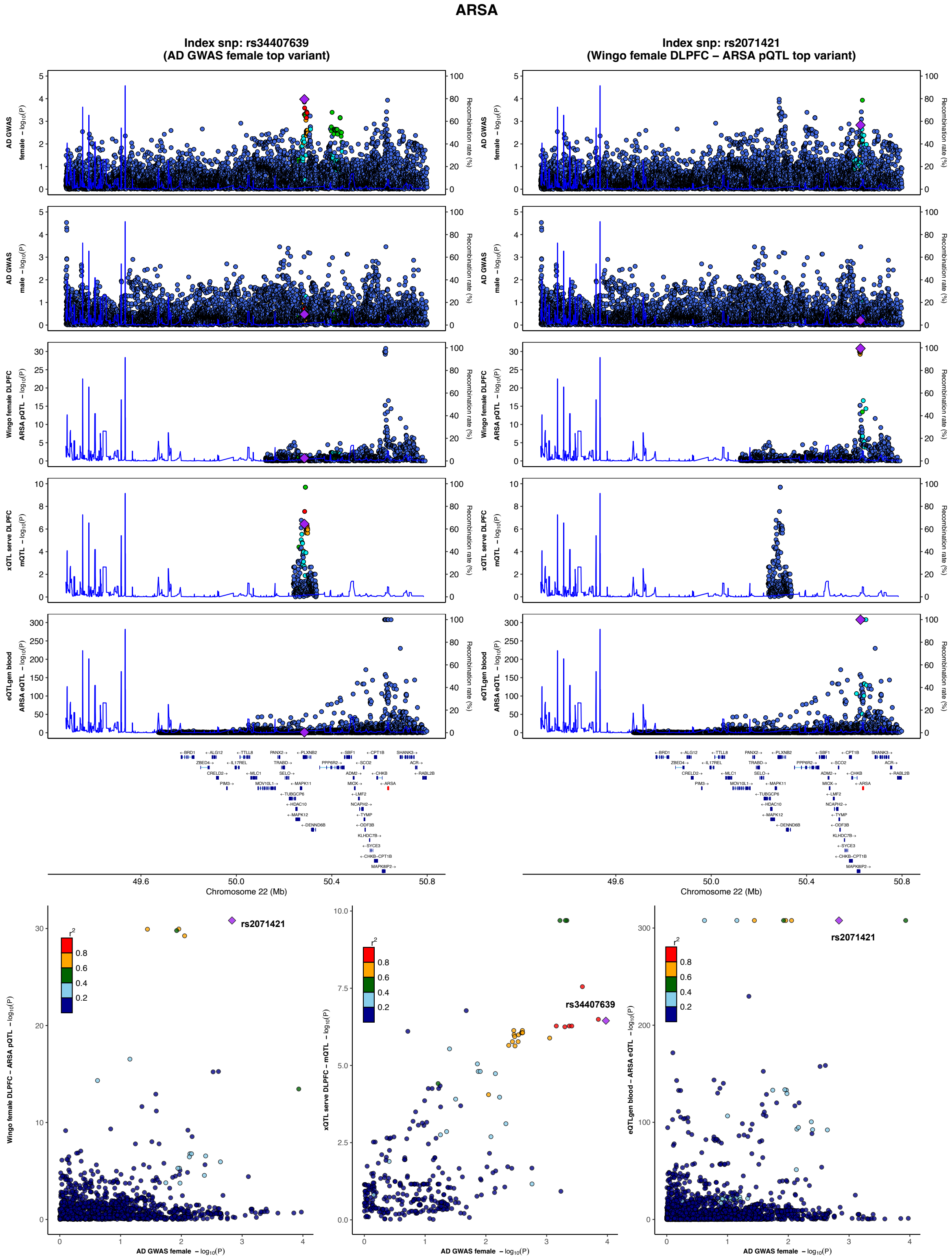

ARSA
Index snp: rs34407639
(AD GWAS female top variant)
Index snp: rs2071421
(Wingo female DLPFC – ARSA pQTL top variant)
AD GWAS female – log10(P)
AD GWAS male – log10(P)
Wingo female DLPFC ARSA pQTL – log10(P)
xQTL serve DLPFC mQTL – log10(P)
eQTLgen blood ARSA eQTL – log10(P)
Recombination rate (%)
Chromosome 22 (Mb)
49.6
50.0
50.4
50.8
rs2071421
rs34407639
r2
0.8
0.6
0.4
0.2
AD GWAS female – log10(P)
Wingo female DLPFC – ARSA pQTL – log10(P)
xQTL serve DLPFC – mQTL – log10(P)
eQTLgen blood – ARSA eQTL – log10(P)

**S20.3**

#### SLC7A5

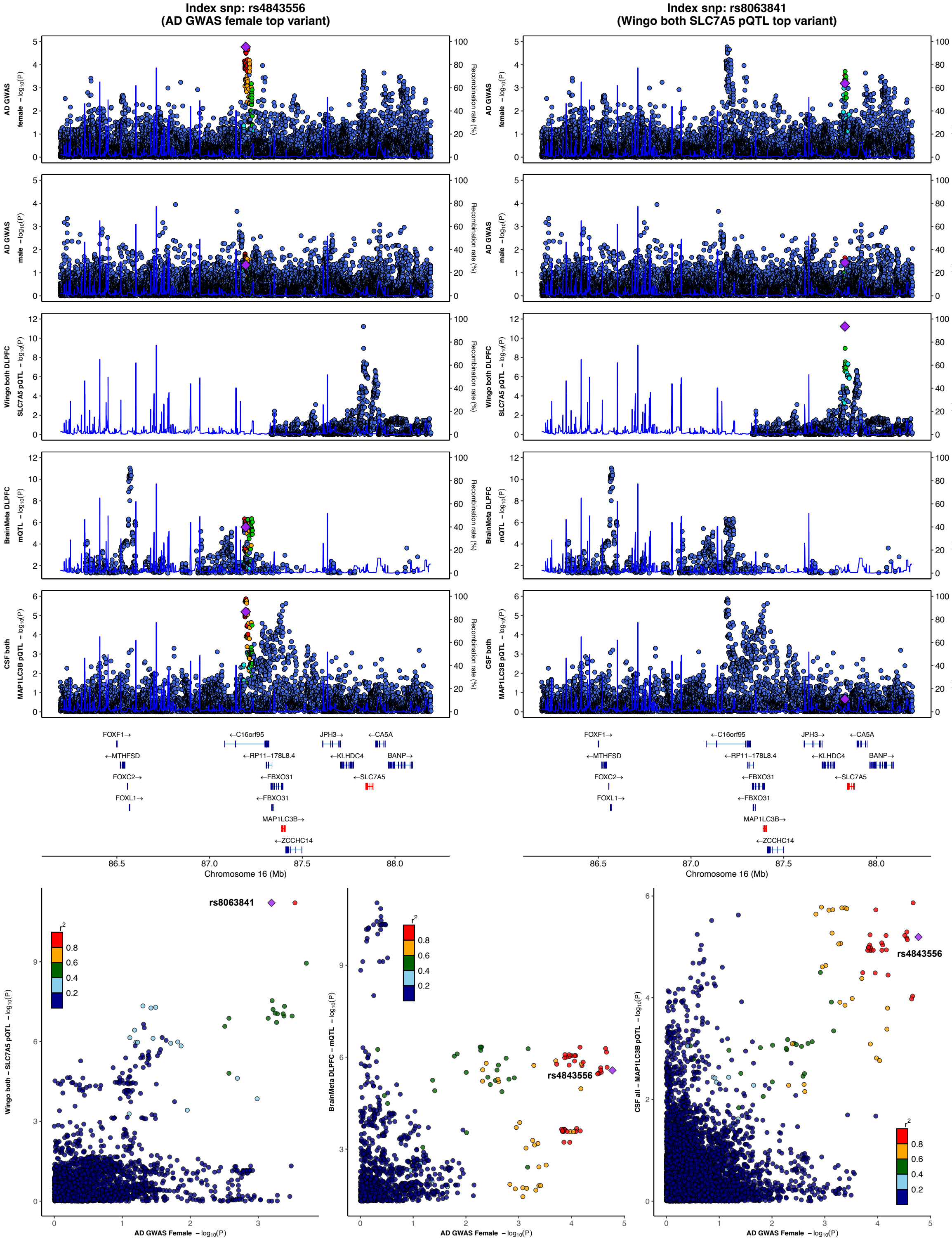

Index snp: rs4843556
(AD GWAS female top variant)
Index snp: rs8063841
(Wingo both SLC7A5 pQTL top variant)
AD GWAS female − log10(P)
AD GWAS male − log10(P)
Wingo both DLPFC SLC7A5 pQTL − log10(P)
BrainMeta DLPFC mQTL − log10(P)
CSF both MAP1LC3B pQTL − log10(P)
Recombination rate (%)
FOXF1→
←MTHFSD
FOXC2→
FOXL1→
←C16orf95
←RP11−178L8.4
←FBXO31
←FBXO31
MAP1LC3B→
←ZCCHC14
JPH3→
←KLHDC4
←CA5A
BANP→
←SLC7A5
Chromosome 16 (Mb)
rs8063841
rs4843556
Wingo both − SLC7A5 pQTL − log10(P)
BrainMeta DLPFC − mQTL − log10(P)
CSF all − MAP1LC3B pQTL − log10(P)
AD GWAS Female − log10(P)

S20.4

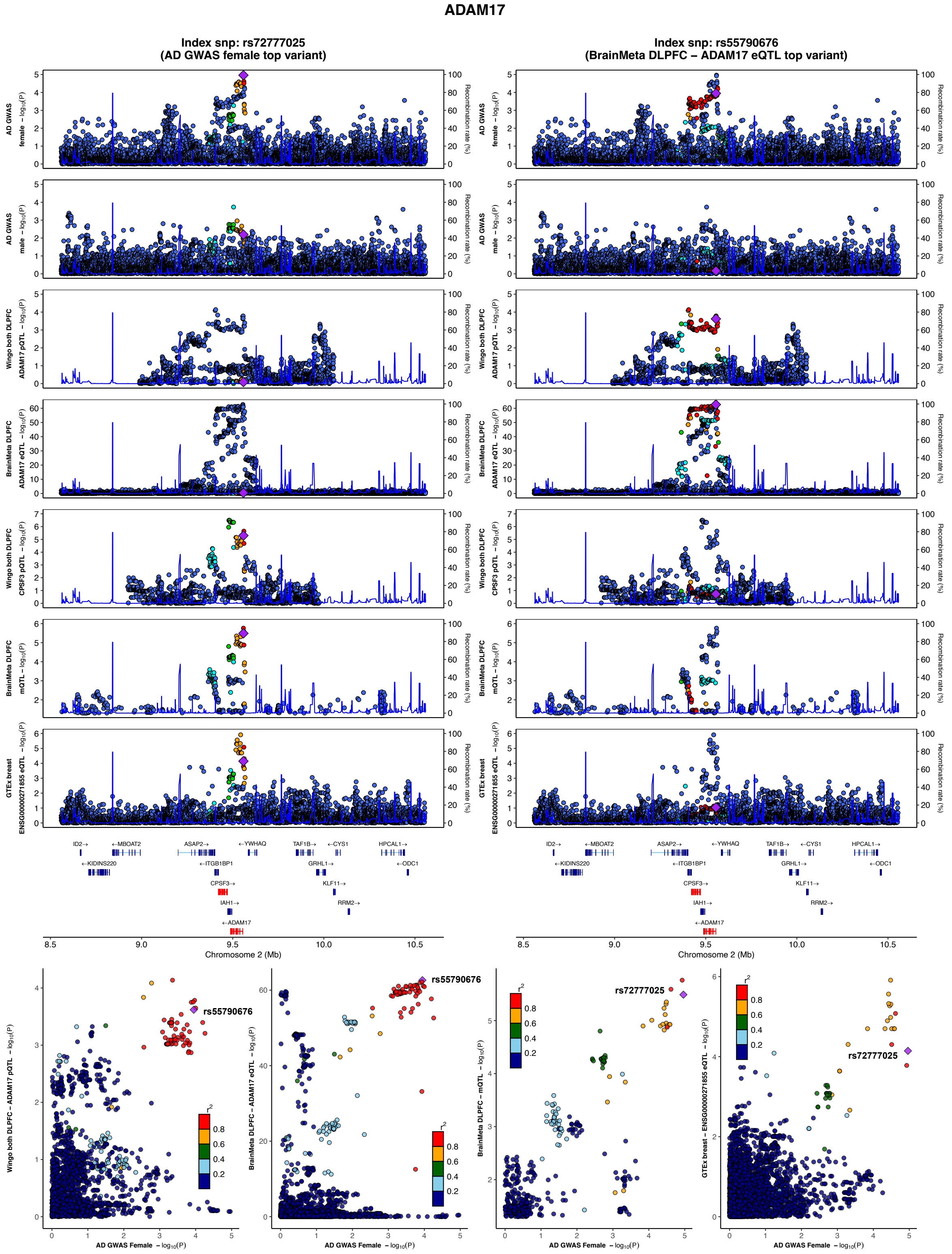
ADAM17
Index snp: rs72777025
(AD GWAS female top variant)
Index snp: rs55790676
(BrainMeta DLPFC – ADAM17 eQTL top variant)
AD GWAS female – log10(P)
AD GWAS male – log10(P)
Wingo both DLPFC ADAM17 pQTL – log10(P)
BrainMeta DLPFC ADAM17 eQTL – log10(P)
Wingo both DLPFC CPSF3 pQTL – log10(P)
BrainMeta DLPFC mQTL – log10(P)
GTEx breast ENSG00000271855 eQTL – log10(P)
Recombination rate (%)
ID2→
←MBOAT2
←KIDINS220
ASAP2→
←ITGB1BP1
CPSF3→
IAH1→
←ADAM17
←YWHAQ
TAF1B→
←CYS1
GRHL1→
KLF11→
RRM2→
HPCAL1→
←ODC1
Chromosome 2 (Mb)
rs55790676
rs72777025
r2
AD GWAS Female – log10(P)
Wingo both DLPFC – ADAM17 pQTL – log10(P)
BrainMeta DLPFC – ADAM17 eQTL – log10(P)
BrainMeta DLPFC – mQTL – log10(P)
GTEx breast – ENSG00000271855 eQTL – log10(P)

S20.5

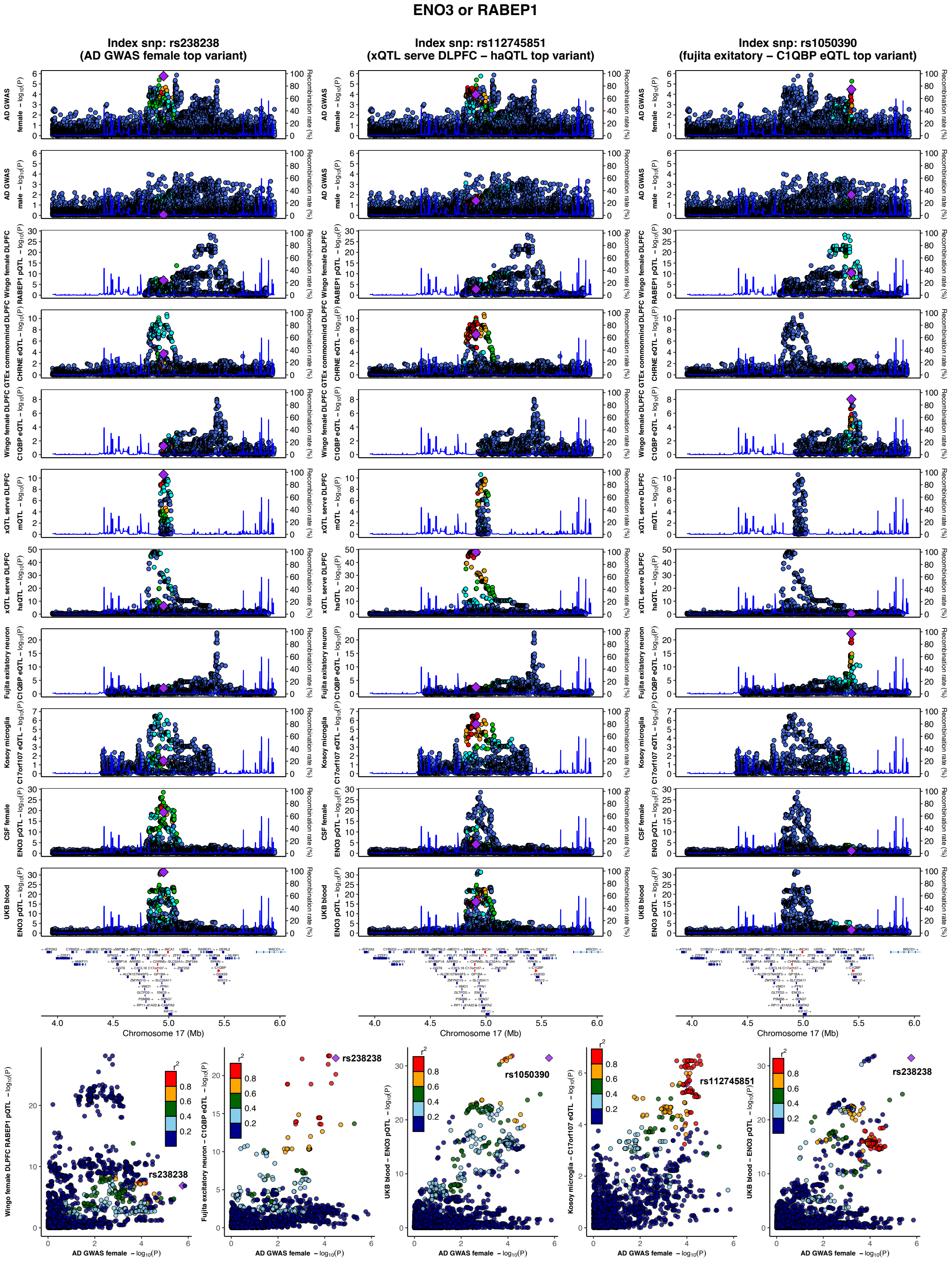

ENO3 or RABEP1
Index snp: rs238238
(AD GWAS female top variant)
Index snp: rs112745851
(xQTL serve DLPFC – haQTL top variant)
Index snp: rs1050390
(fujita exitatory – C1QBP eQTL top variant)
AD GWAS female
AD GWAS male
Wingo female DLPFC RABEP1 pQTL
GTEx commonmind DLPFC CHRNE eQTL
Wingo female DLPFC C1QBP eQTL
xQTL serve DLPFC mQTL
xQTL serve DLPFC haQTL
Fujita exitatory neuron C1QBP eQTL
Kosoy microglia C17orf107 eQTL
CSF female ENO3 pQTL
UKB blood ENO3 pQTL
Recombination rate (%)
Chromosome 17 (Mb)
AD GWAS female – log10(P)
rs238238
rs1050390
rs112745851

**S20.6**

#### PHB1

**Index snp: rs12944684**
**(AD GWAS male top variant)**

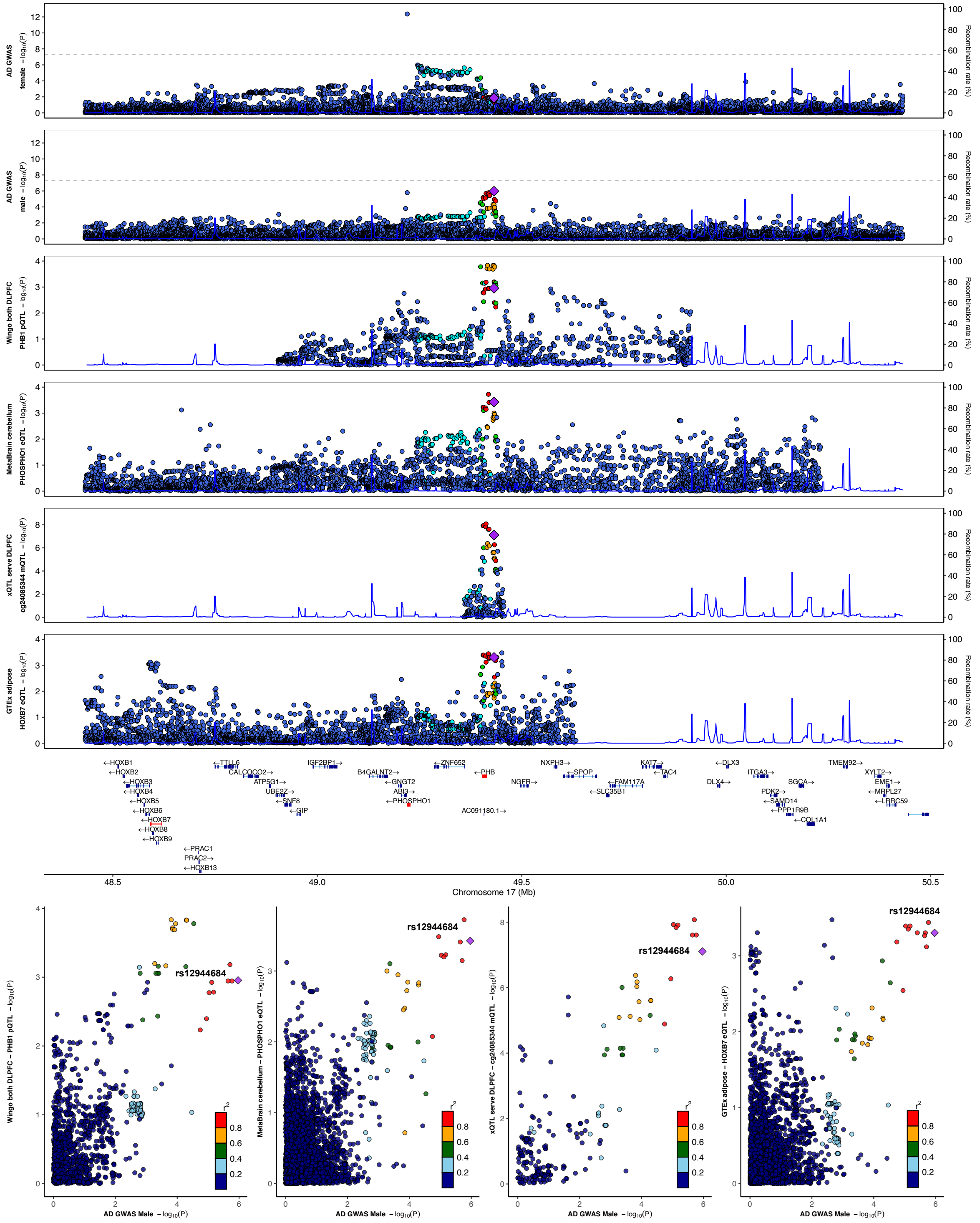

S20.7

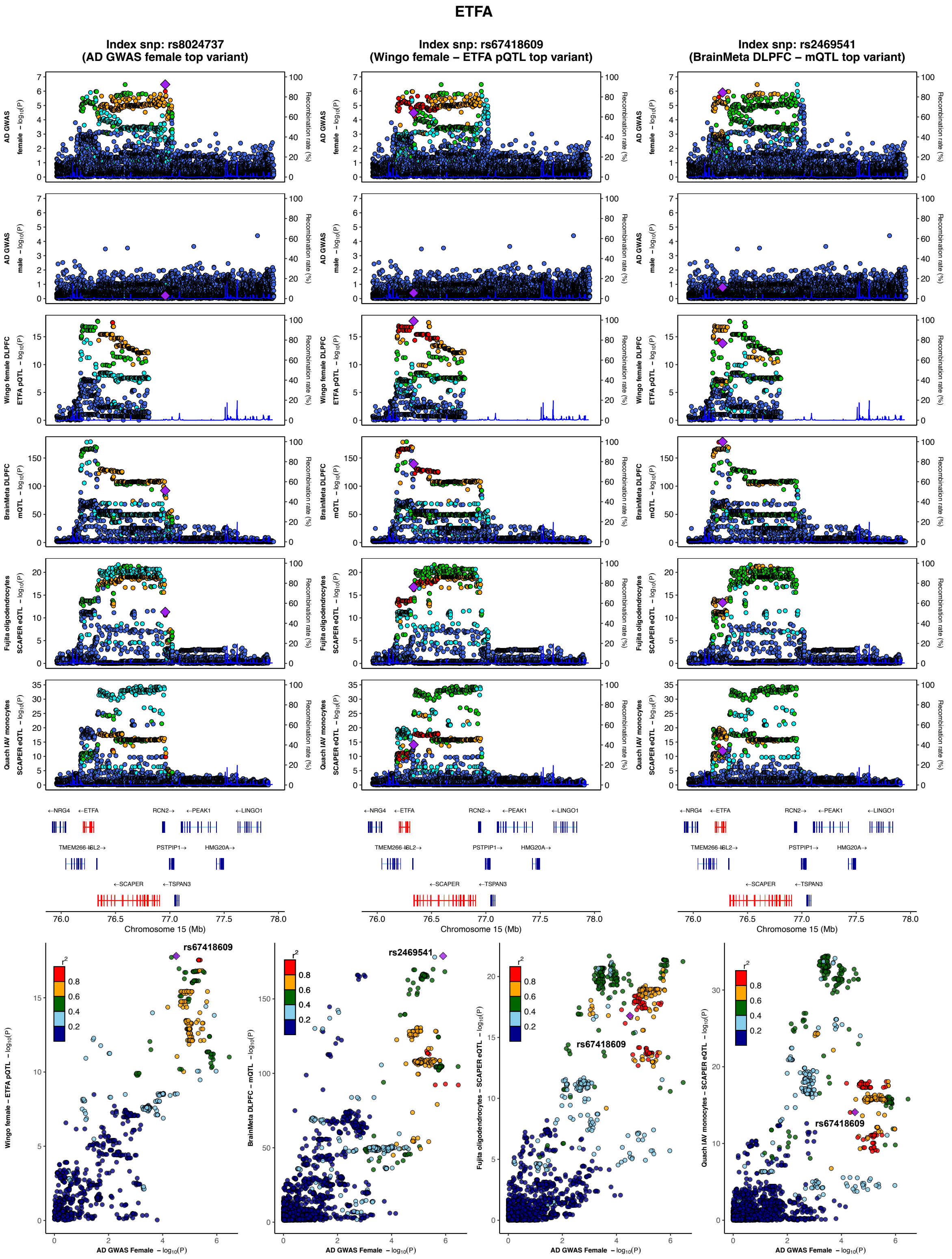
ETFA
Index snp: rs8024737 (AD GWAS female top variant)
Index snp: rs67418609 (Wingo female – ETFA pQTL top variant)
Index snp: rs2469541 (BrainMeta DLPFC – mQTL top variant)
AD GWAS female – log10(P)
AD GWAS male – log10(P)
Wingo female DLPFC ETFA pQTL – log10(P)
BrainMeta DLPFC mQTL – log10(P)
Fujita oligodendrocytes SCAPER eQTL – log10(P)
Quach IAV monocytes SCAPER eQTL – log10(P)
Recombination rate (%)
←NRG4
←ETFA
RCN2→
←PEAK1
←LINGO1
TMEM266
ISL2→
PSTPIP1→
HMG20A→
←SCAPER
←TSPAN3
Chromosome 15 (Mb)
rs67418609
rs2469541
r2
AD GWAS Female – log10(P)
Wingo female – ETFA pQTL – log10(P)
BrainMeta DLPFC – mQTL – log10(P)
Fujita oligodendrocytes – SCAPER eQTL – log10(P)
Quach IAV monocytes – SCAPER eQTL – log10(P)

## S20.8

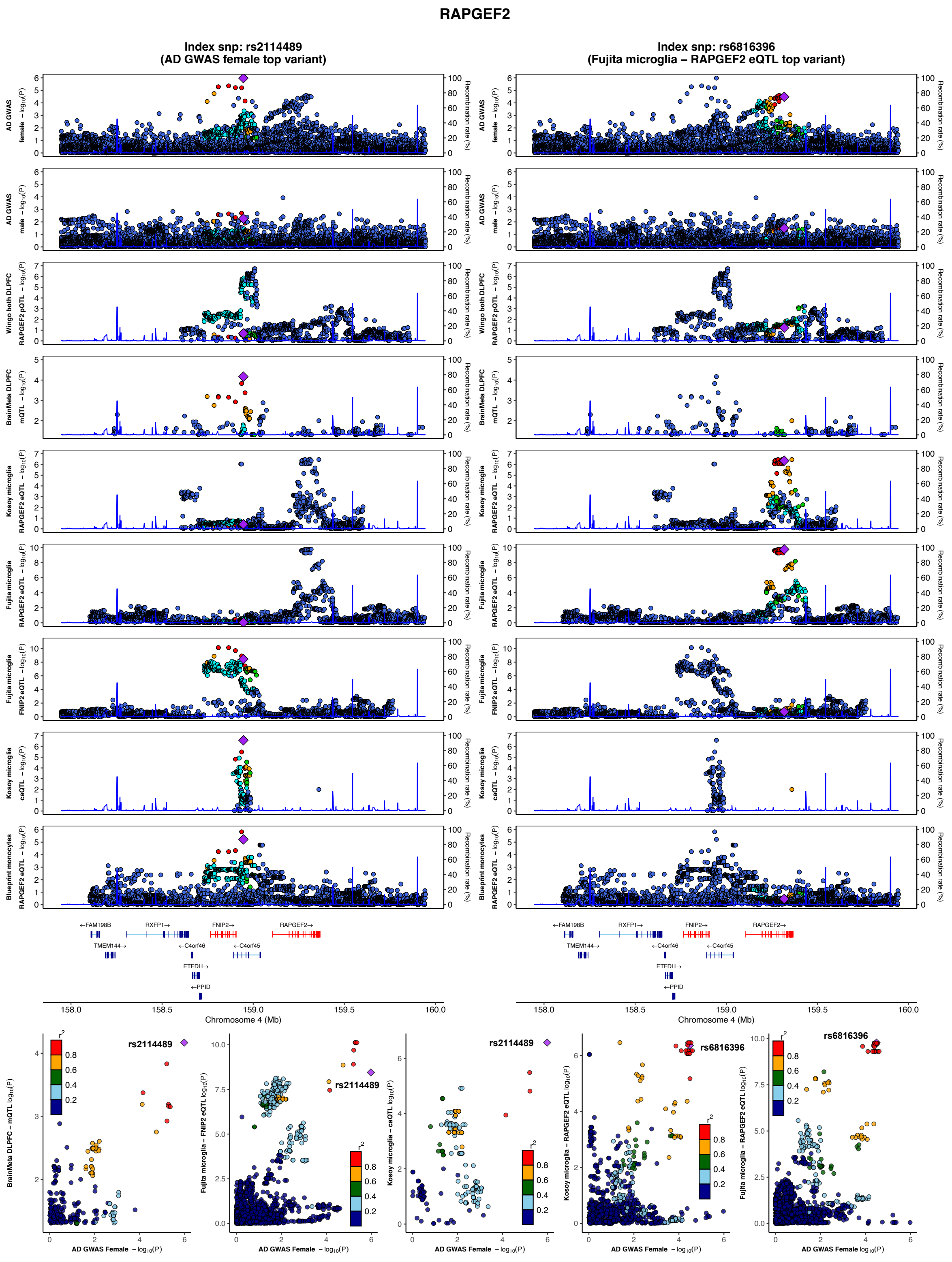
RAPGEF2
Index snp: rs2114489
(AD GWAS female top variant)
Index snp: rs6816396
(Fujita microglia – RAPGEF2 eQTL top variant)
AD GWAS female –log10(P)
AD GWAS male –log10(P)
Wingo both DLPFC RAPGEF2 pQTL –log10(P)
BrainMeta DLPFC mQTL –log10(P)
Kosoy microglia RAPGEF2 eQTL –log10(P)
Fujita microglia RAPGEF2 eQTL –log10(P)
Fujita microglia FNIP2 eQTL –log10(P)
Kosoy microglia caQTL –log10(P)
Blueprint monocytes RAPGEF2 eQTL –log10(P)
Recombination rate (%)
←FAM198B
RXFP1→
FNIP2→
RAPGEF2→
TMEM144→
←C4orf46
←C4orf45
ETFDH→
←PPID
158.0
158.5
159.0
159.5
160.0
Chromosome 4 (Mb)
rs2114489
rs6816396
r2
0.8
0.6
0.4
0.2
BrainMeta DLPFC – mQTL log10(P)
Fujita microglia – FNIP2 eQTL log10(P)
Kosoy microglia – caQTL log10(P)
Kosoy microglia – RAPGEF2 eQTL log10(P)
Fujita microglia – RAPGEF2 eQTL log10(P)
AD GWAS Female – log10(P)

**Supplementary Figure 20. Illustrative AD PWAS locus zoom plots highlighting xQTL colocalizations.** The illustrative PWAS hits included are **1)** TOM1L2; **2)** ARSA; **3)** SLC7A5; **4)** ADAM17; **5)** ENO3/RABEP1; **6)** PHB1; **7)** ETFA; **8)** RAPGEF2. Figures with more than one LocusZoom column indicate the presence of more than one independent signal at the locus contributing to QTL colocalizations. Dot color coding represents LD with tagging variant (purple diamond): red – $R^2$: 0.8-1.0, orange – $R^2$:0.6-0.8, green – $R^2$:0.4-0.6, cyan – $R^2$:0.2-0.4, blue – $R^2$:0.0-0.2.

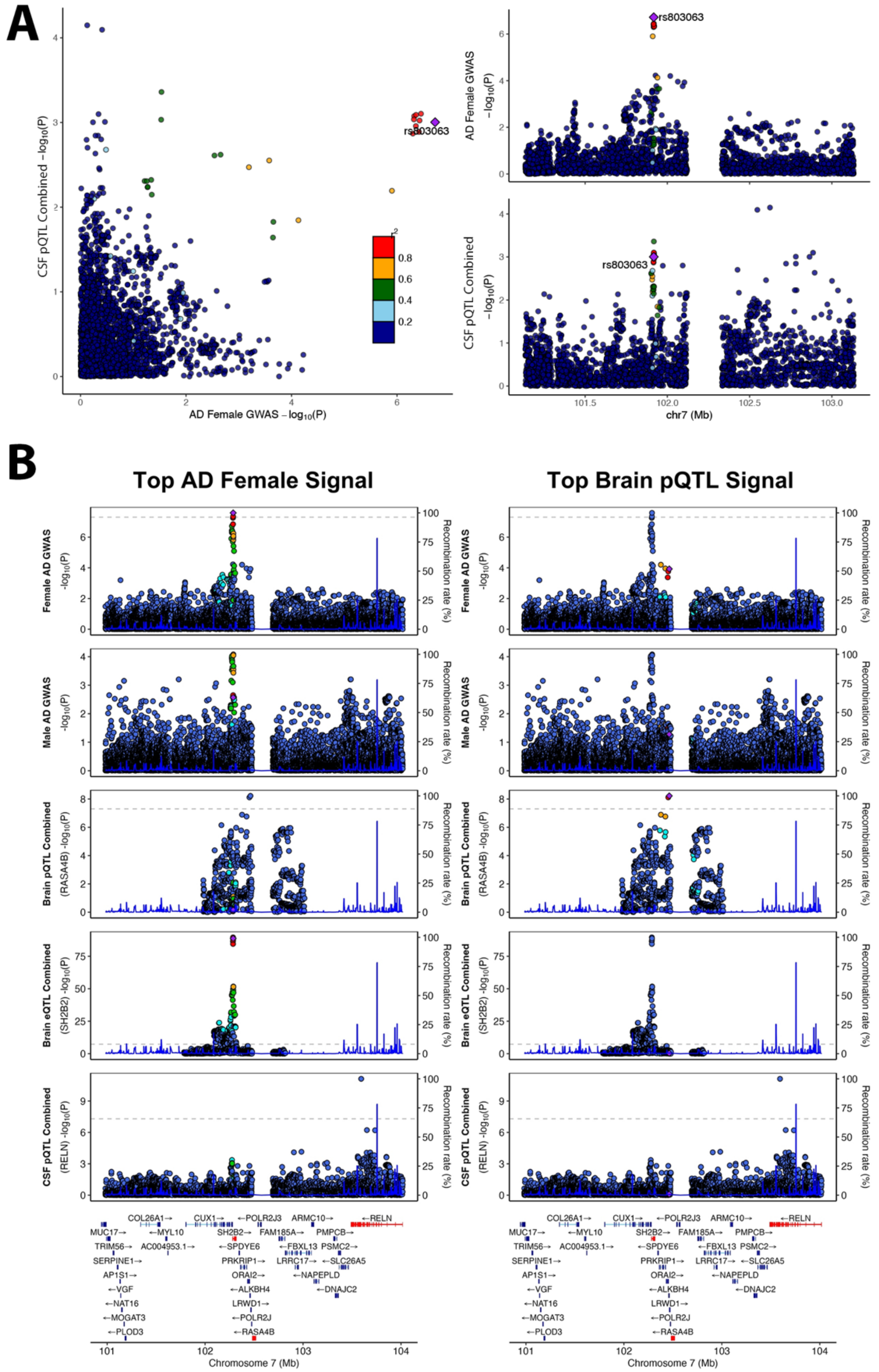
A
rs803063
CSF pQTL Combined -log10(P)
AD Female GWAS -log10(P)
AD Female GWAS -log10(P)
CSF pQTL Combined -log10(P)
chr7 (Mb)
B
Top AD Female Signal
Top Brain pQTL Signal
Female AD GWAS -log10(P)
Male AD GWAS -log10(P)
Brain pQTL Combined (RASA4B) -log10(P)
Brain eQTL Combined (SH2B2) -log10(P)
CSF pQTL Combined (RELN) -log10(P)
Recombination rate (%)
Chromosome 7 (Mb)

**Supplementary Figure 21. Follow-up inspection of the RASA4B PWAS finding that showed colocalization with RELN in CSF pQTL data. A)** In our global approach, QTL colocalization analyses for PWAS findings were performed on a 2MB window centered on the area for which protein weights were available. In this window, colocalization with *RELN* in CSF pQTL data reached PP4=0.71, through a suggestive pQTL signal (*P*~1e-3). Notably, this signal was more than 1Mb away from *RELN*. **B)** Locus zoom plots are shown for the RASA4B PWAS locus, on the left using the top AD female GWAS lead variant as a tagging variant, and on the right using the RASA4B brain pQTL lead variant as a tagging variant. An expanded 3Mb window was used to visualize the location of *RELN*. The RASA4B pQTL signal showed some overlap with a secondary signal in the AD female GWAS, but not the primary stronger AD female GWAS signal (and PP4 values suggested no colocalization). The colocalization with *SH2B2* brain eQTL data was much more striking. In expanding the window, it was apparent that another independent, much stronger CSF pQTL signal was present for RELN, relative to the suggestive pQTL signal that appeared to colocalize with the top AD female GWAS signal. Overall, the evidence appears more in favor that regulation of *SH2B2* brain expression underlies the AD female association but suggests *RASA4B* and *RELN* may also play causal roles at the locus. Dot color coding represents LD with tagging variant (purple diamond): red – $R^2$: 0.8-1.0, orange – $R^2$:0.6-0.8, green – $R^2$:0.4-0.6, cyan – $R^2$:0.2-0.4, blue – $R^2$:0.0-0.2.

### Enrichment analyses

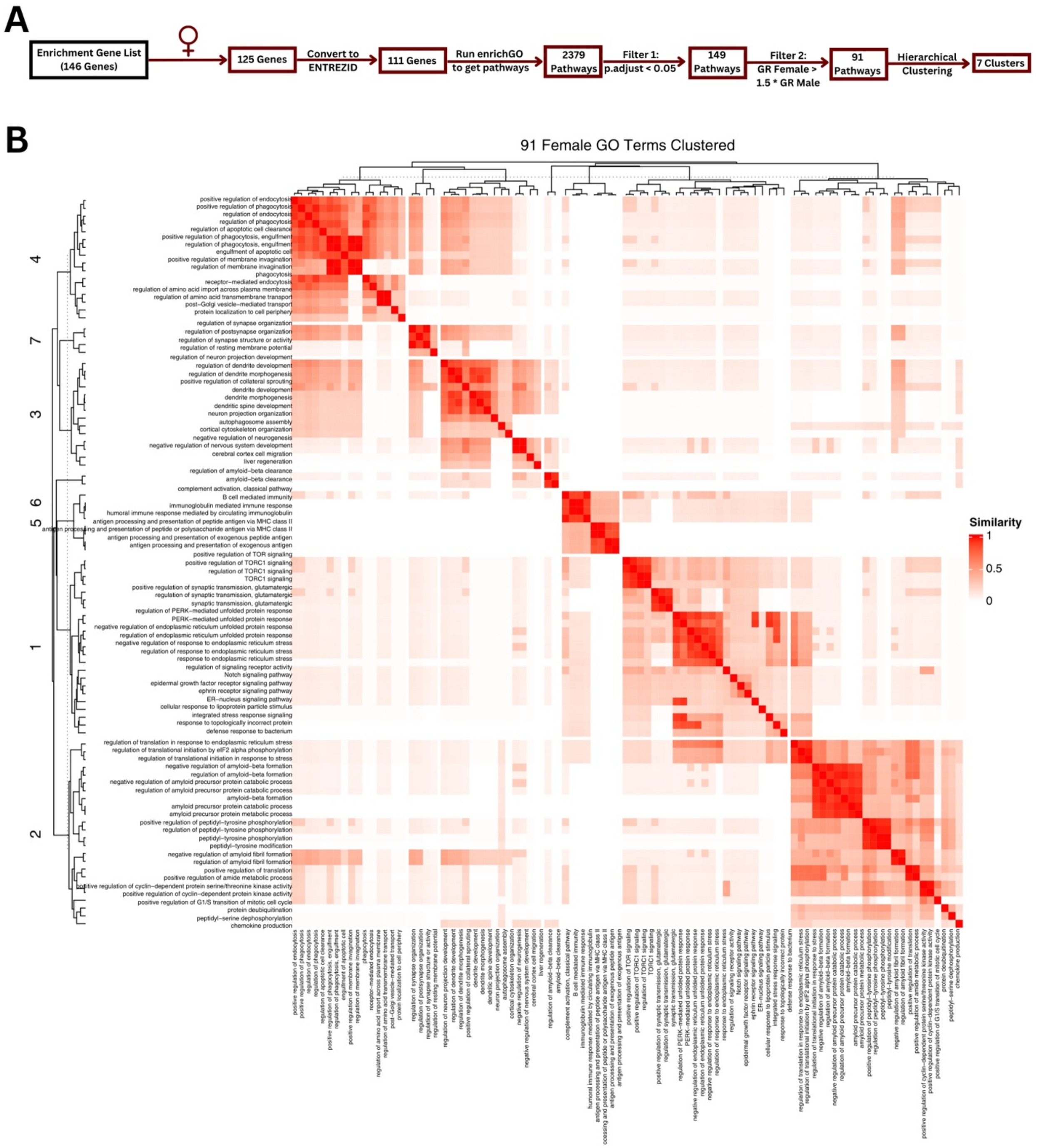

**Supplementary Figure 22. Pathway enrichment filtering and clustering for female-biassed genes. A)** Filtering process for 126 female-bias genes resulting in 7 pathway clusters. Pathways were filtered for sex specificity. **B)** Semantic similarity heat matrix for female-specific pathway clusters.

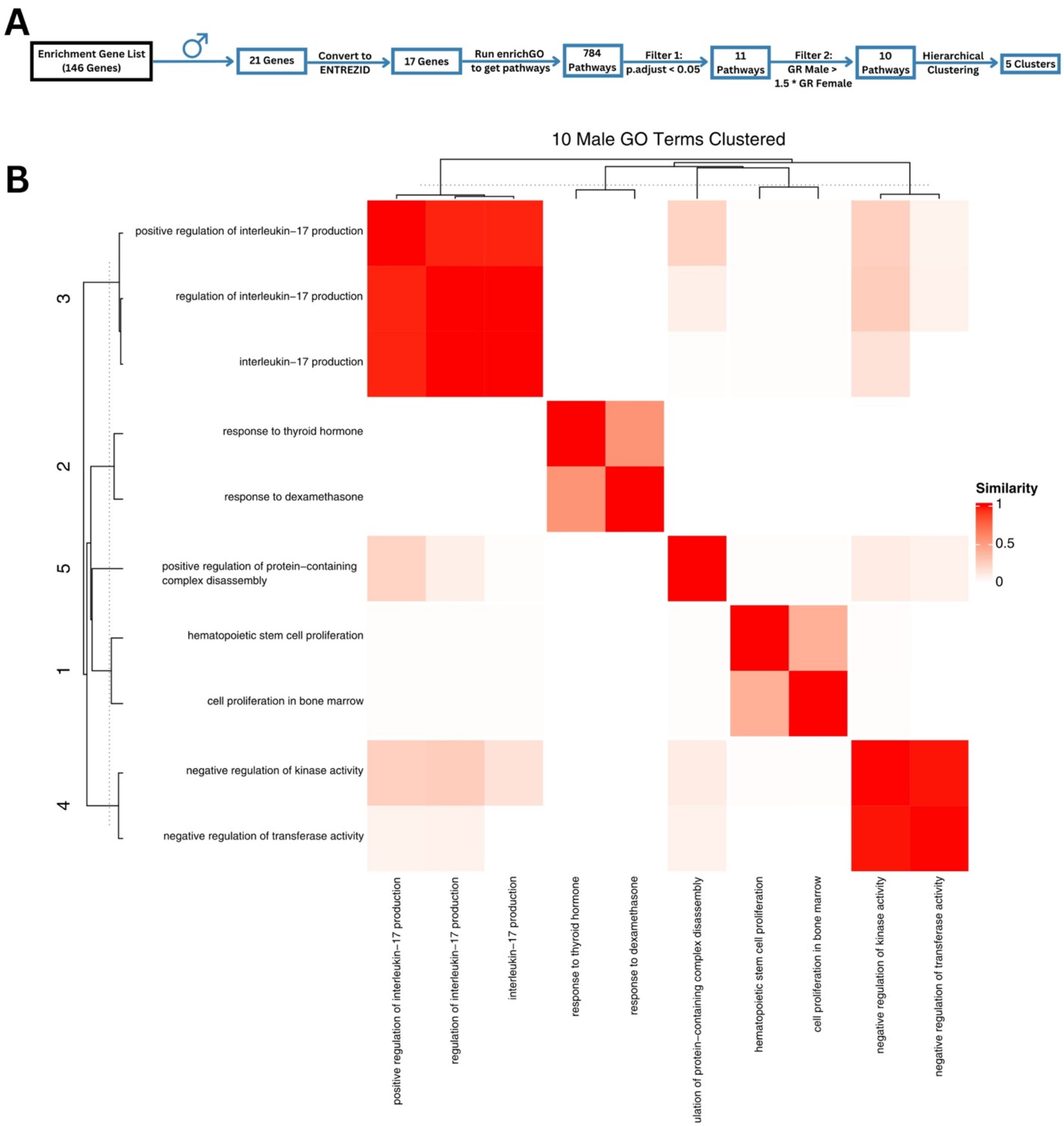

**Supplementary Figure 23. Pathway enrichment filtering and clustering for male-biassed genes. A)** Filtering process for 21 male-bias genes resulting in 5 pathway clusters. Pathways were filtered for sex specificity. **B)** Semantic similarity heat matrix for male-specific pathway clusters.

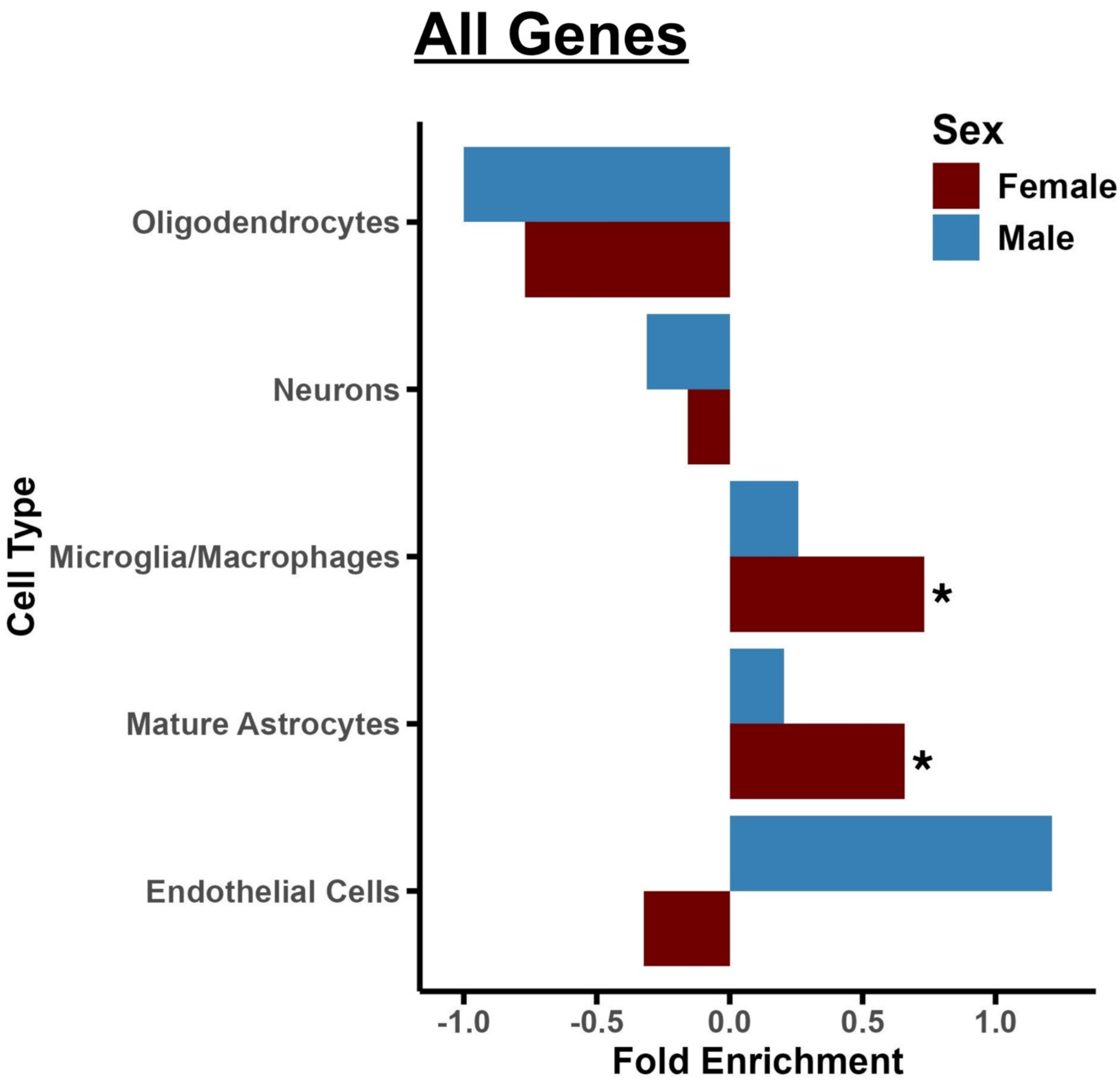

**Supplementary Figure 24. Brain cell-type specific enrichment of prioritized genes across sexes.** Extending on figure 5B, results are shown across all five available cell types (y-axis) with x-axis indicating fold enrichment. Red bars represent female results, while blue bars represent male results. Stars above bars denote a hypergeometric test *P*<0.05.

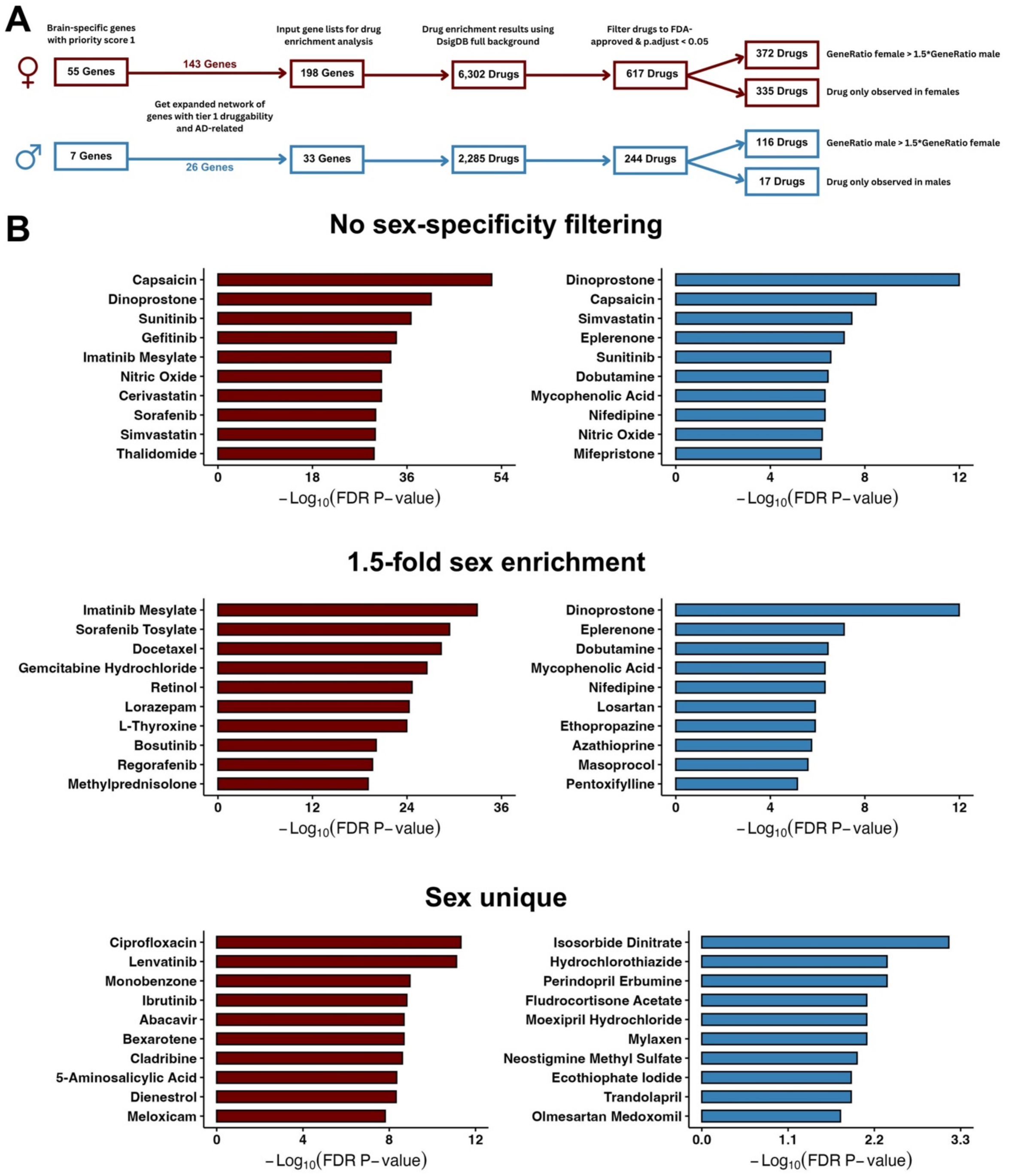

**Supplementary Figure 25. Drug repurposing and enrichment for sex-biased Alzheimer's disease genes. A)** Flow chart illustrating the number of genes and drugs from the drug enrichment analysis, detailing the progression and filtering process. **B)** Bar plots showing the top 10 enriched drugs for females (left, red) and males (right, blue). First row shows results before sex specificity filtering, the second row after applying a 1.5-fold sex-enrichment filter, and the third row highlighting drugs uniquely associated for each sex. Y-axes list drugs and x-axes indicate the -log10($P_{FDR}$; values >1.3 correspond to $P_{FDR}$ < 0.05.

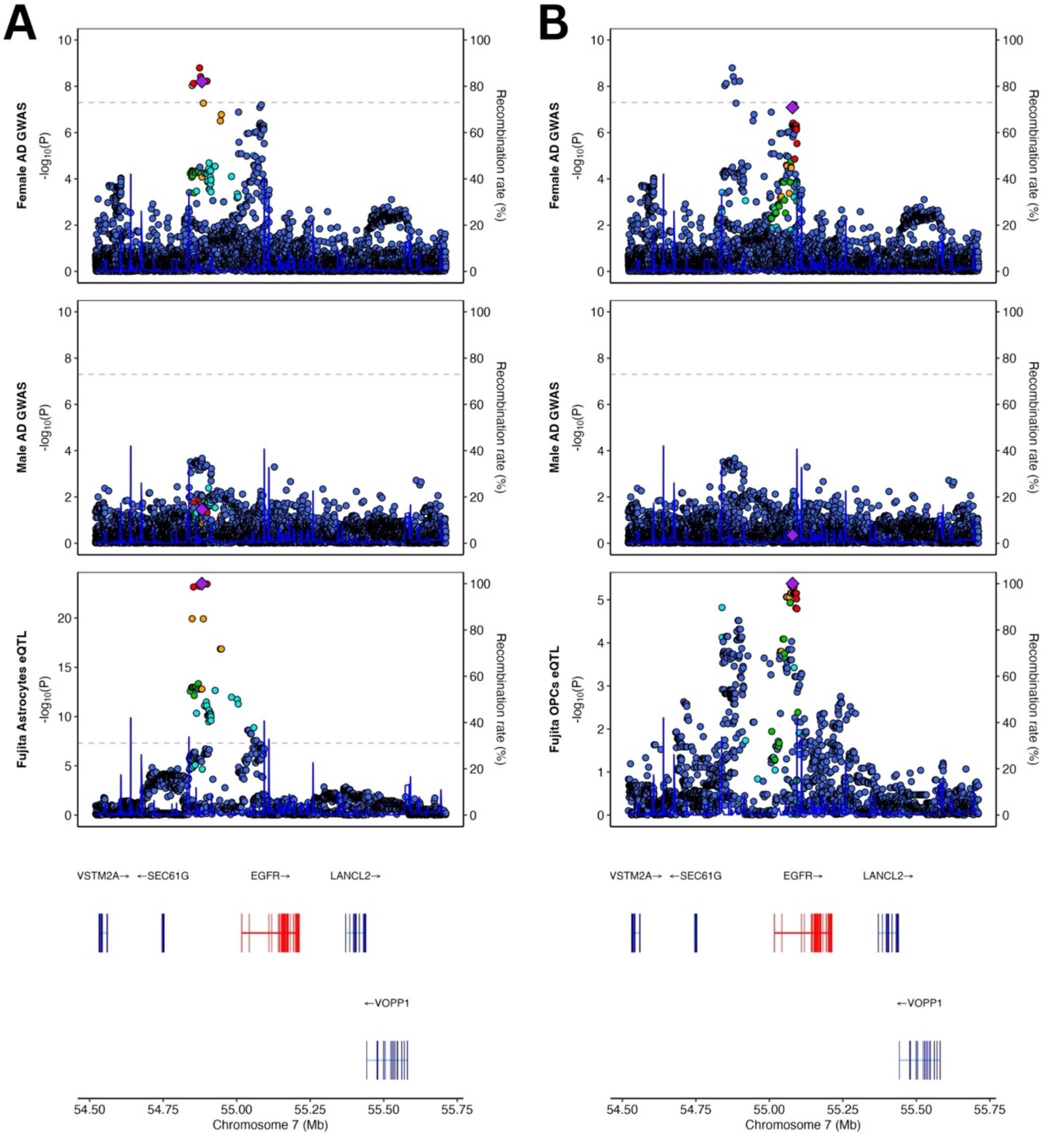

**Supplementary Figure 26. Colocalization of AD GWAS signals with *EGFR* eQTLs across brain cell types.** Locus zoom plots are shown for the *EGFR* locus for female and male AD GWAS signals, highlighting colocalization with *EGFR* eQTLs in astrocytes and oligodendrocyte precursor cells (OPCs). **A)** The top female-biased AD signal colocalized with *EGFR* eQTLs in astrocytes; **B)** A novel female-specific AD signal colocalized with *EGFR* eQTLs in OPCs. Dot color coding represents LD with tagging variant (purple diamond): red – $R^2$: 0.8-1.0, orange – $R^2$:0.6-0.8, green – $R^2$:0.4-0.6, cyan – $R^2$:0.2-0.4, blue – $R^2$:0.0-0.2.

#### Haptoglobin follow-up

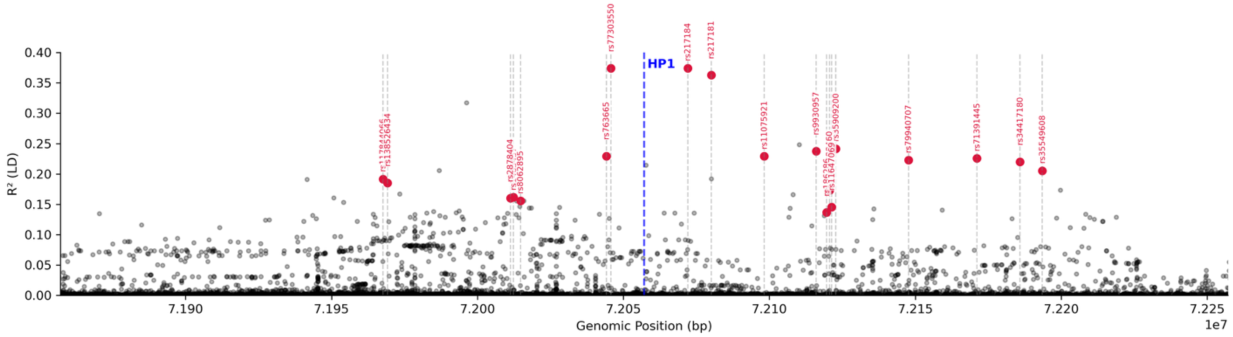

**Supplementary Figure 27. Linkage disequilibrium (LD) landscape within ±200 kbp of the HP1 allele deletion (chr16:72057133, DEL, –1716 bp), measured in the Stanford cohort.** LD is represented as $R^2$ (y-axis) between the deletion and surrounding SNVs. SNVs used for HP1 imputation (in red) are located within 200 kbp of the deletion and present across all AD GWAS stages 1 and 2 cohorts and CSF and plasma proteogenomics cohorts. In the Stanford cohort, SNVs were genotyped via short-read sequencing, while the deletion genotype was called using long-read sequencing.

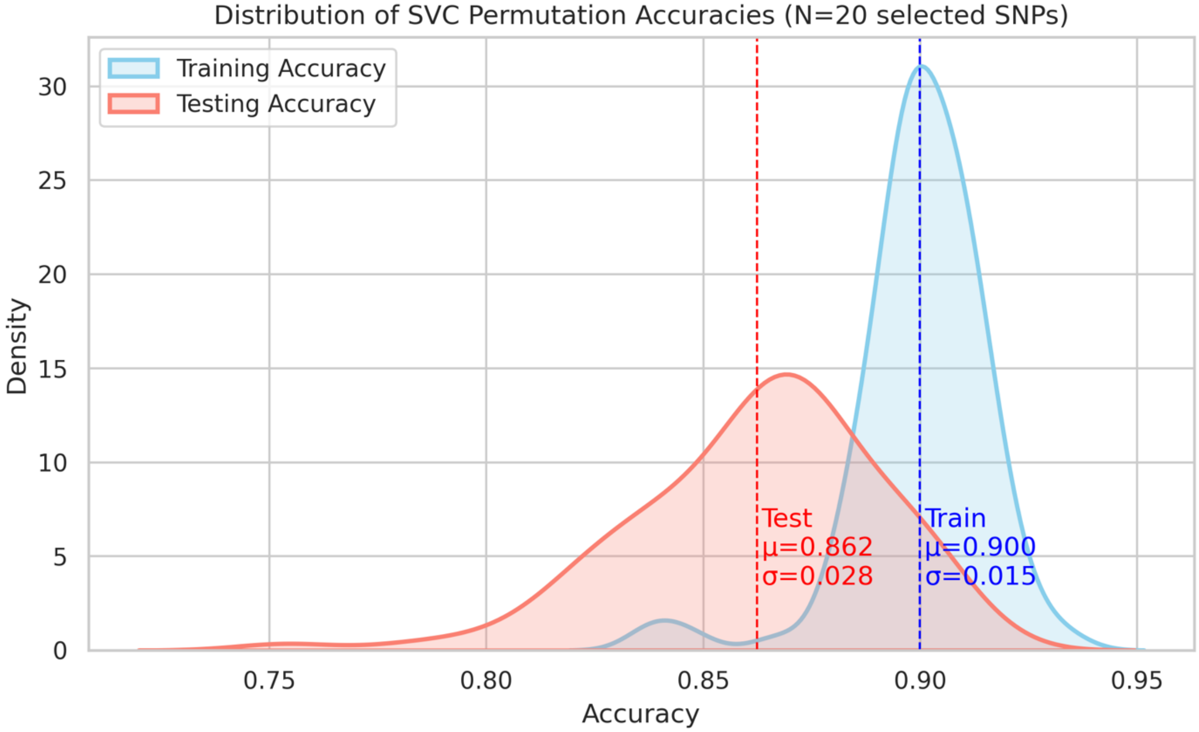

**Supplementary Figure 28. Distribution of support vector classifier (SVC) permutation accuracies for HP1 allele prediction/imputation using the top 20 most frequently selected SNPs.** Kernel density estimates (KDE) are shown for training (blue) and testing (red) accuracy across 100 permutations. Vertical dashed lines represent the mean accuracy for each distribution. Mean and standard deviation ($\mu \pm \sigma$) values are annotated: training ($\mu = 0.900$, $\sigma = 0.015$) and testing ($\mu = 0.862$, $\sigma = 0.028$). The classifier was trained using a polynomial kernel (degree = 3) after variance and univariate feature selection, and stratified 70/30 train-test splits were repeated across permutations.

**Top AD Female Signal (rs79940707)** | **Top CSF & Plasma pQTL Signal (rs77303550)** | **Top Brain pQTL Signal (rs34042070)**

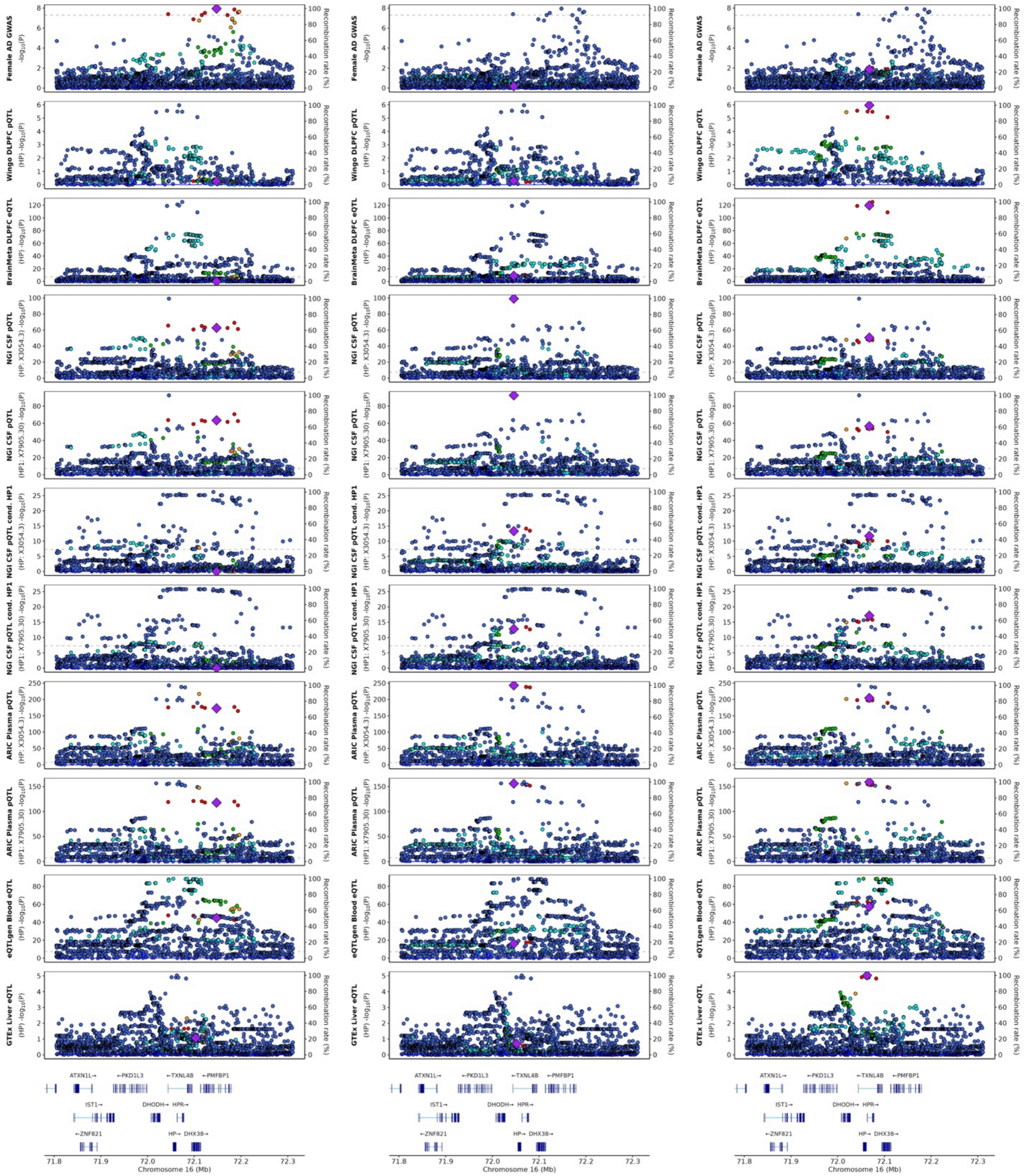

**Supplementary Figure 29. Locus zoom plots for multi-tissue QTL signals at the *HP* locus.** Each column represents a different LD tagging variant: the left column tags the top Female AD GWAS variant (which tags HP1F), the middle column tags the top CSF and plasma pQTL variant (which tags HP1S), and the right column tags the top brain pQTL signal. Each row corresponds to a GWAS or QTL signals, including: Female AD GWAS (row 1), DLPFC pQTL (row 2), DLPFC eQTL (row 3), CSF pQTL for analyte X3054.3 - HP (row 4), CSF pQTL for analyte X7905.30 - HP1 (row 5), CSF pQTL for analyte X3054.3 – HP - conditioned on HP1 genotype (row 6), CSF pQTL for analyte X7905.30 – HP1 - conditioned on HP1 genotype (row 7), Plasma pQTL for analyte X3054.3 - HP (row 8), Plasma pQTL for analyte X7905.30 – HP1 (row 9), Blood eQTL (row 10), and Liver eQTL (row 11). Dot color coding represents LD with tagging variant (purple diamond): red – $R^2$: 0.8-1.0, orange – $R^2$:0.6-0.8, green – $R^2$:0.4-0.6, cyan – $R^2$:0.2-0.4, blue – $R^2$:0.0-0.2.

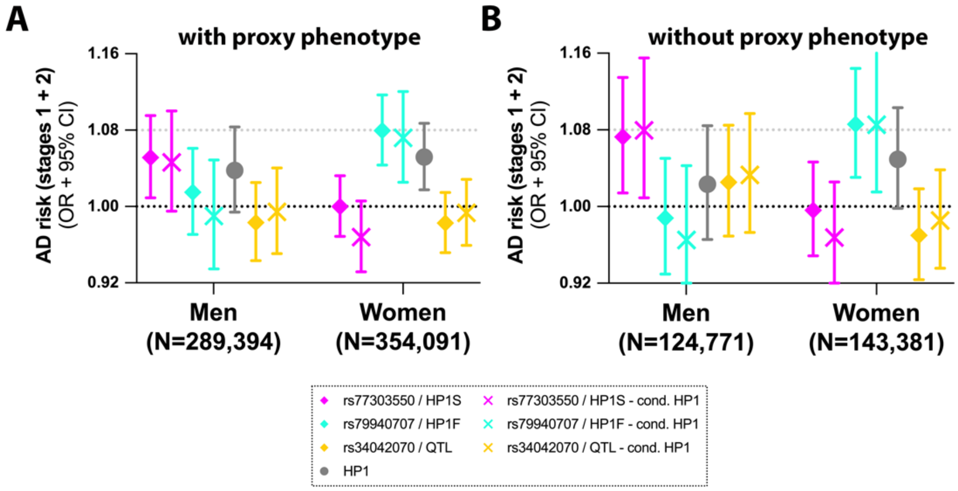

**Supplementary Figure 30. HP1 allele related analyses in AD GWAS data.** This figure extends on main Figure 6, providing AD association results, across stages 1 and 2 of the GWAS data, for HP1, rs79940707 (HP1F), rs77303550 (HP1S), and rs34042070 (QTL). Analyses are shown including **(A)** family proxy AD phenotypes in stage 2, and **(B)** ignoring AD proxy phenotypes in stage 2 but instead using direct health registry data in subjects ages 60 and above.

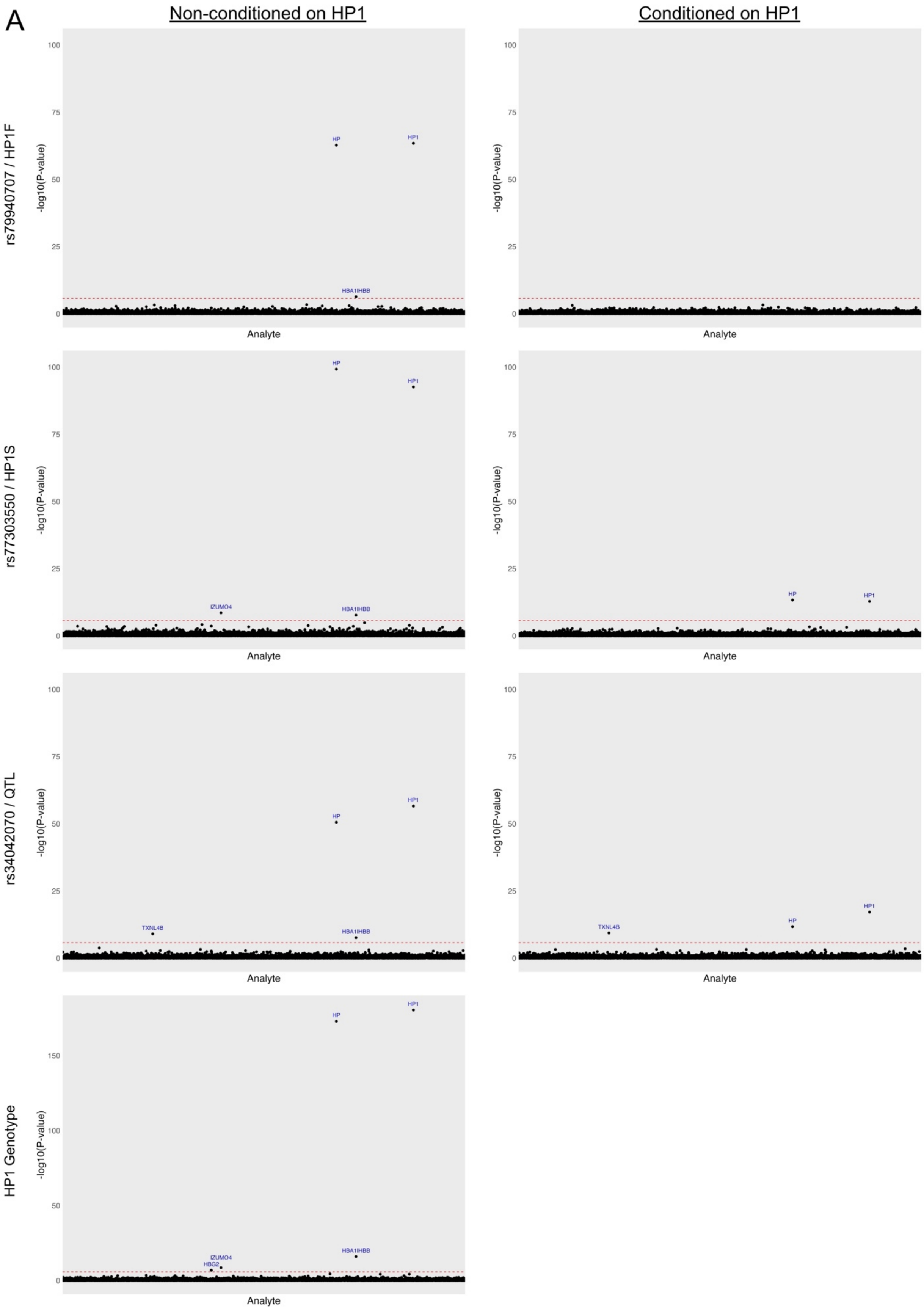
A
Non-conditioned on HP1
Conditioned on HP1
rs79940707 / HP1F
rs77303550 / HP1S
rs34042070 / QTL
HP1 Genotype
-log10(P-value)
Analyte
HP
HP1
HBA1|HBB
IZUMO4
TXNL4B
HBG2

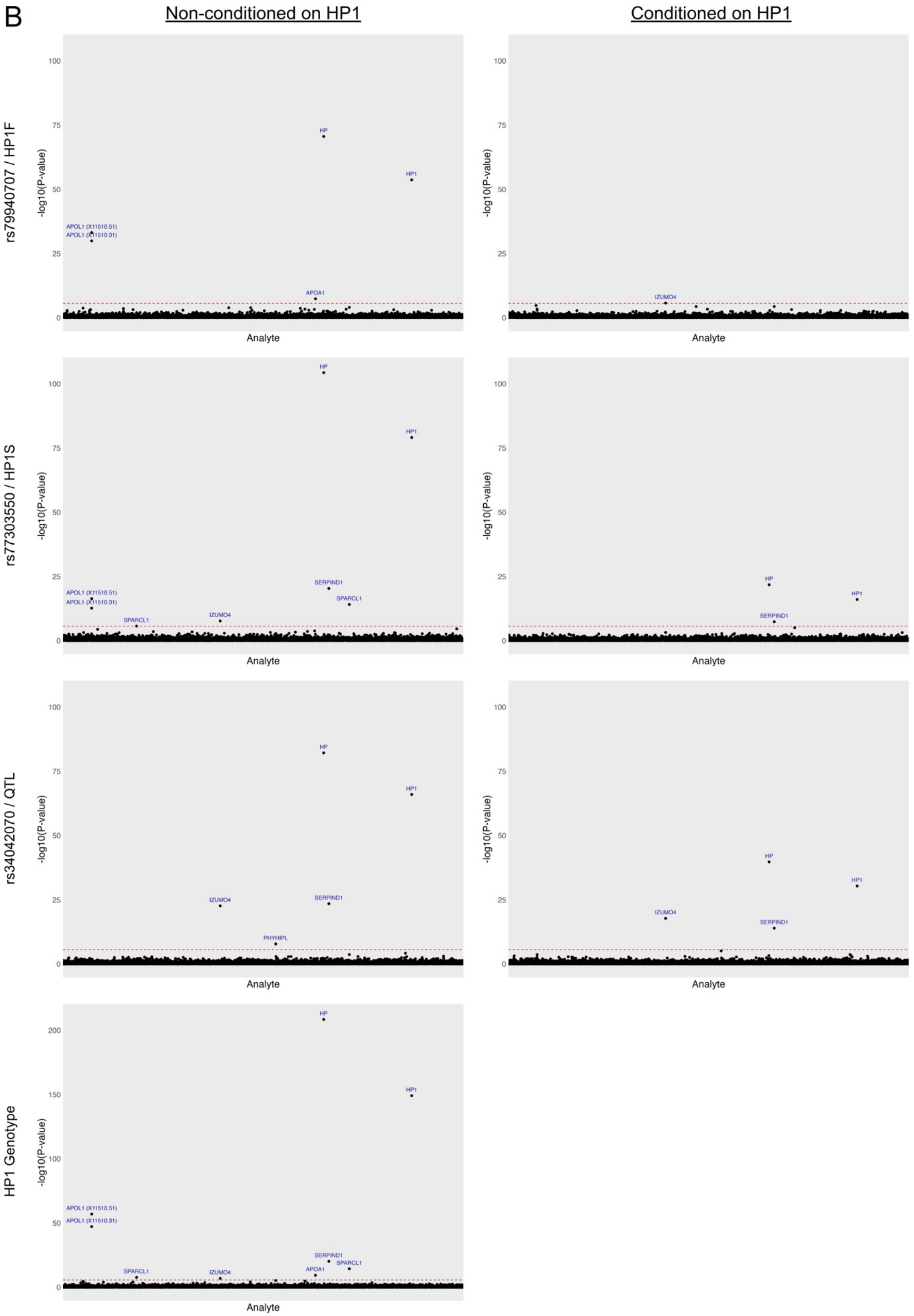
B
Non-conditioned on HP1
Conditioned on HP1
rs79940707 / HP1F
rs77303550 / HP1S
rs34042070 / QTL
HP1 Genotype
-log10(P-value)
Analyte
HP
HP1
APOL1 (X11510.51)
APOL1 (X11510.31)
APOA1
IZUMO4
SERPIND1
SPARCL1
PHYHIPL

**Supplementary Figure 31. PheWAS results for 3 *HP* QTLs and HP1 allele associated with CSF and plasma protein analytes.** Plots visualize genomic positions of proteins on the x-axis and -log10(*P*) on the y-axis for phenome-wide association study (pheWAS) analysis of **A)** CSF and **B)** plasma protein analytes. Points above the red dashed line indicate significance after Bonferroni correction (-log10(0.05/7008/4) for the left column and -log10(0.05/7008/3) for the right column in A and -log10(0.05/6907/4) for the left column and -log10(0.05/6907/3) for the right column in B) are labeled with the associated protein. The left column shows results not conditioned on HP1 allele, while the right column displays pheWAS results conditioning on HP1 allele.

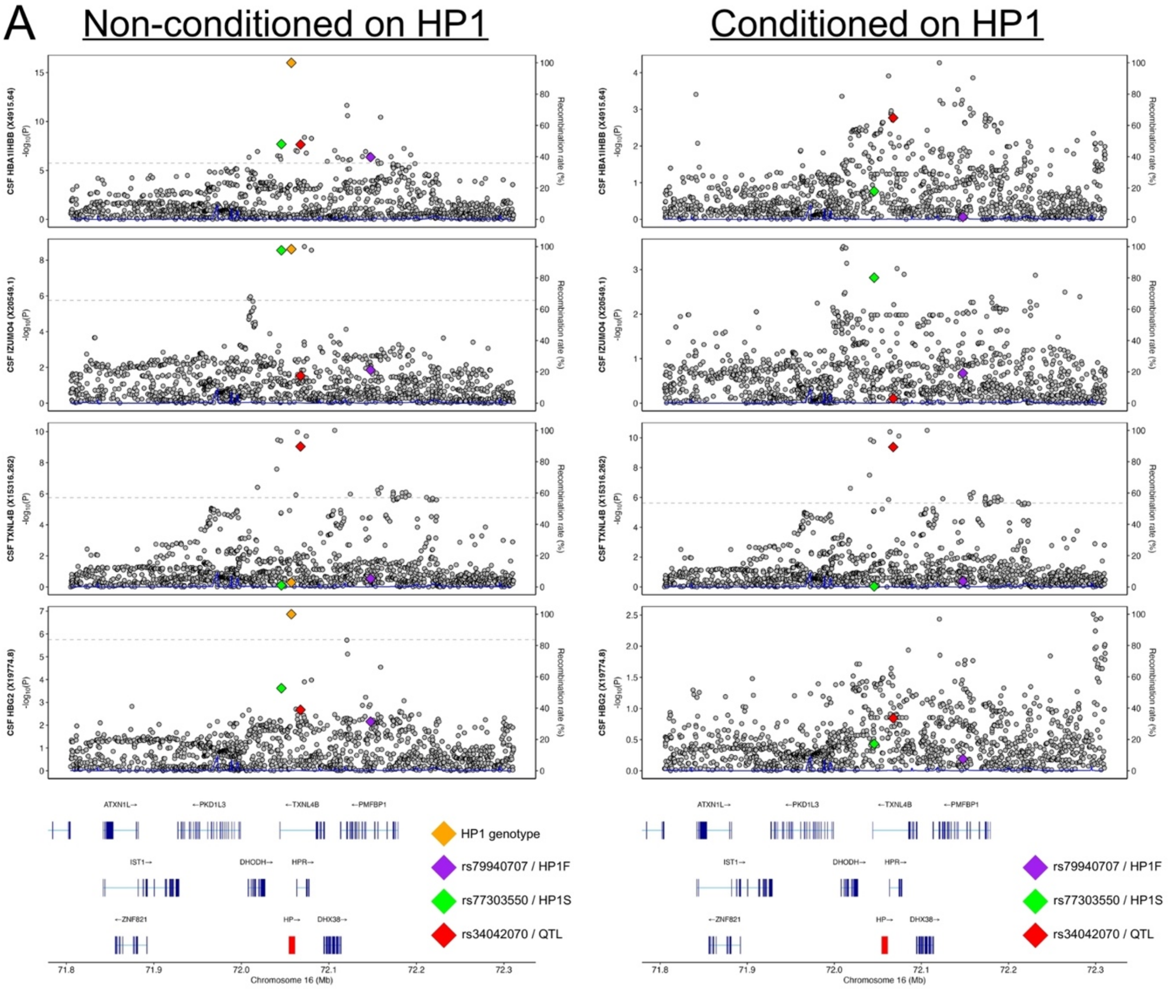
A
Non-conditioned on HP1
Conditioned on HP1
CSF HBA1IHBB (X4915.64)
CSF IZUMO4 (X20549.1)
CSF TXNL4B (X15316.262)
CSF HBG2 (X19774.8)
-log10(P)
Recombination rate (%)
ATXN1L→
←PKD1L3
←TXNL4B
←PMFBP1
IST1→
DHODH→
HPR→
←ZNF821
HP→
DHX38→
HP1 genotype
rs79940707 / HP1F
rs77303550 / HP1S
rs34042070 / QTL
Chromosome 16 (Mb)

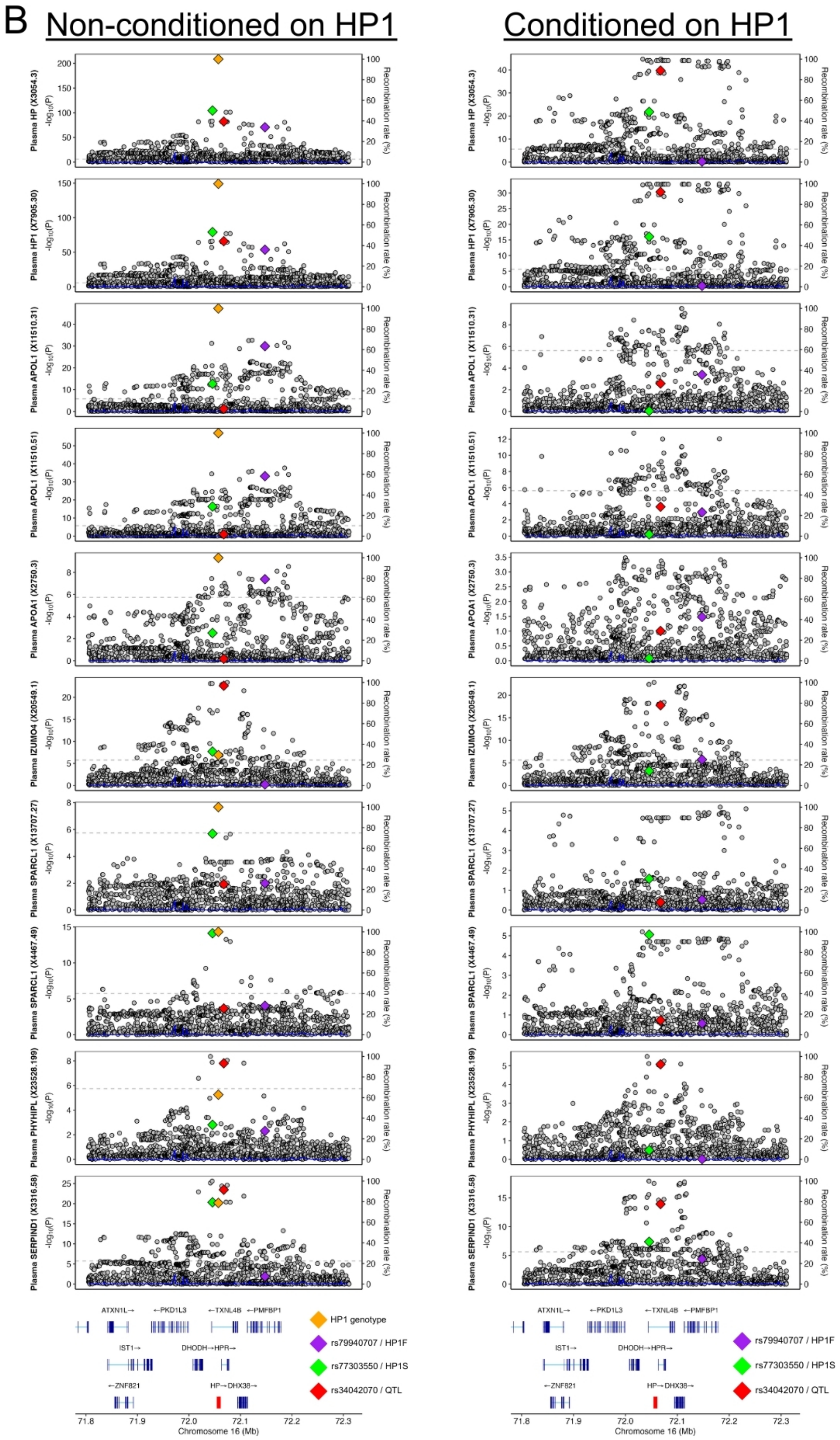
B
Non-conditioned on HP1
Conditioned on HP1
Plasma HP (X3054.3)
Plasma HP1 (X7905.30)
Plasma APOL1 (X11510.31)
Plasma APOL1 (X11510.51)
Plasma APOA1 (X2750.3)
Plasma IZUMO4 (X20549.1)
Plasma SPARCL1 (X13707.27)
Plasma SPARCL1 (X4467.49)
Plasma PHYHIPL (X23528.199)
Plasma SERPIND1 (X3316.58)
$-\log_{10}(P)$
Recombination rate (%)
ATXN1L→
←PKD1L3
←TXNL4B
←PMFBP1
IST1→
DHODH→HPR→
←ZNF821
HP→DHX38→
HP1 genotype
rs79940707 / HP1F
rs77303550 / HP1S
rs34042070 / QTL
Chromosome 16 (Mb)

**Supplementary Figure 32.** ***HP*** **locus zoom plots for significant proteins identified in CSF and plasma pheWAS.** This figure extends on Figure S31; **A)** CSF; **B)** plasma. Left column displays pheWAS results not conditioned on HP1 allele, while the right column shows results when conditioning on HP1 allele. Each plot illustrates the *HP* locus genomic region on the x-axis and -log10(*P*) on the y-axis for pheWAS results. Yellow diamonds represent the HP1 allele, purple diamonds HP1F, green diamonds HP1S, and red diamonds the top brain pQTL variant. Each row corresponds to the significant proteins from pheWAS analysis (cf. Figure S31 & Table S38).

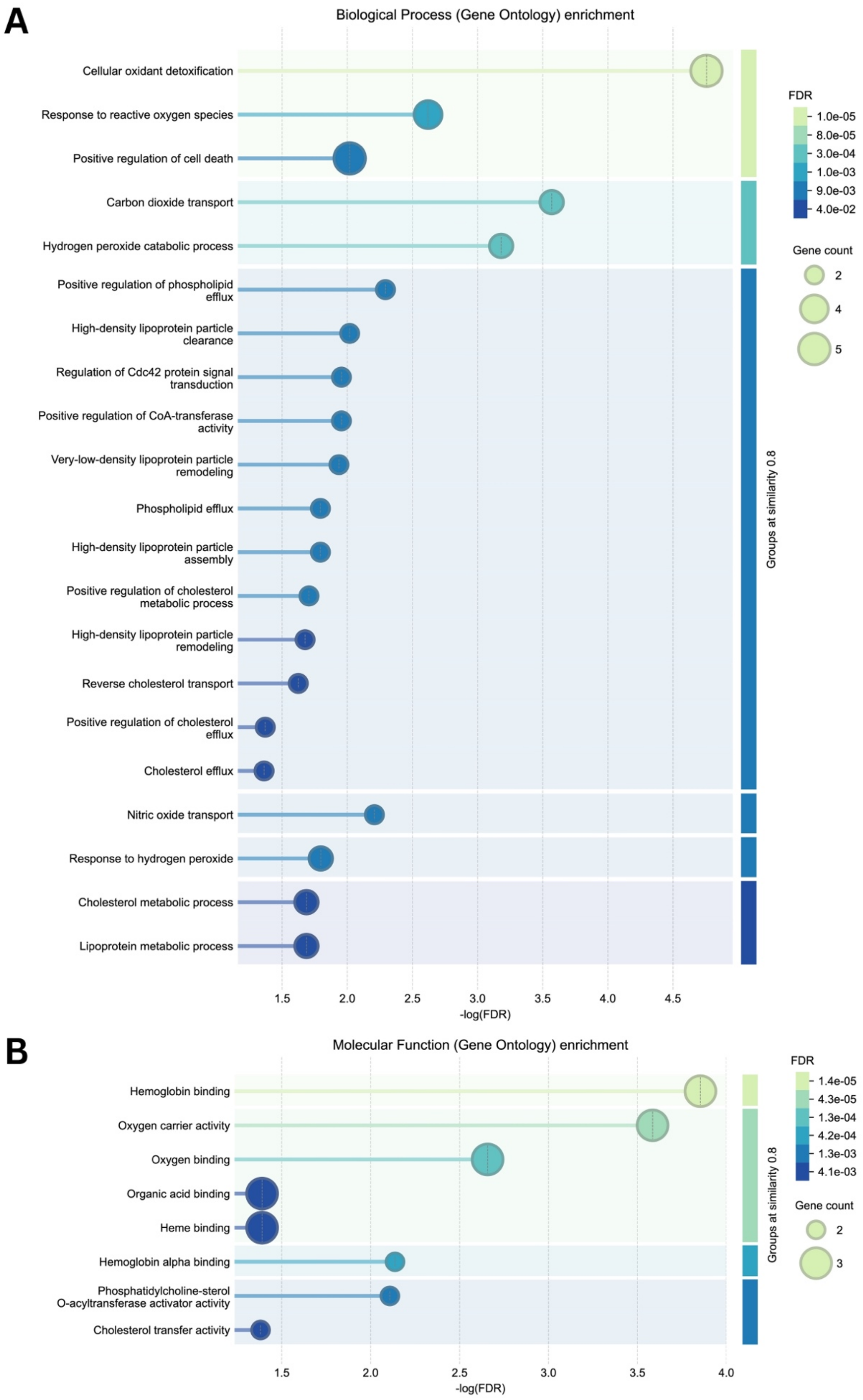
A
Biological Process (Gene Ontology) enrichment
Cellular oxidant detoxification
Response to reactive oxygen species
Positive regulation of cell death
Carbon dioxide transport
Hydrogen peroxide catabolic process
Positive regulation of phospholipid efflux
High-density lipoprotein particle clearance
Regulation of Cdc42 protein signal transduction
Positive regulation of CoA-transferase activity
Very-low-density lipoprotein particle remodeling
Phospholipid efflux
High-density lipoprotein particle assembly
Positive regulation of cholesterol metabolic process
High-density lipoprotein particle remodeling
Reverse cholesterol transport
Positive regulation of cholesterol efflux
Cholesterol efflux
Nitric oxide transport
Response to hydrogen peroxide
Cholesterol metabolic process
Lipoprotein metabolic process
1.5
2.0
2.5
3.0
3.5
4.0
4.5
-log(FDR)
Groups at similarity 0.8
FDR
1.0e-05
8.0e-05
3.0e-04
1.0e-03
9.0e-03
4.0e-02
Gene count
2
4
5
B
Molecular Function (Gene Ontology) enrichment
Hemoglobin binding
Oxygen carrier activity
Oxygen binding
Organic acid binding
Heme binding
Hemoglobin alpha binding
Phosphatidylcholine-sterol O-acyltransferase activator activity
Cholesterol transfer activity
1.5
2.0
2.5
3.0
3.5
4.0
-log(FDR)
Groups at similarity 0.8
FDR
1.4e-05
4.3e-05
1.3e-04
4.2e-04
1.3e-03
4.1e-03
Gene count
2
3

**Supplementary Figure 33. Functional enrichment analysis *HP* locus-associated proteins.** Input proteins for analysis include 10 significant proteins from pheWAS analysis (cf. Figure S31 and Table S38; excluding TXLN4B due to no link in protein-protein interaction network) and APOE due to literature and *APOE*-stratified analysis implicating its link with HP. **A)** Biological process (Gene Ontology) enrichment plot, showing significant biological processes on the y-axis and -log($P_{FDR}$) on the x-axis, with circle size representing gene count. **B)** Molecular Function enrichment plot, displaying significant molecular functions on the y-axis and -log($P_{FDR}$) on the x-axis, with circle size indicating gene count.

### Supplementary Tables

**Supplementary Table 1. Alzheimer's disease GWAS stage 1 ADGC & ADSP cohorts.** Overview of genotyping platforms across all stage 1 AD-related genetic data (ADGC and other cohorts).

**Supplementary Table 2. Alzheimer's disease GWAS stage 1 ADSP sub-cohorts.** Overview of ADSP cohorts available through NIAGADS DSS (NG00067).

**Supplementary Table 3. Alzheimer's disease GWAS EUR cohort demographics.**
*§ Across ADGC and ADSP, 40% of clinically diagnosed cases were additionally verified to have Alzheimer's disease pathology.*
*¶ In UKB, reported sex for proxy cases reflect: (1) Women - subjects with maternal AD status and no sibling or paternal AD status, (2) Men - subjects with paternal AD status and no sibling or maternal AD status.*

**Supplementary Table 4. Alzheimer's disease GWAS EUR quality-controlled variant counts.**
*§ ADGC imputed cohorts were merged and variants filtered to genotyping rate >50% and minor allele count > 20, equivalent to minor allele frequencies >= 0.09% and >= 0.07% in APOE*4 pos and APOE*4 neg respectively.*
*¶ ADSP variants were filtered to genotyping rate >20% and minor allele count > 2, followed by standard and ADSP-specific quality control.*
*‡ UKB and FinnGen variants underwent cohort specific QC and were then filtered to imputation scores > 0.3 and effect allele frequencies >= 0.05%.*
*# Meta-analyses were restricted to variants with genotyping rate >90% across all included cohorts.*

**Supplementary Table 5. Sex-stratified Alzheimer's disease GWAS: Linkage of sex-biased lead variants with prior AD lead variants.** Variants were considered to be in Linkage Disequilibrium (LD) when $R^2$>0.01. For some loci, multiple lead AD variants have been previously reported; known AD variants displayed in the table represent those that showed the highest LD. Two sex-biased lead variants (†) showed $R^2$>0.01 with known AD variants but still appeared to be independent signals (cf. Figure S4).

**Supplementary Table 6. Variant effect prediction (VEP) annotations for the EUR Alzheimer's disease GWAS sex-biased loci.** VEP annotation results for the significant sex-biased lead variants as well as their closely linked variants (LD $R^2$>0.8).

**Supplementary Table 7. Alzheimer's disease GWAS AFR cohort demographics.**

**Supplementary Table 8. Alzheimer's disease GWAS AFR quality-controlled variant counts.**
*§ ADGC and ADSP cohorts, per HISP/non-HISP group, had variants filtered to genotyping rate >95% and minor allele frequencies >= 1% in male and female strata respectively.*
*# Meta-analyses were restricted to variants with genotyping rate >90% across all included cohorts.*

**Supplementary Table 9. Sex-stratified Alzheimer's disease GWAS: Sex-biased European lead variants in African ancestry samples.** Sex heterogeneity consistency in African ancestry samples was determined by observing more significant p-values upon fixed effects meta-analyses of European and African sex heterogeneity effects across cohorts using GWAMA.

**Supplementary Table 10. EUR female Alzheimer's disease PWAS primary discovery results in brain.**

**Supplementary Table 11. EUR female Alzheimer's disease PWAS secondary discovery results in brain.**

**Supplementary Table 12. EUR female Alzheimer's disease PWAS primary discovery results in CSF.**

**Supplementary Table 13. EUR female Alzheimer's disease PWAS secondary discovery results in CSF.**

**Supplementary Table 14. EUR male Alzheimer's disease PWAS primary discovery results in brain.**

**Supplementary Table 15. EUR male Alzheimer's disease PWAS secondary discovery results in brain.**

**Supplementary Table 16. EUR male Alzheimer's disease PWAS primary discovery results in CSF.**

**Supplementary Table 17. EUR male Alzheimer's disease PWAS secondary discovery results in CSF.**

**Supplementary Table 18. Overlap of sex-biased PWAS findings across brain and CSF analyses.** Results for sex-specific genes that had significant ($P_{FDR}$<0.05) results in either brain or CSF PWAS show direction concordance for 5 out of 6 overlapping proteins in respective sex strata, including 3 observations of nominal significance (absolute Z-score > 1.965).

**Supplementary Table 19. Sex-stratified Alzheimer's disease PWAS: Sex-biased European proteins in African ancestry samples.** Observations of sex heterogeneity consistency are indicated. EUR findings were considered consistent in African ancestry analyses if the P-value in the initial female or male discovery improved after meta-analyzing the EUR and AFR findings, while the opposite sex maintained $P$>0.05. The most significant findings across the primary and secondary discoveries are reported when applicable.
[a]*Protein profiled in female-specific proteogenomic data but does not have significant SNP-heritability estimates in non-stratified or male-specific proteogenomic data. Values reported for male stratums were derived from analyses that integrated the male AD GWAS with female protein weights.*

**Supplementary Table 20. Sex-stratified Alzheimer's disease PWAS: Sex-biased European proteins in sensitivity analyses excluding stage 2 (UKB).** Findings were considered consistent in sensitivity analyses if the respective female or male protein discovery maintained $P<0.05$ and the opposite sex maintained $P>0.05$. The most significant findings across primary and secondary discoveries are reported when applicable.
[a]*Protein profiled in female-specific proteogenomic data but does not have significant SNP-heritability estimates in non-stratified or male-specific proteogenomic data. Values reported for male stratums were derived from analyses that integrated the male AD GWAS with female protein weights.*

**Supplementary Table 21. Summary-based Mendelian randomization (SMR) results for significant PWAS findings.** SMR was performed on significant PWAS findings using sex-stratified AD GWAS and pQTL datasets, with matched sex-specific pQTLs applied for primary PWAS discoveries and non-sex-specific pQTLs for secondary PWAS discovery. SMR results were considered significant if pval_FDR_SMR<0.05 and pval_HEIDI>0.05.

**Supplementary Table 22. Variant effect prediction (VEP) annotations for the EUR Alzheimer's disease PWAS sex-biased proteins.** VEP annotations for the top sex-specific pQTL SNPs identified in the primary EUR AD PWAS and variants within high LD ($R^2>0.8$). VEP annotation results for lead pQTL variants, for respective PWAS protein discoveries, as well as their closely linked variants (LD R2>0.8).

**Supplementary Table 23. Table overview of all QTL resources and their public identifiers.**

**Supplementary Table 24. Full xQTL COLOC results for all sex-biased Alzheimer's disease GWAS loci.**

**Supplementary Table 25. Full xQTL COLOC results for all sex-biased Alzheimer's disease PWAS loci.**

**Supplementary Table 26. Sex-biased differential abundance level analyses results.**

**Supplementary Table 27. Overview of prioritized sex-biased Alzheimer's disease genes.** A total of 125 genes were prioritized as female-specific, and 21 genes were prioritized as male-specific (see Methods for details on priority scoring). This list represents the input for enrichment analyses presented in Figure 5.
**ALPL was prioritized by the lead variant being a high impact missense variant (VEP, CADD = 29.6), while all other genes marked with `1*` for the priority score were prioritized based on prior AD literature support.*

**Supplementary Table 28. GO pathway enrichment results for female-biased pathways.** Results for pathway-specific enrichment analysis for gene ontology terms associated with female-biased genes. All pathways with p.adjust<0.05 are shown, with pathways passing sex specificity filter (GeneRatio Female > 1.5*GeneRatio Male) assigned to a cluster.

**Supplementary Table 29. GO pathway enrichment results for male-biased pathways.** Results for pathway-specific enrichment analysis for gene ontology terms associated with male-biased genes. All pathways with p.adjust<0.05 are shown, with pathways passing sex specificity filter (GeneRatio Male > 1.5*GeneRatio Female) assigned to a cluster.

**Supplementary Table 30. Cell-type specificity enrichment results.**

**Supplementary Table 31. Drug repurposing enrichment analysis comprehensive results for female-biased genes.** Summary of drug repurposing enrichment analyses conducted on female-biased genes, filtered for FDA approved drugs and p.adjust < 0.05.

**Supplementary Table 32. Drug repurposing enrichment analysis comprehensive results for male-biased genes.** Summary of drug repurposing enrichment analyses conducted on male-biased genes, filtered for FDA approved drugs and p.adjust < 0.05.

**Supplementary Table 33. Sex-stratified PWAS results for EGFR in brain and CSF.** PWAS results for EGFR, stratified by sex, tissue, and analysis type. *EGFR* was identified as a female-biased GWAS locus. Our PWAS discovery framework required a significant finding in one sex ($P_{FDR}$<0.5) to show no significance in the opposite sex (P>0.05), which is a strict filter to prioritize sex-biased genes. While EGFR did not pass this filter, it still displayed strongly female-biased PWAS results.

**Supplementary Table 34. Summary of minor allele frequency of the HP1 allele across GWAS and pQTL datasets.** Predicted genotype distribution of the HP1 deletion across cohorts using a common set of N = 19 SNVs. The table summarizes the predicted genotype frequencies for the HP1 deletion (chr16:72,057,133, DEL, -1716 bp) across multiple cohorts, including the ADRC/SAMS Stanford dataset (training and prediction) and GWAS and pQTL cohorts. The number of samples with missing predictions is indicated, corresponding to individuals who had at least one of the 19 required SNPs with missing genotype data (NaN), preventing model input.

**Supplementary Table 35. Effect sizes of *HP* locus variants of interest across multiple tissue QTLs.** Comparison of effect sizes and standard errors of three prioritized *HP* locus variants across male and female AD GWASs and various tissue QTL datasets. HP1S (rs77303550) and brain pQTL variant (rs34042070) had opposite effect directions across all QTL datasets.

**Supplementary Table 36. Summary of *HP* locus variants of interest associations with Alzheimer's disease risk by sex.** Comparisons of effect sizes and standard errors for HP1 and 3 prioritized variants at the *HP* locus across men and women, with and without proxy phenotypes in GWAS analyses. Nominally significant effect sizes (*P*<0.05) are highlighted in bold.

**Supplementary Table 37. Summary of *HP* locus variants of interest associations with Alzheimer's disease risk by *APOE* status.** Comparisons of effect sizes and standard errors for HP1 and 3 prioritized variants at the *HP* locus across *APOE* status, without proxy phenotypes in GWAS analyses. Nominally significant effect sizes (*P*<0.05) are highlighted in bold.

**Supplementary Table 38. Summary of significant proteins in CSF and plasma from *HP* locus pheWAS.** Phenome-wide association study (pheWAS) results for significant proteins in CSF ($P$<0.05/7008/4) and plasma ($P$<0.05/6907/4) for variants of interest.

**Supplementary Table 39. FinnGen authors and contact information.**
